# Federal Funding and Citation Metrics of Biomedical Research in the USA

**DOI:** 10.1101/2022.08.31.22279467

**Authors:** John P.A. Ioannidis, Iztok Hozo, Benjamin Djulbegovic

## Abstract

Both citation and funding metrics converge in shaping current perceptions of academic success. We aimed to evaluate what proportion of the most-cited USA-based biomedical scientists are funded by biomedical federal agencies and whether funded scientists are more cited than not funded ones. We linked a Scopus-based database on top-cited researchers (n=75,316 USA-based) and the NIH RePORTER database of 33 biomedical federal agencies (n=204,603 grant records) with matching based on name and institution. The 40,887 USA-based top-cited scientists who were allocated to any of 69 scientific subfields highly related to biomedicine were considered in the main analysis. The proportion of USA-based top-cited biomedical scientists (based on career-long citation impact) who had received any federal funding from biomedical research agencies was 63% for any funding (1996-2022), 21% for recent funding (2015-2022), and 14% for current funding (2021-2022). Respective proportions were 65%, 31%, and 21%, when top-cited scientists based on recent single year impact were considered. There was large variability across scientific subfields. No subfield had more than 31% of its top-cited USA-based scientists (career-long impact) currently funded. Funded top-cited researchers were overall more cited than non-funded top-cited scientists, e.g. mean (median) 14,420 (8983) versus 8,445 (4613) (p<0.001) and a substantial difference remained (, after adjusting for subfield and years since first publication. Differences were more prominent in some specific biomedical subfields. Overall, biomedical federal funding has offered support to approximately two-thirds of the top-cited biomedical scientists at some point during the last quarter century, but only a small minority of top-cited scientists have current federal biomedical funding. The large unevenness across subfields needs to be addressed with ways that improve equity, efficiency, excellence, and translational potential.

## INTRODUCTION

Citation metrics(1) are key indicators of research productivity in the academic reward system, despite their widely acknowledged limitations(2). Research funding is also considered as an important indicator of scientific success, perhaps even more so in the USA. Successful competition for research funding, particularly from the federal agencies such as the National Institutes of Health (NIH) is used by most US academic institutions as metrics in promotion, tenure, and recruitment(3). Institutions also incentivize their faculty to apply for federal funding because they cover multiple expenses through grant-related indirect costs.

Despite a clear importance of funding on research output, to date, there is no good large-scale empirical data on the relationship between funding and citation metrics across biomedical research. Some earlier work has shown that NIH-funded papers tend to attract more citations(4). However, other work has suggested that among scientists with extremely cited papers (those with >1000 citations), a large share are not funded by NIH(5). That analysis focused on extremely cited papers; however, it would be useful to obtain evidence on a large, comprehensive sample of highly-cited scientists considering their entire career impact as well as their recent citation impact. To assess to what extent highly-cited scientists in the USA in the biomedical sciences are funded by federal agencies. we merged a database of most-cited scientists(6) with the NIH RePORTER(7), a repository of the federal funded research projects of 33 US government agencies. Our main analysis assessed how many biomedical scientists had received any listed federal funding at any time since 1996, since 2015, and currently. Secondarily, we compared whether key citation indices differed between funded top-cited scientists and other top-cited scientists.

## METHODS

### Citations database

We used the August 2021 update of the databases of the top-cited scientists across science according to Scopus data(6). Data were used separately for career-long citation impact and for single recent year citation impact (citations received in calendar year 2021 only). The databases and code are freely available in Mendeley (https://dx.doi.org/10.17632/btchxktzyw, https://data.mendeley.com/datasets/btchxktzyw/3).

For details on the construction of the database see refs. 8-10. The database includes all the scientists who rank among the top 100,000 across all fields according to a validated composite citation index^8^ that considers 6 citation indicators (total citations, Hirsch h index, Schreiber co-authorship-adjusted hm index, citations to papers as single author, citations to papers as single or first author, and citations to papers as single, first, or last author), as previously described(8-10) or they are within the top 2% of scientists of their main subfield discipline among authors who have published at least five papers. The classification of science into fields and subfields follows the Science-Metrix classification with machine learning classification of multidisciplinary journals, as previously described(11). All citation metrics are computed without exclusion of self-citations. Less than 5% of the scientists who are in the top 2% of their subfield for career-long impact when self-citations are included are no longer in the top 2% of their subdiscipline when self-citations are excluded, and only 0.01% fall below the top 10%.

There are 174 subfields to which science is divided according to the Science-Metrix classification. We pre-specified 69 subfields as being highly related to biomedicine. For the main analysis, we considered as biomedical scientists those who are assigned primarily to any of these 69 subfields, i.e. the most common subfield for their publications is one of these 69 subfields.

### NIH RePORTER database

The NIH RePORTER(7) is a repository of the federal funded research projects from 33 US government agencies (27 NIH institutes, the Agency for Healthcare Research and Quality, Centers for Disease Control and Prevention, Food and Drug Administration, Health Resources and Services Administration, Administration for Children and Family, and U.S. Department of Veterans Affairs). We downloaded data on 6/11/2022. Funded projects cover the period from 1985 until 2022, but we used only the data for grants from 1996 until 2022 to be more in line with the coverage of the citation database (Scopus data are more complete from 1996 forward).

### Matching process

If the name in the citations database and NIH RePORTER database is exactly the same, we considered the scientists matched. To slightly improve on this “brute force matching” we added three criteria for matching missing middle initials: whenever a name in RePORTER is given with full first and middle names, we try to match it to citations database with the same last name and first name and middle name initial; whenever a name is given in RePORTER with last name and full first name but no middle name or initial, we try to match it to the citations database with the same last name and first name (ignoring any middle name info in the citations database); and whenever a name is given in RePORTER with last name and full first name and a middle name initial, we try and match it to the citations database with the same last name and first name (provided any middle name or initial in the citations database is not inconsistent with the RePORTER middle name initial).

We evaluated random samples of scientists who were matched and of scientists who were not matched, in the matching process between the career-long citation impact database and the entire RePORTER database. This allowed to estimate rates of false matching and of missed matching and they seemed acceptable - for details see Supplementary text.

### Analyses

We considered all 6 possible combinations of matching different citations and funding data. These included the consideration of the career-long citation impact and, separately, the single recent year citation impact; and of funding at any time (having any active grants in years 1996-2022), recent (having any active grant in years 2015-2022), and current (having any active grant in years 2021-2022). In all 6 analyses, the main outcome of interest was the proportion of top-cited scientists who had funding. We evaluated these proportions overall in all biomedical scientists and in each of the 69 Science Metrix-defined subfields highly related to biomedicine.

We also evaluated whether in each analysis funded and not funded scientists differed in the total number of citations and in the composite citation indicator. Comparisons used the Mann Whitney U test and medians are presented (also means and sd for normally-distributed variables). Given that older scientists have more time to obtain grants and also to accrue citations, we also performed regressions adjusting for the publication age of the scientist (number of years since the year of the first publication). We performed linear regression, mixed-effect linear regression and mixed-effect linear regression of log-transformed citation counts. The mixed model added random effect variation which accounts for variability of our measurements for scientists in each subfield. Results are largely similar and here we report mixed-effects results with untransformed citation index values for ease of interpretation.

P-values are 2-tailed with statistical significance at p<0.005. Analyses were run in Stata.

## RESULTS

### Overall proportion of top-cited scientists funded

We started with 204,603 records in the RePORTER database for 1996-2022 and 186,177 records in the career-long citations database, of which 75,316 were USA-based scientists (40,887 classified in the 69 subfields highly related to biomedicine). Among subfields highly related to biomedicine (Table 1), the proportion of top-cited career impact scientists who had received any federal funding from biomedical research agencies was 62.7% (95% confidence interval, 62.3-63.2%) for any funding in 1996-2022, 23.1% (22.7-23.5%) for recent funding, and 14.1% (13.8-14.5%) for current funding. Respective proportions were 64.8% (64.2-65.2%), 31.4% (31.0-31.9%), and 20.9% (20.4-21.3%), when top-cited scientists based on recent single year impact were considered. For the other subfields, biomedical funding was overall very low and did not exceed 15% in any of the analyses (Table 1).

**Table 1.** Proportion funded in 69 subfields of science that are highly related to biomedicine and 105 other subfields of science Funding time is defined as ‘any’: any grant entry in RePORTER in 1996-2022; ‘recent’: any entry in the RePORTER that covers any year in the period 2015 until 2022; ‘current’: any entry in the RePORTER that covers 2021 and/or 2022. Highly related fields (according to the Science-Metrix classification) are 69 subfields, i.e. the 60 subfields within the larger fields of Biomedical Research, Clinical Medicine, Public Health and Health Services, and Psychology and Cognitive Sciences as well as 9 prespecified subfields from other large fields, i.e. Applied Ethics, Bioinformatics, Biomedical Engineering, Biotechnology, Demography, Gender Studies, Medical Informatics, Medicinal and Biomolecular Chemistry, and Veterinary Sciences.

| **Top-cited US-based researchers** | **Funding time** | **Total** | **Funded (%)** |
| --- | --- | --- | --- |
| **Career** | **Any** | 75316 | 30471 (40%) |
| Fields highly related to biomedicine |  | 40887 | 25650 (63%) |
| Other fields |  | 34429 | 4821 (14%) |
| **Career** | **Recent** | 75316 | 1120 (15%) |
| Fields highly related to biomedicine |  | 40887 | 9427 (23%) |
| Other fields |  | 34429 | 1813 (5.3%) |
| **Career** | **Current** | 75316 | 6854 (9.1%) |
| Fields highly related to biomedicine |  | 40887 | 5778 (14%) |
| Other fields |  | 34429 | 1076 (3.1%) |
| **Recent year** | **Any** | 65560 | 27535 (42%) |
| Fields highly related to biomedicine |  | 35787 | 23172 (65%) |
| Other fields |  | 29773 | 4363 (15%) |
| **Recent year** | **Recent** | 65560 | 13195 (20%) |
| Fields highly related to biomedicine |  | 35787 | 11252 (31%) |
| Other fields |  | 29773 | 1943 (6.5%) |
| **Recent year** | **Current** | 65560 | 8656 (13%) |
| Fields highly related to biomedicine |  | 35787 | 7465 (21%) |
| Other fields |  | 29773 | 1191 (4%) |

There was large variability across subfields. Table 2 shows for each analysis the 3 subfields with the highest proportions of funded top-cited scientists and the 3 subfields with the lowest proportions of funded top-cited scientists For example, when career-long impact was considered, almost all top-cited scientists had received such funding at some point in 1996-2022 in the fields of Developmental Biology (a classification subfield in Science-Metrix that includes most -omics) (89%), Substance Abuse (87%) and Immunology (85%), while low funding proportions were seen in several fields, with the lowest being in General Psychology and Cognitive Sciences (12%), Psychoanalysis (8%),and Legal and forensic medicine (3%). For recent funding, the highest proportions were seen in Developmental Biology, Bioinformatics and Geriatrics (40-42%) and the same three subfields toped proportions funded for current funding (29-31%). Conversely, Psychoanalysis, Legal & Forensic Medicine, General Psychology & Cognitive Sciences subfields that are highly related to biomedicine had less than 5% of their top-cited scientists funded recently and Dentistry, Veterinary Sciences, Social Psychology, Anatomy & Morphology, General Clinical Medicine, Human Factors, Psychoanalysis, Legal & Forensic Medicine, General Psychology & Cognitive Sciences, Gender Studies subfields that are highly related to biomedicine had less than 5% of their top-cited scientists currently funded. When single recent year impact was considered, the highest proportions of funding were dominated by Geriatrics, Gerontology, Substance Abuse, Developmental Biology, and (for current funding) Medical Informatics, while several subfields that are highly related to biomedicine had negligible proportions of their top-cited scientists being funded. Anatomy and Morphology, Gender studies, Legal and Forensic Medicine, General Psychology and Cognitive Sciences and Human Factors fared the worst (Table 2).

**Table 2.** Subfields with the highest proportions of funded top-cited scientists and subfields with the lowest proportions of funded top-cited scientists among the 69 subfields pre-specified as highly related to biomedicine Funding time is defined as ‘any’: any grant entry in RePORTER in 1996-2022; ‘recent’: any entry in the RePORTER that covers any year in the period 2015 until 2022; ‘current’: any entry in the RePORTER that covers 2021 and/or 2022. Highly related fields (according to the Science-Metrix classification) are 69 subfields, i.e. the 60 subfields within the larger fields of Biomedical Research, Clinical Medicine, Public Health and Health Services, and Psychology and Cognitive Sciences as well as 9 prespecified subfields from other large fields, i.e. Applied Ethics, Bioinformatics, Biomedical Engineering, Biotechnology, Demography, Gender Studies, Medical Informatics, Medicinal and Biomolecular Chemistry, and Veterinary Sciences.

| Analysis | Three subfields with highest proportions of funded top-cited scientists | Three subfields with lowest proportions of funded top-cited scientists |
| --- | --- | --- |
| **Career-long impact, Any funding** | Developmental Biology 89% (1566/1769) | General Psychology & Cognitive Sciences 12% (8/65) |
|  | Substance Abuse 87% (240/277) | Psychoanalysis 8% (4/50) |
|  | Immunology 85% (1247/1474) | Legal & Forensic Medicine 3% (2/66) |
| **Career-long impact, Recent funding** | Developmental Biology 42% (744/1769) | Psychoanalysis 2% (1/50) |
|  | Bioinformatics 41% (81/196) | Legal & Forensic Medicine 1.5% (1/66) |
|  | Geriatrics 40% (39/97) | General Psychology & Cognitive Sciences 0% (0/65) |
| **Career-long impact, Current funding** | Geriatrics 31% (30/97) | Legal & Forensic Medicine 0% (0/66) |
|  | Bioinformatics 30% (58/196) | General Psychology & Cognitive Sciences 0% (0/65) |
|  | Developmental Biology 29% (510/1769) | Gender Studies 0% (0/13) |
| **Recent year impact, Any funding** | Geriatrics 88% (66/75) | Anatomy & Morphology 14% (8/56) |
|  | Gerontology 87% (97/112) | Gender Studies 13% (2/15) |
|  | Substance Abuse 86% (184/214) | Legal & Forensic Medicine 5.7% (3/53) |
| **Recent year impact, Recent funding** | Geriatrics 55% (41/75) | General Psychology & Cognitive Sciences 4.7% (3/64) |
|  | Substance Abuse 50% (106/214) | Legal & Forensic Medicine 3.8% (2/53) |
|  | Developmental Biology 49% (799/1621) | Anatomy & Morphology 3.6% (2/56) |
| **Recent year impact, Current funding** | Geriatrics 43% (32/75) | General Psychology & Cognitive Sciences 3.1% (2/64) |
|  | Developmental Biology 36% (590/1621) | Human Factors 2.3% (4/177) |
|  | Medical Informatics 34% (41/119) | Gender Studies 0% (0/15) |

Among the 105 other subfields that we had considered a priori as not highly related to biomedicine, most had very low or even zero proportions of their top-cited scientists funded. However, exceptions existed, in particular for Family Studies, Organic Chemistry, Analytical Chemistry, Statistics and Probability, and Nanoscience and Nanotechnology (Supplementary Tables).

### Difference is citation metrics between funded and not funded top-cited scientists

Funded scientists had higher citation counts and composite citation indices than not funded scientists (Figure 1, Table 3). The difference in median career-long citations was more substantial when any funding was considered (+4222 citations for the 69 highly related fields) and it was smaller when recent funding was considered (+3068 citations) and even smaller when current funding was considered (+2833 citations). Most of the 69 subfields showed higher citations in funded versus not funded scientists, but exceptions occurred (Supplementary Tables).

**Table 3.** Comparison of citation counts and of composite citation index metrics in funded and not funded scientists in scientific subfields highly related to biomedicine Funding time is defined as ‘any’: any grant entry in RePORTER in 1996-2022; ‘recent’: any entry in the RePORTER that covers any year in the period 2015 until 2022; ‘current’: any entry in the RePORTER that covers 2021 and/or 2022. Highly related fields (according to the Science-Metrix classification) are 69 subfields, i.e. the 60 subfields within the larger fields of Biomedical Research, Clinical Medicine, Public Health and Health Services, and Psychology and Cognitive Sciences as well as 9 prespecified subfields from other large fields, i.e. Applied Ethics, Bioinformatics, Biomedical Engineering, Biotechnology, Demography, Gender Studies, Medical Informatics, Medicinal and Biomolecular Chemistry, and Veterinary Sciences.

| Top-cited US-based researchers | Funding time | Citations for funded, median | Citations for non-funded, median | p-value | Composite index for funded, median | Composite index for non-funded, median | p-value |
| --- | --- | --- | --- | --- | --- | --- | --- |
| Career-long impact | Any funding | 9594 | 5352 | <0.001 | 3.69 | 3.51 | <0.001 |
| Career-long impact | Recent funding | 10258 | 7190 | <0.001 | 3.69 | 3.6 | <0.001 |
| Career-long impact | Current funding | 10312 | 7479 | <0.001 | 3.68 | 3.61 | <0.001 |
| Recent year impact | Any funding | 1265 | 820 | <0.001 | 3.01 | 2.85 | <0.001 |
| Recent year impact | Recent funding | 1346 | 985 | <0.001 | 3 | 2.93 | <0.001 |
| Recent year impact | Current funding | 1376 | 1026 | <0.001 | 3 | 2.94 | <0.001 |

**Figure 1.**
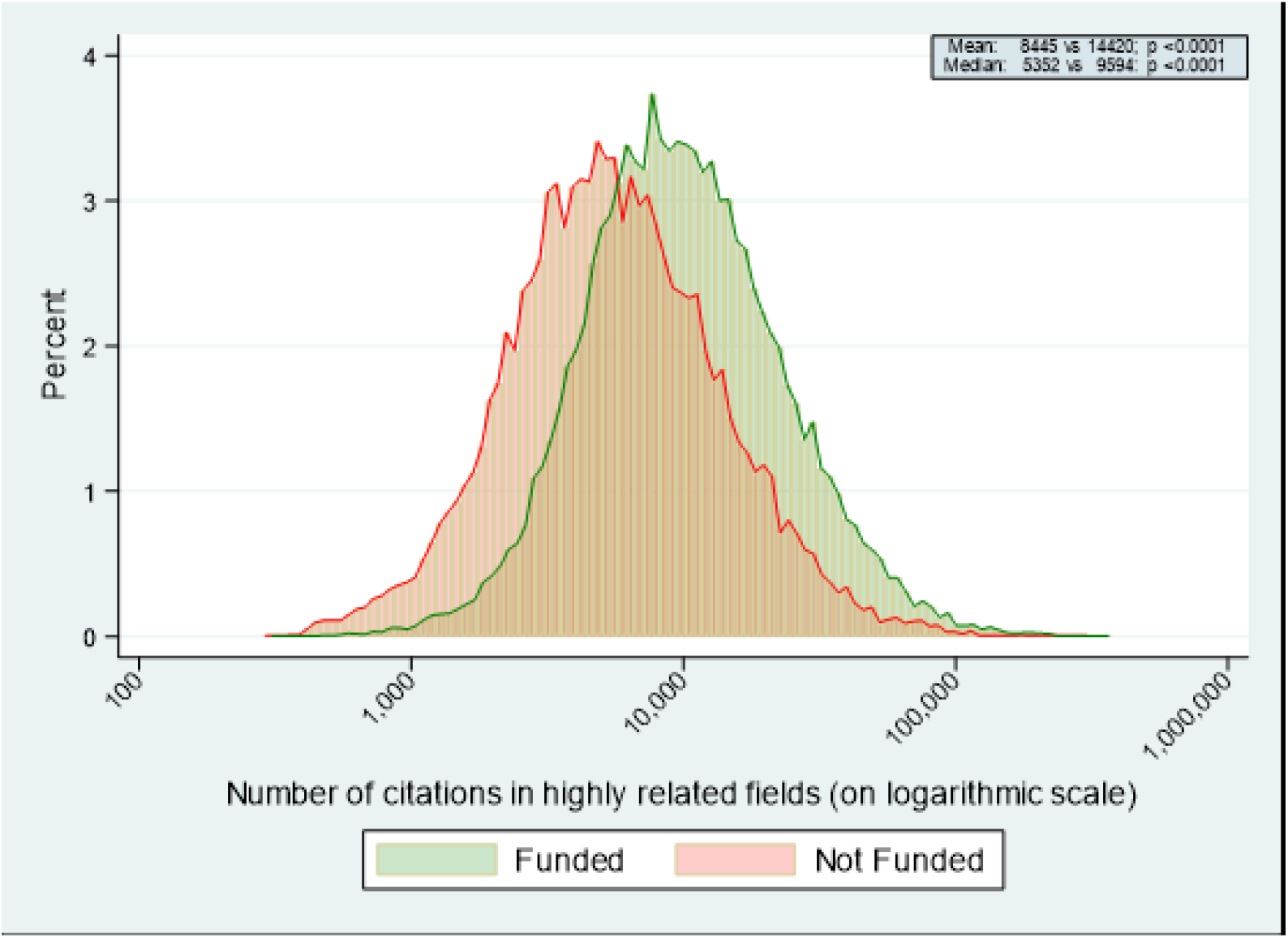
Distribution of citations (career-long) for top-cited scientists in biomedical subfields of science according to whether they have received or not federal biomedical funding.

After adjusting for subfield and the publication age of the scientist, the funded scientists still had higher citation metrics than the not funded ones. The differences tended to become smaller, e.g. +2069 citations in career-impact for those with versus those without current funding and similar pictures emerged also for subfield-specific analyses (Supplementary Tables).

## DISCUSSION

Biomedical federal funding has offered support as principal investigators to about two-thirds of the top-cited biomedical scientists at some point during the last quarter century. However, the rates of support are smaller, when only recent funding is considered and only a small minority of top-cited scientists have current federal biomedical funding as principal investigators. There is very large variability across scientific subfields, with some subfields having had almost ubiquitous funding of their top-cited scientists at least at some point during the last quarter century, and other subfields having negligible funding coverage for their top-cited scientists from biomedical federal agencies. Funded top-cited scientists attract substantially more citations than not funded ones, after adjusting for scientific subfield. Citation differences persist also when further adjustment is made for publication age of scientists.

These data highlight the great importance of federal funding for the most impactful researchers in the bibliometrics of the scientific literature. However, they also demonstrate that the large majority of top-cited biomedical scientists do not have current federal funding from biomedical federal agencies as principal investigators. This would suggest the need for further strengthening of the federal research budget. Investment in biomedical science is vital. Economic and improved health outcomes such as longer life expectancy are well documented benefits of research investment(12-14). There is concern that many capable scientists do not receive federal funding or receive federal funding for the first time very late in their career – prompting suggestions for funding reform(15-17). Moreover, there is great unevenness across subfields and it is unclear whether this unevenness is justified. It is possible that some subfields are more able to attract funding from other federal (e.g. National Science Foundation) and non-federal sources, e.g. the industry, foundations, institutions, philanthropy, etc. However, it is likely that there is genuine imbalance in funding that has been driven by the funding agency leadership choices, the ability of some subfields to make stronger claims for the support of their work, the existence (or perceived existence) of major opportunities for discoveries, and the fact that study section members tend to prefer funding work that they feel more familiar with.^5^ The extremely low rates of funding of some subfields focused on psychology, cognitive sciences, gender studies, legal and forensic medicine, and human factors is problematic, because these fields may also have difficulty to attract other funding (e.g. from industry). What may emerge may be a strengthening of some disciplines, while others (with crucial potential contributions) are abandoned. Fields that receive the lion’ s share of funding may not necessarily be the best investments and inertia may delay refocusing of research priorities.^15^ Scientists in underfunded fields often make pleas for more attention to their discipline,^16^ but fragmented efforts may not allow rational, comprehensive reform.

The observed association of higher citation indices with federal funding is congruent with these considerations. Correlations between citation indices and federal funding have also been previously observed in more limited analyses focused on specific specialties, e.g. radiology or neurosurgery(20-22). However, the direction of a causal relationship, if any, in this association in unclear. Bidirectional effects are plausible. More funding could lead to more productivity and citations. Concurrently more productivity and citations offer ammunition to claim and secure more funding. We should caution that we focused only on the top-cited scientists and there is no guarantee that less cited scientists show the same difference between funded and non-funded ones. However, top-cited scientists are the ones whose work is more widely used in the scientific literature, so they represent a critically influential sample. Of course, neither citations nor funding guarantee quality and reproducibility of the work.

Some limitations of our work should be discussed. First, matching of names across the two databases is imperfect. We performed in-depth evaluation of random samples of names for validation purposes. These validations suggest that matching errors are relatively small and the same applies for redundancy in the databases (duplicate entries of the same person are extremely uncommon^8^ in the citations database and relatively uncommon in RePORTER). If one were to adjust for the magnitude of the potential matching and duplication errors, the proportion funded would be similar to the crude estimates we have recorded or only slightly larger. Data for women may have the extra problem of changes in name upon change in marital status. Second, some scientists in the citations database are currently well beyond their prime (and very few may also have passed away), therefore they cannot be considered eligible for current or even recent funding, but the analysis of any funding at any time is not affected by this consideration. Third, recording of funding in RePORTER does not mean that the funding has covered a substantial portion of a researcher’ s investigational agenda and effort. Many federal grants offer very limited money and we did not try to analyze amounts of funding as this would be precarious to do across fields with very different cost requirements and over decades. Fourth, RePORTER catalogues the principal investigators of extramural grants (including multiple principal investigators). Some researchers may have received funding through other roles (co-investigators, subaward, intramural NIH, contractors) and not be retrievable in RePORTER. Fifth, some top-cited biomedical researchers may have received other federal funding (e.g. National Science Foundation or Department of Defense) not included in RePORTER. Sixth, the citations database does have some errors in accuracy and precision, but overall these errors have been documented to be small in previous validation exercises(8-10); we refer to previous work(8-10) on more details. While other scientists who perform important work may not be among those included in the top-cited lists, the included scientists clearly have large impact in the literature – even if the true deep scientific quality of this impact cannot be certain and is often even intangible long after scientific work is published.

Both funding success and citation metrics have major limitations as measures of impact, novelty, or good scientific work. However, they are widely used in academia and beyond and they can shape, extol, or ruin careers and shape science and allocation of precious resources. Our analysis offers insights about the likely bidirectional relationship of funding and citation impact and provides a map of US science in these two important dimensions. The findings that only a small minority of top-cited scientists have current federal biomedical funding calls for thoughtful inclusive redesign of the future funding agenda.

## Data Availability

Both the citation and funding databases are publicly available.

## SUPPLEMENTARY TEXT

### Manual validation of accuracy of the matching process

We randomly selected 100 scientists from the RePORTER database who were matched to the career-long citation impact database using our algorithm, and manually checked each of them for validity. Out of these 100 scientists, 85 were verified to be correct matches (same name, same institution or affiliated institutions), 3 had the same name but it was impossible to check institution match, because the RePORTER database has missing institution data), and 12 were wrongly matched (same name, but actually different scientists). Thus false matching rate was 12-15%.

We also randomly selected 100 scientists from the RePORTER database who were not matched to the career-long citation impact database using our algorithm. We manually checked each of them whether the citations database includes them with different spelling or different country. Out of 100 scientists, 4 were missed matching and for another 2 we could not be certain. Thus missed matching rate was 4-6%.

Finally, some scientists appear more than one time in the RePORTER database, because they may be entered with different spelling. This means that the number of scientists who are not matched may be lower, if many duplicates exist. To assess the magnitude of this problem, we also randomly selected 100 scientists from the RePORTER database who were not matched using our algorithm, and manually checked whether each of them appears as a duplicate in the RePORTER database: Out of these 100 scientists, 94 were uniquely identified; 1 was a possible duplicate (RePORTER had another entry with same initials, and ambiguous institution data, so one cannot confirm uniqueness), and 5 were clearly repeated entries (same scientist, different spelling). Thus duplicate rate was 5-6%.

