## Supplementary Tables for "Federal Funding and Citation Metrics of Biomedical Research in the USA"

**Supplementary Table 1.1: Percentage funded by field for Career: Any funding time**

| **Top-cited US-based researchers: Subfield** | **Funding time** | **Total** | **Funded (%)** |
| --- | --- | --- | --- |
| **Career: Developmental Biology** | **Any** | 1769 | 1566( 89%) |
| **Career: Substance Abuse** | **Any** | 277 | 240( 87%) |
| **Career: Immunology** | **Any** | 1474 | 1247( 85%) |
| **Career: Geriatrics** | **Any** | 97 | 82( 85%) |
| **Career: Biochemistry & Molecular Biology** | **Any** | 2173 | 1756( 81%) |
| **Career: Endocrinology & Metabolism** | **Any** | 993 | 800( 81%) |
| **Career: Gerontology** | **Any** | 126 | 101( 80%) |
| **Career: Virology** | **Any** | 726 | 578( 80%) |
| **Career: Neurology & Neurosurgery** | **Any** | 3337 | 2635( 79%) |
| **Career: Genetics & Heredity** | **Any** | 353 | 278( 79%) |
| **Career: Bioinformatics** | **Any** | 196 | 150( 77%) |
| **Career: Psychiatry** | **Any** | 927 | 707( 76%) |
| **Career: Oncology & Carcinogenesis** | **Any** | 2894 | 2144( 74%) |
| **Career: Public Health** | **Any** | 692 | 507( 73%) |
| **Career: Medical Informatics** | **Any** | 126 | 90( 71%) |
| **Career: Demography** | **Any** | 42 | 30( 71%) |
| **Career: Physiology** | **Any** | 247 | 176( 71%) |
| **Career: Epidemiology** | **Any** | 156 | 111( 71%) |
| **Career: Biomedical Engineering** | **Any** | 599 | 425( 71%) |
| **Career: Developmental & Child Psychology** | **Any** | 459 | 323( 70%) |
| **Career: Arthritis & Rheumatology** | **Any** | 229 | 160( 70%) |
| **Career: Allergy** | **Any** | 142 | 98( 69%) |
| **Career: Biophysics** | **Any** | 186 | 128( 69%) |
| **Career: Respiratory System** | **Any** | 591 | 389( 66%) |
| **Career: Experimental Psychology** | **Any** | 633 | 412( 65%) |
| **Career: Gastroenterology & Hepatology** | **Any** | 700 | 454( 65%) |
| **Career: Cardiovascular System & Hematology** | **Any** | 1876 | 1207( 64%) |
| **Career: Health Policy & Services** | **Any** | 214 | 135( 63%) |
| **Career: Urology & Nephrology** | **Any** | 814 | 510( 63%) |
| **Career: Speech-Language Pathology & Audiology** | **Any** | 111 | 68( 61%) |
| **Career: Clinical Psychology** | **Any** | 215 | 129( 60%) |
| **Career: Pediatrics** | **Any** | 752 | 448( 60%) |
| **Career: Microbiology** | **Any** | 1471 | 876( 60%) |
| **Career: Family Studies** | **Any** | 56 | 33( 59%) |
| **Career: Applied Ethics** | **Any** | 71 | 41( 58%) |
| **Career: Nutrition & Dietetics** | **Any** | 367 | 211( 57%) |
| **Career: Ophthalmology & Optometry** | **Any** | 771 | 443( 57%) |
| **Career: Organic Chemistry** | **Any** | 725 | 413( 57%) |
| **Career: Environmental & Occupational Health** | **Any** | 123 | 70( 57%) |
| **Career: Toxicology** | **Any** | 541 | 301( 56%) |
| **Career: Nursing** | **Any** | 919 | 502( 55%) |
| **Career: Obstetrics & Reproductive Medicine** | **Any** | 734 | 400( 54%) |
| **Career: Emergency & Critical Care Medicine** | **Any** | 339 | 181( 53%) |
| **Career: Rehabilitation** | **Any** | 230 | 122( 53%) |
| **Career: Pharmacology & Pharmacy** | **Any** | 833 | 425( 51%) |
| **Career: Behavioral Science & Comparative Psychology** | **Any** | 207 | 103( 50%) |
| **Career: Analytical Chemistry** | **Any** | 527 | 256( 49%) |
| **Career: Nuclear Medicine & Medical Imaging** | **Any** | 1184 | 562( 47%) |
| **Career: Otorhinolaryngology** | **Any** | 522 | 232( 44%) |
| **Career: Dentistry** | **Any** | 564 | 250( 44%) |
| **Career: Tropical Medicine** | **Any** | 217 | 95( 44%) |
| **Career: Mycology & Parasitology** | **Any** | 146 | 63( 43%) |
| **Career: Statistics & Probability** | **Any** | 249 | 107( 43%) |
| **Career: Medicinal & Biomolecular Chemistry** | **Any** | 652 | 280( 43%) |
| **Career: Anesthesiology** | **Any** | 435 | 186( 43%) |
| **Career: Dermatology & Venereal Diseases** | **Any** | 381 | 160( 42%) |
| **Career: Biotechnology** | **Any** | 270 | 110( 41%) |
| **Career: Social Psychology** | **Any** | 482 | 196( 41%) |
| **Career: Surgery** | **Any** | 1261 | 504( 40%) |
| **Career: General & Internal Medicine** | **Any** | 1574 | 624( 40%) |
| **Career: Complementary & Alternative Medicine** | **Any** | 81 | 32( 40%) |
| **Career: Sport Sciences** | **Any** | 186 | 68( 37%) |
| **Career: Acoustics** | **Any** | 310 | 113( 36%) |
| **Career: History of Social Sciences** | **Any** | 23 | 8( 35%) |
| **Career: Nanoscience & Nanotechnology** | **Any** | 488 | 169( 35%) |
| **Career: General Chemistry** | **Any** | 490 | 167( 34%) |
| **Career: Microscopy** | **Any** | 30 | 10( 33%) |
| **Career: Criminology** | **Any** | 153 | 47( 31%) |
| **Career: Sociology** | **Any** | 157 | 48( 31%) |
| **Career: Optics** | **Any** | 484 | 146( 30%) |
| **Career: Pathology** | **Any** | 293 | 87( 30%) |
| **Career: Social Work** | **Any** | 88 | 26( 30%) |
| **Career: Evolutionary Biology** | **Any** | 398 | 117( 29%) |
| **Career: Orthopedics** | **Any** | 777 | 214( 28%) |
| **Career: Social Sciences Methods** | **Any** | 77 | 21( 27%) |
| **Career: Chemical Physics** | **Any** | 890 | 238( 27%) |
| **Career: General Clinical Medicine** | **Any** | 128 | 31( 24%) |
| **Career: Plant Biology & Botany** | **Any** | 812 | 193( 24%) |
| **Career: Inorganic & Nuclear Chemistry** | **Any** | 318 | 75( 24%) |
| **Career: Veterinary Sciences** | **Any** | 558 | 131( 23%) |
| **Career: Environmental Sciences** | **Any** | 422 | 88( 21%) |
| **Career: Human Factors** | **Any** | 166 | 31( 19%) |
| **Career: Ornithology** | **Any** | 33 | 6( 18%) |
| **Career: Urban & Regional Planning** | **Any** | 61 | 11( 18%) |
| **Career: Economics** | **Any** | 453 | 81( 18%) |
| **Career: Artificial Intelligence & Image Processing** | **Any** | 1439 | 249( 17%) |
| **Career: Polymers** | **Any** | 531 | 89( 17%) |
| **Career: Economic Theory** | **Any** | 18 | 3( 17%) |
| **Career: Development Studies** | **Any** | 24 | 4( 17%) |
| **Career: Anatomy & Morphology** | **Any** | 54 | 9( 17%) |
| **Career: Distributed Computing** | **Any** | 139 | 23( 17%) |
| **Career: Entomology** | **Any** | 294 | 46( 16%) |
| **Career: Gender Studies** | **Any** | 13 | 2( 15%) |
| **Career: History of Science, Technology & Medicine** | **Any** | 20 | 3( 15%) |
| **Career: Software Engineering** | **Any** | 228 | 34( 15%) |
| **Career: Education** | **Any** | 732 | 108( 15%) |
| **Career: Food Science** | **Any** | 268 | 39( 15%) |
| **Career: Industrial Engineering & Automation** | **Any** | 628 | 90( 14%) |
| **Career: Logistics & Transportation** | **Any** | 183 | 26( 14%) |
| **Career: Geography** | **Any** | 92 | 13( 14%) |
| **Career: Sport, Leisure & Tourism** | **Any** | 46 | 6( 13%) |
| **Career: Numerical & Computational Mathematics** | **Any** | 120 | 15( 13%) |
| **Career: Communication & Media Studies** | **Any** | 154 | 19( 12%) |
| **Career: General Psychology & Cognitive Sciences** | **Any** | 65 | 8( 12%) |
| **Career: Fluids & Plasmas** | **Any** | 372 | 45( 12%) |
| **Career: Information & Library Sciences** | **Any** | 117 | 14( 12%) |
| **Career: Dairy & Animal Science** | **Any** | 454 | 54( 12%) |
| **Career: Optoelectronics & Photonics** | **Any** | 933 | 108( 12%) |
| **Career: Anthropology** | **Any** | 123 | 14( 11%) |
| **Career: Religions & Theology** | **Any** | 90 | 10( 11%) |
| **Career: Drama & Theater** | **Any** | 9 | 1( 11%) |
| **Career: Design Practice & Management** | **Any** | 100 | 11( 11%) |
| **Career: Environmental Engineering** | **Any** | 410 | 45( 11%) |
| **Career: Chemical Engineering** | **Any** | 330 | 36( 11%) |
| **Career: Applied Mathematics** | **Any** | 138 | 15( 11%) |
| **Career: Marketing** | **Any** | 129 | 14( 11%) |
| **Career: Zoology** | **Any** | 99 | 10( 10%) |
| **Career: Architecture** | **Any** | 10 | 1( 10%) |
| **Career: Ecology** | **Any** | 743 | 72( 9.7%) |
| **Career: Marine Biology & Hydrobiology** | **Any** | 331 | 32( 9.7%) |
| **Career: Networking & Telecommunications** | **Any** | 1476 | 140( 9.5%) |
| **Career: Computer Hardware & Architecture** | **Any** | 227 | 21( 9.3%) |
| **Career: Electrical & Electronic Engineering** | **Any** | 501 | 46( 9.2%) |
| **Career: Geological & Geomatics Engineering** | **Any** | 357 | 32( 9%) |
| **Career: Information Systems** | **Any** | 209 | 18( 8.6%) |
| **Career: Law** | **Any** | 106 | 9( 8.5%) |
| **Career: Mathematical Physics** | **Any** | 24 | 2( 8.3%) |
| **Career: Strategic, Defence & Security Studies** | **Any** | 182 | 15( 8.2%) |
| **Career: Agricultural Economics & Policy** | **Any** | 73 | 6( 8.2%) |
| **Career: General Physics** | **Any** | 488 | 40( 8.2%) |
| **Career: Languages & Linguistics** | **Any** | 110 | 9( 8.2%) |
| **Career: Mining & Metallurgy** | **Any** | 62 | 5( 8.1%) |
| **Career: Psychoanalysis** | **Any** | 50 | 4( 8%) |
| **Career: Political Science & Public Administration** | **Any** | 228 | 18( 7.9%) |
| **Career: Building & Construction** | **Any** | 128 | 10( 7.8%) |
| **Career: Oceanography** | **Any** | 185 | 14( 7.6%) |
| **Career: Literary Studies** | **Any** | 159 | 12( 7.5%) |
| **Career: Cultural Studies** | **Any** | 54 | 4( 7.4%) |
| **Career: Materials** | **Any** | 1363 | 99( 7.3%) |
| **Career: Applied Physics** | **Any** | 1918 | 138( 7.2%) |
| **Career: Econometrics** | **Any** | 42 | 3( 7.1%) |
| **Career: Mechanical Engineering & Transports** | **Any** | 706 | 50( 7.1%) |
| **Career: Operations Research** | **Any** | 205 | 14( 6.8%) |
| **Career: Finance** | **Any** | 132 | 9( 6.8%) |
| **Career: Business & Management** | **Any** | 526 | 34( 6.5%) |
| **Career: Meteorology & Atmospheric Sciences** | **Any** | 923 | 59( 6.4%) |
| **Career: General Mathematics** | **Any** | 502 | 32( 6.4%) |
| **Career: Computation Theory & Mathematics** | **Any** | 177 | 11( 6.2%) |
| **Career: Fisheries** | **Any** | 227 | 14( 6.2%) |
| **Career: Philosophy** | **Any** | 130 | 8( 6.2%) |
| **Career: Archaeology** | **Any** | 114 | 7( 6.1%) |
| **Career: Paleontology** | **Any** | 168 | 10( 6%) |
| **Career: Energy** | **Any** | 1212 | 72( 5.9%) |
| **Career: Agronomy & Agriculture** | **Any** | 456 | 27( 5.9%) |
| **Career: Aerospace & Aeronautics** | **Any** | 754 | 44( 5.8%) |
| **Career: Astronomy & Astrophysics** | **Any** | 789 | 44( 5.6%) |
| **Career: Classics** | **Any** | 18 | 1( 5.6%) |
| **Career: Geology** | **Any** | 92 | 5( 5.4%) |
| **Career: Forestry** | **Any** | 209 | 11( 5.3%) |
| **Career: Geochemistry & Geophysics** | **Any** | 843 | 41( 4.9%) |
| **Career: Nuclear & Particle Physics** | **Any** | 1030 | 48( 4.7%) |
| **Career: History** | **Any** | 111 | 5( 4.5%) |
| **Career: International Relations** | **Any** | 68 | 3( 4.4%) |
| **Career: Physical Chemistry** | **Any** | 114 | 5( 4.4%) |
| **Career: Science Studies** | **Any** | 26 | 1( 3.8%) |
| **Career: Civil Engineering** | **Any** | 275 | 10( 3.6%) |
| **Career: Legal & Forensic Medicine** | **Any** | 66 | 2( 3%) |
| **Career: Music** | **Any** | 42 | 1( 2.4%) |
| **Career: Accounting** | **Any** | 42 | 1( 2.4%) |
| **Career: Horticulture** | **Any** | 75 | 1( 1.3%) |
| **Career: Industrial Relations** | **Any** | 8 | 0( 0%) |
| **Career: Folklore** | **Any** | 6 | 0( 0%) |
| **Career: Automobile Design & Engineering** | **Any** | 7 | 0( 0%) |
| **Career: Art Practice, History & Theory** | **Any** | 32 | 0( 0%) |

**Supplementary Table 1.2: Percentage funded by field for Career: Recent funding time**

| **Top-cited US-based researchers: Subfield** | **Funding time** | **Total** | **Funded (%)** |
| --- | --- | --- | --- |
| **Career: Developmental Biology** | **Recent** | 1769 | 744( 42%) |
| **Career: Bioinformatics** | **Recent** | 196 | 81( 41%) |
| **Career: Geriatrics** | **Recent** | 97 | 39( 40%) |
| **Career: Substance Abuse** | **Recent** | 277 | 100( 36%) |
| **Career: Biomedical Engineering** | **Recent** | 599 | 205( 34%) |
| **Career: Medical Informatics** | **Recent** | 126 | 43( 34%) |
| **Career: Immunology** | **Recent** | 1474 | 502( 34%) |
| **Career: Epidemiology** | **Recent** | 156 | 53( 34%) |
| **Career: Virology** | **Recent** | 726 | 246( 34%) |
| **Career: Neurology & Neurosurgery** | **Recent** | 3337 | 1060( 32%) |
| **Career: Public Health** | **Recent** | 692 | 212( 31%) |
| **Career: Gerontology** | **Recent** | 126 | 38( 30%) |
| **Career: Oncology & Carcinogenesis** | **Recent** | 2894 | 861( 30%) |
| **Career: Genetics & Heredity** | **Recent** | 353 | 102( 29%) |
| **Career: Demography** | **Recent** | 42 | 11( 26%) |
| **Career: Health Policy & Services** | **Recent** | 214 | 56( 26%) |
| **Career: Developmental & Child Psychology** | **Recent** | 459 | 117( 25%) |
| **Career: Applied Ethics** | **Recent** | 71 | 18( 25%) |
| **Career: Gastroenterology & Hepatology** | **Recent** | 700 | 176( 25%) |
| **Career: Endocrinology & Metabolism** | **Recent** | 993 | 246( 25%) |
| **Career: Biophysics** | **Recent** | 186 | 46( 25%) |
| **Career: Psychiatry** | **Recent** | 927 | 229( 25%) |
| **Career: Arthritis & Rheumatology** | **Recent** | 229 | 55( 24%) |
| **Career: Emergency & Critical Care Medicine** | **Recent** | 339 | 81( 24%) |
| **Career: Microbiology** | **Recent** | 1471 | 343( 23%) |
| **Career: Allergy** | **Recent** | 142 | 33( 23%) |
| **Career: Respiratory System** | **Recent** | 591 | 136( 23%) |
| **Career: Clinical Psychology** | **Recent** | 215 | 49( 23%) |
| **Career: Cardiovascular System & Hematology** | **Recent** | 1876 | 423( 23%) |
| **Career: Biochemistry & Molecular Biology** | **Recent** | 2173 | 483( 22%) |
| **Career: Physiology** | **Recent** | 247 | 54( 22%) |
| **Career: Ophthalmology & Optometry** | **Recent** | 771 | 164( 21%) |
| **Career: Analytical Chemistry** | **Recent** | 527 | 112( 21%) |
| **Career: Urology & Nephrology** | **Recent** | 814 | 166( 20%) |
| **Career: Rehabilitation** | **Recent** | 230 | 46( 20%) |
| **Career: Microscopy** | **Recent** | 30 | 6( 20%) |
| **Career: Tropical Medicine** | **Recent** | 217 | 43( 20%) |
| **Career: Statistics & Probability** | **Recent** | 249 | 49( 20%) |
| **Career: Family Studies** | **Recent** | 56 | 11( 20%) |
| **Career: Toxicology** | **Recent** | 541 | 106( 20%) |
| **Career: Nutrition & Dietetics** | **Recent** | 367 | 71( 19%) |
| **Career: Medicinal & Biomolecular Chemistry** | **Recent** | 652 | 126( 19%) |
| **Career: Nuclear Medicine & Medical Imaging** | **Recent** | 1184 | 228( 19%) |
| **Career: Nanoscience & Nanotechnology** | **Recent** | 488 | 91( 19%) |
| **Career: Pediatrics** | **Recent** | 752 | 140( 19%) |
| **Career: Pharmacology & Pharmacy** | **Recent** | 833 | 151( 18%) |
| **Career: Optics** | **Recent** | 484 | 87( 18%) |
| **Career: Anesthesiology** | **Recent** | 435 | 78( 18%) |
| **Career: Obstetrics & Reproductive Medicine** | **Recent** | 734 | 130( 18%) |
| **Career: Experimental Psychology** | **Recent** | 633 | 111( 18%) |
| **Career: Speech-Language Pathology & Audiology** | **Recent** | 111 | 19( 17%) |
| **Career: Biotechnology** | **Recent** | 270 | 45( 17%) |
| **Career: Organic Chemistry** | **Recent** | 725 | 116( 16%) |
| **Career: Nursing** | **Recent** | 919 | 147( 16%) |
| **Career: Environmental & Occupational Health** | **Recent** | 123 | 19( 15%) |
| **Career: Behavioral Science & Comparative Psychology** | **Recent** | 207 | 27( 13%) |
| **Career: Mycology & Parasitology** | **Recent** | 146 | 19( 13%) |
| **Career: General & Internal Medicine** | **Recent** | 1574 | 204( 13%) |
| **Career: Otorhinolaryngology** | **Recent** | 522 | 67( 13%) |
| **Career: Sociology** | **Recent** | 157 | 20( 13%) |
| **Career: Surgery** | **Recent** | 1261 | 159( 13%) |
| **Career: Development Studies** | **Recent** | 24 | 3( 13%) |
| **Career: Drama & Theater** | **Recent** | 9 | 1( 11%) |
| **Career: General Chemistry** | **Recent** | 490 | 54( 11%) |
| **Career: Dermatology & Venereal Diseases** | **Recent** | 381 | 41( 11%) |
| **Career: Acoustics** | **Recent** | 310 | 33( 11%) |
| **Career: Criminology** | **Recent** | 153 | 16( 10%) |
| **Career: Evolutionary Biology** | **Recent** | 398 | 41( 10%) |
| **Career: Orthopedics** | **Recent** | 777 | 79( 10%) |
| **Career: Environmental Sciences** | **Recent** | 422 | 41( 9.7%) |
| **Career: Distributed Computing** | **Recent** | 139 | 13( 9.4%) |
| **Career: Social Work** | **Recent** | 88 | 8( 9.1%) |
| **Career: Social Sciences Methods** | **Recent** | 77 | 7( 9.1%) |
| **Career: Artificial Intelligence & Image Processing** | **Recent** | 1439 | 128( 8.9%) |
| **Career: Complementary & Alternative Medicine** | **Recent** | 81 | 7( 8.6%) |
| **Career: Dentistry** | **Recent** | 564 | 45( 8%) |
| **Career: Veterinary Sciences** | **Recent** | 558 | 44( 7.9%) |
| **Career: Pathology** | **Recent** | 293 | 23( 7.8%) |
| **Career: Plant Biology & Botany** | **Recent** | 812 | 63( 7.8%) |
| **Career: Gender Studies** | **Recent** | 13 | 1( 7.7%) |
| **Career: Social Psychology** | **Recent** | 482 | 37( 7.7%) |
| **Career: Geography** | **Recent** | 92 | 7( 7.6%) |
| **Career: Sport Sciences** | **Recent** | 186 | 14( 7.5%) |
| **Career: Industrial Engineering & Automation** | **Recent** | 628 | 46( 7.3%) |
| **Career: Chemical Physics** | **Recent** | 890 | 65( 7.3%) |
| **Career: Urban & Regional Planning** | **Recent** | 61 | 4( 6.6%) |
| **Career: Sport, Leisure & Tourism** | **Recent** | 46 | 3( 6.5%) |
| **Career: Entomology** | **Recent** | 294 | 18( 6.1%) |
| **Career: Human Factors** | **Recent** | 166 | 10( 6%) |
| **Career: Information & Library Sciences** | **Recent** | 117 | 7( 6%) |
| **Career: Numerical & Computational Mathematics** | **Recent** | 120 | 7( 5.8%) |
| **Career: Economics** | **Recent** | 453 | 26( 5.7%) |
| **Career: Economic Theory** | **Recent** | 18 | 1( 5.6%) |
| **Career: Classics** | **Recent** | 18 | 1( 5.6%) |
| **Career: Anatomy & Morphology** | **Recent** | 54 | 3( 5.6%) |
| **Career: General Clinical Medicine** | **Recent** | 128 | 7( 5.5%) |
| **Career: Logistics & Transportation** | **Recent** | 183 | 10( 5.5%) |
| **Career: Computer Hardware & Architecture** | **Recent** | 227 | 12( 5.3%) |
| **Career: Polymers** | **Recent** | 531 | 28( 5.3%) |
| **Career: Communication & Media Studies** | **Recent** | 154 | 8( 5.2%) |
| **Career: Fluids & Plasmas** | **Recent** | 372 | 19( 5.1%) |
| **Career: Software Engineering** | **Recent** | 228 | 11( 4.8%) |
| **Career: Electrical & Electronic Engineering** | **Recent** | 501 | 24( 4.8%) |
| **Career: Optoelectronics & Photonics** | **Recent** | 933 | 44( 4.7%) |
| **Career: Marketing** | **Recent** | 129 | 6( 4.7%) |
| **Career: Languages & Linguistics** | **Recent** | 110 | 5( 4.5%) |
| **Career: Education** | **Recent** | 732 | 33( 4.5%) |
| **Career: Networking & Telecommunications** | **Recent** | 1476 | 66( 4.5%) |
| **Career: Religions & Theology** | **Recent** | 90 | 4( 4.4%) |
| **Career: Archaeology** | **Recent** | 114 | 5( 4.4%) |
| **Career: History of Social Sciences** | **Recent** | 23 | 1( 4.3%) |
| **Career: Applied Mathematics** | **Recent** | 138 | 6( 4.3%) |
| **Career: Mathematical Physics** | **Recent** | 24 | 1( 4.2%) |
| **Career: Agricultural Economics & Policy** | **Recent** | 73 | 3( 4.1%) |
| **Career: Food Science** | **Recent** | 268 | 11( 4.1%) |
| **Career: Anthropology** | **Recent** | 123 | 5( 4.1%) |
| **Career: Science Studies** | **Recent** | 26 | 1( 3.8%) |
| **Career: Information Systems** | **Recent** | 209 | 8( 3.8%) |
| **Career: Finance** | **Recent** | 132 | 5( 3.8%) |
| **Career: Cultural Studies** | **Recent** | 54 | 2( 3.7%) |
| **Career: General Physics** | **Recent** | 488 | 18( 3.7%) |
| **Career: Chemical Engineering** | **Recent** | 330 | 12( 3.6%) |
| **Career: Mechanical Engineering & Transports** | **Recent** | 706 | 25( 3.5%) |
| **Career: Ecology** | **Recent** | 743 | 26( 3.5%) |
| **Career: Operations Research** | **Recent** | 205 | 7( 3.4%) |
| **Career: Dairy & Animal Science** | **Recent** | 454 | 15( 3.3%) |
| **Career: Mining & Metallurgy** | **Recent** | 62 | 2( 3.2%) |
| **Career: Building & Construction** | **Recent** | 128 | 4( 3.1%) |
| **Career: Philosophy** | **Recent** | 130 | 4( 3.1%) |
| **Career: Political Science & Public Administration** | **Recent** | 228 | 7( 3.1%) |
| **Career: Zoology** | **Recent** | 99 | 3( 3%) |
| **Career: Ornithology** | **Recent** | 33 | 1( 3%) |
| **Career: Design Practice & Management** | **Recent** | 100 | 3( 3%) |
| **Career: Materials** | **Recent** | 1363 | 40( 2.9%) |
| **Career: Environmental Engineering** | **Recent** | 410 | 12( 2.9%) |
| **Career: Inorganic & Nuclear Chemistry** | **Recent** | 318 | 9( 2.8%) |
| **Career: Geological & Geomatics Engineering** | **Recent** | 357 | 10( 2.8%) |
| **Career: Oceanography** | **Recent** | 185 | 5( 2.7%) |
| **Career: Literary Studies** | **Recent** | 159 | 4( 2.5%) |
| **Career: Meteorology & Atmospheric Sciences** | **Recent** | 923 | 23( 2.5%) |
| **Career: Applied Physics** | **Recent** | 1918 | 46( 2.4%) |
| **Career: Econometrics** | **Recent** | 42 | 1( 2.4%) |
| **Career: Business & Management** | **Recent** | 526 | 12( 2.3%) |
| **Career: Computation Theory & Mathematics** | **Recent** | 177 | 4( 2.3%) |
| **Career: Aerospace & Aeronautics** | **Recent** | 754 | 17( 2.3%) |
| **Career: Strategic, Defence & Security Studies** | **Recent** | 182 | 4( 2.2%) |
| **Career: General Mathematics** | **Recent** | 502 | 11( 2.2%) |
| **Career: Civil Engineering** | **Recent** | 275 | 6( 2.2%) |
| **Career: Geology** | **Recent** | 92 | 2( 2.2%) |
| **Career: Energy** | **Recent** | 1212 | 26( 2.1%) |
| **Career: Psychoanalysis** | **Recent** | 50 | 1( 2%) |
| **Career: Law** | **Recent** | 106 | 2( 1.9%) |
| **Career: Marine Biology & Hydrobiology** | **Recent** | 331 | 6( 1.8%) |
| **Career: Astronomy & Astrophysics** | **Recent** | 789 | 14( 1.8%) |
| **Career: Fisheries** | **Recent** | 227 | 4( 1.8%) |
| **Career: Legal & Forensic Medicine** | **Recent** | 66 | 1( 1.5%) |
| **Career: International Relations** | **Recent** | 68 | 1( 1.5%) |
| **Career: Nuclear & Particle Physics** | **Recent** | 1030 | 13( 1.3%) |
| **Career: Geochemistry & Geophysics** | **Recent** | 843 | 10( 1.2%) |
| **Career: Forestry** | **Recent** | 209 | 2( .96%) |
| **Career: Physical Chemistry** | **Recent** | 114 | 1( .88%) |
| **Career: Agronomy & Agriculture** | **Recent** | 456 | 4( .88%) |
| **Career: Paleontology** | **Recent** | 168 | 0( 0%) |
| **Career: Music** | **Recent** | 42 | 0( 0%) |
| **Career: Industrial Relations** | **Recent** | 8 | 0( 0%) |
| **Career: Horticulture** | **Recent** | 75 | 0( 0%) |
| **Career: History of Science, Technology & Medicine** | **Recent** | 20 | 0( 0%) |
| **Career: History** | **Recent** | 111 | 0( 0%) |
| **Career: General Psychology & Cognitive Sciences** | **Recent** | 65 | 0( 0%) |
| **Career: Folklore** | **Recent** | 6 | 0( 0%) |
| **Career: Automobile Design & Engineering** | **Recent** | 7 | 0( 0%) |
| **Career: Art Practice, History & Theory** | **Recent** | 32 | 0( 0%) |
| **Career: Architecture** | **Recent** | 10 | 0( 0%) |
| **Career: Accounting** | **Recent** | 42 | 0( 0%) |

**Supplementary Table 1.3: Percentage funded by field for Career: Current funding time**

| **Top-cited US-based researchers: Subfield** | **Funding time** | **Total** | **Funded (%)** |
| --- | --- | --- | --- |
| **Career: Geriatrics** | **Current** | 97 | 30( 31%) |
| **Career: Bioinformatics** | **Current** | 196 | 58( 30%) |
| **Career: Developmental Biology** | **Current** | 1769 | 510( 29%) |
| **Career: Substance Abuse** | **Current** | 277 | 64( 23%) |
| **Career: Medical Informatics** | **Current** | 126 | 29( 23%) |
| **Career: Virology** | **Current** | 726 | 159( 22%) |
| **Career: Biomedical Engineering** | **Current** | 599 | 131( 22%) |
| **Career: Neurology & Neurosurgery** | **Current** | 3337 | 692( 21%) |
| **Career: Immunology** | **Current** | 1474 | 300( 20%) |
| **Career: Gerontology** | **Current** | 126 | 25( 20%) |
| **Career: Applied Ethics** | **Current** | 71 | 14( 20%) |
| **Career: Public Health** | **Current** | 692 | 134( 19%) |
| **Career: Demography** | **Current** | 42 | 8( 19%) |
| **Career: Oncology & Carcinogenesis** | **Current** | 2894 | 517( 18%) |
| **Career: Gastroenterology & Hepatology** | **Current** | 700 | 116( 17%) |
| **Career: Allergy** | **Current** | 142 | 23( 16%) |
| **Career: Biophysics** | **Current** | 186 | 30( 16%) |
| **Career: Developmental & Child Psychology** | **Current** | 459 | 73( 16%) |
| **Career: Health Policy & Services** | **Current** | 214 | 34( 16%) |
| **Career: Physiology** | **Current** | 247 | 39( 16%) |
| **Career: Arthritis & Rheumatology** | **Current** | 229 | 36( 16%) |
| **Career: Epidemiology** | **Current** | 156 | 24( 15%) |
| **Career: Psychiatry** | **Current** | 927 | 142( 15%) |
| **Career: Genetics & Heredity** | **Current** | 353 | 54( 15%) |
| **Career: Emergency & Critical Care Medicine** | **Current** | 339 | 51( 15%) |
| **Career: Endocrinology & Metabolism** | **Current** | 993 | 148( 15%) |
| **Career: Statistics & Probability** | **Current** | 249 | 36( 14%) |
| **Career: Ophthalmology & Optometry** | **Current** | 771 | 110( 14%) |
| **Career: Microbiology** | **Current** | 1471 | 209( 14%) |
| **Career: Cardiovascular System & Hematology** | **Current** | 1876 | 260( 14%) |
| **Career: Respiratory System** | **Current** | 591 | 79( 13%) |
| **Career: Clinical Psychology** | **Current** | 215 | 28( 13%) |
| **Career: Nuclear Medicine & Medical Imaging** | **Current** | 1184 | 153( 13%) |
| **Career: Analytical Chemistry** | **Current** | 527 | 68( 13%) |
| **Career: Medicinal & Biomolecular Chemistry** | **Current** | 652 | 81( 12%) |
| **Career: Nanoscience & Nanotechnology** | **Current** | 488 | 60( 12%) |
| **Career: Urology & Nephrology** | **Current** | 814 | 100( 12%) |
| **Career: Optics** | **Current** | 484 | 58( 12%) |
| **Career: Biochemistry & Molecular Biology** | **Current** | 2173 | 250( 12%) |
| **Career: Nutrition & Dietetics** | **Current** | 367 | 42( 11%) |
| **Career: Experimental Psychology** | **Current** | 633 | 72( 11%) |
| **Career: Rehabilitation** | **Current** | 230 | 26( 11%) |
| **Career: Toxicology** | **Current** | 541 | 61( 11%) |
| **Career: Drama & Theater** | **Current** | 9 | 1( 11%) |
| **Career: Obstetrics & Reproductive Medicine** | **Current** | 734 | 80( 11%) |
| **Career: Pediatrics** | **Current** | 752 | 80( 11%) |
| **Career: Anesthesiology** | **Current** | 435 | 46( 11%) |
| **Career: Pharmacology & Pharmacy** | **Current** | 833 | 83( 10%) |
| **Career: Speech-Language Pathology & Audiology** | **Current** | 111 | 11( 9.9%) |
| **Career: Organic Chemistry** | **Current** | 725 | 71( 9.8%) |
| **Career: Environmental & Occupational Health** | **Current** | 123 | 12( 9.8%) |
| **Career: Family Studies** | **Current** | 56 | 5( 8.9%) |
| **Career: Development Studies** | **Current** | 24 | 2( 8.3%) |
| **Career: Mycology & Parasitology** | **Current** | 146 | 12( 8.2%) |
| **Career: Nursing** | **Current** | 919 | 73( 7.9%) |
| **Career: Surgery** | **Current** | 1261 | 99( 7.9%) |
| **Career: Tropical Medicine** | **Current** | 217 | 17( 7.8%) |
| **Career: General Chemistry** | **Current** | 490 | 38( 7.8%) |
| **Career: Complementary & Alternative Medicine** | **Current** | 81 | 6( 7.4%) |
| **Career: General & Internal Medicine** | **Current** | 1574 | 112( 7.1%) |
| **Career: Biotechnology** | **Current** | 270 | 19( 7%) |
| **Career: Microscopy** | **Current** | 30 | 2( 6.7%) |
| **Career: Acoustics** | **Current** | 310 | 20( 6.5%) |
| **Career: Sociology** | **Current** | 157 | 10( 6.4%) |
| **Career: Otorhinolaryngology** | **Current** | 522 | 33( 6.3%) |
| **Career: Behavioral Science & Comparative Psychology** | **Current** | 207 | 13( 6.3%) |
| **Career: Orthopedics** | **Current** | 777 | 46( 5.9%) |
| **Career: Sport Sciences** | **Current** | 186 | 11( 5.9%) |
| **Career: Social Work** | **Current** | 88 | 5( 5.7%) |
| **Career: Economic Theory** | **Current** | 18 | 1( 5.6%) |
| **Career: Evolutionary Biology** | **Current** | 398 | 22( 5.5%) |
| **Career: Dermatology & Venereal Diseases** | **Current** | 381 | 21( 5.5%) |
| **Career: Artificial Intelligence & Image Processing** | **Current** | 1439 | 79( 5.5%) |
| **Career: Environmental Sciences** | **Current** | 422 | 23( 5.5%) |
| **Career: Plant Biology & Botany** | **Current** | 812 | 42( 5.2%) |
| **Career: Pathology** | **Current** | 293 | 15( 5.1%) |
| **Career: Chemical Physics** | **Current** | 890 | 43( 4.8%) |
| **Career: Industrial Engineering & Automation** | **Current** | 628 | 30( 4.8%) |
| **Career: Entomology** | **Current** | 294 | 14( 4.8%) |
| **Career: Criminology** | **Current** | 153 | 7( 4.6%) |
| **Career: Religions & Theology** | **Current** | 90 | 4( 4.4%) |
| **Career: Sport, Leisure & Tourism** | **Current** | 46 | 2( 4.3%) |
| **Career: Distributed Computing** | **Current** | 139 | 6( 4.3%) |
| **Career: Dentistry** | **Current** | 564 | 24( 4.3%) |
| **Career: Social Sciences Methods** | **Current** | 77 | 3( 3.9%) |
| **Career: Science Studies** | **Current** | 26 | 1( 3.8%) |
| **Career: Logistics & Transportation** | **Current** | 183 | 7( 3.8%) |
| **Career: Veterinary Sciences** | **Current** | 558 | 21( 3.8%) |
| **Career: Social Psychology** | **Current** | 482 | 18( 3.7%) |
| **Career: Anatomy & Morphology** | **Current** | 54 | 2( 3.7%) |
| **Career: Applied Mathematics** | **Current** | 138 | 5( 3.6%) |
| **Career: Polymers** | **Current** | 531 | 19( 3.6%) |
| **Career: Information & Library Sciences** | **Current** | 117 | 4( 3.4%) |
| **Career: Communication & Media Studies** | **Current** | 154 | 5( 3.2%) |
| **Career: Mining & Metallurgy** | **Current** | 62 | 2( 3.2%) |
| **Career: General Clinical Medicine** | **Current** | 128 | 4( 3.1%) |
| **Career: Marketing** | **Current** | 129 | 4( 3.1%) |
| **Career: Economics** | **Current** | 453 | 14( 3.1%) |
| **Career: Software Engineering** | **Current** | 228 | 7( 3.1%) |
| **Career: Ornithology** | **Current** | 33 | 1( 3%) |
| **Career: Finance** | **Current** | 132 | 4( 3%) |
| **Career: Human Factors** | **Current** | 166 | 5( 3%) |
| **Career: Agricultural Economics & Policy** | **Current** | 73 | 2( 2.7%) |
| **Career: Computer Hardware & Architecture** | **Current** | 227 | 6( 2.6%) |
| **Career: Networking & Telecommunications** | **Current** | 1476 | 39( 2.6%) |
| **Career: Political Science & Public Administration** | **Current** | 228 | 6( 2.6%) |
| **Career: Food Science** | **Current** | 268 | 7( 2.6%) |
| **Career: Education** | **Current** | 732 | 19( 2.6%) |
| **Career: Dairy & Animal Science** | **Current** | 454 | 11( 2.4%) |
| **Career: Econometrics** | **Current** | 42 | 1( 2.4%) |
| **Career: Mechanical Engineering & Transports** | **Current** | 706 | 16( 2.3%) |
| **Career: Strategic, Defence & Security Studies** | **Current** | 182 | 4( 2.2%) |
| **Career: Electrical & Electronic Engineering** | **Current** | 501 | 11( 2.2%) |
| **Career: Geography** | **Current** | 92 | 2( 2.2%) |
| **Career: Chemical Engineering** | **Current** | 330 | 7( 2.1%) |
| **Career: General Physics** | **Current** | 488 | 10( 2%) |
| **Career: Optoelectronics & Photonics** | **Current** | 933 | 19( 2%) |
| **Career: Psychoanalysis** | **Current** | 50 | 1( 2%) |
| **Career: Design Practice & Management** | **Current** | 100 | 2( 2%) |
| **Career: Operations Research** | **Current** | 205 | 4( 2%) |
| **Career: Environmental Engineering** | **Current** | 410 | 8( 2%) |
| **Career: Information Systems** | **Current** | 209 | 4( 1.9%) |
| **Career: Literary Studies** | **Current** | 159 | 3( 1.9%) |
| **Career: Cultural Studies** | **Current** | 54 | 1( 1.9%) |
| **Career: Languages & Linguistics** | **Current** | 110 | 2( 1.8%) |
| **Career: Fisheries** | **Current** | 227 | 4( 1.8%) |
| **Career: Archaeology** | **Current** | 114 | 2( 1.8%) |
| **Career: Computation Theory & Mathematics** | **Current** | 177 | 3( 1.7%) |
| **Career: Numerical & Computational Mathematics** | **Current** | 120 | 2( 1.7%) |
| **Career: Urban & Regional Planning** | **Current** | 61 | 1( 1.6%) |
| **Career: Anthropology** | **Current** | 123 | 2( 1.6%) |
| **Career: Materials** | **Current** | 1363 | 22( 1.6%) |
| **Career: Fluids & Plasmas** | **Current** | 372 | 6( 1.6%) |
| **Career: General Mathematics** | **Current** | 502 | 8( 1.6%) |
| **Career: Building & Construction** | **Current** | 128 | 2( 1.6%) |
| **Career: Philosophy** | **Current** | 130 | 2( 1.5%) |
| **Career: Meteorology & Atmospheric Sciences** | **Current** | 923 | 14( 1.5%) |
| **Career: Aerospace & Aeronautics** | **Current** | 754 | 11( 1.5%) |
| **Career: Energy** | **Current** | 1212 | 17( 1.4%) |
| **Career: Geological & Geomatics Engineering** | **Current** | 357 | 5( 1.4%) |
| **Career: Ecology** | **Current** | 743 | 10( 1.3%) |
| **Career: Oceanography** | **Current** | 185 | 2( 1.1%) |
| **Career: Business & Management** | **Current** | 526 | 5( .95%) |
| **Career: Law** | **Current** | 106 | 1( .94%) |
| **Career: Applied Physics** | **Current** | 1918 | 18( .94%) |
| **Career: Marine Biology & Hydrobiology** | **Current** | 331 | 3( .91%) |
| **Career: Physical Chemistry** | **Current** | 114 | 1( .88%) |
| **Career: Nuclear & Particle Physics** | **Current** | 1030 | 8( .78%) |
| **Career: Astronomy & Astrophysics** | **Current** | 789 | 6( .76%) |
| **Career: Civil Engineering** | **Current** | 275 | 2( .73%) |
| **Career: Geochemistry & Geophysics** | **Current** | 843 | 6( .71%) |
| **Career: Agronomy & Agriculture** | **Current** | 456 | 3( .66%) |
| **Career: Inorganic & Nuclear Chemistry** | **Current** | 318 | 2( .63%) |
| **Career: Forestry** | **Current** | 209 | 1( .48%) |
| **Career: Zoology** | **Current** | 99 | 0( 0%) |
| **Career: Paleontology** | **Current** | 168 | 0( 0%) |
| **Career: Music** | **Current** | 42 | 0( 0%) |
| **Career: Mathematical Physics** | **Current** | 24 | 0( 0%) |
| **Career: Legal & Forensic Medicine** | **Current** | 66 | 0( 0%) |
| **Career: International Relations** | **Current** | 68 | 0( 0%) |
| **Career: Industrial Relations** | **Current** | 8 | 0( 0%) |
| **Career: Horticulture** | **Current** | 75 | 0( 0%) |
| **Career: History of Social Sciences** | **Current** | 23 | 0( 0%) |
| **Career: History of Science, Technology & Medicine** | **Current** | 20 | 0( 0%) |
| **Career: History** | **Current** | 111 | 0( 0%) |
| **Career: Geology** | **Current** | 92 | 0( 0%) |
| **Career: General Psychology & Cognitive Sciences** | **Current** | 65 | 0( 0%) |
| **Career: Gender Studies** | **Current** | 13 | 0( 0%) |
| **Career: Folklore** | **Current** | 6 | 0( 0%) |
| **Career: Classics** | **Current** | 18 | 0( 0%) |
| **Career: Automobile Design & Engineering** | **Current** | 7 | 0( 0%) |
| **Career: Art Practice, History & Theory** | **Current** | 32 | 0( 0%) |
| **Career: Architecture** | **Current** | 10 | 0( 0%) |
| **Career: Accounting** | **Current** | 42 | 0( 0%) |

**Supplementary Table 1.4: Percentage funded by field for Recent year: Any funding time**

| **Top-cited US-based researchers: Subfield** | **Funding time** | **Total** | **Funded (%)** |
| --- | --- | --- | --- |
| **Recent year: Geriatrics** | **Any** | 75 | 66( 88%) |
| **Recent year: Gerontology** | **Any** | 112 | 97( 87%) |
| **Recent year: Substance Abuse** | **Any** | 214 | 184( 86%) |
| **Recent year: Developmental Biology** | **Any** | 1621 | 1388( 86%) |
| **Recent year: Endocrinology & Metabolism** | **Any** | 729 | 607( 83%) |
| **Recent year: Immunology** | **Any** | 1329 | 1103( 83%) |
| **Recent year: Neurology & Neurosurgery** | **Any** | 2793 | 2283( 82%) |
| **Recent year: Biochemistry & Molecular Biology** | **Any** | 1718 | 1395( 81%) |
| **Recent year: Virology** | **Any** | 685 | 552( 81%) |
| **Recent year: Psychiatry** | **Any** | 757 | 610( 81%) |
| **Recent year: Genetics & Heredity** | **Any** | 335 | 263( 79%) |
| **Recent year: Allergy** | **Any** | 122 | 91( 75%) |
| **Recent year: Biophysics** | **Any** | 169 | 125( 74%) |
| **Recent year: Epidemiology** | **Any** | 129 | 95( 74%) |
| **Recent year: Biomedical Engineering** | **Any** | 526 | 387( 74%) |
| **Recent year: Arthritis & Rheumatology** | **Any** | 206 | 151( 73%) |
| **Recent year: Oncology & Carcinogenesis** | **Any** | 2746 | 1990( 72%) |
| **Recent year: Developmental & Child Psychology** | **Any** | 513 | 368( 72%) |
| **Recent year: Medical Informatics** | **Any** | 119 | 85( 71%) |
| **Recent year: Physiology** | **Any** | 228 | 162( 71%) |
| **Recent year: Public Health** | **Any** | 637 | 452( 71%) |
| **Recent year: Gastroenterology & Hepatology** | **Any** | 631 | 436( 69%) |
| **Recent year: Urology & Nephrology** | **Any** | 737 | 494( 67%) |
| **Recent year: Respiratory System** | **Any** | 546 | 365( 67%) |
| **Recent year: Pediatrics** | **Any** | 714 | 476( 67%) |
| **Recent year: Bioinformatics** | **Any** | 180 | 120( 67%) |
| **Recent year: Cardiovascular System & Hematology** | **Any** | 1694 | 1119( 66%) |
| **Recent year: Health Policy & Services** | **Any** | 191 | 123( 64%) |
| **Recent year: Experimental Psychology** | **Any** | 610 | 388( 64%) |
| **Recent year: Emergency & Critical Care Medicine** | **Any** | 261 | 165( 63%) |
| **Recent year: Clinical Psychology** | **Any** | 234 | 147( 63%) |
| **Recent year: Ophthalmology & Optometry** | **Any** | 695 | 432( 62%) |
| **Recent year: Family Studies** | **Any** | 55 | 34( 62%) |
| **Recent year: Speech-Language Pathology & Audiology** | **Any** | 114 | 70( 61%) |
| **Recent year: Rehabilitation** | **Any** | 199 | 122( 61%) |
| **Recent year: Obstetrics & Reproductive Medicine** | **Any** | 637 | 385( 60%) |
| **Recent year: Toxicology** | **Any** | 433 | 259( 60%) |
| **Recent year: Environmental & Occupational Health** | **Any** | 98 | 58( 59%) |
| **Recent year: Nutrition & Dietetics** | **Any** | 320 | 189( 59%) |
| **Recent year: Microbiology** | **Any** | 1319 | 779( 59%) |
| **Recent year: Nursing** | **Any** | 764 | 447( 59%) |
| **Recent year: Demography** | **Any** | 36 | 21( 58%) |
| **Recent year: Analytical Chemistry** | **Any** | 333 | 193( 58%) |
| **Recent year: Nuclear Medicine & Medical Imaging** | **Any** | 1007 | 576( 57%) |
| **Recent year: Organic Chemistry** | **Any** | 574 | 323( 56%) |
| **Recent year: Pharmacology & Pharmacy** | **Any** | 624 | 333( 53%) |
| **Recent year: Medicinal & Biomolecular Chemistry** | **Any** | 384 | 196( 51%) |
| **Recent year: Complementary & Alternative Medicine** | **Any** | 50 | 25( 50%) |
| **Recent year: Behavioral Science & Comparative Psychology** | **Any** | 127 | 62( 49%) |
| **Recent year: Anesthesiology** | **Any** | 403 | 195( 48%) |
| **Recent year: Dentistry** | **Any** | 441 | 210( 48%) |
| **Recent year: Otorhinolaryngology** | **Any** | 506 | 239( 47%) |
| **Recent year: Tropical Medicine** | **Any** | 213 | 99( 46%) |
| **Recent year: Applied Ethics** | **Any** | 54 | 25( 46%) |
| **Recent year: General & Internal Medicine** | **Any** | 1293 | 592( 46%) |
| **Recent year: Statistics & Probability** | **Any** | 270 | 121( 45%) |
| **Recent year: Surgery** | **Any** | 1125 | 499( 44%) |
| **Recent year: Mycology & Parasitology** | **Any** | 105 | 46( 44%) |
| **Recent year: Dermatology & Venereal Diseases** | **Any** | 341 | 144( 42%) |
| **Recent year: History of Social Sciences** | **Any** | 17 | 7( 41%) |
| **Recent year: Biotechnology** | **Any** | 185 | 76( 41%) |
| **Recent year: General Chemistry** | **Any** | 415 | 167( 40%) |
| **Recent year: Sport Sciences** | **Any** | 159 | 61( 38%) |
| **Recent year: Social Psychology** | **Any** | 562 | 214( 38%) |
| **Recent year: Acoustics** | **Any** | 233 | 86( 37%) |
| **Recent year: Optics** | **Any** | 437 | 138( 32%) |
| **Recent year: Orthopedics** | **Any** | 716 | 224( 31%) |
| **Recent year: Chemical Physics** | **Any** | 697 | 208( 30%) |
| **Recent year: Evolutionary Biology** | **Any** | 366 | 109( 30%) |
| **Recent year: Nanoscience & Nanotechnology** | **Any** | 595 | 173( 29%) |
| **Recent year: Sociology** | **Any** | 245 | 69( 28%) |
| **Recent year: Pathology** | **Any** | 299 | 84( 28%) |
| **Recent year: Plant Biology & Botany** | **Any** | 683 | 186( 27%) |
| **Recent year: General Clinical Medicine** | **Any** | 103 | 28( 27%) |
| **Recent year: Veterinary Sciences** | **Any** | 483 | 130( 27%) |
| **Recent year: Environmental Sciences** | **Any** | 234 | 62( 26%) |
| **Recent year: Social Sciences Methods** | **Any** | 100 | 25( 25%) |
| **Recent year: Criminology** | **Any** | 188 | 47( 25%) |
| **Recent year: Microscopy** | **Any** | 33 | 8( 24%) |
| **Recent year: Inorganic & Nuclear Chemistry** | **Any** | 205 | 47( 23%) |
| **Recent year: Social Work** | **Any** | 83 | 19( 23%) |
| **Recent year: Polymers** | **Any** | 375 | 79( 21%) |
| **Recent year: Drama & Theater** | **Any** | 10 | 2( 20%) |
| **Recent year: Economics** | **Any** | 586 | 112( 19%) |
| **Recent year: Food Science** | **Any** | 169 | 32( 19%) |
| **Recent year: Artificial Intelligence & Image Processing** | **Any** | 1230 | 228( 19%) |
| **Recent year: Distributed Computing** | **Any** | 103 | 19( 18%) |
| **Recent year: Industrial Engineering & Automation** | **Any** | 492 | 83( 17%) |
| **Recent year: Entomology** | **Any** | 259 | 43( 17%) |
| **Recent year: Design Practice & Management** | **Any** | 67 | 11( 16%) |
| **Recent year: Human Factors** | **Any** | 177 | 29( 16%) |
| **Recent year: Information & Library Sciences** | **Any** | 95 | 15( 16%) |
| **Recent year: Fluids & Plasmas** | **Any** | 325 | 51( 16%) |
| **Recent year: General Psychology & Cognitive Sciences** | **Any** | 64 | 10( 16%) |
| **Recent year: Urban & Regional Planning** | **Any** | 71 | 11( 15%) |
| **Recent year: Education** | **Any** | 699 | 108( 15%) |
| **Recent year: Psychoanalysis** | **Any** | 33 | 5( 15%) |
| **Recent year: Development Studies** | **Any** | 27 | 4( 15%) |
| **Recent year: Anatomy & Morphology** | **Any** | 56 | 8( 14%) |
| **Recent year: Geography** | **Any** | 120 | 17( 14%) |
| **Recent year: Ornithology** | **Any** | 29 | 4( 14%) |
| **Recent year: History of Science, Technology & Medicine** | **Any** | 15 | 2( 13%) |
| **Recent year: Gender Studies** | **Any** | 15 | 2( 13%) |
| **Recent year: Dairy & Animal Science** | **Any** | 336 | 43( 13%) |
| **Recent year: Logistics & Transportation** | **Any** | 151 | 19( 13%) |
| **Recent year: Economic Theory** | **Any** | 16 | 2( 13%) |
| **Recent year: Optoelectronics & Photonics** | **Any** | 697 | 87( 12%) |
| **Recent year: Software Engineering** | **Any** | 178 | 22( 12%) |
| **Recent year: Marine Biology & Hydrobiology** | **Any** | 287 | 35( 12%) |
| **Recent year: Anthropology** | **Any** | 116 | 14( 12%) |
| **Recent year: Numerical & Computational Mathematics** | **Any** | 101 | 12( 12%) |
| **Recent year: Geological & Geomatics Engineering** | **Any** | 282 | 31( 11%) |
| **Recent year: Electrical & Electronic Engineering** | **Any** | 364 | 40( 11%) |
| **Recent year: Physical Chemistry** | **Any** | 107 | 11( 10%) |
| **Recent year: Econometrics** | **Any** | 49 | 5( 10%) |
| **Recent year: Computer Hardware & Architecture** | **Any** | 226 | 23( 10%) |
| **Recent year: Communication & Media Studies** | **Any** | 187 | 19( 10%) |
| **Recent year: Environmental Engineering** | **Any** | 316 | 32( 10%) |
| **Recent year: Networking & Telecommunications** | **Any** | 1008 | 102( 10%) |
| **Recent year: Archaeology** | **Any** | 101 | 10( 9.9%) |
| **Recent year: Zoology** | **Any** | 92 | 9( 9.8%) |
| **Recent year: Religions & Theology** | **Any** | 82 | 8( 9.8%) |
| **Recent year: Chemical Engineering** | **Any** | 188 | 18( 9.6%) |
| **Recent year: Ecology** | **Any** | 811 | 76( 9.4%) |
| **Recent year: Materials** | **Any** | 980 | 91( 9.3%) |
| **Recent year: General Physics** | **Any** | 449 | 41( 9.1%) |
| **Recent year: Oceanography** | **Any** | 168 | 15( 8.9%) |
| **Recent year: Building & Construction** | **Any** | 102 | 9( 8.8%) |
| **Recent year: Sport, Leisure & Tourism** | **Any** | 104 | 9( 8.7%) |
| **Recent year: Applied Mathematics** | **Any** | 104 | 9( 8.7%) |
| **Recent year: Mechanical Engineering & Transports** | **Any** | 487 | 42( 8.6%) |
| **Recent year: Strategic, Defence & Security Studies** | **Any** | 130 | 11( 8.5%) |
| **Recent year: Languages & Linguistics** | **Any** | 99 | 8( 8.1%) |
| **Recent year: Computation Theory & Mathematics** | **Any** | 176 | 14( 8%) |
| **Recent year: Marketing** | **Any** | 254 | 20( 7.9%) |
| **Recent year: Law** | **Any** | 92 | 7( 7.6%) |
| **Recent year: Meteorology & Atmospheric Sciences** | **Any** | 1047 | 77( 7.4%) |
| **Recent year: Applied Physics** | **Any** | 1547 | 113( 7.3%) |
| **Recent year: Operations Research** | **Any** | 167 | 12( 7.2%) |
| **Recent year: Finance** | **Any** | 181 | 13( 7.2%) |
| **Recent year: Political Science & Public Administration** | **Any** | 345 | 24( 7%) |
| **Recent year: Forestry** | **Any** | 175 | 12( 6.9%) |
| **Recent year: Business & Management** | **Any** | 818 | 54( 6.6%) |
| **Recent year: Literary Studies** | **Any** | 138 | 9( 6.5%) |
| **Recent year: General Mathematics** | **Any** | 423 | 26( 6.1%) |
| **Recent year: Aerospace & Aeronautics** | **Any** | 638 | 39( 6.1%) |
| **Recent year: Geology** | **Any** | 82 | 5( 6.1%) |
| **Recent year: History** | **Any** | 101 | 6( 5.9%) |
| **Recent year: Information Systems** | **Any** | 169 | 10( 5.9%) |
| **Recent year: Energy** | **Any** | 850 | 50( 5.9%) |
| **Recent year: Science Studies** | **Any** | 35 | 2( 5.7%) |
| **Recent year: Legal & Forensic Medicine** | **Any** | 53 | 3( 5.7%) |
| **Recent year: Cultural Studies** | **Any** | 36 | 2( 5.6%) |
| **Recent year: Astronomy & Astrophysics** | **Any** | 638 | 35( 5.5%) |
| **Recent year: Accounting** | **Any** | 73 | 4( 5.5%) |
| **Recent year: Music** | **Any** | 37 | 2( 5.4%) |
| **Recent year: Geochemistry & Geophysics** | **Any** | 741 | 40( 5.4%) |
| **Recent year: Agronomy & Agriculture** | **Any** | 361 | 19( 5.3%) |
| **Recent year: Nuclear & Particle Physics** | **Any** | 945 | 46( 4.9%) |
| **Recent year: Fisheries** | **Any** | 187 | 9( 4.8%) |
| **Recent year: Classics** | **Any** | 21 | 1( 4.8%) |
| **Recent year: Mining & Metallurgy** | **Any** | 43 | 2( 4.7%) |
| **Recent year: Paleontology** | **Any** | 136 | 6( 4.4%) |
| **Recent year: Agricultural Economics & Policy** | **Any** | 69 | 3( 4.3%) |
| **Recent year: Philosophy** | **Any** | 127 | 5( 3.9%) |
| **Recent year: International Relations** | **Any** | 56 | 2( 3.6%) |
| **Recent year: Civil Engineering** | **Any** | 209 | 6( 2.9%) |
| **Recent year: Mathematical Physics** | **Any** | 21 | 0( 0%) |
| **Recent year: Industrial Relations** | **Any** | 7 | 0( 0%) |
| **Recent year: Horticulture** | **Any** | 60 | 0( 0%) |
| **Recent year: Folklore** | **Any** | 7 | 0( 0%) |
| **Recent year: Automobile Design & Engineering** | **Any** | 7 | 0( 0%) |
| **Recent year: Art Practice, History & Theory** | **Any** | 24 | 0( 0%) |
| **Recent year: Architecture** | **Any** | 10 | 0( 0%) |

**Supplementary Table 1.5: Percentage funded by field for Recent year: Recent funding time**

| **Top-cited US-based researchers: Subfield** | **Funding time** | **Total** | **Funded (%)** |
| --- | --- | --- | --- |
| **Recent year: Geriatrics** | **Recent** | 75 | 41( 55%) |
| **Recent year: Substance Abuse** | **Recent** | 214 | 106( 50%) |
| **Recent year: Developmental Biology** | **Recent** | 1621 | 799( 49%) |
| **Recent year: Medical Informatics** | **Recent** | 119 | 57( 48%) |
| **Recent year: Bioinformatics** | **Recent** | 180 | 83( 46%) |
| **Recent year: Virology** | **Recent** | 685 | 311( 45%) |
| **Recent year: Immunology** | **Recent** | 1329 | 592( 45%) |
| **Recent year: Gerontology** | **Recent** | 112 | 49( 44%) |
| **Recent year: Biomedical Engineering** | **Recent** | 526 | 230( 44%) |
| **Recent year: Neurology & Neurosurgery** | **Recent** | 2793 | 1160( 42%) |
| **Recent year: Oncology & Carcinogenesis** | **Recent** | 2746 | 1088( 40%) |
| **Recent year: Public Health** | **Recent** | 637 | 245( 38%) |
| **Recent year: Genetics & Heredity** | **Recent** | 335 | 128( 38%) |
| **Recent year: Emergency & Critical Care Medicine** | **Recent** | 261 | 99( 38%) |
| **Recent year: Epidemiology** | **Recent** | 129 | 47( 36%) |
| **Recent year: Psychiatry** | **Recent** | 757 | 269( 36%) |
| **Recent year: Gastroenterology & Hepatology** | **Recent** | 631 | 222( 35%) |
| **Recent year: Analytical Chemistry** | **Recent** | 333 | 117( 35%) |
| **Recent year: Arthritis & Rheumatology** | **Recent** | 206 | 70( 34%) |
| **Recent year: Allergy** | **Recent** | 122 | 41( 34%) |
| **Recent year: Developmental & Child Psychology** | **Recent** | 513 | 171( 33%) |
| **Recent year: Endocrinology & Metabolism** | **Recent** | 729 | 241( 33%) |
| **Recent year: Respiratory System** | **Recent** | 546 | 179( 33%) |
| **Recent year: Nuclear Medicine & Medical Imaging** | **Recent** | 1007 | 318( 32%) |
| **Recent year: Cardiovascular System & Hematology** | **Recent** | 1694 | 534( 32%) |
| **Recent year: Health Policy & Services** | **Recent** | 191 | 60( 31%) |
| **Recent year: Microbiology** | **Recent** | 1319 | 413( 31%) |
| **Recent year: Toxicology** | **Recent** | 433 | 135( 31%) |
| **Recent year: Pediatrics** | **Recent** | 714 | 221( 31%) |
| **Recent year: Biophysics** | **Recent** | 169 | 52( 31%) |
| **Recent year: Rehabilitation** | **Recent** | 199 | 60( 30%) |
| **Recent year: Urology & Nephrology** | **Recent** | 737 | 221( 30%) |
| **Recent year: Ophthalmology & Optometry** | **Recent** | 695 | 208( 30%) |
| **Recent year: Physiology** | **Recent** | 228 | 67( 29%) |
| **Recent year: Biochemistry & Molecular Biology** | **Recent** | 1718 | 503( 29%) |
| **Recent year: Clinical Psychology** | **Recent** | 234 | 66( 28%) |
| **Recent year: Medicinal & Biomolecular Chemistry** | **Recent** | 384 | 108( 28%) |
| **Recent year: Obstetrics & Reproductive Medicine** | **Recent** | 637 | 176( 28%) |
| **Recent year: Anesthesiology** | **Recent** | 403 | 110( 27%) |
| **Recent year: Tropical Medicine** | **Recent** | 213 | 57( 27%) |
| **Recent year: Pharmacology & Pharmacy** | **Recent** | 624 | 158( 25%) |
| **Recent year: Nutrition & Dietetics** | **Recent** | 320 | 80( 25%) |
| **Recent year: Nursing** | **Recent** | 764 | 191( 25%) |
| **Recent year: Demography** | **Recent** | 36 | 9( 25%) |
| **Recent year: Speech-Language Pathology & Audiology** | **Recent** | 114 | 28( 25%) |
| **Recent year: Statistics & Probability** | **Recent** | 270 | 65( 24%) |
| **Recent year: Family Studies** | **Recent** | 55 | 13( 24%) |
| **Recent year: Environmental & Occupational Health** | **Recent** | 98 | 23( 23%) |
| **Recent year: Experimental Psychology** | **Recent** | 610 | 137( 22%) |
| **Recent year: Applied Ethics** | **Recent** | 54 | 12( 22%) |
| **Recent year: Organic Chemistry** | **Recent** | 574 | 126( 22%) |
| **Recent year: Optics** | **Recent** | 437 | 95( 22%) |
| **Recent year: Microscopy** | **Recent** | 33 | 7( 21%) |
| **Recent year: General & Internal Medicine** | **Recent** | 1293 | 269( 21%) |
| **Recent year: Biotechnology** | **Recent** | 185 | 38( 21%) |
| **Recent year: Otorhinolaryngology** | **Recent** | 506 | 103( 20%) |
| **Recent year: Surgery** | **Recent** | 1125 | 227( 20%) |
| **Recent year: General Chemistry** | **Recent** | 415 | 77( 19%) |
| **Recent year: Mycology & Parasitology** | **Recent** | 105 | 19( 18%) |
| **Recent year: Nanoscience & Nanotechnology** | **Recent** | 595 | 106( 18%) |
| **Recent year: Behavioral Science & Comparative Psychology** | **Recent** | 127 | 22( 17%) |
| **Recent year: Dermatology & Venereal Diseases** | **Recent** | 341 | 55( 16%) |
| **Recent year: Acoustics** | **Recent** | 233 | 37( 16%) |
| **Recent year: Orthopedics** | **Recent** | 716 | 108( 15%) |
| **Recent year: Environmental Sciences** | **Recent** | 234 | 35( 15%) |
| **Recent year: Sociology** | **Recent** | 245 | 33( 13%) |
| **Recent year: Complementary & Alternative Medicine** | **Recent** | 50 | 6( 12%) |
| **Recent year: Dentistry** | **Recent** | 441 | 52( 12%) |
| **Recent year: Distributed Computing** | **Recent** | 103 | 12( 12%) |
| **Recent year: Pathology** | **Recent** | 299 | 34( 11%) |
| **Recent year: Evolutionary Biology** | **Recent** | 366 | 41( 11%) |
| **Recent year: Artificial Intelligence & Image Processing** | **Recent** | 1230 | 134( 11%) |
| **Recent year: Veterinary Sciences** | **Recent** | 483 | 51( 11%) |
| **Recent year: Plant Biology & Botany** | **Recent** | 683 | 71( 10%) |
| **Recent year: Chemical Physics** | **Recent** | 697 | 71( 10%) |
| **Recent year: Industrial Engineering & Automation** | **Recent** | 492 | 50( 10%) |
| **Recent year: Sport Sciences** | **Recent** | 159 | 16( 10%) |
| **Recent year: Drama & Theater** | **Recent** | 10 | 1( 10%) |
| **Recent year: Social Work** | **Recent** | 83 | 8( 9.6%) |
| **Recent year: Criminology** | **Recent** | 188 | 18( 9.6%) |
| **Recent year: Design Practice & Management** | **Recent** | 67 | 6( 9%) |
| **Recent year: Polymers** | **Recent** | 375 | 32( 8.5%) |
| **Recent year: General Clinical Medicine** | **Recent** | 103 | 8( 7.8%) |
| **Recent year: Geography** | **Recent** | 120 | 9( 7.5%) |
| **Recent year: Social Psychology** | **Recent** | 562 | 42( 7.5%) |
| **Recent year: Development Studies** | **Recent** | 27 | 2( 7.4%) |
| **Recent year: Economics** | **Recent** | 586 | 42( 7.2%) |
| **Recent year: Fluids & Plasmas** | **Recent** | 325 | 23( 7.1%) |
| **Recent year: Social Sciences Methods** | **Recent** | 100 | 7( 7%) |
| **Recent year: Entomology** | **Recent** | 259 | 18( 6.9%) |
| **Recent year: Numerical & Computational Mathematics** | **Recent** | 101 | 7( 6.9%) |
| **Recent year: Electrical & Electronic Engineering** | **Recent** | 364 | 25( 6.9%) |
| **Recent year: Gender Studies** | **Recent** | 15 | 1( 6.7%) |
| **Recent year: Computer Hardware & Architecture** | **Recent** | 226 | 15( 6.6%) |
| **Recent year: Logistics & Transportation** | **Recent** | 151 | 10( 6.6%) |
| **Recent year: Food Science** | **Recent** | 169 | 11( 6.5%) |
| **Recent year: Information & Library Sciences** | **Recent** | 95 | 6( 6.3%) |
| **Recent year: Psychoanalysis** | **Recent** | 33 | 2( 6.1%) |
| **Recent year: Education** | **Recent** | 699 | 42( 6%) |
| **Recent year: Archaeology** | **Recent** | 101 | 6( 5.9%) |
| **Recent year: Optoelectronics & Photonics** | **Recent** | 697 | 41( 5.9%) |
| **Recent year: History of Social Sciences** | **Recent** | 17 | 1( 5.9%) |
| **Recent year: Science Studies** | **Recent** | 35 | 2( 5.7%) |
| **Recent year: Human Factors** | **Recent** | 177 | 10( 5.6%) |
| **Recent year: Urban & Regional Planning** | **Recent** | 71 | 4( 5.6%) |
| **Recent year: Networking & Telecommunications** | **Recent** | 1008 | 52( 5.2%) |
| **Recent year: Dairy & Animal Science** | **Recent** | 336 | 17( 5.1%) |
| **Recent year: Languages & Linguistics** | **Recent** | 99 | 5( 5.1%) |
| **Recent year: Building & Construction** | **Recent** | 102 | 5( 4.9%) |
| **Recent year: Religions & Theology** | **Recent** | 82 | 4( 4.9%) |
| **Recent year: Inorganic & Nuclear Chemistry** | **Recent** | 205 | 10( 4.9%) |
| **Recent year: Applied Mathematics** | **Recent** | 104 | 5( 4.8%) |
| **Recent year: Mechanical Engineering & Transports** | **Recent** | 487 | 23( 4.7%) |
| **Recent year: General Psychology & Cognitive Sciences** | **Recent** | 64 | 3( 4.7%) |
| **Recent year: Mining & Metallurgy** | **Recent** | 43 | 2( 4.7%) |
| **Recent year: Software Engineering** | **Recent** | 178 | 8( 4.5%) |
| **Recent year: Anthropology** | **Recent** | 116 | 5( 4.3%) |
| **Recent year: Materials** | **Recent** | 980 | 42( 4.3%) |
| **Recent year: Chemical Engineering** | **Recent** | 188 | 8( 4.3%) |
| **Recent year: Accounting** | **Recent** | 73 | 3( 4.1%) |
| **Recent year: Econometrics** | **Recent** | 49 | 2( 4.1%) |
| **Recent year: Ecology** | **Recent** | 811 | 32( 3.9%) |
| **Recent year: Sport, Leisure & Tourism** | **Recent** | 104 | 4( 3.8%) |
| **Recent year: Legal & Forensic Medicine** | **Recent** | 53 | 2( 3.8%) |
| **Recent year: Communication & Media Studies** | **Recent** | 187 | 7( 3.7%) |
| **Recent year: Physical Chemistry** | **Recent** | 107 | 4( 3.7%) |
| **Recent year: Operations Research** | **Recent** | 167 | 6( 3.6%) |
| **Recent year: Anatomy & Morphology** | **Recent** | 56 | 2( 3.6%) |
| **Recent year: Information Systems** | **Recent** | 169 | 6( 3.6%) |
| **Recent year: Marketing** | **Recent** | 254 | 9( 3.5%) |
| **Recent year: Ornithology** | **Recent** | 29 | 1( 3.4%) |
| **Recent year: General Physics** | **Recent** | 449 | 15( 3.3%) |
| **Recent year: Finance** | **Recent** | 181 | 6( 3.3%) |
| **Recent year: Zoology** | **Recent** | 92 | 3( 3.3%) |
| **Recent year: Oceanography** | **Recent** | 168 | 5( 3%) |
| **Recent year: Meteorology & Atmospheric Sciences** | **Recent** | 1047 | 31( 3%) |
| **Recent year: Applied Physics** | **Recent** | 1547 | 44( 2.8%) |
| **Recent year: Geological & Geomatics Engineering** | **Recent** | 282 | 8( 2.8%) |
| **Recent year: Aerospace & Aeronautics** | **Recent** | 638 | 18( 2.8%) |
| **Recent year: Marine Biology & Hydrobiology** | **Recent** | 287 | 8( 2.8%) |
| **Recent year: Cultural Studies** | **Recent** | 36 | 1( 2.8%) |
| **Recent year: Energy** | **Recent** | 850 | 22( 2.6%) |
| **Recent year: Environmental Engineering** | **Recent** | 316 | 8( 2.5%) |
| **Recent year: Geology** | **Recent** | 82 | 2( 2.4%) |
| **Recent year: Business & Management** | **Recent** | 818 | 19( 2.3%) |
| **Recent year: Strategic, Defence & Security Studies** | **Recent** | 130 | 3( 2.3%) |
| **Recent year: Computation Theory & Mathematics** | **Recent** | 176 | 4( 2.3%) |
| **Recent year: Law** | **Recent** | 92 | 2( 2.2%) |
| **Recent year: Political Science & Public Administration** | **Recent** | 345 | 7( 2%) |
| **Recent year: History** | **Recent** | 101 | 2( 2%) |
| **Recent year: International Relations** | **Recent** | 56 | 1( 1.8%) |
| **Recent year: Astronomy & Astrophysics** | **Recent** | 638 | 11( 1.7%) |
| **Recent year: General Mathematics** | **Recent** | 423 | 7( 1.7%) |
| **Recent year: Fisheries** | **Recent** | 187 | 3( 1.6%) |
| **Recent year: Philosophy** | **Recent** | 127 | 2( 1.6%) |
| **Recent year: Literary Studies** | **Recent** | 138 | 2( 1.4%) |
| **Recent year: Agricultural Economics & Policy** | **Recent** | 69 | 1( 1.4%) |
| **Recent year: Civil Engineering** | **Recent** | 209 | 3( 1.4%) |
| **Recent year: Geochemistry & Geophysics** | **Recent** | 741 | 10( 1.3%) |
| **Recent year: Forestry** | **Recent** | 175 | 2( 1.1%) |
| **Recent year: Nuclear & Particle Physics** | **Recent** | 945 | 10( 1.1%) |
| **Recent year: Agronomy & Agriculture** | **Recent** | 361 | 3( .83%) |
| **Recent year: Paleontology** | **Recent** | 136 | 1( .74%) |
| **Recent year: Music** | **Recent** | 37 | 0( 0%) |
| **Recent year: Mathematical Physics** | **Recent** | 21 | 0( 0%) |
| **Recent year: Industrial Relations** | **Recent** | 7 | 0( 0%) |
| **Recent year: Horticulture** | **Recent** | 60 | 0( 0%) |
| **Recent year: History of Science, Technology & Medicine** | **Recent** | 15 | 0( 0%) |
| **Recent year: Folklore** | **Recent** | 7 | 0( 0%) |
| **Recent year: Economic Theory** | **Recent** | 16 | 0( 0%) |
| **Recent year: Classics** | **Recent** | 21 | 0( 0%) |
| **Recent year: Automobile Design & Engineering** | **Recent** | 7 | 0( 0%) |
| **Recent year: Art Practice, History & Theory** | **Recent** | 24 | 0( 0%) |
| **Recent year: Architecture** | **Recent** | 10 | 0( 0%) |

**Supplementary Table 1.6: Percentage funded by field for Recent year: Current funding time**

| **Top-cited US-based researchers: Subfield** | **Funding time** | **Total** | **Funded (%)** |
| --- | --- | --- | --- |
| **Recent year: Geriatrics** | **Current** | 75 | 32( 43%) |
| **Recent year: Developmental Biology** | **Current** | 1621 | 590( 36%) |
| **Recent year: Medical Informatics** | **Current** | 119 | 41( 34%) |
| **Recent year: Bioinformatics** | **Current** | 180 | 61( 34%) |
| **Recent year: Substance Abuse** | **Current** | 214 | 67( 31%) |
| **Recent year: Gerontology** | **Current** | 112 | 35( 31%) |
| **Recent year: Biomedical Engineering** | **Current** | 526 | 160( 30%) |
| **Recent year: Virology** | **Current** | 685 | 206( 30%) |
| **Recent year: Immunology** | **Current** | 1329 | 394( 30%) |
| **Recent year: Neurology & Neurosurgery** | **Current** | 2793 | 801( 29%) |
| **Recent year: Allergy** | **Current** | 122 | 33( 27%) |
| **Recent year: Emergency & Critical Care Medicine** | **Current** | 261 | 70( 27%) |
| **Recent year: Oncology & Carcinogenesis** | **Current** | 2746 | 725( 26%) |
| **Recent year: Public Health** | **Current** | 637 | 163( 26%) |
| **Recent year: Gastroenterology & Hepatology** | **Current** | 631 | 154( 24%) |
| **Recent year: Psychiatry** | **Current** | 757 | 177( 23%) |
| **Recent year: Arthritis & Rheumatology** | **Current** | 206 | 48( 23%) |
| **Recent year: Epidemiology** | **Current** | 129 | 29( 22%) |
| **Recent year: Genetics & Heredity** | **Current** | 335 | 75( 22%) |
| **Recent year: Analytical Chemistry** | **Current** | 333 | 74( 22%) |
| **Recent year: Ophthalmology & Optometry** | **Current** | 695 | 153( 22%) |
| **Recent year: Developmental & Child Psychology** | **Current** | 513 | 112( 22%) |
| **Recent year: Nuclear Medicine & Medical Imaging** | **Current** | 1007 | 219( 22%) |
| **Recent year: Endocrinology & Metabolism** | **Current** | 729 | 156( 21%) |
| **Recent year: Biophysics** | **Current** | 169 | 36( 21%) |
| **Recent year: Toxicology** | **Current** | 433 | 91( 21%) |
| **Recent year: Cardiovascular System & Hematology** | **Current** | 1694 | 354( 21%) |
| **Recent year: Respiratory System** | **Current** | 546 | 114( 21%) |
| **Recent year: Physiology** | **Current** | 228 | 47( 21%) |
| **Recent year: Rehabilitation** | **Current** | 199 | 41( 21%) |
| **Recent year: Microbiology** | **Current** | 1319 | 269( 20%) |
| **Recent year: Urology & Nephrology** | **Current** | 737 | 148( 20%) |
| **Recent year: Medicinal & Biomolecular Chemistry** | **Current** | 384 | 77( 20%) |
| **Recent year: Health Policy & Services** | **Current** | 191 | 38( 20%) |
| **Recent year: Pediatrics** | **Current** | 714 | 136( 19%) |
| **Recent year: Obstetrics & Reproductive Medicine** | **Current** | 637 | 119( 19%) |
| **Recent year: Anesthesiology** | **Current** | 403 | 72( 18%) |
| **Recent year: Statistics & Probability** | **Current** | 270 | 47( 17%) |
| **Recent year: Biochemistry & Molecular Biology** | **Current** | 1718 | 290( 17%) |
| **Recent year: Speech-Language Pathology & Audiology** | **Current** | 114 | 19( 17%) |
| **Recent year: Demography** | **Current** | 36 | 6( 17%) |
| **Recent year: Experimental Psychology** | **Current** | 610 | 97( 16%) |
| **Recent year: Pharmacology & Pharmacy** | **Current** | 624 | 96( 15%) |
| **Recent year: Clinical Psychology** | **Current** | 234 | 35( 15%) |
| **Recent year: Applied Ethics** | **Current** | 54 | 8( 15%) |
| **Recent year: Organic Chemistry** | **Current** | 574 | 85( 15%) |
| **Recent year: Nutrition & Dietetics** | **Current** | 320 | 46( 14%) |
| **Recent year: Mycology & Parasitology** | **Current** | 105 | 15( 14%) |
| **Recent year: Nursing** | **Current** | 764 | 109( 14%) |
| **Recent year: Optics** | **Current** | 437 | 61( 14%) |
| **Recent year: General Chemistry** | **Current** | 415 | 56( 13%) |
| **Recent year: General & Internal Medicine** | **Current** | 1293 | 173( 13%) |
| **Recent year: Surgery** | **Current** | 1125 | 150( 13%) |
| **Recent year: Nanoscience & Nanotechnology** | **Current** | 595 | 75( 13%) |
| **Recent year: Otorhinolaryngology** | **Current** | 506 | 62( 12%) |
| **Recent year: Environmental & Occupational Health** | **Current** | 98 | 12( 12%) |
| **Recent year: Tropical Medicine** | **Current** | 213 | 26( 12%) |
| **Recent year: Microscopy** | **Current** | 33 | 4( 12%) |
| **Recent year: Biotechnology** | **Current** | 185 | 22( 12%) |
| **Recent year: Drama & Theater** | **Current** | 10 | 1( 10%) |
| **Recent year: Complementary & Alternative Medicine** | **Current** | 50 | 5( 10%) |
| **Recent year: Acoustics** | **Current** | 233 | 23( 9.9%) |
| **Recent year: Dermatology & Venereal Diseases** | **Current** | 341 | 33( 9.7%) |
| **Recent year: Orthopedics** | **Current** | 716 | 68( 9.5%) |
| **Recent year: Family Studies** | **Current** | 55 | 5( 9.1%) |
| **Recent year: Environmental Sciences** | **Current** | 234 | 21( 9%) |
| **Recent year: Behavioral Science & Comparative Psychology** | **Current** | 127 | 11( 8.7%) |
| **Recent year: Sociology** | **Current** | 245 | 19( 7.8%) |
| **Recent year: Pathology** | **Current** | 299 | 23( 7.7%) |
| **Recent year: Design Practice & Management** | **Current** | 67 | 5( 7.5%) |
| **Recent year: Dentistry** | **Current** | 441 | 32( 7.3%) |
| **Recent year: Plant Biology & Botany** | **Current** | 683 | 49( 7.2%) |
| **Recent year: Evolutionary Biology** | **Current** | 366 | 26( 7.1%) |
| **Recent year: Artificial Intelligence & Image Processing** | **Current** | 1230 | 83( 6.7%) |
| **Recent year: Chemical Physics** | **Current** | 697 | 47( 6.7%) |
| **Recent year: Industrial Engineering & Automation** | **Current** | 492 | 33( 6.7%) |
| **Recent year: Criminology** | **Current** | 188 | 12( 6.4%) |
| **Recent year: Sport Sciences** | **Current** | 159 | 10( 6.3%) |
| **Recent year: Veterinary Sciences** | **Current** | 483 | 30( 6.2%) |
| **Recent year: Psychoanalysis** | **Current** | 33 | 2( 6.1%) |
| **Recent year: Social Work** | **Current** | 83 | 5( 6%) |
| **Recent year: Polymers** | **Current** | 375 | 22( 5.9%) |
| **Recent year: General Clinical Medicine** | **Current** | 103 | 6( 5.8%) |
| **Recent year: Entomology** | **Current** | 259 | 15( 5.8%) |
| **Recent year: Religions & Theology** | **Current** | 82 | 4( 4.9%) |
| **Recent year: Distributed Computing** | **Current** | 103 | 5( 4.9%) |
| **Recent year: Computer Hardware & Architecture** | **Current** | 226 | 10( 4.4%) |
| **Recent year: Information & Library Sciences** | **Current** | 95 | 4( 4.2%) |
| **Recent year: Education** | **Current** | 699 | 28( 4%) |
| **Recent year: Logistics & Transportation** | **Current** | 151 | 6( 4%) |
| **Recent year: Social Psychology** | **Current** | 562 | 22( 3.9%) |
| **Recent year: Applied Mathematics** | **Current** | 104 | 4( 3.8%) |
| **Recent year: Legal & Forensic Medicine** | **Current** | 53 | 2( 3.8%) |
| **Recent year: Physical Chemistry** | **Current** | 107 | 4( 3.7%) |
| **Recent year: Development Studies** | **Current** | 27 | 1( 3.7%) |
| **Recent year: Economics** | **Current** | 586 | 21( 3.6%) |
| **Recent year: Anatomy & Morphology** | **Current** | 56 | 2( 3.6%) |
| **Recent year: Food Science** | **Current** | 169 | 6( 3.6%) |
| **Recent year: Ornithology** | **Current** | 29 | 1( 3.4%) |
| **Recent year: Geography** | **Current** | 120 | 4( 3.3%) |
| **Recent year: Electrical & Electronic Engineering** | **Current** | 364 | 12( 3.3%) |
| **Recent year: Networking & Telecommunications** | **Current** | 1008 | 33( 3.3%) |
| **Recent year: Dairy & Animal Science** | **Current** | 336 | 11( 3.3%) |
| **Recent year: General Psychology & Cognitive Sciences** | **Current** | 64 | 2( 3.1%) |
| **Recent year: Fluids & Plasmas** | **Current** | 325 | 10( 3.1%) |
| **Recent year: Social Sciences Methods** | **Current** | 100 | 3( 3%) |
| **Recent year: Numerical & Computational Mathematics** | **Current** | 101 | 3( 3%) |
| **Recent year: Building & Construction** | **Current** | 102 | 3( 2.9%) |
| **Recent year: Mechanical Engineering & Transports** | **Current** | 487 | 14( 2.9%) |
| **Recent year: Science Studies** | **Current** | 35 | 1( 2.9%) |
| **Recent year: Cultural Studies** | **Current** | 36 | 1( 2.8%) |
| **Recent year: Marketing** | **Current** | 254 | 7( 2.8%) |
| **Recent year: Anthropology** | **Current** | 116 | 3( 2.6%) |
| **Recent year: Materials** | **Current** | 980 | 24( 2.4%) |
| **Recent year: Operations Research** | **Current** | 167 | 4( 2.4%) |
| **Recent year: Mining & Metallurgy** | **Current** | 43 | 1( 2.3%) |
| **Recent year: Optoelectronics & Photonics** | **Current** | 697 | 16( 2.3%) |
| **Recent year: Human Factors** | **Current** | 177 | 4( 2.3%) |
| **Recent year: Software Engineering** | **Current** | 178 | 4( 2.2%) |
| **Recent year: Finance** | **Current** | 181 | 4( 2.2%) |
| **Recent year: Meteorology & Atmospheric Sciences** | **Current** | 1047 | 23( 2.2%) |
| **Recent year: Communication & Media Studies** | **Current** | 187 | 4( 2.1%) |
| **Recent year: Econometrics** | **Current** | 49 | 1( 2%) |
| **Recent year: Political Science & Public Administration** | **Current** | 345 | 7( 2%) |
| **Recent year: Languages & Linguistics** | **Current** | 99 | 2( 2%) |
| **Recent year: General Physics** | **Current** | 449 | 9( 2%) |
| **Recent year: Archaeology** | **Current** | 101 | 2( 2%) |
| **Recent year: Sport, Leisure & Tourism** | **Current** | 104 | 2( 1.9%) |
| **Recent year: Environmental Engineering** | **Current** | 316 | 6( 1.9%) |
| **Recent year: Energy** | **Current** | 850 | 16( 1.9%) |
| **Recent year: Marine Biology & Hydrobiology** | **Current** | 287 | 5( 1.7%) |
| **Recent year: Ecology** | **Current** | 811 | 13( 1.6%) |
| **Recent year: Chemical Engineering** | **Current** | 188 | 3( 1.6%) |
| **Recent year: Philosophy** | **Current** | 127 | 2( 1.6%) |
| **Recent year: Aerospace & Aeronautics** | **Current** | 638 | 10( 1.6%) |
| **Recent year: Inorganic & Nuclear Chemistry** | **Current** | 205 | 3( 1.5%) |
| **Recent year: Agricultural Economics & Policy** | **Current** | 69 | 1( 1.4%) |
| **Recent year: Geological & Geomatics Engineering** | **Current** | 282 | 4( 1.4%) |
| **Recent year: Accounting** | **Current** | 73 | 1( 1.4%) |
| **Recent year: Applied Physics** | **Current** | 1547 | 19( 1.2%) |
| **Recent year: Business & Management** | **Current** | 818 | 10( 1.2%) |
| **Recent year: Information Systems** | **Current** | 169 | 2( 1.2%) |
| **Recent year: Computation Theory & Mathematics** | **Current** | 176 | 2( 1.1%) |
| **Recent year: Astronomy & Astrophysics** | **Current** | 638 | 6( .94%) |
| **Recent year: Geochemistry & Geophysics** | **Current** | 741 | 6( .81%) |
| **Recent year: Strategic, Defence & Security Studies** | **Current** | 130 | 1( .77%) |
| **Recent year: Paleontology** | **Current** | 136 | 1( .74%) |
| **Recent year: Literary Studies** | **Current** | 138 | 1( .72%) |
| **Recent year: General Mathematics** | **Current** | 423 | 3( .71%) |
| **Recent year: Nuclear & Particle Physics** | **Current** | 945 | 6( .63%) |
| **Recent year: Oceanography** | **Current** | 168 | 1( .6%) |
| **Recent year: Agronomy & Agriculture** | **Current** | 361 | 2( .55%) |
| **Recent year: Fisheries** | **Current** | 187 | 1( .53%) |
| **Recent year: Zoology** | **Current** | 92 | 0( 0%) |
| **Recent year: Urban & Regional Planning** | **Current** | 71 | 0( 0%) |
| **Recent year: Music** | **Current** | 37 | 0( 0%) |
| **Recent year: Mathematical Physics** | **Current** | 21 | 0( 0%) |
| **Recent year: Law** | **Current** | 92 | 0( 0%) |
| **Recent year: International Relations** | **Current** | 56 | 0( 0%) |
| **Recent year: Industrial Relations** | **Current** | 7 | 0( 0%) |
| **Recent year: Horticulture** | **Current** | 60 | 0( 0%) |
| **Recent year: History of Social Sciences** | **Current** | 17 | 0( 0%) |
| **Recent year: History of Science, Technology & Medicine** | **Current** | 15 | 0( 0%) |
| **Recent year: History** | **Current** | 101 | 0( 0%) |
| **Recent year: Geology** | **Current** | 82 | 0( 0%) |
| **Recent year: Gender Studies** | **Current** | 15 | 0( 0%) |
| **Recent year: Forestry** | **Current** | 175 | 0( 0%) |
| **Recent year: Folklore** | **Current** | 7 | 0( 0%) |
| **Recent year: Economic Theory** | **Current** | 16 | 0( 0%) |
| **Recent year: Classics** | **Current** | 21 | 0( 0%) |
| **Recent year: Civil Engineering** | **Current** | 209 | 0( 0%) |
| **Recent year: Automobile Design & Engineering** | **Current** | 7 | 0( 0%) |
| **Recent year: Art Practice, History & Theory** | **Current** | 24 | 0( 0%) |
| **Recent year: Architecture** | **Current** | 10 | 0( 0%) |

**Supplementary Table 2.1 : Career-long impact, Funding time any funding citation counts and composite citation indices for each subfield (ordered by percentage funded)**

| **Top-cited US-based researchers: Subfield (perc. funded)** | **Classification** | **Citations for funded, median** | **Citations for non-funded, median** | **p-value** | **Composite index for funded, median** | **Composite index for non-funded, median** | **p-value** |
| --- | --- | --- | --- | --- | --- | --- | --- |
| **Developmental Biology ( 89%)** | Highly related fields | 12842 | 10942 | 0.003 | 3.82 | 3.76 | 0.014 |
| **Substance Abuse ( 87%)** | Highly related fields | 7834 | 5392 | 0.005 | 3.73 | 3.64 | 0.063 |
| **Immunology ( 85%)** | Highly related fields | 13888 | 12865 | 0.012 | 3.81 | 3.68 | <0.001 |
| **Geriatrics ( 85%)** | Highly related fields | 9847 | 4994 | 0.003 | 3.64 | 3.53 | 0.142 |
| **Biochemistry & Molecular Biology ( 81%)** | Highly related fields | 10284 | 7192 | <0.001 | 3.79 | 3.7 | <0.001 |
| **Endocrinology & Metabolism ( 81%)** | Highly related fields | 12233 | 9650 | <0.001 | 3.79 | 3.72 | <0.001 |
| **Gerontology ( 80%)** | Highly related fields | 7152 | 3317 | 0.001 | 3.74 | 3.5 | 0.004 |
| **Virology ( 80%)** | Highly related fields | 10352 | 7624 | <0.001 | 3.63 | 3.55 | <0.001 |
| **Neurology & Neurosurgery ( 79%)** | Highly related fields | 11476 | 8561 | <0.001 | 3.81 | 3.7 | <0.001 |
| **Genetics & Heredity ( 79%)** | Highly related fields | 13806 | 8860 | <0.001 | 3.68 | 3.62 | 0.063 |
| **Bioinformatics ( 77%)** | Highly related fields | 12262 | 7761 | 0.012 | 3.63 | 3.52 | 0.019 |
| **Psychiatry ( 76%)** | Highly related fields | 12995 | 8047 | <0.001 | 3.86 | 3.72 | <0.001 |
| **Oncology & Carcinogenesis ( 74%)** | Highly related fields | 14252 | 11172 | <0.001 | 3.7 | 3.59 | <0.001 |
| **Public Health ( 73%)** | Highly related fields | 7667 | 6619 | 0.001 | 3.68 | 3.61 | 0.004 |
| **Medical Informatics ( 71%)** | Highly related fields | 4352 | 3404 | 0.041 | 3.34 | 3.17 | 0.009 |
| **Demography ( 71%)** | Highly related fields | 4286 | 2260 | 0.058 | 3.6 | 3.56 | 0.911 |
| **Physiology ( 71%)** | Highly related fields | 6838 | 4617 | <0.001 | 3.75 | 3.66 | 0.013 |
| **Epidemiology ( 71%)** | Highly related fields | 16480 | 10546 | 0.005 | 3.89 | 3.73 | 0.237 |
| **Biomedical Engineering ( 71%)** | Highly related fields | 6113 | 4070 | <0.001 | 3.39 | 3.29 | 0.001 |
| **Developmental & Child Psychology ( 70%)** | Highly related fields | 8495 | 5279 | <0.001 | 3.81 | 3.65 | <0.001 |
| **Arthritis & Rheumatology ( 70%)** | Highly related fields | 13517 | 12504 | 0.049 | 3.79 | 3.73 | 0.021 |
| **Allergy ( 69%)** | Highly related fields | 9587 | 7563 | 0.003 | 3.77 | 3.66 | 0.030 |
| **Biophysics ( 69%)** | Highly related fields | 7676 | 2939 | <0.001 | 3.66 | 3.35 | <0.001 |
| **Respiratory System ( 66%)** | Highly related fields | 11186 | 9769 | 0.013 | 3.69 | 3.67 | 0.489 |
| **Experimental Psychology ( 65%)** | Highly related fields | 7137 | 5142 | <0.001 | 3.8 | 3.73 | 0.004 |
| **Gastroenterology & Hepatology ( 65%)** | Highly related fields | 11005 | 8016 | <0.001 | 3.74 | 3.66 | <0.001 |
| **Cardiovascular System & Hematology ( 64%)** | Highly related fields | 14741 | 12270 | <0.001 | 3.79 | 3.69 | <0.001 |
| **Health Policy & Services ( 63%)** | Highly related fields | 7915 | 4993 | 0.002 | 3.65 | 3.54 | 0.013 |
| **Urology & Nephrology ( 63%)** | Highly related fields | 10090 | 7520 | <0.001 | 3.71 | 3.58 | <0.001 |
| **Speech-Language Pathology & Audiology ( 61%)** | Highly related fields | 3170 | 3904 | 0.347 | 3.48 | 3.5 | 0.417 |
| **Clinical Psychology ( 60%)** | Highly related fields | 10172 | 5683 | <0.001 | 3.82 | 3.7 | 0.004 |
| **Pediatrics ( 60%)** | Highly related fields | 6112 | 4322 | <0.001 | 3.47 | 3.36 | <0.001 |
| **Microbiology ( 60%)** | Highly related fields | 8973 | 7541 | <0.001 | 3.68 | 3.62 | <0.001 |
| **Family Studies ( 59%)** | Other fields | 3575 | 2278 | 0.007 | 3.59 | 3.39 | 0.048 |
| **Applied Ethics ( 58%)** | Highly related fields | 4638 | 2669 | 0.003 | 3.64 | 3.47 | 0.056 |
| **Nutrition & Dietetics ( 57%)** | Highly related fields | 7903 | 5857 | <0.001 | 3.73 | 3.6 | 0.001 |
| **Ophthalmology & Optometry ( 57%)** | Highly related fields | 7416 | 4649 | <0.001 | 3.58 | 3.47 | <0.001 |
| **Organic Chemistry ( 57%)** | Other fields | 7187 | 5401 | <0.001 | 3.67 | 3.55 | <0.001 |
| **Environmental & Occupational Health ( 57%)** | Highly related fields | 4184 | 3353 | 0.012 | 3.38 | 3.26 | 0.005 |
| **Toxicology ( 56%)** | Highly related fields | 6442 | 3944 | <0.001 | 3.49 | 3.39 | <0.001 |
| **Nursing ( 55%)** | Highly related fields | 2018 | 1258 | <0.001 | 3.12 | 3.03 | <0.001 |
| **Obstetrics & Reproductive Medicine ( 54%)** | Highly related fields | 7616 | 5005 | <0.001 | 3.61 | 3.46 | <0.001 |
| **Emergency & Critical Care Medicine ( 53%)** | Highly related fields | 8576 | 5130 | <0.001 | 3.52 | 3.43 | 0.001 |
| **Rehabilitation ( 53%)** | Highly related fields | 4662 | 3686 | 0.001 | 3.47 | 3.35 | 0.003 |
| **Pharmacology & Pharmacy ( 51%)** | Highly related fields | 5538 | 3620 | <0.001 | 3.45 | 3.31 | <0.001 |
| **Behavioral Science & Comparative Psychology ( 50%)** | Highly related fields | 5072 | 3916 | <0.001 | 3.78 | 3.69 | 0.048 |
| **Analytical Chemistry ( 49%)** | Other fields | 6609 | 3696 | <0.001 | 3.48 | 3.36 | <0.001 |
| **Nuclear Medicine & Medical Imaging ( 47%)** | Highly related fields | 7837 | 5258 | <0.001 | 3.55 | 3.39 | <0.001 |
| **Otorhinolaryngology ( 44%)** | Highly related fields | 3940 | 3319 | 0.001 | 3.38 | 3.33 | 0.004 |
| **Dentistry ( 44%)** | Highly related fields | 4572 | 2814 | <0.001 | 3.46 | 3.34 | <0.001 |
| **Tropical Medicine ( 44%)** | Highly related fields | 6738 | 4014 | <0.001 | 3.52 | 3.32 | <0.001 |
| **Mycology & Parasitology ( 43%)** | Highly related fields | 4813 | 4646 | 0.539 | 3.41 | 3.49 | 0.998 |
| **Statistics & Probability ( 43%)** | Other fields | 11431 | 5598 | <0.001 | 3.78 | 3.66 | 0.004 |
| **Medicinal & Biomolecular Chemistry ( 43%)** | Highly related fields | 4644 | 3581 | <0.001 | 3.28 | 3.14 | <0.001 |
| **Anesthesiology ( 43%)** | Highly related fields | 6281 | 4140 | <0.001 | 3.47 | 3.37 | <0.001 |
| **Dermatology & Venereal Diseases ( 42%)** | Highly related fields | 8008 | 4697 | <0.001 | 3.68 | 3.54 | <0.001 |
| **Biotechnology ( 41%)** | Highly related fields | 6671 | 4219 | <0.001 | 3.44 | 3.27 | 0.004 |
| **Social Psychology ( 41%)** | Highly related fields | 10567 | 6215 | <0.001 | 3.93 | 3.75 | <0.001 |
| **Surgery ( 40%)** | Highly related fields | 8570 | 5061 | <0.001 | 3.47 | 3.38 | <0.001 |
| **General & Internal Medicine ( 40%)** | Highly related fields | 5786 | 2779 | <0.001 | 3.35 | 3.14 | <0.001 |
| **Complementary & Alternative Medicine ( 40%)** | Highly related fields | 2674 | 981 | <0.001 | 3.01 | 2.86 | 0.042 |
| **Sport Sciences ( 37%)** | Highly related fields | 6500 | 4860 | 0.003 | 3.56 | 3.52 | 0.045 |
| **Acoustics ( 36%)** | Other fields | 3620 | 2680 | 0.001 | 3.44 | 3.4 | 0.215 |
| **History of Social Sciences ( 35%)** | Other fields | 1745 | 1740 | 0.651 | 3.46 | 3.25 | 0.796 |
| **Nanoscience & Nanotechnology ( 35%)** | Other fields | 14211 | 12246 | 0.032 | 3.63 | 3.53 | 0.079 |
| **General Chemistry ( 34%)** | Other fields | 7990 | 3518 | <0.001 | 3.63 | 3.3 | <0.001 |
| **Microscopy ( 33%)** | Highly related fields | 9950 | 4837 | 0.035 | 3.57 | 3.45 | 0.202 |
| **Criminology ( 31%)** | Other fields | 4848 | 3566 | 0.001 | 3.62 | 3.6 | 0.255 |
| **Sociology ( 31%)** | Other fields | 4018 | 3670 | 0.193 | 3.66 | 3.7 | 0.291 |
| **Optics ( 30%)** | Other fields | 6704 | 5176 | <0.001 | 3.38 | 3.3 | 0.088 |
| **Pathology ( 30%)** | Highly related fields | 10145 | 8933 | 0.039 | 3.69 | 3.61 | 0.321 |
| **Social Work ( 30%)** | Other fields | 2179 | 1526 | 0.058 | 3.26 | 3.23 | 0.100 |
| **Evolutionary Biology ( 29%)** | Other fields | 8799 | 6563 | <0.001 | 3.9 | 3.78 | 0.002 |
| **Orthopedics ( 28%)** | Highly related fields | 8903 | 4973 | <0.001 | 3.64 | 3.45 | <0.001 |
| **Social Sciences Methods ( 27%)** | Other fields | 9838 | 4916 | 0.001 | 4.02 | 3.71 | 0.002 |
| **Chemical Physics ( 27%)** | Other fields | 11822 | 7682 | <0.001 | 3.88 | 3.77 | <0.001 |
| **General Clinical Medicine ( 24%)** | Highly related fields | 3716 | 2040 | <0.001 | 3.44 | 3.03 | <0.001 |
| **Plant Biology & Botany ( 24%)** | Other fields | 8824 | 5160 | <0.001 | 3.69 | 3.47 | <0.001 |
| **Inorganic & Nuclear Chemistry ( 24%)** | Other fields | 7773 | 5586 | 0.023 | 3.65 | 3.52 | 0.005 |
| **Veterinary Sciences ( 23%)** | Highly related fields | 4095 | 2952 | <0.001 | 3.37 | 3.3 | 0.041 |
| **Environmental Sciences ( 21%)** | Other fields | 7461 | 4139 | <0.001 | 3.49 | 3.36 | 0.001 |
| **Human Factors ( 19%)** | Highly related fields | 5312 | 4990 | 0.411 | 3.62 | 3.58 | 0.702 |
| **Ornithology ( 18%)** | Other fields | 2028 | 3711 | 0.148 | 3.4 | 3.44 | 0.852 |
| **Urban & Regional Planning ( 18%)** | Other fields | 3257 | 2278 | 0.268 | 3.5 | 3.54 | 0.388 |
| **Economics ( 18%)** | Other fields | 5788 | 4649 | 0.021 | 3.74 | 3.72 | 0.247 |
| **Artificial Intelligence & Image Processing ( 17%)** | Other fields | 6266 | 4047 | <0.001 | 3.47 | 3.33 | <0.001 |
| **Polymers ( 17%)** | Other fields | 7916 | 5343 | <0.001 | 3.57 | 3.48 | 0.164 |
| **Economic Theory ( 17%)** | Other fields | 3669 | 2701 | 0.859 | 3.56 | 3.61 | 0.953 |
| **Development Studies ( 17%)** | Other fields | 3826 | 3565 | 0.757 | 3.56 | 3.57 | 0.535 |
| **Anatomy & Morphology ( 17%)** | Highly related fields | 2612 | 1419 | 0.088 | 3.02 | 3.07 | 0.198 |
| **Distributed Computing ( 17%)** | Other fields | 4890 | 3099 | 0.001 | 3.24 | 3.13 | 0.072 |
| **Entomology ( 16%)** | Other fields | 4967 | 3189 | <0.001 | 3.59 | 3.46 | 0.036 |
| **Gender Studies ( 15%)** | Highly related fields | 2960 | 2284 | 0.324 | 3.61 | 3.5 | 0.236 |
| **History of Science, Technology & Medicine ( 15%)** | Other fields | 957 | 761 | 0.874 | 3.15 | 3.12 | 0.958 |
| **Software Engineering ( 15%)** | Other fields | 4660 | 4504 | 0.191 | 3.48 | 3.46 | 0.437 |
| **Education ( 15%)** | Other fields | 3374 | 2290 | <0.001 | 3.44 | 3.35 | <0.001 |
| **Food Science ( 15%)** | Other fields | 6002 | 3877 | <0.001 | 3.57 | 3.36 | 0.005 |
| **Industrial Engineering & Automation ( 14%)** | Other fields | 5380 | 4123 | 0.002 | 3.52 | 3.43 | 0.165 |
| **Logistics & Transportation ( 14%)** | Other fields | 2504 | 2848 | 0.353 | 3.19 | 3.29 | 0.055 |
| **Geography ( 14%)** | Other fields | 4866 | 3227 | 0.002 | 3.83 | 3.66 | 0.005 |
| **Sport, Leisure & Tourism ( 13%)** | Other fields | 3071 | 3757 | 0.267 | 3.59 | 3.56 | 0.453 |
| **Numerical & Computational Mathematics ( 13%)** | Other fields | 4001 | 3507 | 0.163 | 3.59 | 3.54 | 0.821 |
| **Communication & Media Studies ( 12%)** | Other fields | 3009 | 2781 | 0.369 | 3.54 | 3.48 | 0.357 |
| **General Psychology & Cognitive Sciences ( 12%)** | Highly related fields | 3202 | 3365 | 0.811 | 3.6 | 3.47 | 0.348 |
| **Fluids & Plasmas ( 12%)** | Other fields | 8612 | 6618 | 0.014 | 3.8 | 3.72 | 0.114 |
| **Information & Library Sciences ( 12%)** | Other fields | 2294 | 1245 | 0.012 | 3.44 | 3.14 | 0.051 |
| **Dairy & Animal Science ( 12%)** | Other fields | 4046 | 3011 | 0.005 | 3.34 | 3.24 | 0.158 |
| **Optoelectronics & Photonics ( 12%)** | Other fields | 3848 | 2806 | <0.001 | 3.15 | 3.11 | 0.137 |
| **Anthropology ( 11%)** | Other fields | 4001 | 3204 | 0.327 | 3.73 | 3.6 | 0.588 |
| **Religions & Theology ( 11%)** | Other fields | 784 | 475 | 0.508 | 2.89 | 2.85 | 0.729 |
| **Drama & Theater ( 11%)** | Other fields | 606 | 357 | 0.439 | 2.8 | 2.67 | 0.439 |
| **Design Practice & Management ( 11%)** | Other fields | 4323 | 2892 | 0.563 | 3.24 | 3.28 | 0.882 |
| **Environmental Engineering ( 11%)** | Other fields | 5009 | 4072 | 0.042 | 3.42 | 3.44 | 0.723 |
| **Chemical Engineering ( 11%)** | Other fields | 4285 | 4074 | 0.295 | 3.44 | 3.43 | 0.811 |
| **Applied Mathematics ( 11%)** | Other fields | 4476 | 4595 | 0.835 | 3.52 | 3.61 | 0.926 |
| **Marketing ( 11%)** | Other fields | 6545 | 5722 | 0.293 | 3.77 | 3.64 | 0.368 |
| **Zoology ( 10%)** | Other fields | 2070 | 2115 | 0.862 | 3.24 | 3.19 | 0.312 |
| **Architecture ( 10%)** | Other fields | 465 | 238 | 0.384 | 2.45 | 2.48 | 0.602 |
| **Ecology ( 9.7%)** | Other fields | 7557 | 7548 | 0.412 | 3.83 | 3.74 | 0.019 |
| **Marine Biology & Hydrobiology ( 9.7%)** | Other fields | 6058 | 6136 | 0.768 | 3.66 | 3.7 | 0.726 |
| **Networking & Telecommunications ( 9.5%)** | Other fields | 3715 | 3691 | 0.192 | 3.29 | 3.25 | 0.497 |
| **Computer Hardware & Architecture ( 9.3%)** | Other fields | 4918 | 4403 | 0.823 | 3.25 | 3.33 | 0.505 |
| **Electrical & Electronic Engineering ( 9.2%)** | Other fields | 3502 | 2503 | 0.009 | 3.04 | 3.04 | 0.740 |
| **Geological & Geomatics Engineering ( 9%)** | Other fields | 6690 | 4526 | 0.060 | 3.71 | 3.44 | 0.034 |
| **Information Systems ( 8.6%)** | Other fields | 6086 | 5645 | 0.197 | 3.61 | 3.58 | 0.990 |
| **Law ( 8.5%)** | Other fields | 1025 | 1252 | 0.764 | 3.23 | 3.22 | 0.764 |
| **Mathematical Physics ( 8.3%)** | Other fields | 2324 | 3075 | 0.465 | 3.37 | 3.53 | 0.144 |
| **Strategic, Defence & Security Studies ( 8.2%)** | Other fields | 2495 | 2076 | 0.183 | 3.49 | 3.26 | 0.009 |
| **Agricultural Economics & Policy ( 8.2%)** | Other fields | 3642 | 4084 | 0.888 | 3.58 | 3.58 | 0.534 |
| **General Physics ( 8.2%)** | Other fields | 6484 | 5568 | 0.295 | 3.6 | 3.57 | 0.903 |
| **Languages & Linguistics ( 8.2%)** | Other fields | 2937 | 1820 | 0.082 | 3.59 | 3.42 | 0.061 |
| **Mining & Metallurgy ( 8.1%)** | Other fields | 1218 | 954 | 0.544 | 2.74 | 2.76 | 0.826 |
| **Psychoanalysis ( 8%)** | Highly related fields | 1245 | 1338 | 0.943 | 3.23 | 3.31 | 0.543 |
| **Political Science & Public Administration ( 7.9%)** | Other fields | 5258 | 3489 | 0.023 | 3.76 | 3.63 | 0.231 |
| **Building & Construction ( 7.8%)** | Other fields | 3477 | 2960 | 0.135 | 3.32 | 3.32 | 0.887 |
| **Oceanography ( 7.6%)** | Other fields | 6151 | 4641 | 0.239 | 3.68 | 3.58 | 0.247 |
| **Literary Studies ( 7.5%)** | Other fields | 382 | 351 | 0.686 | 2.76 | 2.7 | 0.620 |
| **Cultural Studies ( 7.4%)** | Other fields | 874 | 795 | 0.597 | 2.99 | 3.11 | 0.092 |
| **Materials ( 7.3%)** | Other fields | 4898 | 3408 | <0.001 | 3.45 | 3.27 | 0.002 |
| **Applied Physics ( 7.2%)** | Other fields | 7915 | 6332 | 0.007 | 3.64 | 3.53 | 0.002 |
| **Econometrics ( 7.1%)** | Other fields | 17980 | 6910 | 0.213 | 3.88 | 3.97 | 0.574 |
| **Mechanical Engineering & Transports ( 7.1%)** | Other fields | 3309 | 2869 | 0.343 | 3.4 | 3.36 | 0.747 |
| **Operations Research ( 6.8%)** | Other fields | 3871 | 4231 | 0.201 | 3.55 | 3.6 | 0.229 |
| **Finance ( 6.8%)** | Other fields | 3236 | 4958 | 0.314 | 3.53 | 3.61 | 0.530 |
| **Business & Management ( 6.5%)** | Other fields | 6365 | 6058 | 0.535 | 3.75 | 3.69 | 0.244 |
| **Meteorology & Atmospheric Sciences ( 6.4%)** | Other fields | 8805 | 8746 | 0.863 | 3.76 | 3.74 | 0.918 |
| **General Mathematics ( 6.4%)** | Other fields | 1961 | 2247 | 0.781 | 3.42 | 3.43 | 0.577 |
| **Computation Theory & Mathematics ( 6.2%)** | Other fields | 5982 | 4894 | 0.316 | 3.68 | 3.62 | 0.535 |
| **Fisheries ( 6.2%)** | Other fields | 3816 | 3591 | 0.876 | 3.34 | 3.41 | 0.950 |
| **Philosophy ( 6.2%)** | Other fields | 1662 | 1353 | 0.492 | 3.43 | 3.38 | 0.608 |
| **Archaeology ( 6.1%)** | Other fields | 1927 | 1923 | 0.986 | 3.18 | 3.31 | 0.090 |
| **Paleontology ( 6%)** | Other fields | 6741 | 5589 | 0.573 | 3.84 | 3.71 | 0.410 |
| **Energy ( 5.9%)** | Other fields | 4465 | 3323 | 0.056 | 3.28 | 3.26 | 0.768 |
| **Agronomy & Agriculture ( 5.9%)** | Other fields | 5820 | 3930 | 0.248 | 3.46 | 3.39 | 0.508 |
| **Aerospace & Aeronautics ( 5.8%)** | Other fields | 2062 | 1839 | 0.170 | 3.13 | 3.13 | 0.903 |
| **Astronomy & Astrophysics ( 5.6%)** | Other fields | 10241 | 10762 | 0.789 | 3.77 | 3.74 | 0.185 |
| **Classics ( 5.6%)** | Other fields | 549 | 534 | 0.772 | 2.86 | 2.92 | 0.289 |
| **Geology ( 5.4%)** | Other fields | 3832 | 3385 | 0.938 | 3.52 | 3.48 | 0.636 |
| **Forestry ( 5.3%)** | Other fields | 2454 | 2711 | 0.935 | 3.29 | 3.22 | 0.598 |
| **Geochemistry & Geophysics ( 4.9%)** | Other fields | 6027 | 6442 | 0.993 | 3.8 | 3.73 | 0.851 |
| **Nuclear & Particle Physics ( 4.7%)** | Other fields | 6028 | 6744 | 0.215 | 3.61 | 3.62 | 0.668 |
| **History ( 4.5%)** | Other fields | 543 | 530 | 0.733 | 2.92 | 2.88 | 0.798 |
| **International Relations ( 4.4%)** | Other fields | 2949 | 2338 | 0.362 | 3.54 | 3.5 | 0.378 |
| **Physical Chemistry ( 4.4%)** | Other fields | 15148 | 6278 | 0.107 | 3.87 | 3.51 | 0.210 |
| **Science Studies ( 3.8%)** | Other fields | 4961 | 4004 | 0.549 | 3.59 | 3.69 | 0.257 |
| **Civil Engineering ( 3.6%)** | Other fields | 1610 | 2205 | 0.242 | 3.04 | 3.17 | 0.139 |
| **Legal & Forensic Medicine ( 3%)** | Highly related fields | 1132 | 1332 | 0.654 | 2.85 | 2.98 | 0.190 |
| **Music ( 2.4%)** | Other fields | 575 | 437 | 0.302 | 3.03 | 2.78 | 0.127 |
| **Accounting ( 2.4%)** | Other fields | 10654 | 5375 | 0.173 | 3.9 | 3.54 | 0.173 |
| **Horticulture ( 1.3%)** | Other fields | 1584 | 2110 | 0.644 | 3.12 | 3.14 | 0.853 |
| **Industrial Relations ( 0%)** | Other fields | . | 1524 |  | . | 3.34 |  |
| **Folklore ( 0%)** | Other fields | . | 310 |  | . | 2.61 |  |
| **Automobile Design & Engineering ( 0%)** | Other fields | . | 860 |  | . | 2.69 |  |
| **Art Practice, History & Theory ( 0%)** | Other fields | . | 210 |  | . | 2.47 |  |

**Supplementary Table 2.2 : Career-long impact, Funding time recent funding citation counts and composite citation indices for each subfield (ordered by percentage funded)**

| **Top-cited US-based researchers: Subfield (perc. funded)** | **Classification** | **Citations for funded, median** | **Citations for non-funded, median** | **p-value** | **Composite index for funded, median** | **Composite index for non-funded, median** | **p-value** |
| --- | --- | --- | --- | --- | --- | --- | --- |
| **Developmental Biology ( 42%)** | Highly related fields | 12967 | 12512 | 0.048 | 3.8 | 3.82 | 0.433 |
| **Bioinformatics ( 41%)** | Highly related fields | 15297 | 8222 | <0.001 | 3.69 | 3.54 | 0.056 |
| **Geriatrics ( 40%)** | Highly related fields | 12950 | 8067 | 0.010 | 3.71 | 3.62 | 0.306 |
| **Substance Abuse ( 36%)** | Highly related fields | 8516 | 7193 | 0.016 | 3.68 | 3.73 | 0.422 |
| **Biomedical Engineering ( 34%)** | Highly related fields | 6113 | 5039 | 0.011 | 3.37 | 3.36 | 0.958 |
| **Medical Informatics ( 34%)** | Highly related fields | 4194 | 3967 | 0.275 | 3.35 | 3.3 | 0.155 |
| **Immunology ( 34%)** | Highly related fields | 14470 | 13093 | 0.003 | 3.83 | 3.77 | 0.024 |
| **Epidemiology ( 34%)** | Highly related fields | 15811 | 14374 | 0.671 | 3.74 | 3.86 | 0.519 |
| **Virology ( 34%)** | Highly related fields | 1.0e+04 | 9705 | 0.109 | 3.6 | 3.61 | 0.753 |
| **Neurology & Neurosurgery ( 32%)** | Highly related fields | 11562 | 10342 | <0.001 | 3.78 | 3.78 | 0.728 |
| **Public Health ( 31%)** | Highly related fields | 7869 | 7198 | 0.005 | 3.71 | 3.63 | 0.015 |
| **Gerontology ( 30%)** | Highly related fields | 7551 | 6134 | 0.017 | 3.74 | 3.62 | 0.122 |
| **Oncology & Carcinogenesis ( 30%)** | Highly related fields | 15064 | 12526 | <0.001 | 3.66 | 3.68 | 0.319 |
| **Genetics & Heredity ( 29%)** | Highly related fields | 15385 | 11562 | 0.001 | 3.7 | 3.67 | 0.548 |
| **Demography ( 26%)** | Highly related fields | 2837 | 3432 | 0.830 | 3.51 | 3.61 | 0.466 |
| **Health Policy & Services ( 26%)** | Highly related fields | 7301 | 7555 | 0.177 | 3.62 | 3.59 | 0.209 |
| **Developmental & Child Psychology ( 25%)** | Highly related fields | 9051 | 6643 | <0.001 | 3.86 | 3.73 | <0.001 |
| **Applied Ethics ( 25%)** | Highly related fields | 6159 | 3077 | <0.001 | 3.67 | 3.55 | 0.110 |
| **Gastroenterology & Hepatology ( 25%)** | Highly related fields | 12311 | 9249 | 0.001 | 3.73 | 3.7 | 0.076 |
| **Endocrinology & Metabolism ( 25%)** | Highly related fields | 11786 | 11565 | 0.417 | 3.78 | 3.77 | 0.685 |
| **Biophysics ( 25%)** | Highly related fields | 6072 | 5727 | 0.170 | 3.56 | 3.5 | 0.752 |
| **Psychiatry ( 25%)** | Highly related fields | 14867 | 10684 | <0.001 | 3.9 | 3.81 | 0.031 |
| **Arthritis & Rheumatology ( 24%)** | Highly related fields | 13201 | 13569 | 0.867 | 3.78 | 3.78 | 0.484 |
| **Emergency & Critical Care Medicine ( 24%)** | Highly related fields | 10183 | 6102 | <0.001 | 3.54 | 3.46 | 0.043 |
| **Microbiology ( 23%)** | Highly related fields | 9152 | 8088 | 0.010 | 3.66 | 3.65 | 0.419 |
| **Allergy ( 23%)** | Highly related fields | 9374 | 8456 | 0.298 | 3.65 | 3.75 | 0.418 |
| **Respiratory System ( 23%)** | Highly related fields | 11174 | 10570 | 0.577 | 3.65 | 3.69 | 0.583 |
| **Clinical Psychology ( 23%)** | Highly related fields | 10384 | 7198 | 0.002 | 3.88 | 3.74 | 0.029 |
| **Cardiovascular System & Hematology ( 23%)** | Highly related fields | 15696 | 13298 | <0.001 | 3.77 | 3.75 | 0.165 |
| **Biochemistry & Molecular Biology ( 22%)** | Highly related fields | 10813 | 9176 | <0.001 | 3.77 | 3.76 | 0.697 |
| **Physiology ( 22%)** | Highly related fields | 6498 | 6188 | 0.112 | 3.74 | 3.71 | 0.457 |
| **Ophthalmology & Optometry ( 21%)** | Highly related fields | 7658 | 5548 | <0.001 | 3.6 | 3.52 | <0.001 |
| **Analytical Chemistry ( 21%)** | Other fields | 7039 | 4381 | <0.001 | 3.47 | 3.4 | 0.014 |
| **Urology & Nephrology ( 20%)** | Highly related fields | 10675 | 8539 | 0.010 | 3.68 | 3.66 | 0.186 |
| **Rehabilitation ( 20%)** | Highly related fields | 4524 | 4084 | 0.209 | 3.43 | 3.42 | 0.368 |
| **Microscopy ( 20%)** | Highly related fields | 10562 | 4837 | 0.017 | 3.67 | 3.45 | 0.108 |
| **Tropical Medicine ( 20%)** | Highly related fields | 7007 | 4532 | 0.003 | 3.53 | 3.37 | 0.104 |
| **Statistics & Probability ( 20%)** | Other fields | 13714 | 6898 | <0.001 | 3.86 | 3.7 | 0.053 |
| **Family Studies ( 20%)** | Other fields | 4770 | 2920 | 0.056 | 3.65 | 3.48 | 0.051 |
| **Toxicology ( 20%)** | Highly related fields | 7044 | 4733 | <0.001 | 3.51 | 3.44 | 0.016 |
| **Nutrition & Dietetics ( 19%)** | Highly related fields | 9204 | 6728 | 0.014 | 3.71 | 3.67 | 0.720 |
| **Medicinal & Biomolecular Chemistry ( 19%)** | Highly related fields | 4633 | 3930 | 0.007 | 3.24 | 3.18 | 0.040 |
| **Nuclear Medicine & Medical Imaging ( 19%)** | Highly related fields | 8202 | 5983 | <0.001 | 3.52 | 3.44 | <0.001 |
| **Nanoscience & Nanotechnology ( 19%)** | Other fields | 14139 | 12558 | 0.216 | 3.61 | 3.55 | 0.564 |
| **Pediatrics ( 19%)** | Highly related fields | 6967 | 4993 | <0.001 | 3.47 | 3.4 | 0.003 |
| **Pharmacology & Pharmacy ( 18%)** | Highly related fields | 6252 | 4316 | <0.001 | 3.45 | 3.37 | 0.001 |
| **Optics ( 18%)** | Other fields | 6752 | 5290 | 0.001 | 3.36 | 3.32 | 0.502 |
| **Anesthesiology ( 18%)** | Highly related fields | 6464 | 4597 | <0.001 | 3.45 | 3.41 | 0.057 |
| **Obstetrics & Reproductive Medicine ( 18%)** | Highly related fields | 7686 | 5819 | <0.001 | 3.59 | 3.51 | 0.113 |
| **Experimental Psychology ( 18%)** | Highly related fields | 7891 | 6229 | <0.001 | 3.81 | 3.77 | 0.029 |
| **Speech-Language Pathology & Audiology ( 17%)** | Highly related fields | 2806 | 3457 | 0.469 | 3.45 | 3.5 | 0.536 |
| **Biotechnology ( 17%)** | Highly related fields | 6095 | 5049 | 0.043 | 3.42 | 3.31 | 0.201 |
| **Organic Chemistry ( 16%)** | Other fields | 8620 | 6297 | <0.001 | 3.67 | 3.61 | 0.040 |
| **Nursing ( 16%)** | Highly related fields | 2337 | 1503 | <0.001 | 3.13 | 3.06 | 0.001 |
| **Environmental & Occupational Health ( 15%)** | Highly related fields | 3855 | 3745 | 0.758 | 3.33 | 3.3 | 0.758 |
| **Behavioral Science & Comparative Psychology ( 13%)** | Highly related fields | 5918 | 4397 | 0.024 | 3.79 | 3.71 | 0.649 |
| **Mycology & Parasitology ( 13%)** | Highly related fields | 4781 | 4675 | 0.621 | 3.42 | 3.47 | 0.970 |
| **General & Internal Medicine ( 13%)** | Highly related fields | 6007 | 3456 | <0.001 | 3.28 | 3.2 | 0.001 |
| **Otorhinolaryngology ( 13%)** | Highly related fields | 4290 | 3479 | 0.006 | 3.4 | 3.35 | 0.042 |
| **Sociology ( 13%)** | Other fields | 4492 | 3783 | 0.414 | 3.69 | 3.68 | 0.461 |
| **Surgery ( 13%)** | Highly related fields | 8499 | 5842 | <0.001 | 3.51 | 3.4 | 0.001 |
| **Development Studies ( 13%)** | Other fields | 2051 | 4223 | 0.631 | 3.56 | 3.57 | 0.315 |
| **Drama & Theater ( 11%)** | Other fields | 606 | 357 | 0.439 | 2.8 | 2.67 | 0.439 |
| **General Chemistry ( 11%)** | Other fields | 8799 | 4255 | <0.001 | 3.66 | 3.34 | 0.001 |
| **Dermatology & Venereal Diseases ( 11%)** | Highly related fields | 10273 | 5810 | <0.001 | 3.76 | 3.6 | 0.007 |
| **Acoustics ( 11%)** | Other fields | 3241 | 2923 | 0.090 | 3.36 | 3.42 | 0.510 |
| **Criminology ( 10%)** | Other fields | 5521 | 3764 | 0.016 | 3.63 | 3.6 | 0.414 |
| **Evolutionary Biology ( 10%)** | Other fields | 8705 | 6844 | 0.018 | 3.86 | 3.8 | 0.497 |
| **Orthopedics ( 10%)** | Highly related fields | 9151 | 5472 | <0.001 | 3.63 | 3.5 | <0.001 |
| **Environmental Sciences ( 9.7%)** | Other fields | 7291 | 4399 | 0.001 | 3.45 | 3.37 | 0.153 |
| **Distributed Computing ( 9.4%)** | Other fields | 4890 | 3192 | 0.012 | 3.13 | 3.13 | 0.510 |
| **Social Work ( 9.1%)** | Other fields | 2259 | 1587 | 0.087 | 3.26 | 3.24 | 0.159 |
| **Social Sciences Methods ( 9.1%)** | Other fields | 10839 | 5163 | 0.040 | 4.1 | 3.72 | 0.018 |
| **Artificial Intelligence & Image Processing ( 8.9%)** | Other fields | 5809 | 4172 | <0.001 | 3.45 | 3.34 | 0.034 |
| **Complementary & Alternative Medicine ( 8.6%)** | Highly related fields | 3306 | 1372 | 0.010 | 3.09 | 2.86 | 0.064 |
| **Dentistry ( 8%)** | Highly related fields | 4670 | 3403 | 0.005 | 3.46 | 3.39 | 0.319 |
| **Veterinary Sciences ( 7.9%)** | Highly related fields | 4201 | 3062 | 0.002 | 3.35 | 3.31 | 0.599 |
| **Pathology ( 7.8%)** | Highly related fields | 12052 | 9349 | 0.138 | 3.67 | 3.62 | 0.564 |
| **Plant Biology & Botany ( 7.8%)** | Other fields | 9493 | 5688 | <0.001 | 3.66 | 3.5 | <0.001 |
| **Gender Studies ( 7.7%)** | Highly related fields | 3202 | 2400 | 0.285 | 3.66 | 3.5 | 0.181 |
| **Social Psychology ( 7.7%)** | Highly related fields | 10753 | 7178 | 0.085 | 3.94 | 3.8 | 0.211 |
| **Geography ( 7.6%)** | Other fields | 4576 | 3281 | 0.055 | 3.74 | 3.68 | 0.188 |
| **Sport Sciences ( 7.5%)** | Highly related fields | 6253 | 5240 | 0.706 | 3.47 | 3.53 | 0.718 |
| **Industrial Engineering & Automation ( 7.3%)** | Other fields | 5599 | 4187 | 0.047 | 3.5 | 3.44 | 0.379 |
| **Chemical Physics ( 7.3%)** | Other fields | 13599 | 8323 | <0.001 | 3.93 | 3.79 | 0.001 |
| **Urban & Regional Planning ( 6.6%)** | Other fields | 5110 | 2308 | 0.308 | 3.69 | 3.53 | 0.662 |
| **Sport, Leisure & Tourism ( 6.5%)** | Other fields | 2884 | 3753 | 0.519 | 3.54 | 3.57 | 0.982 |
| **Entomology ( 6.1%)** | Other fields | 7467 | 3331 | 0.001 | 3.71 | 3.46 | 0.036 |
| **Human Factors ( 6%)** | Highly related fields | 5070 | 5089 | 0.489 | 3.64 | 3.58 | 0.207 |
| **Information & Library Sciences ( 6%)** | Other fields | 2095 | 1293 | 0.317 | 3.19 | 3.16 | 0.863 |
| **Numerical & Computational Mathematics ( 5.8%)** | Other fields | 9289 | 3452 | 0.064 | 3.63 | 3.54 | 0.542 |
| **Economics ( 5.7%)** | Other fields | 8352 | 4727 | 0.005 | 3.92 | 3.71 | 0.033 |
| **Economic Theory ( 5.6%)** | Other fields | 3859 | 2701 | 0.772 | 3.51 | 3.61 | 0.386 |
| **Classics ( 5.6%)** | Other fields | 361 | 538 | 0.101 | 2.85 | 2.92 | 0.101 |
| **Anatomy & Morphology ( 5.6%)** | Highly related fields | 1231 | 1583 | 0.985 | 2.81 | 3.07 | 0.015 |
| **General Clinical Medicine ( 5.5%)** | Highly related fields | 6849 | 2662 | 0.021 | 3.61 | 3.08 | 0.013 |
| **Logistics & Transportation ( 5.5%)** | Other fields | 2506 | 2803 | 0.589 | 3.19 | 3.27 | 0.428 |
| **Computer Hardware & Architecture ( 5.3%)** | Other fields | 5481 | 4368 | 0.545 | 3.23 | 3.33 | 0.201 |
| **Polymers ( 5.3%)** | Other fields | 7412 | 5524 | 0.017 | 3.6 | 3.49 | 0.506 |
| **Communication & Media Studies ( 5.2%)** | Other fields | 3023 | 2784 | 0.430 | 3.57 | 3.48 | 0.536 |
| **Fluids & Plasmas ( 5.1%)** | Other fields | 11761 | 6618 | 0.001 | 3.88 | 3.73 | 0.074 |
| **Software Engineering ( 4.8%)** | Other fields | 6139 | 4522 | 0.275 | 3.48 | 3.46 | 0.516 |
| **Electrical & Electronic Engineering ( 4.8%)** | Other fields | 3410 | 2535 | 0.081 | 3.03 | 3.04 | 0.857 |
| **Optoelectronics & Photonics ( 4.7%)** | Other fields | 4096 | 2880 | 0.017 | 3.11 | 3.11 | 0.496 |
| **Marketing ( 4.7%)** | Other fields | 5556 | 6018 | 0.884 | 3.63 | 3.64 | 0.561 |
| **Languages & Linguistics ( 4.5%)** | Other fields | 2937 | 1832 | 0.184 | 3.58 | 3.42 | 0.426 |
| **Education ( 4.5%)** | Other fields | 4463 | 2366 | <0.001 | 3.58 | 3.36 | <0.001 |
| **Networking & Telecommunications ( 4.5%)** | Other fields | 4059 | 3680 | 0.530 | 3.17 | 3.26 | 0.196 |
| **Religions & Theology ( 4.4%)** | Other fields | 911 | 463 | 0.106 | 3.06 | 2.85 | 0.210 |
| **Archaeology ( 4.4%)** | Other fields | 1927 | 1923 | 0.906 | 3.35 | 3.3 | 0.443 |
| **History of Social Sciences ( 4.3%)** | Other fields | 1613 | 1743 | 0.880 | 3.31 | 3.31 | 1.000 |
| **Applied Mathematics ( 4.3%)** | Other fields | 7299 | 4499 | 0.321 | 3.73 | 3.61 | 0.638 |
| **Mathematical Physics ( 4.2%)** | Other fields | 2455 | 2830 | 0.718 | 3.38 | 3.49 | 0.427 |
| **Agricultural Economics & Policy ( 4.1%)** | Other fields | 3582 | 4088 | 0.221 | 3.58 | 3.58 | 0.453 |
| **Food Science ( 4.1%)** | Other fields | 5487 | 3996 | 0.067 | 3.46 | 3.39 | 0.681 |
| **Anthropology ( 4.1%)** | Other fields | 3487 | 3228 | 0.828 | 3.52 | 3.61 | 0.391 |
| **Science Studies ( 3.8%)** | Other fields | 4961 | 4004 | 0.549 | 3.59 | 3.69 | 0.257 |
| **Information Systems ( 3.8%)** | Other fields | 5605 | 5798 | 0.501 | 3.61 | 3.58 | 0.563 |
| **Finance ( 3.8%)** | Other fields | 5679 | 4919 | 0.882 | 3.67 | 3.59 | 0.571 |
| **Cultural Studies ( 3.7%)** | Other fields | 814 | 795 | 0.410 | 3.13 | 3.1 | 0.680 |
| **General Physics ( 3.7%)** | Other fields | 6681 | 5581 | 0.465 | 3.62 | 3.57 | 0.819 |
| **Chemical Engineering ( 3.6%)** | Other fields | 5097 | 4079 | 0.476 | 3.55 | 3.43 | 0.504 |
| **Mechanical Engineering & Transports ( 3.5%)** | Other fields | 3801 | 2884 | 0.291 | 3.24 | 3.36 | 0.789 |
| **Ecology ( 3.5%)** | Other fields | 7263 | 7548 | 0.860 | 3.9 | 3.75 | 0.172 |
| **Operations Research ( 3.4%)** | Other fields | 4391 | 4226 | 0.645 | 3.5 | 3.6 | 0.371 |
| **Dairy & Animal Science ( 3.3%)** | Other fields | 4212 | 3062 | 0.035 | 3.4 | 3.25 | 0.452 |
| **Mining & Metallurgy ( 3.2%)** | Other fields | 5030 | 1020 | 0.605 | 3.25 | 2.76 | 0.661 |
| **Building & Construction ( 3.1%)** | Other fields | 5319 | 2994 | 0.087 | 3.21 | 3.32 | 0.603 |
| **Philosophy ( 3.1%)** | Other fields | 2367 | 1358 | 0.686 | 3.51 | 3.38 | 1.000 |
| **Political Science & Public Administration ( 3.1%)** | Other fields | 4718 | 3499 | 0.225 | 3.57 | 3.64 | 0.910 |
| **Zoology ( 3%)** | Other fields | 4056 | 2076 | 0.090 | 3.34 | 3.2 | 0.221 |
| **Ornithology ( 3%)** | Other fields | 2139 | 3168 | 0.462 | 3.37 | 3.43 | 0.674 |
| **Design Practice & Management ( 3%)** | Other fields | 5320 | 3001 | 0.498 | 3.24 | 3.28 | 0.992 |
| **Materials ( 2.9%)** | Other fields | 5960 | 3443 | 0.001 | 3.52 | 3.28 | 0.004 |
| **Environmental Engineering ( 2.9%)** | Other fields | 3537 | 4225 | 0.629 | 3.4 | 3.44 | 0.665 |
| **Inorganic & Nuclear Chemistry ( 2.8%)** | Other fields | 4382 | 5908 | 0.470 | 3.36 | 3.55 | 0.084 |
| **Geological & Geomatics Engineering ( 2.8%)** | Other fields | 10138 | 4568 | 0.023 | 3.76 | 3.45 | 0.064 |
| **Oceanography ( 2.7%)** | Other fields | 8622 | 4618 | 0.141 | 3.75 | 3.58 | 0.168 |
| **Literary Studies ( 2.5%)** | Other fields | 469 | 351 | 0.428 | 2.81 | 2.7 | 0.409 |
| **Meteorology & Atmospheric Sciences ( 2.5%)** | Other fields | 7792 | 8800 | 0.145 | 3.71 | 3.74 | 0.675 |
| **Applied Physics ( 2.4%)** | Other fields | 8254 | 6372 | 0.051 | 3.65 | 3.53 | 0.065 |
| **Econometrics ( 2.4%)** | Other fields | 17980 | 6910 | 0.302 | 3.88 | 3.97 | 0.902 |
| **Business & Management ( 2.3%)** | Other fields | 5650 | 6071 | 0.941 | 3.71 | 3.7 | 0.943 |
| **Computation Theory & Mathematics ( 2.3%)** | Other fields | 8243 | 4906 | 0.203 | 3.71 | 3.62 | 0.483 |
| **Aerospace & Aeronautics ( 2.3%)** | Other fields | 2463 | 1884 | 0.301 | 3.13 | 3.12 | 0.927 |
| **Strategic, Defence & Security Studies ( 2.2%)** | Other fields | 3126 | 2117 | 0.833 | 3.59 | 3.27 | 0.074 |
| **General Mathematics ( 2.2%)** | Other fields | 1878 | 2240 | 0.636 | 3.37 | 3.43 | 0.371 |
| **Civil Engineering ( 2.2%)** | Other fields | 3434 | 2152 | 0.424 | 3.06 | 3.17 | 0.674 |
| **Geology ( 2.2%)** | Other fields | 2839 | 3402 | 0.407 | 3.53 | 3.48 | 0.728 |
| **Energy ( 2.1%)** | Other fields | 4554 | 3374 | 0.377 | 3.23 | 3.26 | 0.979 |
| **Psychoanalysis ( 2%)** | Highly related fields | 4061 | 1330 | 0.136 | 3.67 | 3.29 | 0.136 |
| **Law ( 1.9%)** | Other fields | 1289 | 1221 | 1.000 | 3.19 | 3.22 | 0.577 |
| **Marine Biology & Hydrobiology ( 1.8%)** | Other fields | 5379 | 6136 | 0.766 | 3.61 | 3.7 | 0.718 |
| **Astronomy & Astrophysics ( 1.8%)** | Other fields | 8979 | 10830 | 0.202 | 3.79 | 3.74 | 0.522 |
| **Fisheries ( 1.8%)** | Other fields | 4535 | 3591 | 0.504 | 3.52 | 3.4 | 0.307 |
| **Legal & Forensic Medicine ( 1.5%)** | Highly related fields | 1275 | 1327 | 0.937 | 2.93 | 2.98 | 0.854 |
| **International Relations ( 1.5%)** | Other fields | 3418 | 2448 | 0.558 | 3.67 | 3.52 | 0.460 |
| **Nuclear & Particle Physics ( 1.3%)** | Other fields | 6051 | 6735 | 0.529 | 3.54 | 3.62 | 0.318 |
| **Geochemistry & Geophysics ( 1.2%)** | Other fields | 6012 | 6385 | 0.538 | 3.63 | 3.73 | 0.087 |
| **Forestry ( .96%)** | Other fields | 2201 | 2708 | 0.488 | 3.21 | 3.23 | 0.833 |
| **Physical Chemistry ( .88%)** | Other fields | 3764 | 6576 | 0.338 | 3.48 | 3.51 | 0.891 |
| **Agronomy & Agriculture ( .88%)** | Other fields | 5300 | 3948 | 0.885 | 3.45 | 3.39 | 0.970 |
| **Paleontology ( 0%)** | Other fields | . | 5643 |  | . | 3.72 |  |
| **Music ( 0%)** | Other fields | . | 446 |  | . | 2.79 |  |
| **Industrial Relations ( 0%)** | Other fields | . | 1524 |  | . | 3.34 |  |
| **Horticulture ( 0%)** | Other fields | . | 2081 |  | . | 3.14 |  |
| **History of Science, Technology & Medicine ( 0%)** | Other fields | . | 782 |  | . | 3.13 |  |
| **History ( 0%)** | Other fields | . | 532 |  | . | 2.88 |  |
| **General Psychology & Cognitive Sciences ( 0%)** | Highly related fields | . | 3365 |  | . | 3.48 |  |
| **Folklore ( 0%)** | Other fields | . | 310 |  | . | 2.61 |  |
| **Automobile Design & Engineering ( 0%)** | Other fields | . | 860 |  | . | 2.69 |  |
| **Art Practice, History & Theory ( 0%)** | Other fields | . | 210 |  | . | 2.47 |  |
| **Architecture ( 0%)** | Other fields | . | 259 |  | . | 2.47 |  |
| **Accounting ( 0%)** | Other fields | . | 5445 |  | . | 3.54 |  |

**Supplementary Table 2.3 : Career-long impact, Funding time current funding citation counts and composite citation indices for each subfield (ordered by percentage funded)**

| **Top-cited US-based researchers: Subfield (perc. funded)** | **Classification** | **Citations for funded, median** | **Citations for non-funded, median** | **p-value** | **Composite index for funded, median** | **Composite index for non-funded, median** | **p-value** |
| --- | --- | --- | --- | --- | --- | --- | --- |
| **Geriatrics ( 31%)** | Highly related fields | 10839 | 9133 | 0.217 | 3.57 | 3.64 | 0.845 |
| **Bioinformatics ( 30%)** | Highly related fields | 14836 | 9462 | 0.007 | 3.69 | 3.54 | 0.118 |
| **Developmental Biology ( 29%)** | Highly related fields | 13168 | 12541 | 0.037 | 3.79 | 3.82 | 0.104 |
| **Substance Abuse ( 23%)** | Highly related fields | 8603 | 7416 | 0.121 | 3.68 | 3.73 | 0.211 |
| **Medical Informatics ( 23%)** | Highly related fields | 4194 | 4026 | 0.202 | 3.5 | 3.3 | 0.156 |
| **Virology ( 22%)** | Highly related fields | 10016 | 9720 | 0.304 | 3.6 | 3.6 | 0.955 |
| **Biomedical Engineering ( 22%)** | Highly related fields | 6113 | 5174 | 0.060 | 3.31 | 3.37 | 0.539 |
| **Neurology & Neurosurgery ( 21%)** | Highly related fields | 11459 | 10579 | 0.007 | 3.77 | 3.78 | 0.638 |
| **Immunology ( 20%)** | Highly related fields | 15484 | 13119 | 0.001 | 3.88 | 3.77 | 0.005 |
| **Gerontology ( 20%)** | Highly related fields | 7573 | 6230 | 0.143 | 3.75 | 3.62 | 0.140 |
| **Applied Ethics ( 20%)** | Highly related fields | 6906 | 3197 | 0.002 | 3.62 | 3.56 | 0.418 |
| **Public Health ( 19%)** | Highly related fields | 8145 | 7301 | 0.009 | 3.71 | 3.65 | 0.066 |
| **Demography ( 19%)** | Highly related fields | 3532 | 3282 | 0.949 | 3.53 | 3.59 | 0.423 |
| **Oncology & Carcinogenesis ( 18%)** | Highly related fields | 14737 | 13030 | <0.001 | 3.65 | 3.68 | 0.053 |
| **Gastroenterology & Hepatology ( 17%)** | Highly related fields | 12537 | 9605 | 0.049 | 3.72 | 3.7 | 0.327 |
| **Allergy ( 16%)** | Highly related fields | 9374 | 8456 | 0.491 | 3.75 | 3.75 | 0.589 |
| **Biophysics ( 16%)** | Highly related fields | 5430 | 6020 | 0.717 | 3.48 | 3.52 | 0.559 |
| **Developmental & Child Psychology ( 16%)** | Highly related fields | 9033 | 6918 | 0.008 | 3.86 | 3.74 | 0.002 |
| **Health Policy & Services ( 16%)** | Highly related fields | 6978 | 7567 | 0.811 | 3.58 | 3.61 | 0.966 |
| **Physiology ( 16%)** | Highly related fields | 5972 | 6492 | 0.657 | 3.73 | 3.72 | 0.930 |
| **Arthritis & Rheumatology ( 16%)** | Highly related fields | 12811 | 13497 | 0.640 | 3.76 | 3.79 | 0.402 |
| **Epidemiology ( 15%)** | Highly related fields | 13964 | 15301 | 0.926 | 3.76 | 3.84 | 0.666 |
| **Psychiatry ( 15%)** | Highly related fields | 15417 | 11068 | <0.001 | 3.9 | 3.82 | 0.053 |
| **Genetics & Heredity ( 15%)** | Highly related fields | 15832 | 11983 | 0.011 | 3.64 | 3.67 | 0.431 |
| **Emergency & Critical Care Medicine ( 15%)** | Highly related fields | 9522 | 6216 | 0.001 | 3.55 | 3.47 | 0.151 |
| **Endocrinology & Metabolism ( 15%)** | Highly related fields | 11919 | 11565 | 0.390 | 3.77 | 3.78 | 0.536 |
| **Statistics & Probability ( 14%)** | Other fields | 10685 | 7455 | 0.033 | 3.8 | 3.7 | 0.347 |
| **Ophthalmology & Optometry ( 14%)** | Highly related fields | 7668 | 5621 | <0.001 | 3.6 | 3.53 | 0.007 |
| **Microbiology ( 14%)** | Highly related fields | 8839 | 8261 | 0.038 | 3.66 | 3.65 | 0.351 |
| **Cardiovascular System & Hematology ( 14%)** | Highly related fields | 15934 | 13421 | 0.006 | 3.76 | 3.76 | 0.847 |
| **Respiratory System ( 13%)** | Highly related fields | 9910 | 10639 | 0.687 | 3.61 | 3.69 | 0.339 |
| **Clinical Psychology ( 13%)** | Highly related fields | 10228 | 7700 | 0.204 | 3.82 | 3.74 | 0.311 |
| **Nuclear Medicine & Medical Imaging ( 13%)** | Highly related fields | 8077 | 6067 | <0.001 | 3.51 | 3.45 | 0.032 |
| **Analytical Chemistry ( 13%)** | Other fields | 7187 | 4504 | <0.001 | 3.39 | 3.42 | 0.754 |
| **Medicinal & Biomolecular Chemistry ( 12%)** | Highly related fields | 4694 | 3986 | 0.027 | 3.19 | 3.19 | 0.264 |
| **Nanoscience & Nanotechnology ( 12%)** | Other fields | 12566 | 12694 | 0.607 | 3.65 | 3.55 | 0.572 |
| **Urology & Nephrology ( 12%)** | Highly related fields | 11108 | 8675 | 0.026 | 3.69 | 3.66 | 0.321 |
| **Optics ( 12%)** | Other fields | 6211 | 5527 | 0.024 | 3.3 | 3.34 | 0.862 |
| **Biochemistry & Molecular Biology ( 12%)** | Highly related fields | 11189 | 9351 | <0.001 | 3.79 | 3.76 | 0.143 |
| **Nutrition & Dietetics ( 11%)** | Highly related fields | 9888 | 6824 | 0.044 | 3.7 | 3.67 | 0.847 |
| **Experimental Psychology ( 11%)** | Highly related fields | 9469 | 6183 | <0.001 | 3.91 | 3.76 | 0.002 |
| **Rehabilitation ( 11%)** | Highly related fields | 5546 | 4032 | 0.028 | 3.45 | 3.42 | 0.206 |
| **Toxicology ( 11%)** | Highly related fields | 7828 | 5089 | <0.001 | 3.47 | 3.45 | 0.113 |
| **Drama & Theater ( 11%)** | Other fields | 606 | 357 | 0.439 | 2.8 | 2.67 | 0.439 |
| **Obstetrics & Reproductive Medicine ( 11%)** | Highly related fields | 7559 | 5916 | 0.022 | 3.59 | 3.52 | 0.237 |
| **Pediatrics ( 11%)** | Highly related fields | 6146 | 5109 | <0.001 | 3.47 | 3.41 | 0.098 |
| **Anesthesiology ( 11%)** | Highly related fields | 6764 | 4696 | 0.001 | 3.48 | 3.41 | 0.120 |
| **Pharmacology & Pharmacy ( 10%)** | Highly related fields | 5538 | 4508 | 0.002 | 3.39 | 3.38 | 0.287 |
| **Speech-Language Pathology & Audiology ( 9.9%)** | Highly related fields | 2857 | 3380 | 0.459 | 3.38 | 3.49 | 0.547 |
| **Organic Chemistry ( 9.8%)** | Other fields | 8436 | 6344 | <0.001 | 3.7 | 3.61 | 0.067 |
| **Environmental & Occupational Health ( 9.8%)** | Highly related fields | 3363 | 3847 | 0.545 | 3.31 | 3.3 | 0.772 |
| **Family Studies ( 8.9%)** | Other fields | 4688 | 2976 | 0.381 | 3.54 | 3.5 | 0.943 |
| **Development Studies ( 8.3%)** | Other fields | 3741 | 3565 | 0.917 | 3.48 | 3.57 | 0.296 |
| **Mycology & Parasitology ( 8.2%)** | Highly related fields | 5086 | 4661 | 0.593 | 3.45 | 3.46 | 0.960 |
| **Nursing ( 7.9%)** | Highly related fields | 2865 | 1561 | <0.001 | 3.17 | 3.07 | 0.002 |
| **Surgery ( 7.9%)** | Highly related fields | 8494 | 6014 | <0.001 | 3.46 | 3.41 | 0.232 |
| **Tropical Medicine ( 7.8%)** | Highly related fields | 7167 | 4979 | 0.046 | 3.55 | 3.39 | 0.173 |
| **General Chemistry ( 7.8%)** | Other fields | 8763 | 4364 | <0.001 | 3.73 | 3.34 | 0.005 |
| **Complementary & Alternative Medicine ( 7.4%)** | Highly related fields | 3645 | 1421 | 0.010 | 3.13 | 2.86 | 0.043 |
| **General & Internal Medicine ( 7.1%)** | Highly related fields | 6242 | 3586 | <0.001 | 3.32 | 3.2 | 0.015 |
| **Biotechnology ( 7%)** | Highly related fields | 6265 | 5049 | 0.166 | 3.5 | 3.31 | 0.171 |
| **Microscopy ( 6.7%)** | Highly related fields | 10312 | 5644 | 0.280 | 3.78 | 3.46 | 0.135 |
| **Acoustics ( 6.5%)** | Other fields | 3213 | 2976 | 0.284 | 3.34 | 3.42 | 0.441 |
| **Sociology ( 6.4%)** | Other fields | 4332 | 3815 | 0.527 | 3.7 | 3.68 | 0.790 |
| **Otorhinolaryngology ( 6.3%)** | Highly related fields | 4490 | 3530 | 0.024 | 3.37 | 3.36 | 0.213 |
| **Behavioral Science & Comparative Psychology ( 6.3%)** | Highly related fields | 6282 | 4557 | 0.137 | 3.79 | 3.71 | 0.867 |
| **Orthopedics ( 5.9%)** | Highly related fields | 9267 | 5580 | <0.001 | 3.65 | 3.5 | 0.003 |
| **Sport Sciences ( 5.9%)** | Highly related fields | 7114 | 5198 | 0.404 | 3.45 | 3.53 | 0.788 |
| **Social Work ( 5.7%)** | Other fields | 2382 | 1638 | 0.308 | 3.28 | 3.24 | 0.245 |
| **Economic Theory ( 5.6%)** | Other fields | 3859 | 2701 | 0.772 | 3.51 | 3.61 | 0.386 |
| **Evolutionary Biology ( 5.5%)** | Other fields | 9058 | 6924 | 0.017 | 3.89 | 3.8 | 0.151 |
| **Dermatology & Venereal Diseases ( 5.5%)** | Highly related fields | 8804 | 5984 | 0.008 | 3.67 | 3.61 | 0.211 |
| **Artificial Intelligence & Image Processing ( 5.5%)** | Other fields | 4867 | 4293 | 0.102 | 3.44 | 3.35 | 0.224 |
| **Environmental Sciences ( 5.5%)** | Other fields | 7291 | 4504 | 0.015 | 3.44 | 3.37 | 0.340 |
| **Plant Biology & Botany ( 5.2%)** | Other fields | 9646 | 5794 | <0.001 | 3.72 | 3.51 | 0.001 |
| **Pathology ( 5.1%)** | Highly related fields | 10145 | 9393 | 0.891 | 3.55 | 3.63 | 0.584 |
| **Chemical Physics ( 4.8%)** | Other fields | 13002 | 8433 | 0.011 | 3.93 | 3.79 | 0.066 |
| **Industrial Engineering & Automation ( 4.8%)** | Other fields | 5400 | 4234 | 0.149 | 3.52 | 3.44 | 0.337 |
| **Entomology ( 4.8%)** | Other fields | 7706 | 3312 | <0.001 | 3.76 | 3.46 | 0.006 |
| **Criminology ( 4.6%)** | Other fields | 4788 | 3799 | 0.089 | 3.61 | 3.6 | 0.582 |
| **Religions & Theology ( 4.4%)** | Other fields | 911 | 463 | 0.106 | 3.06 | 2.85 | 0.210 |
| **Sport, Leisure & Tourism ( 4.3%)** | Other fields | 3629 | 3732 | 0.829 | 3.61 | 3.57 | 0.829 |
| **Distributed Computing ( 4.3%)** | Other fields | 6213 | 3356 | 0.012 | 3.23 | 3.13 | 0.836 |
| **Dentistry ( 4.3%)** | Highly related fields | 4451 | 3419 | 0.087 | 3.45 | 3.39 | 0.614 |
| **Social Sciences Methods ( 3.9%)** | Other fields | 30480 | 5254 | 0.073 | 4.39 | 3.72 | 0.040 |
| **Science Studies ( 3.8%)** | Other fields | 4961 | 4004 | 0.549 | 3.59 | 3.69 | 0.257 |
| **Logistics & Transportation ( 3.8%)** | Other fields | 2116 | 2775 | 0.319 | 3.16 | 3.28 | 0.024 |
| **Veterinary Sciences ( 3.8%)** | Highly related fields | 5021 | 3105 | 0.003 | 3.4 | 3.31 | 0.149 |
| **Social Psychology ( 3.7%)** | Highly related fields | 11577 | 7178 | 0.086 | 3.95 | 3.8 | 0.083 |
| **Anatomy & Morphology ( 3.7%)** | Highly related fields | 3295 | 1505 | 0.156 | 2.95 | 3.06 | 0.272 |
| **Applied Mathematics ( 3.6%)** | Other fields | 6279 | 4521 | 0.745 | 3.46 | 3.61 | 0.789 |
| **Polymers ( 3.6%)** | Other fields | 7233 | 5559 | 0.102 | 3.57 | 3.49 | 0.762 |
| **Information & Library Sciences ( 3.4%)** | Other fields | 1095 | 1299 | 0.631 | 3.12 | 3.16 | 0.893 |
| **Communication & Media Studies ( 3.2%)** | Other fields | 2875 | 2784 | 0.325 | 3.54 | 3.49 | 0.479 |
| **Mining & Metallurgy ( 3.2%)** | Other fields | 5030 | 1020 | 0.605 | 3.25 | 2.76 | 0.661 |
| **General Clinical Medicine ( 3.1%)** | Highly related fields | 10192 | 2712 | 0.018 | 3.63 | 3.08 | 0.013 |
| **Marketing ( 3.1%)** | Other fields | 6267 | 5797 | 0.606 | 3.72 | 3.64 | 0.714 |
| **Economics ( 3.1%)** | Other fields | 9915 | 4739 | 0.001 | 3.96 | 3.71 | 0.009 |
| **Software Engineering ( 3.1%)** | Other fields | 6139 | 4522 | 0.308 | 3.59 | 3.46 | 0.275 |
| **Ornithology ( 3%)** | Other fields | 2139 | 3168 | 0.462 | 3.37 | 3.43 | 0.674 |
| **Finance ( 3%)** | Other fields | 6458 | 4901 | 0.700 | 3.69 | 3.59 | 0.418 |
| **Human Factors ( 3%)** | Highly related fields | 4766 | 5142 | 0.441 | 3.67 | 3.58 | 0.155 |
| **Agricultural Economics & Policy ( 2.7%)** | Other fields | 3180 | 4084 | 0.265 | 3.58 | 3.58 | 1.000 |
| **Computer Hardware & Architecture ( 2.6%)** | Other fields | 4513 | 4437 | 0.696 | 3.26 | 3.32 | 0.990 |
| **Networking & Telecommunications ( 2.6%)** | Other fields | 4203 | 3679 | 0.306 | 3.16 | 3.25 | 0.629 |
| **Political Science & Public Administration ( 2.6%)** | Other fields | 5295 | 3497 | 0.089 | 3.67 | 3.64 | 0.661 |
| **Food Science ( 2.6%)** | Other fields | 4023 | 4018 | 0.558 | 3.23 | 3.41 | 0.532 |
| **Education ( 2.6%)** | Other fields | 4463 | 2375 | <0.001 | 3.64 | 3.36 | 0.001 |
| **Dairy & Animal Science ( 2.4%)** | Other fields | 3802 | 3067 | 0.299 | 3.35 | 3.25 | 0.988 |
| **Econometrics ( 2.4%)** | Other fields | 17980 | 6910 | 0.302 | 3.88 | 3.97 | 0.902 |
| **Mechanical Engineering & Transports ( 2.3%)** | Other fields | 3934 | 2911 | 0.739 | 3.15 | 3.36 | 0.127 |
| **Strategic, Defence & Security Studies ( 2.2%)** | Other fields | 3126 | 2117 | 0.773 | 3.57 | 3.27 | 0.103 |
| **Electrical & Electronic Engineering ( 2.2%)** | Other fields | 3080 | 2546 | 0.350 | 2.89 | 3.04 | 0.279 |
| **Geography ( 2.2%)** | Other fields | 3516 | 3345 | 0.872 | 3.6 | 3.68 | 0.453 |
| **Chemical Engineering ( 2.1%)** | Other fields | 3832 | 4079 | 0.802 | 3.37 | 3.43 | 0.951 |
| **General Physics ( 2%)** | Other fields | 7034 | 5581 | 0.232 | 3.62 | 3.57 | 0.744 |
| **Optoelectronics & Photonics ( 2%)** | Other fields | 3883 | 2901 | 0.278 | 3.15 | 3.11 | 0.506 |
| **Psychoanalysis ( 2%)** | Highly related fields | 4061 | 1330 | 0.136 | 3.67 | 3.29 | 0.136 |
| **Design Practice & Management ( 2%)** | Other fields | 5309 | 3034 | 0.787 | 3.37 | 3.27 | 0.588 |
| **Operations Research ( 2%)** | Other fields | 4628 | 4221 | 0.708 | 3.68 | 3.6 | 0.785 |
| **Environmental Engineering ( 2%)** | Other fields | 4128 | 4209 | 0.916 | 3.4 | 3.44 | 0.986 |
| **Information Systems ( 1.9%)** | Other fields | 9535 | 5645 | 0.137 | 3.69 | 3.58 | 0.321 |
| **Literary Studies ( 1.9%)** | Other fields | 591 | 349 | 0.355 | 2.92 | 2.7 | 0.349 |
| **Cultural Studies ( 1.9%)** | Other fields | 1102 | 794 | 0.585 | 3.29 | 3.1 | 0.352 |
| **Languages & Linguistics ( 1.8%)** | Other fields | 2577 | 1838 | 0.408 | 3.62 | 3.43 | 0.194 |
| **Fisheries ( 1.8%)** | Other fields | 6517 | 3591 | 0.265 | 3.59 | 3.4 | 0.174 |
| **Archaeology ( 1.8%)** | Other fields | 2125 | 1922 | 0.796 | 3.29 | 3.31 | 0.666 |
| **Computation Theory & Mathematics ( 1.7%)** | Other fields | 3448 | 4911 | 0.699 | 3.68 | 3.63 | 0.991 |
| **Numerical & Computational Mathematics ( 1.7%)** | Other fields | 9914 | 3498 | 0.062 | 3.95 | 3.54 | 0.085 |
| **Urban & Regional Planning ( 1.6%)** | Other fields | 3463 | 2360 | 0.570 | 3.37 | 3.54 | 0.281 |
| **Anthropology ( 1.6%)** | Other fields | 4446 | 3247 | 0.368 | 3.61 | 3.61 | 0.704 |
| **Materials ( 1.6%)** | Other fields | 7834 | 3448 | 0.001 | 3.67 | 3.28 | 0.004 |
| **Fluids & Plasmas ( 1.6%)** | Other fields | 13640 | 6643 | 0.013 | 3.91 | 3.73 | 0.080 |
| **General Mathematics ( 1.6%)** | Other fields | 1729 | 2247 | 0.161 | 3.37 | 3.43 | 0.084 |
| **Building & Construction ( 1.6%)** | Other fields | 5601 | 3016 | 0.120 | 3.46 | 3.32 | 0.939 |
| **Philosophy ( 1.5%)** | Other fields | 5532 | 1353 | 0.020 | 4.02 | 3.38 | 0.023 |
| **Meteorology & Atmospheric Sciences ( 1.5%)** | Other fields | 7974 | 8765 | 0.550 | 3.74 | 3.74 | 0.956 |
| **Aerospace & Aeronautics ( 1.5%)** | Other fields | 2674 | 1884 | 0.500 | 3.07 | 3.13 | 0.681 |
| **Energy ( 1.4%)** | Other fields | 4706 | 3378 | 0.410 | 3.13 | 3.26 | 0.586 |
| **Geological & Geomatics Engineering ( 1.4%)** | Other fields | 12243 | 4575 | 0.028 | 3.84 | 3.45 | 0.176 |
| **Ecology ( 1.3%)** | Other fields | 6631 | 7570 | 0.697 | 4.02 | 3.75 | 0.195 |
| **Oceanography ( 1.1%)** | Other fields | 10377 | 4641 | 0.058 | 3.78 | 3.58 | 0.319 |
| **Business & Management ( .95%)** | Other fields | 7503 | 6052 | 0.388 | 3.86 | 3.7 | 0.414 |
| **Law ( .94%)** | Other fields | 1025 | 1223 | 0.524 | 3.23 | 3.22 | 0.961 |
| **Applied Physics ( .94%)** | Other fields | 8020 | 6408 | 0.484 | 3.64 | 3.53 | 0.151 |
| **Marine Biology & Hydrobiology ( .91%)** | Other fields | 11873 | 6074 | 0.219 | 4.02 | 3.69 | 0.221 |
| **Physical Chemistry ( .88%)** | Other fields | 3764 | 6576 | 0.338 | 3.48 | 3.51 | 0.891 |
| **Nuclear & Particle Physics ( .78%)** | Other fields | 7802 | 6724 | 0.538 | 3.63 | 3.62 | 0.871 |
| **Astronomy & Astrophysics ( .76%)** | Other fields | 8250 | 10762 | 0.465 | 3.88 | 3.74 | 0.217 |
| **Civil Engineering ( .73%)** | Other fields | 3434 | 2152 | 0.445 | 3.39 | 3.16 | 0.865 |
| **Geochemistry & Geophysics ( .71%)** | Other fields | 5389 | 6405 | 0.404 | 3.54 | 3.73 | 0.031 |
| **Agronomy & Agriculture ( .66%)** | Other fields | 7813 | 3940 | 0.540 | 3.69 | 3.39 | 0.475 |
| **Inorganic & Nuclear Chemistry ( .63%)** | Other fields | 4341 | 5884 | 0.418 | 3.4 | 3.55 | 0.388 |
| **Forestry ( .48%)** | Other fields | 1063 | 2711 | 0.136 | 3.08 | 3.23 | 0.353 |
| **Zoology ( 0%)** | Other fields | . | 2100 |  | . | 3.21 |  |
| **Paleontology ( 0%)** | Other fields | . | 5643 |  | . | 3.72 |  |
| **Music ( 0%)** | Other fields | . | 446 |  | . | 2.79 |  |
| **Mathematical Physics ( 0%)** | Other fields | . | 2774 |  | . | 3.49 |  |
| **Legal & Forensic Medicine ( 0%)** | Highly related fields | . | 1326 |  | . | 2.97 |  |
| **International Relations ( 0%)** | Other fields | . | 2518 |  | . | 3.52 |  |
| **Industrial Relations ( 0%)** | Other fields | . | 1524 |  | . | 3.34 |  |
| **Horticulture ( 0%)** | Other fields | . | 2081 |  | . | 3.14 |  |
| **History of Social Sciences ( 0%)** | Other fields | . | 1740 |  | . | 3.31 |  |
| **History of Science, Technology & Medicine ( 0%)** | Other fields | . | 782 |  | . | 3.13 |  |
| **History ( 0%)** | Other fields | . | 532 |  | . | 2.88 |  |
| **Geology ( 0%)** | Other fields | . | 3402 |  | . | 3.48 |  |
| **General Psychology & Cognitive Sciences ( 0%)** | Highly related fields | . | 3365 |  | . | 3.48 |  |
| **Gender Studies ( 0%)** | Highly related fields | . | 2515 |  | . | 3.51 |  |
| **Folklore ( 0%)** | Other fields | . | 310 |  | . | 2.61 |  |
| **Classics ( 0%)** | Other fields | . | 536 |  | . | 2.92 |  |
| **Automobile Design & Engineering ( 0%)** | Other fields | . | 860 |  | . | 2.69 |  |
| **Art Practice, History & Theory ( 0%)** | Other fields | . | 210 |  | . | 2.47 |  |
| **Architecture ( 0%)** | Other fields | . | 259 |  | . | 2.47 |  |
| **Accounting ( 0%)** | Other fields | . | 5445 |  | . | 3.54 |  |

**Supplementary Table 2.4 : Recent year impact, Funding time any funding citation counts and composite citation indices for each subfield (ordered by percentage funded)**

| **Top-cited US-based researchers: Subfield (perc. funded)** | **Classification** | **Citations for funded, median** | **Citations for non-funded, median** | **p-value** | **Composite index for funded, median** | **Composite index for non-funded, median** | **p-value** |
| --- | --- | --- | --- | --- | --- | --- | --- |
| **Geriatrics ( 88%)** | Highly related fields | 1567 | 1003 | 0.015 | 3.11 | 2.97 | 0.033 |
| **Gerontology ( 87%)** | Highly related fields | 1210 | 536 | 0.002 | 3.16 | 2.98 | 0.023 |
| **Substance Abuse ( 86%)** | Highly related fields | 1155 | 919 | 0.048 | 3.09 | 3.06 | 0.472 |
| **Developmental Biology ( 86%)** | Highly related fields | 1826 | 2005 | 0.102 | 3.15 | 3.07 | <0.001 |
| **Endocrinology & Metabolism ( 83%)** | Highly related fields | 1579 | 1290 | 0.003 | 3.16 | 3.05 | <0.001 |
| **Immunology ( 83%)** | Highly related fields | 1825 | 2049 | 0.271 | 3.12 | 2.99 | <0.001 |
| **Neurology & Neurosurgery ( 82%)** | Highly related fields | 1561 | 1311 | <0.001 | 3.14 | 3.03 | <0.001 |
| **Biochemistry & Molecular Biology ( 81%)** | Highly related fields | 985 | 756 | <0.001 | 2.98 | 2.84 | <0.001 |
| **Virology ( 81%)** | Highly related fields | 1204 | 1158 | 0.486 | 2.86 | 2.82 | 0.018 |
| **Psychiatry ( 81%)** | Highly related fields | 1724 | 1174 | <0.001 | 3.2 | 3.1 | 0.023 |
| **Genetics & Heredity ( 79%)** | Highly related fields | 1477 | 1277 | 0.097 | 2.85 | 2.82 | 0.103 |
| **Allergy ( 75%)** | Highly related fields | 1704 | 1215 | 0.019 | 3.19 | 3.02 | 0.059 |
| **Biophysics ( 74%)** | Highly related fields | 715 | 479 | 0.001 | 2.83 | 2.68 | <0.001 |
| **Epidemiology ( 74%)** | Highly related fields | 2056 | 1906 | 0.477 | 3.14 | 3.18 | 0.688 |
| **Biomedical Engineering ( 74%)** | Highly related fields | 999 | 713 | <0.001 | 2.86 | 2.73 | <0.001 |
| **Arthritis & Rheumatology ( 73%)** | Highly related fields | 2153 | 1718 | 0.004 | 3.18 | 3.08 | 0.026 |
| **Oncology & Carcinogenesis ( 72%)** | Highly related fields | 2165 | 1827 | <0.001 | 3.03 | 2.93 | <0.001 |
| **Developmental & Child Psychology ( 72%)** | Highly related fields | 1132 | 761 | <0.001 | 3.2 | 3.1 | 0.007 |
| **Medical Informatics ( 71%)** | Highly related fields | 703 | 586 | 0.088 | 2.71 | 2.7 | 0.625 |
| **Physiology ( 71%)** | Highly related fields | 763 | 517 | <0.001 | 2.96 | 2.94 | 0.404 |
| **Public Health ( 71%)** | Highly related fields | 1243 | 1004 | 0.001 | 3.13 | 3.08 | 0.046 |
| **Gastroenterology & Hepatology ( 69%)** | Highly related fields | 1415 | 1151 | <0.001 | 3 | 2.91 | <0.001 |
| **Urology & Nephrology ( 67%)** | Highly related fields | 1222 | 1043 | 0.001 | 2.95 | 2.81 | <0.001 |
| **Respiratory System ( 67%)** | Highly related fields | 1550 | 1363 | 0.001 | 2.97 | 2.93 | 0.031 |
| **Pediatrics ( 67%)** | Highly related fields | 831 | 598 | <0.001 | 2.72 | 2.64 | <0.001 |
| **Bioinformatics ( 67%)** | Highly related fields | 2402 | 1840 | 0.089 | 3.03 | 2.98 | 0.237 |
| **Cardiovascular System & Hematology ( 66%)** | Highly related fields | 1799 | 1762 | 0.037 | 3.06 | 2.98 | <0.001 |
| **Health Policy & Services ( 64%)** | Highly related fields | 1131 | 853 | 0.021 | 3.01 | 2.92 | 0.005 |
| **Experimental Psychology ( 64%)** | Highly related fields | 981 | 724 | <0.001 | 3.21 | 3.15 | 0.025 |
| **Emergency & Critical Care Medicine ( 63%)** | Highly related fields | 1313 | 794 | <0.001 | 2.98 | 2.76 | <0.001 |
| **Clinical Psychology ( 63%)** | Highly related fields | 1270 | 895 | <0.001 | 3.17 | 3.14 | 0.093 |
| **Ophthalmology & Optometry ( 62%)** | Highly related fields | 978 | 688 | <0.001 | 2.87 | 2.73 | <0.001 |
| **Family Studies ( 62%)** | Other fields | 529 | 310 | 0.007 | 3.04 | 2.94 | 0.323 |
| **Speech-Language Pathology & Audiology ( 61%)** | Highly related fields | 465 | 597 | 0.273 | 2.87 | 2.9 | 0.834 |
| **Rehabilitation ( 61%)** | Highly related fields | 784 | 677 | 0.144 | 2.84 | 2.84 | 0.706 |
| **Obstetrics & Reproductive Medicine ( 60%)** | Highly related fields | 874 | 689 | <0.001 | 2.84 | 2.74 | <0.001 |
| **Toxicology ( 60%)** | Highly related fields | 1051 | 674 | <0.001 | 2.86 | 2.76 | <0.001 |
| **Environmental & Occupational Health ( 59%)** | Highly related fields | 565 | 491 | 0.216 | 2.65 | 2.58 | 0.107 |
| **Nutrition & Dietetics ( 59%)** | Highly related fields | 1268 | 921 | <0.001 | 3.12 | 3.01 | 0.006 |
| **Microbiology ( 59%)** | Highly related fields | 1339 | 1275 | 0.169 | 3.02 | 2.96 | <0.001 |
| **Nursing ( 59%)** | Highly related fields | 345 | 211 | <0.001 | 2.49 | 2.47 | 0.085 |
| **Demography ( 58%)** | Highly related fields | 533 | 339 | 0.033 | 3.06 | 2.99 | 0.642 |
| **Analytical Chemistry ( 58%)** | Other fields | 881 | 626 | <0.001 | 2.81 | 2.77 | 0.085 |
| **Nuclear Medicine & Medical Imaging ( 57%)** | Highly related fields | 960 | 727 | <0.001 | 2.75 | 2.65 | <0.001 |
| **Organic Chemistry ( 56%)** | Other fields | 1023 | 848 | 0.005 | 2.98 | 2.83 | <0.001 |
| **Pharmacology & Pharmacy ( 53%)** | Highly related fields | 718 | 519 | <0.001 | 2.75 | 2.62 | <0.001 |
| **Medicinal & Biomolecular Chemistry ( 51%)** | Highly related fields | 801 | 591 | <0.001 | 2.68 | 2.58 | <0.001 |
| **Complementary & Alternative Medicine ( 50%)** | Highly related fields | 520 | 242 | <0.001 | 2.65 | 2.53 | 0.273 |
| **Behavioral Science & Comparative Psychology ( 49%)** | Highly related fields | 727 | 614 | 0.085 | 3.11 | 3.07 | 0.169 |
| **Anesthesiology ( 48%)** | Highly related fields | 743 | 602 | <0.001 | 2.77 | 2.64 | <0.001 |
| **Dentistry ( 48%)** | Highly related fields | 663 | 423 | <0.001 | 2.73 | 2.67 | 0.002 |
| **Otorhinolaryngology ( 47%)** | Highly related fields | 528 | 507 | 0.442 | 2.66 | 2.57 | 0.005 |
| **Tropical Medicine ( 46%)** | Highly related fields | 908 | 634 | 0.001 | 2.77 | 2.67 | 0.001 |
| **Applied Ethics ( 46%)** | Highly related fields | 686 | 489 | 0.081 | 3.04 | 2.92 | 0.461 |
| **General & Internal Medicine ( 46%)** | Highly related fields | 776 | 536 | <0.001 | 2.73 | 2.49 | <0.001 |
| **Statistics & Probability ( 45%)** | Other fields | 1447 | 758 | <0.001 | 3.18 | 3.09 | 0.093 |
| **Surgery ( 44%)** | Highly related fields | 1034 | 686 | <0.001 | 2.75 | 2.61 | <0.001 |
| **Mycology & Parasitology ( 44%)** | Highly related fields | 695 | 778 | 0.321 | 2.79 | 2.82 | 0.181 |
| **Dermatology & Venereal Diseases ( 42%)** | Highly related fields | 1012 | 673 | <0.001 | 2.87 | 2.73 | <0.001 |
| **History of Social Sciences ( 41%)** | Other fields | 184 | 262 | 0.526 | 2.75 | 2.76 | 0.696 |
| **Biotechnology ( 41%)** | Highly related fields | 1046 | 945 | 0.343 | 2.94 | 2.82 | 0.112 |
| **General Chemistry ( 40%)** | Other fields | 806 | 728 | 0.052 | 2.83 | 2.64 | <0.001 |
| **Sport Sciences ( 38%)** | Highly related fields | 1102 | 898 | 0.020 | 3.01 | 3.02 | 0.710 |
| **Social Psychology ( 38%)** | Highly related fields | 1363 | 820 | <0.001 | 3.39 | 3.21 | <0.001 |
| **Acoustics ( 37%)** | Other fields | 505 | 436 | 0.072 | 2.77 | 2.77 | 0.990 |
| **Optics ( 32%)** | Other fields | 987 | 907 | 0.100 | 2.77 | 2.76 | 0.130 |
| **Orthopedics ( 31%)** | Highly related fields | 1159 | 810 | <0.001 | 2.95 | 2.83 | <0.001 |
| **Chemical Physics ( 30%)** | Other fields | 1629 | 1075 | <0.001 | 3.18 | 3.08 | 0.002 |
| **Evolutionary Biology ( 30%)** | Other fields | 1011 | 916 | 0.003 | 3.24 | 3.13 | 0.005 |
| **Nanoscience & Nanotechnology ( 29%)** | Other fields | 2493 | 2400 | 0.121 | 3.22 | 3.12 | 0.001 |
| **Sociology ( 28%)** | Other fields | 467 | 415 | 0.132 | 3.05 | 3.11 | 0.471 |
| **Pathology ( 28%)** | Highly related fields | 1232 | 1032 | 0.030 | 2.89 | 2.8 | 0.105 |
| **Plant Biology & Botany ( 27%)** | Other fields | 1183 | 863 | <0.001 | 3.02 | 2.94 | 0.001 |
| **General Clinical Medicine ( 27%)** | Highly related fields | 571 | 276 | <0.001 | 2.57 | 2.3 | 0.008 |
| **Veterinary Sciences ( 27%)** | Highly related fields | 532 | 377 | <0.001 | 2.59 | 2.55 | 0.027 |
| **Environmental Sciences ( 26%)** | Other fields | 1648 | 1154 | 0.002 | 3.16 | 3.08 | 0.089 |
| **Social Sciences Methods ( 25%)** | Other fields | 1099 | 719 | 0.003 | 3.47 | 3.15 | 0.007 |
| **Criminology ( 25%)** | Other fields | 805 | 474 | <0.001 | 3.14 | 3.04 | 0.026 |
| **Microscopy ( 24%)** | Highly related fields | 1146 | 603 | 0.027 | 2.9 | 2.74 | 0.141 |
| **Inorganic & Nuclear Chemistry ( 23%)** | Other fields | 923 | 690 | 0.142 | 2.89 | 2.76 | 0.011 |
| **Social Work ( 23%)** | Other fields | 468 | 271 | 0.001 | 2.88 | 2.71 | 0.004 |
| **Polymers ( 21%)** | Other fields | 1353 | 791 | <0.001 | 2.99 | 2.89 | 0.074 |
| **Drama & Theater ( 20%)** | Other fields | 52.5 | 50 | 1.000 | 2.12 | 2.1 | 0.602 |
| **Economics ( 19%)** | Other fields | 746 | 597 | 0.001 | 3.2 | 3.13 | 0.033 |
| **Food Science ( 19%)** | Other fields | 1271 | 828 | 0.026 | 3.07 | 2.96 | 0.052 |
| **Artificial Intelligence & Image Processing ( 19%)** | Other fields | 1028 | 867 | 0.001 | 2.87 | 2.86 | 0.224 |
| **Distributed Computing ( 18%)** | Other fields | 665 | 450 | 0.010 | 2.52 | 2.39 | 0.332 |
| **Industrial Engineering & Automation ( 17%)** | Other fields | 860 | 714 | 0.017 | 2.96 | 2.86 | 0.099 |
| **Entomology ( 17%)** | Other fields | 918 | 503 | <0.001 | 3.04 | 2.8 | <0.001 |
| **Design Practice & Management ( 16%)** | Other fields | 577 | 515 | 0.839 | 2.83 | 2.77 | 0.973 |
| **Human Factors ( 16%)** | Highly related fields | 994 | 712 | 0.024 | 3.01 | 3.02 | 0.167 |
| **Information & Library Sciences ( 16%)** | Other fields | 257 | 173 | 0.117 | 2.73 | 2.54 | 0.098 |
| **Fluids & Plasmas ( 16%)** | Other fields | 943 | 802 | 0.198 | 3.11 | 3.08 | 0.434 |
| **General Psychology & Cognitive Sciences ( 16%)** | Highly related fields | 402 | 589 | 0.631 | 2.94 | 2.99 | 0.882 |
| **Urban & Regional Planning ( 15%)** | Other fields | 499 | 422 | 0.348 | 3.3 | 3.13 | 0.162 |
| **Education ( 15%)** | Other fields | 616 | 407 | <0.001 | 2.95 | 2.93 | 0.299 |
| **Psychoanalysis ( 15%)** | Highly related fields | 151 | 121 | 0.269 | 2.33 | 2.4 | 0.547 |
| **Development Studies ( 15%)** | Other fields | 720 | 605 | 0.633 | 3.13 | 3.14 | 0.891 |
| **Anatomy & Morphology ( 14%)** | Highly related fields | 260 | 180 | 0.374 | 2.44 | 2.4 | 0.656 |
| **Geography ( 14%)** | Other fields | 465 | 401 | 0.128 | 3.29 | 3.05 | 0.071 |
| **Ornithology ( 14%)** | Other fields | 287 | 361 | 0.569 | 2.75 | 2.68 | 0.752 |
| **History of Science, Technology & Medicine ( 13%)** | Other fields | 91.5 | 68 | 0.932 | 2.32 | 2.27 | 1.000 |
| **Gender Studies ( 13%)** | Highly related fields | 345 | 360 | 0.497 | 3.05 | 3.02 | 0.734 |
| **Dairy & Animal Science ( 13%)** | Other fields | 576 | 495 | 0.140 | 2.67 | 2.64 | 0.715 |
| **Logistics & Transportation ( 13%)** | Other fields | 481 | 604 | 0.265 | 2.89 | 2.96 | 0.130 |
| **Economic Theory ( 13%)** | Other fields | 308 | 358 | 0.427 | 3 | 2.86 | 0.525 |
| **Optoelectronics & Photonics ( 12%)** | Other fields | 469 | 368 | 0.040 | 2.44 | 2.41 | 0.524 |
| **Software Engineering ( 12%)** | Other fields | 485 | 630 | 0.183 | 2.7 | 2.8 | 0.209 |
| **Marine Biology & Hydrobiology ( 12%)** | Other fields | 920 | 943 | 0.963 | 3.01 | 3.08 | 0.258 |
| **Anthropology ( 12%)** | Other fields | 525 | 400 | 0.218 | 3.01 | 3 | 0.564 |
| **Numerical & Computational Mathematics ( 12%)** | Other fields | 995 | 515 | 0.061 | 2.93 | 3.01 | 0.950 |
| **Geological & Geomatics Engineering ( 11%)** | Other fields | 1035 | 977 | 0.332 | 3.16 | 3.07 | 0.255 |
| **Electrical & Electronic Engineering ( 11%)** | Other fields | 557 | 450 | 0.067 | 2.55 | 2.53 | 0.464 |
| **Physical Chemistry ( 10%)** | Other fields | 2102 | 1328 | 0.043 | 2.94 | 2.98 | 0.813 |
| **Econometrics ( 10%)** | Other fields | 898 | 1069 | 0.668 | 3.3 | 3.5 | 0.355 |
| **Computer Hardware & Architecture ( 10%)** | Other fields | 726 | 496 | 0.028 | 2.62 | 2.51 | 0.035 |
| **Communication & Media Studies ( 10%)** | Other fields | 520 | 446 | 0.388 | 3.13 | 3.07 | 0.452 |
| **Environmental Engineering ( 10%)** | Other fields | 1230 | 754 | <0.001 | 3.01 | 2.97 | 0.215 |
| **Networking & Telecommunications ( 10%)** | Other fields | 549 | 564 | 0.759 | 2.61 | 2.63 | 0.635 |
| **Archaeology ( 9.9%)** | Other fields | 388 | 350 | 0.645 | 2.75 | 2.79 | 0.351 |
| **Zoology ( 9.8%)** | Other fields | 284 | 345 | 0.703 | 2.61 | 2.57 | 0.901 |
| **Religions & Theology ( 9.8%)** | Other fields | 94.5 | 70 | 0.340 | 2.28 | 2.26 | 0.220 |
| **Chemical Engineering ( 9.6%)** | Other fields | 1385 | 854 | 0.079 | 3.08 | 2.96 | 0.278 |
| **Ecology ( 9.4%)** | Other fields | 1160 | 1222 | 0.989 | 3.24 | 3.15 | 0.131 |
| **Materials ( 9.3%)** | Other fields | 831 | 716 | 0.036 | 2.89 | 2.82 | 0.307 |
| **General Physics ( 9.1%)** | Other fields | 751 | 661 | 0.149 | 2.86 | 2.82 | 0.842 |
| **Oceanography ( 8.9%)** | Other fields | 796 | 641 | 0.104 | 3.15 | 2.92 | 0.313 |
| **Building & Construction ( 8.8%)** | Other fields | 1431 | 843 | 0.044 | 3.08 | 3.05 | 0.536 |
| **Sport, Leisure & Tourism ( 8.7%)** | Other fields | 550 | 675 | 0.275 | 3.1 | 3.09 | 0.959 |
| **Applied Mathematics ( 8.7%)** | Other fields | 1594 | 930 | 0.028 | 3.45 | 3.13 | 0.088 |
| **Mechanical Engineering & Transports ( 8.6%)** | Other fields | 562 | 556 | 0.776 | 2.81 | 2.86 | 0.224 |
| **Strategic, Defence & Security Studies ( 8.5%)** | Other fields | 602 | 373 | 0.157 | 2.84 | 2.86 | 0.661 |
| **Languages & Linguistics ( 8.1%)** | Other fields | 503 | 360 | 0.186 | 3.33 | 3.05 | 0.106 |
| **Computation Theory & Mathematics ( 8%)** | Other fields | 598 | 517 | 0.577 | 2.97 | 2.94 | 0.678 |
| **Marketing ( 7.9%)** | Other fields | 947 | 678 | 0.571 | 3.25 | 3.09 | 0.161 |
| **Law ( 7.6%)** | Other fields | 128 | 142 | 0.918 | 2.57 | 2.47 | 0.591 |
| **Meteorology & Atmospheric Sciences ( 7.4%)** | Other fields | 1419 | 1357 | 0.371 | 3.12 | 3.13 | 0.431 |
| **Applied Physics ( 7.3%)** | Other fields | 948 | 876 | 0.148 | 2.89 | 2.82 | 0.106 |
| **Operations Research ( 7.2%)** | Other fields | 530 | 707 | 0.492 | 3.12 | 3.15 | 0.780 |
| **Finance ( 7.2%)** | Other fields | 634 | 773 | 0.231 | 3.01 | 3.12 | 0.099 |
| **Political Science & Public Administration ( 7%)** | Other fields | 601 | 442 | 0.122 | 3.18 | 3.06 | 0.174 |
| **Forestry ( 6.9%)** | Other fields | 466 | 492 | 0.654 | 2.81 | 2.64 | 0.354 |
| **Business & Management ( 6.6%)** | Other fields | 965 | 776 | 0.162 | 3.19 | 3.17 | 0.607 |
| **Literary Studies ( 6.5%)** | Other fields | 45 | 44 | 0.836 | 2.12 | 2 | 0.921 |
| **General Mathematics ( 6.1%)** | Other fields | 229 | 302 | 0.110 | 2.84 | 2.85 | 0.511 |
| **Aerospace & Aeronautics ( 6.1%)** | Other fields | 345 | 289 | 0.533 | 2.49 | 2.5 | 0.320 |
| **Geology ( 6.1%)** | Other fields | 381 | 505 | 0.388 | 2.73 | 2.89 | 0.218 |
| **History ( 5.9%)** | Other fields | 57 | 47 | 0.278 | 2.17 | 2.07 | 0.239 |
| **Information Systems ( 5.9%)** | Other fields | 880 | 900 | 0.615 | 3.07 | 3.06 | 0.699 |
| **Energy ( 5.9%)** | Other fields | 953 | 780 | 0.032 | 2.91 | 2.87 | 0.655 |
| **Science Studies ( 5.7%)** | Other fields | 415 | 552 | 0.477 | 2.99 | 3.16 | 0.155 |
| **Legal & Forensic Medicine ( 5.7%)** | Highly related fields | 245 | 209 | 0.847 | 2.31 | 2.29 | 0.729 |
| **Cultural Studies ( 5.6%)** | Other fields | 84 | 128 | 0.189 | 2.46 | 2.54 | 0.407 |
| **Astronomy & Astrophysics ( 5.5%)** | Other fields | 1406 | 1769 | 0.427 | 3.11 | 3.14 | 0.995 |
| **Accounting ( 5.5%)** | Other fields | 809 | 687 | 0.396 | 3.04 | 3.1 | 0.827 |
| **Music ( 5.4%)** | Other fields | 49 | 52 | 0.788 | 2.15 | 2.06 | 0.893 |
| **Geochemistry & Geophysics ( 5.4%)** | Other fields | 729 | 874 | 0.329 | 3.16 | 3.15 | 0.608 |
| **Agronomy & Agriculture ( 5.3%)** | Other fields | 1018 | 765 | 0.154 | 3.01 | 2.89 | 0.110 |
| **Nuclear & Particle Physics ( 4.9%)** | Other fields | 830 | 757 | 0.865 | 2.93 | 2.9 | 0.391 |
| **Fisheries ( 4.8%)** | Other fields | 491 | 618 | 0.399 | 2.69 | 2.81 | 0.334 |
| **Classics ( 4.8%)** | Other fields | 67 | 40.5 | 0.186 | 2.32 | 2.03 | 0.186 |
| **Mining & Metallurgy ( 4.7%)** | Other fields | 241 | 252 | 0.908 | 2.54 | 2.36 | 0.686 |
| **Paleontology ( 4.4%)** | Other fields | 829 | 803 | 0.832 | 3.08 | 3.1 | 0.560 |
| **Agricultural Economics & Policy ( 4.3%)** | Other fields | 911 | 732 | 0.617 | 3.23 | 3.08 | 0.814 |
| **Philosophy ( 3.9%)** | Other fields | 415 | 201 | 0.246 | 3.16 | 2.85 | 0.118 |
| **International Relations ( 3.6%)** | Other fields | 389 | 343 | 0.791 | 2.92 | 3.01 | 0.402 |
| **Civil Engineering ( 2.9%)** | Other fields | 642 | 561 | 0.681 | 2.82 | 2.84 | 0.763 |
| **Mathematical Physics ( 0%)** | Other fields | . | 358 |  | . | 2.88 |  |
| **Industrial Relations ( 0%)** | Other fields | . | 218 |  | . | 2.87 |  |
| **Horticulture ( 0%)** | Other fields | . | 396 |  | . | 2.64 |  |
| **Folklore ( 0%)** | Other fields | . | 45 |  | . | 2.1 |  |
| **Automobile Design & Engineering ( 0%)** | Other fields | . | 122 |  | . | 2.18 |  |
| **Art Practice, History & Theory ( 0%)** | Other fields | . | 36 |  | . | 1.88 |  |
| **Architecture ( 0%)** | Other fields | . | 74 |  | . | 2.12 |  |

**Supplementary Table 2.5 : Recent year impact, Funding time recent funding citation counts and composite citation indices for each subfield (ordered by percentage funded)**

| **Top-cited US-based researchers: Subfield (perc. funded)** | **Classification** | **Citations for funded, median** | **Citations for non-funded, median** | **p-value** | **Composite index for funded, median** | **Composite index for non-funded, median** | **p-value** |
| --- | --- | --- | --- | --- | --- | --- | --- |
| **Geriatrics ( 55%)** | Highly related fields | 1681 | 1190 | 0.154 | 3.09 | 3.07 | 0.655 |
| **Substance Abuse ( 50%)** | Highly related fields | 1208 | 1014 | 0.353 | 3.06 | 3.12 | 0.113 |
| **Developmental Biology ( 49%)** | Highly related fields | 1905 | 1789 | 0.057 | 3.14 | 3.14 | 0.710 |
| **Medical Informatics ( 48%)** | Highly related fields | 712 | 624 | 0.171 | 2.73 | 2.7 | 0.690 |
| **Bioinformatics ( 46%)** | Highly related fields | 3039 | 1714 | <0.001 | 3.04 | 2.98 | 0.128 |
| **Virology ( 45%)** | Highly related fields | 1171 | 1229 | 0.577 | 2.86 | 2.84 | 0.695 |
| **Immunology ( 45%)** | Highly related fields | 1917 | 1827 | 0.358 | 3.11 | 3.09 | 0.272 |
| **Gerontology ( 44%)** | Highly related fields | 1240 | 1005 | 0.044 | 3.16 | 3.12 | 0.723 |
| **Biomedical Engineering ( 44%)** | Highly related fields | 990 | 806 | 0.001 | 2.84 | 2.83 | 0.194 |
| **Neurology & Neurosurgery ( 42%)** | Highly related fields | 1581 | 1462 | 0.004 | 3.13 | 3.12 | 0.879 |
| **Oncology & Carcinogenesis ( 40%)** | Highly related fields | 2297 | 1905 | <0.001 | 3.02 | 2.98 | 0.057 |
| **Public Health ( 38%)** | Highly related fields | 1260 | 1095 | 0.100 | 3.13 | 3.1 | 0.724 |
| **Genetics & Heredity ( 38%)** | Highly related fields | 1953 | 1279 | <0.001 | 2.86 | 2.84 | 0.137 |
| **Emergency & Critical Care Medicine ( 38%)** | Highly related fields | 1438 | 975 | <0.001 | 2.98 | 2.82 | 0.001 |
| **Epidemiology ( 36%)** | Highly related fields | 2172 | 1906 | 0.198 | 3.14 | 3.15 | 0.938 |
| **Psychiatry ( 36%)** | Highly related fields | 1746 | 1523 | 0.002 | 3.18 | 3.18 | 0.647 |
| **Gastroenterology & Hepatology ( 35%)** | Highly related fields | 1440 | 1259 | 0.019 | 3.01 | 2.95 | 0.080 |
| **Analytical Chemistry ( 35%)** | Other fields | 867 | 707 | <0.001 | 2.8 | 2.79 | 0.404 |
| **Arthritis & Rheumatology ( 34%)** | Highly related fields | 1818 | 2052 | 0.637 | 3.11 | 3.13 | 0.754 |
| **Allergy ( 34%)** | Highly related fields | 1704 | 1344 | 0.114 | 3.15 | 3.17 | 0.822 |
| **Developmental & Child Psychology ( 33%)** | Highly related fields | 1244 | 892 | <0.001 | 3.18 | 3.16 | 0.213 |
| **Endocrinology & Metabolism ( 33%)** | Highly related fields | 1458 | 1542 | 0.667 | 3.13 | 3.15 | 0.394 |
| **Respiratory System ( 33%)** | Highly related fields | 1531 | 1438 | 0.190 | 2.95 | 2.95 | 0.535 |
| **Nuclear Medicine & Medical Imaging ( 32%)** | Highly related fields | 1057 | 786 | <0.001 | 2.78 | 2.68 | <0.001 |
| **Cardiovascular System & Hematology ( 32%)** | Highly related fields | 1990 | 1723 | <0.001 | 3.07 | 3.01 | 0.003 |
| **Health Policy & Services ( 31%)** | Highly related fields | 1183 | 1013 | 0.041 | 3.03 | 2.95 | 0.051 |
| **Microbiology ( 31%)** | Highly related fields | 1377 | 1280 | 0.151 | 2.99 | 2.98 | 0.211 |
| **Toxicology ( 31%)** | Highly related fields | 1129 | 777 | <0.001 | 2.85 | 2.79 | 0.021 |
| **Pediatrics ( 31%)** | Highly related fields | 873 | 698 | <0.001 | 2.73 | 2.67 | 0.007 |
| **Biophysics ( 31%)** | Highly related fields | 707 | 587 | 0.352 | 2.81 | 2.79 | 0.553 |
| **Rehabilitation ( 30%)** | Highly related fields | 786 | 700 | 0.277 | 2.82 | 2.86 | 0.360 |
| **Urology & Nephrology ( 30%)** | Highly related fields | 1279 | 1108 | 0.016 | 2.97 | 2.85 | <0.001 |
| **Ophthalmology & Optometry ( 30%)** | Highly related fields | 1062 | 754 | <0.001 | 2.91 | 2.77 | <0.001 |
| **Physiology ( 29%)** | Highly related fields | 765 | 628 | 0.036 | 2.91 | 2.96 | 0.765 |
| **Biochemistry & Molecular Biology ( 29%)** | Highly related fields | 1064 | 888 | <0.001 | 2.97 | 2.93 | 0.504 |
| **Clinical Psychology ( 28%)** | Highly related fields | 1307 | 1078 | 0.007 | 3.17 | 3.15 | 0.392 |
| **Medicinal & Biomolecular Chemistry ( 28%)** | Highly related fields | 780 | 672 | 0.010 | 2.64 | 2.62 | 0.353 |
| **Obstetrics & Reproductive Medicine ( 28%)** | Highly related fields | 878 | 747 | 0.031 | 2.81 | 2.79 | 0.338 |
| **Anesthesiology ( 27%)** | Highly related fields | 798 | 627 | <0.001 | 2.76 | 2.68 | 0.059 |
| **Tropical Medicine ( 27%)** | Highly related fields | 897 | 697 | 0.022 | 2.75 | 2.69 | 0.292 |
| **Pharmacology & Pharmacy ( 25%)** | Highly related fields | 772 | 588 | <0.001 | 2.75 | 2.66 | 0.002 |
| **Nutrition & Dietetics ( 25%)** | Highly related fields | 1386 | 995 | <0.001 | 3.16 | 3.04 | 0.179 |
| **Nursing ( 25%)** | Highly related fields | 376 | 250 | <0.001 | 2.52 | 2.46 | 0.007 |
| **Demography ( 25%)** | Highly related fields | 533 | 446 | 0.149 | 3.07 | 2.97 | 0.454 |
| **Speech-Language Pathology & Audiology ( 25%)** | Highly related fields | 485 | 490 | 0.707 | 2.86 | 2.88 | 0.449 |
| **Statistics & Probability ( 24%)** | Other fields | 1532 | 835 | <0.001 | 3.19 | 3.12 | 0.226 |
| **Family Studies ( 24%)** | Other fields | 609 | 434 | 0.005 | 3.14 | 2.94 | 0.003 |
| **Environmental & Occupational Health ( 23%)** | Highly related fields | 626 | 508 | 0.335 | 2.65 | 2.62 | 0.753 |
| **Experimental Psychology ( 22%)** | Highly related fields | 1078 | 825 | <0.001 | 3.17 | 3.2 | 0.425 |
| **Applied Ethics ( 22%)** | Highly related fields | 847 | 493 | 0.010 | 3.06 | 2.94 | 0.204 |
| **Organic Chemistry ( 22%)** | Other fields | 1158 | 856 | <0.001 | 2.97 | 2.9 | 0.038 |
| **Optics ( 22%)** | Other fields | 985 | 918 | 0.146 | 2.76 | 2.77 | 0.756 |
| **Microscopy ( 21%)** | Highly related fields | 1129 | 608 | 0.045 | 2.8 | 2.74 | 0.291 |
| **General & Internal Medicine ( 21%)** | Highly related fields | 777 | 613 | <0.001 | 2.71 | 2.55 | <0.001 |
| **Biotechnology ( 21%)** | Highly related fields | 1022 | 959 | 0.997 | 2.91 | 2.85 | 0.742 |
| **Otorhinolaryngology ( 20%)** | Highly related fields | 605 | 497 | 0.042 | 2.67 | 2.6 | 0.156 |
| **Surgery ( 20%)** | Highly related fields | 1074 | 752 | <0.001 | 2.75 | 2.65 | 0.001 |
| **General Chemistry ( 19%)** | Other fields | 903 | 728 | 0.046 | 2.82 | 2.68 | 0.122 |
| **Mycology & Parasitology ( 18%)** | Highly related fields | 734 | 755 | 0.825 | 2.88 | 2.79 | 0.641 |
| **Nanoscience & Nanotechnology ( 18%)** | Other fields | 2394 | 2441 | 0.351 | 3.2 | 3.14 | 0.052 |
| **Behavioral Science & Comparative Psychology ( 17%)** | Highly related fields | 796 | 668 | 0.240 | 3.04 | 3.1 | 0.863 |
| **Dermatology & Venereal Diseases ( 16%)** | Highly related fields | 976 | 751 | 0.001 | 2.81 | 2.78 | 0.181 |
| **Acoustics ( 16%)** | Other fields | 566 | 436 | 0.014 | 2.74 | 2.78 | 0.460 |
| **Orthopedics ( 15%)** | Highly related fields | 1083 | 871 | 0.001 | 2.95 | 2.85 | 0.031 |
| **Environmental Sciences ( 15%)** | Other fields | 1987 | 1277 | <0.001 | 3.21 | 3.08 | 0.082 |
| **Sociology ( 13%)** | Other fields | 489 | 438 | 0.499 | 3.14 | 3.09 | 0.833 |
| **Complementary & Alternative Medicine ( 12%)** | Highly related fields | 653 | 290 | 0.008 | 2.79 | 2.55 | 0.073 |
| **Dentistry ( 12%)** | Highly related fields | 793 | 490 | <0.001 | 2.77 | 2.7 | 0.014 |
| **Distributed Computing ( 12%)** | Other fields | 605 | 454 | 0.080 | 2.37 | 2.41 | 0.773 |
| **Pathology ( 11%)** | Highly related fields | 1579 | 1032 | 0.020 | 2.89 | 2.83 | 0.208 |
| **Evolutionary Biology ( 11%)** | Other fields | 1194 | 931 | 0.014 | 3.26 | 3.15 | 0.109 |
| **Artificial Intelligence & Image Processing ( 11%)** | Other fields | 987 | 895 | 0.134 | 2.85 | 2.86 | 0.677 |
| **Veterinary Sciences ( 11%)** | Highly related fields | 534 | 398 | <0.001 | 2.63 | 2.55 | 0.012 |
| **Plant Biology & Botany ( 10%)** | Other fields | 1190 | 895 | <0.001 | 3.01 | 2.96 | 0.085 |
| **Chemical Physics ( 10%)** | Other fields | 1648 | 1200 | 0.003 | 3.16 | 3.1 | 0.068 |
| **Industrial Engineering & Automation ( 10%)** | Other fields | 810 | 721 | 0.344 | 2.93 | 2.87 | 0.993 |
| **Sport Sciences ( 10%)** | Highly related fields | 1264 | 941 | 0.109 | 3.02 | 3.01 | 0.663 |
| **Drama & Theater ( 10%)** | Other fields | 73 | 48 | 0.384 | 2.2 | 2.08 | 0.223 |
| **Social Work ( 9.6%)** | Other fields | 527 | 275 | 0.004 | 2.97 | 2.72 | 0.014 |
| **Criminology ( 9.6%)** | Other fields | 910 | 514 | 0.001 | 3.14 | 3.07 | 0.107 |
| **Design Practice & Management ( 9%)** | Other fields | 622 | 508 | 0.442 | 2.81 | 2.78 | 0.878 |
| **Polymers ( 8.5%)** | Other fields | 1668 | 852 | <0.001 | 3.03 | 2.89 | 0.211 |
| **General Clinical Medicine ( 7.8%)** | Highly related fields | 577 | 295 | 0.059 | 2.57 | 2.36 | 0.423 |
| **Geography ( 7.5%)** | Other fields | 465 | 401 | 0.206 | 3.29 | 3.05 | 0.405 |
| **Social Psychology ( 7.5%)** | Highly related fields | 1276 | 936 | 0.090 | 3.48 | 3.26 | 0.061 |
| **Development Studies ( 7.4%)** | Other fields | 553 | 605 | 0.711 | 3.02 | 3.18 | 0.267 |
| **Economics ( 7.2%)** | Other fields | 680 | 610 | 0.008 | 3.26 | 3.14 | 0.059 |
| **Fluids & Plasmas ( 7.1%)** | Other fields | 1286 | 796 | 0.003 | 3.3 | 3.07 | 0.040 |
| **Social Sciences Methods ( 7%)** | Other fields | 2253 | 740 | 0.013 | 3.63 | 3.16 | 0.006 |
| **Entomology ( 6.9%)** | Other fields | 1083 | 524 | <0.001 | 3.22 | 2.82 | <0.001 |
| **Numerical & Computational Mathematics ( 6.9%)** | Other fields | 1112 | 512 | 0.033 | 2.92 | 3 | 0.957 |
| **Electrical & Electronic Engineering ( 6.9%)** | Other fields | 489 | 452 | 0.277 | 2.5 | 2.54 | 0.706 |
| **Gender Studies ( 6.7%)** | Highly related fields | 326 | 362 | 0.247 | 3.14 | 2.99 | 0.355 |
| **Computer Hardware & Architecture ( 6.6%)** | Other fields | 772 | 496 | 0.040 | 2.62 | 2.52 | 0.218 |
| **Logistics & Transportation ( 6.6%)** | Other fields | 635 | 596 | 0.994 | 2.93 | 2.96 | 0.334 |
| **Food Science ( 6.5%)** | Other fields | 1381 | 845 | 0.066 | 3.05 | 2.97 | 0.422 |
| **Information & Library Sciences ( 6.3%)** | Other fields | 188 | 181 | 0.467 | 2.79 | 2.55 | 0.058 |
| **Psychoanalysis ( 6.1%)** | Highly related fields | 334 | 121 | 0.070 | 2.77 | 2.38 | 0.258 |
| **Education ( 6%)** | Other fields | 724 | 413 | <0.001 | 2.96 | 2.94 | 0.294 |
| **Archaeology ( 5.9%)** | Other fields | 382 | 354 | 0.513 | 2.76 | 2.79 | 0.698 |
| **Optoelectronics & Photonics ( 5.9%)** | Other fields | 501 | 369 | 0.020 | 2.48 | 2.41 | 0.286 |
| **History of Social Sciences ( 5.9%)** | Other fields | 184 | 215 | 0.838 | 2.58 | 2.77 | 0.414 |
| **Science Studies ( 5.7%)** | Other fields | 415 | 552 | 0.477 | 2.99 | 3.16 | 0.155 |
| **Human Factors ( 5.6%)** | Highly related fields | 991 | 748 | 0.284 | 3.06 | 3.01 | 0.457 |
| **Urban & Regional Planning ( 5.6%)** | Other fields | 799 | 426 | 0.307 | 3.43 | 3.16 | 0.178 |
| **Networking & Telecommunications ( 5.2%)** | Other fields | 537 | 564 | 0.840 | 2.6 | 2.63 | 0.237 |
| **Dairy & Animal Science ( 5.1%)** | Other fields | 701 | 492 | 0.026 | 2.57 | 2.64 | 0.938 |
| **Languages & Linguistics ( 5.1%)** | Other fields | 365 | 362 | 0.936 | 3.2 | 3.06 | 0.987 |
| **Building & Construction ( 4.9%)** | Other fields | 1504 | 843 | 0.036 | 3.08 | 3.05 | 0.846 |
| **Religions & Theology ( 4.9%)** | Other fields | 98.5 | 70 | 0.185 | 2.24 | 2.28 | 0.532 |
| **Inorganic & Nuclear Chemistry ( 4.9%)** | Other fields | 922 | 739 | 0.550 | 2.8 | 2.78 | 0.904 |
| **Applied Mathematics ( 4.8%)** | Other fields | 1594 | 930 | 0.027 | 3.55 | 3.13 | 0.164 |
| **Mechanical Engineering & Transports ( 4.7%)** | Other fields | 538 | 559 | 0.812 | 2.81 | 2.86 | 0.246 |
| **General Psychology & Cognitive Sciences ( 4.7%)** | Highly related fields | 501 | 578 | 0.886 | 2.79 | 2.98 | 0.260 |
| **Mining & Metallurgy ( 4.7%)** | Other fields | 836 | 252 | 0.453 | 2.88 | 2.36 | 0.299 |
| **Software Engineering ( 4.5%)** | Other fields | 719 | 622 | 0.744 | 2.76 | 2.79 | 0.589 |
| **Anthropology ( 4.3%)** | Other fields | 592 | 400 | 0.146 | 2.88 | 3.01 | 0.426 |
| **Materials ( 4.3%)** | Other fields | 1249 | 716 | 0.003 | 2.95 | 2.82 | 0.251 |
| **Chemical Engineering ( 4.3%)** | Other fields | 1394 | 870 | 0.403 | 3.02 | 2.96 | 0.652 |
| **Accounting ( 4.1%)** | Other fields | 805 | 700 | 0.911 | 2.97 | 3.11 | 0.266 |
| **Econometrics ( 4.1%)** | Other fields | 1326 | 1065 | 0.649 | 3.1 | 3.48 | 0.157 |
| **Ecology ( 3.9%)** | Other fields | 1006 | 1226 | 0.459 | 3.16 | 3.16 | 0.514 |
| **Sport, Leisure & Tourism ( 3.8%)** | Other fields | 584 | 658 | 0.761 | 3.09 | 3.1 | 0.866 |
| **Legal & Forensic Medicine ( 3.8%)** | Highly related fields | 172 | 211 | 0.608 | 2.3 | 2.3 | 0.963 |
| **Communication & Media Studies ( 3.7%)** | Other fields | 718 | 446 | 0.126 | 3.12 | 3.08 | 0.639 |
| **Physical Chemistry ( 3.7%)** | Other fields | 1724 | 1358 | 0.490 | 2.96 | 2.98 | 0.743 |
| **Operations Research ( 3.6%)** | Other fields | 680 | 693 | 0.686 | 3.16 | 3.14 | 0.618 |
| **Anatomy & Morphology ( 3.6%)** | Highly related fields | 373 | 184 | 1.000 | 2.39 | 2.4 | 0.895 |
| **Information Systems ( 3.6%)** | Other fields | 837 | 900 | 0.822 | 3.17 | 3.06 | 0.865 |
| **Marketing ( 3.5%)** | Other fields | 724 | 686 | 0.859 | 3.23 | 3.09 | 0.683 |
| **Ornithology ( 3.4%)** | Other fields | 401 | 353 | 0.811 | 2.95 | 2.68 | 0.189 |
| **General Physics ( 3.3%)** | Other fields | 760 | 679 | 0.288 | 2.86 | 2.82 | 0.471 |
| **Finance ( 3.3%)** | Other fields | 698 | 738 | 0.449 | 3.1 | 3.11 | 0.623 |
| **Zoology ( 3.3%)** | Other fields | 590 | 327 | 0.156 | 2.67 | 2.57 | 0.517 |
| **Oceanography ( 3%)** | Other fields | 1059 | 653 | 0.100 | 3.16 | 2.92 | 0.127 |
| **Meteorology & Atmospheric Sciences ( 3%)** | Other fields | 1305 | 1360 | 0.448 | 3.27 | 3.12 | 0.080 |
| **Applied Physics ( 2.8%)** | Other fields | 1189 | 879 | 0.019 | 2.81 | 2.82 | 0.569 |
| **Geological & Geomatics Engineering ( 2.8%)** | Other fields | 1880 | 935 | 0.064 | 3.46 | 3.07 | 0.037 |
| **Aerospace & Aeronautics ( 2.8%)** | Other fields | 306 | 291 | 0.840 | 2.48 | 2.5 | 0.533 |
| **Marine Biology & Hydrobiology ( 2.8%)** | Other fields | 919 | 940 | 0.576 | 2.97 | 3.07 | 0.296 |
| **Cultural Studies ( 2.8%)** | Other fields | 100 | 124 | 0.810 | 2.52 | 2.54 | 0.962 |
| **Energy ( 2.6%)** | Other fields | 1018 | 788 | 0.053 | 2.94 | 2.87 | 0.548 |
| **Environmental Engineering ( 2.5%)** | Other fields | 971 | 787 | 0.178 | 2.9 | 2.97 | 0.715 |
| **Geology ( 2.4%)** | Other fields | 378 | 490 | 0.321 | 2.83 | 2.89 | 0.400 |
| **Business & Management ( 2.3%)** | Other fields | 932 | 790 | 0.260 | 3.31 | 3.17 | 0.288 |
| **Strategic, Defence & Security Studies ( 2.3%)** | Other fields | 225 | 408 | 0.587 | 2.84 | 2.86 | 0.480 |
| **Computation Theory & Mathematics ( 2.3%)** | Other fields | 862 | 507 | 0.082 | 3.25 | 2.93 | 0.104 |
| **Law ( 2.2%)** | Other fields | 219 | 138 | 0.470 | 2.55 | 2.48 | 0.851 |
| **Political Science & Public Administration ( 2%)** | Other fields | 1694 | 446 | 0.044 | 3.43 | 3.07 | 0.059 |
| **History ( 2%)** | Other fields | 48.5 | 50 | 0.678 | 2.08 | 2.08 | 0.715 |
| **International Relations ( 1.8%)** | Other fields | 519 | 333 | 0.439 | 2.98 | 3.01 | 0.734 |
| **Astronomy & Astrophysics ( 1.7%)** | Other fields | 1280 | 1769 | 0.297 | 3.11 | 3.14 | 0.906 |
| **General Mathematics ( 1.7%)** | Other fields | 251 | 299 | 0.648 | 2.95 | 2.85 | 0.847 |
| **Fisheries ( 1.6%)** | Other fields | 1170 | 593 | 0.169 | 2.99 | 2.8 | 0.263 |
| **Philosophy ( 1.6%)** | Other fields | 736 | 200 | 0.023 | 3.65 | 2.86 | 0.020 |
| **Literary Studies ( 1.4%)** | Other fields | 95.5 | 44 | 0.285 | 2.44 | 1.99 | 0.165 |
| **Agricultural Economics & Policy ( 1.4%)** | Other fields | 911 | 732 | 0.651 | 3.23 | 3.08 | 0.547 |
| **Civil Engineering ( 1.4%)** | Other fields | 1074 | 563 | 0.215 | 2.83 | 2.84 | 0.855 |
| **Geochemistry & Geophysics ( 1.3%)** | Other fields | 733 | 869 | 0.420 | 3.14 | 3.15 | 0.499 |
| **Forestry ( 1.1%)** | Other fields | 489 | 492 | 0.720 | 2.73 | 2.65 | 0.888 |
| **Nuclear & Particle Physics ( 1.1%)** | Other fields | 890 | 759 | 0.708 | 2.9 | 2.9 | 0.969 |
| **Agronomy & Agriculture ( .83%)** | Other fields | 761 | 772 | 0.383 | 3.23 | 2.89 | 0.129 |
| **Paleontology ( .74%)** | Other fields | 496 | 803 | 0.279 | 2.88 | 3.1 | 0.143 |
| **Music ( 0%)** | Other fields | . | 52 |  | . | 2.06 |  |
| **Mathematical Physics ( 0%)** | Other fields | . | 358 |  | . | 2.88 |  |
| **Industrial Relations ( 0%)** | Other fields | . | 218 |  | . | 2.87 |  |
| **Horticulture ( 0%)** | Other fields | . | 396 |  | . | 2.64 |  |
| **History of Science, Technology & Medicine ( 0%)** | Other fields | . | 68 |  | . | 2.27 |  |
| **Folklore ( 0%)** | Other fields | . | 45 |  | . | 2.1 |  |
| **Economic Theory ( 0%)** | Other fields | . | 323 |  | . | 2.93 |  |
| **Classics ( 0%)** | Other fields | . | 41 |  | . | 2.04 |  |
| **Automobile Design & Engineering ( 0%)** | Other fields | . | 122 |  | . | 2.18 |  |
| **Art Practice, History & Theory ( 0%)** | Other fields | . | 36 |  | . | 1.88 |  |
| **Architecture ( 0%)** | Other fields | . | 74 |  | . | 2.12 |  |

**Supplementary Table 2.6 : Recent year impact, Funding time current funding citation counts and composite citation indices for each subfield (ordered by percentage funded)**

| **Top-cited US-based researchers: Subfield (perc. funded)** | **Classification** | **Citations for funded, median** | **Citations for non-funded, median** | **p-value** | **Composite index for funded, median** | **Composite index for non-funded, median** | **p-value** |
| --- | --- | --- | --- | --- | --- | --- | --- |
| **Geriatrics ( 43%)** | Highly related fields | 1755 | 1287 | 0.380 | 3.07 | 3.08 | 0.881 |
| **Developmental Biology ( 36%)** | Highly related fields | 1956 | 1802 | 0.054 | 3.13 | 3.15 | 0.390 |
| **Medical Informatics ( 34%)** | Highly related fields | 708 | 637 | 0.238 | 2.74 | 2.69 | 0.195 |
| **Bioinformatics ( 34%)** | Highly related fields | 2949 | 1739 | 0.004 | 3.06 | 2.98 | 0.079 |
| **Substance Abuse ( 31%)** | Highly related fields | 1356 | 1028 | 0.093 | 3.06 | 3.1 | 0.280 |
| **Gerontology ( 31%)** | Highly related fields | 1240 | 1066 | 0.442 | 3.07 | 3.14 | 0.600 |
| **Biomedical Engineering ( 30%)** | Highly related fields | 988 | 831 | 0.016 | 2.83 | 2.83 | 0.682 |
| **Virology ( 30%)** | Highly related fields | 1198 | 1202 | 0.335 | 2.88 | 2.84 | 0.224 |
| **Immunology ( 30%)** | Highly related fields | 1925 | 1841 | 0.561 | 3.1 | 3.09 | 0.706 |
| **Neurology & Neurosurgery ( 29%)** | Highly related fields | 1577 | 1479 | 0.005 | 3.13 | 3.11 | 0.423 |
| **Allergy ( 27%)** | Highly related fields | 1600 | 1362 | 0.851 | 3.15 | 3.17 | 0.753 |
| **Emergency & Critical Care Medicine ( 27%)** | Highly related fields | 1381 | 1013 | 0.001 | 2.97 | 2.84 | 0.023 |
| **Oncology & Carcinogenesis ( 26%)** | Highly related fields | 2330 | 1978 | <0.001 | 3.02 | 2.99 | 0.625 |
| **Public Health ( 26%)** | Highly related fields | 1298 | 1132 | 0.051 | 3.12 | 3.11 | 0.759 |
| **Gastroenterology & Hepatology ( 24%)** | Highly related fields | 1433 | 1281 | 0.092 | 2.99 | 2.97 | 0.319 |
| **Psychiatry ( 23%)** | Highly related fields | 1729 | 1573 | 0.005 | 3.16 | 3.19 | 0.631 |
| **Arthritis & Rheumatology ( 23%)** | Highly related fields | 2088 | 1964 | 0.951 | 3.07 | 3.14 | 0.447 |
| **Epidemiology ( 22%)** | Highly related fields | 1990 | 2028 | 0.977 | 3.06 | 3.17 | 0.157 |
| **Genetics & Heredity ( 22%)** | Highly related fields | 1770 | 1357 | 0.006 | 2.84 | 2.84 | 0.870 |
| **Analytical Chemistry ( 22%)** | Other fields | 973 | 729 | <0.001 | 2.8 | 2.79 | 0.751 |
| **Ophthalmology & Optometry ( 22%)** | Highly related fields | 1123 | 783 | <0.001 | 2.88 | 2.79 | 0.002 |
| **Developmental & Child Psychology ( 22%)** | Highly related fields | 1221 | 950 | 0.005 | 3.18 | 3.16 | 0.953 |
| **Nuclear Medicine & Medical Imaging ( 22%)** | Highly related fields | 1108 | 810 | <0.001 | 2.76 | 2.7 | <0.001 |
| **Endocrinology & Metabolism ( 21%)** | Highly related fields | 1444 | 1541 | 0.529 | 3.13 | 3.15 | 0.985 |
| **Biophysics ( 21%)** | Highly related fields | 707 | 601 | 0.608 | 2.78 | 2.8 | 0.839 |
| **Toxicology ( 21%)** | Highly related fields | 1129 | 818 | <0.001 | 2.83 | 2.8 | 0.246 |
| **Cardiovascular System & Hematology ( 21%)** | Highly related fields | 1984 | 1754 | 0.001 | 3.06 | 3.02 | 0.043 |
| **Respiratory System ( 21%)** | Highly related fields | 1446 | 1449 | 0.427 | 2.92 | 2.96 | 0.184 |
| **Physiology ( 21%)** | Highly related fields | 705 | 663 | 0.590 | 2.91 | 2.96 | 0.784 |
| **Rehabilitation ( 21%)** | Highly related fields | 815 | 715 | 0.137 | 2.8 | 2.85 | 0.209 |
| **Microbiology ( 20%)** | Highly related fields | 1396 | 1277 | 0.021 | 3 | 2.98 | 0.066 |
| **Urology & Nephrology ( 20%)** | Highly related fields | 1324 | 1109 | 0.070 | 2.97 | 2.87 | 0.003 |
| **Medicinal & Biomolecular Chemistry ( 20%)** | Highly related fields | 806 | 679 | 0.023 | 2.63 | 2.62 | 0.686 |
| **Health Policy & Services ( 20%)** | Highly related fields | 1144 | 1020 | 0.192 | 3.01 | 2.98 | 0.553 |
| **Pediatrics ( 19%)** | Highly related fields | 845 | 714 | 0.004 | 2.73 | 2.68 | 0.078 |
| **Obstetrics & Reproductive Medicine ( 19%)** | Highly related fields | 791 | 766 | 0.634 | 2.8 | 2.79 | 0.776 |
| **Anesthesiology ( 18%)** | Highly related fields | 781 | 644 | 0.004 | 2.73 | 2.7 | 0.298 |
| **Statistics & Probability ( 17%)** | Other fields | 1491 | 873 | 0.001 | 3.19 | 3.12 | 0.360 |
| **Biochemistry & Molecular Biology ( 17%)** | Highly related fields | 1141 | 901 | <0.001 | 2.99 | 2.93 | 0.085 |
| **Speech-Language Pathology & Audiology ( 17%)** | Highly related fields | 607 | 469 | 0.574 | 2.89 | 2.87 | 0.799 |
| **Demography ( 17%)** | Highly related fields | 625 | 445 | 0.149 | 3.09 | 2.96 | 0.373 |
| **Experimental Psychology ( 16%)** | Highly related fields | 1154 | 818 | <0.001 | 3.23 | 3.19 | 0.060 |
| **Pharmacology & Pharmacy ( 15%)** | Highly related fields | 775 | 608 | <0.001 | 2.74 | 2.67 | 0.213 |
| **Clinical Psychology ( 15%)** | Highly related fields | 1232 | 1126 | 0.207 | 3.16 | 3.15 | 0.919 |
| **Applied Ethics ( 15%)** | Highly related fields | 988 | 549 | 0.021 | 3.07 | 2.96 | 0.252 |
| **Organic Chemistry ( 15%)** | Other fields | 1175 | 873 | 0.005 | 2.92 | 2.9 | 0.262 |
| **Nutrition & Dietetics ( 14%)** | Highly related fields | 1378 | 1018 | 0.019 | 3.14 | 3.06 | 0.397 |
| **Mycology & Parasitology ( 14%)** | Highly related fields | 734 | 755 | 0.996 | 2.81 | 2.8 | 0.660 |
| **Nursing ( 14%)** | Highly related fields | 409 | 266 | <0.001 | 2.5 | 2.47 | 0.119 |
| **Optics ( 14%)** | Other fields | 931 | 927 | 0.322 | 2.73 | 2.77 | 0.760 |
| **General Chemistry ( 13%)** | Other fields | 911 | 730 | 0.064 | 2.84 | 2.69 | 0.064 |
| **General & Internal Medicine ( 13%)** | Highly related fields | 804 | 632 | <0.001 | 2.71 | 2.56 | <0.001 |
| **Surgery ( 13%)** | Highly related fields | 1067 | 770 | <0.001 | 2.75 | 2.66 | 0.011 |
| **Nanoscience & Nanotechnology ( 13%)** | Other fields | 2256 | 2447 | 0.903 | 3.15 | 3.15 | 0.530 |
| **Otorhinolaryngology ( 12%)** | Highly related fields | 708 | 499 | 0.007 | 2.63 | 2.61 | 0.490 |
| **Environmental & Occupational Health ( 12%)** | Highly related fields | 594 | 538 | 0.749 | 2.66 | 2.62 | 0.991 |
| **Tropical Medicine ( 12%)** | Highly related fields | 873 | 736 | 0.190 | 2.76 | 2.69 | 0.354 |
| **Microscopy ( 12%)** | Highly related fields | 1046 | 664 | 0.408 | 2.9 | 2.75 | 0.544 |
| **Biotechnology ( 12%)** | Highly related fields | 966 | 971 | 0.885 | 2.87 | 2.86 | 0.932 |
| **Drama & Theater ( 10%)** | Other fields | 73 | 48 | 0.384 | 2.2 | 2.08 | 0.223 |
| **Complementary & Alternative Medicine ( 10%)** | Highly related fields | 669 | 294 | 0.017 | 2.8 | 2.53 | 0.025 |
| **Acoustics ( 9.9%)** | Other fields | 558 | 448 | 0.161 | 2.74 | 2.77 | 0.586 |
| **Dermatology & Venereal Diseases ( 9.7%)** | Highly related fields | 976 | 768 | 0.034 | 2.81 | 2.79 | 0.646 |
| **Orthopedics ( 9.5%)** | Highly related fields | 1121 | 880 | 0.004 | 2.97 | 2.85 | 0.025 |
| **Family Studies ( 9.1%)** | Other fields | 643 | 442 | 0.114 | 3.05 | 2.97 | 0.482 |
| **Environmental Sciences ( 9%)** | Other fields | 2081 | 1297 | 0.001 | 3.25 | 3.08 | 0.083 |
| **Behavioral Science & Comparative Psychology ( 8.7%)** | Highly related fields | 1129 | 671 | 0.278 | 3.06 | 3.08 | 0.751 |
| **Sociology ( 7.8%)** | Other fields | 418 | 445 | 0.472 | 3.03 | 3.1 | 0.219 |
| **Pathology ( 7.7%)** | Highly related fields | 1656 | 1079 | 0.182 | 2.89 | 2.83 | 0.479 |
| **Design Practice & Management ( 7.5%)** | Other fields | 577 | 515 | 0.519 | 2.94 | 2.77 | 0.633 |
| **Dentistry ( 7.3%)** | Highly related fields | 713 | 499 | 0.002 | 2.71 | 2.7 | 0.220 |
| **Plant Biology & Botany ( 7.2%)** | Other fields | 1262 | 909 | <0.001 | 3 | 2.96 | 0.251 |
| **Evolutionary Biology ( 7.1%)** | Other fields | 1247 | 934 | 0.025 | 3.3 | 3.15 | 0.039 |
| **Artificial Intelligence & Image Processing ( 6.7%)** | Other fields | 908 | 904 | 0.630 | 2.8 | 2.86 | 0.410 |
| **Chemical Physics ( 6.7%)** | Other fields | 1648 | 1222 | 0.059 | 3.2 | 3.1 | 0.124 |
| **Industrial Engineering & Automation ( 6.7%)** | Other fields | 794 | 728 | 0.537 | 2.89 | 2.88 | 0.828 |
| **Criminology ( 6.4%)** | Other fields | 733 | 523 | 0.031 | 3.13 | 3.07 | 0.456 |
| **Sport Sciences ( 6.3%)** | Highly related fields | 1591 | 941 | 0.018 | 3.08 | 3.01 | 0.395 |
| **Veterinary Sciences ( 6.2%)** | Highly related fields | 615 | 411 | 0.011 | 2.67 | 2.55 | 0.065 |
| **Psychoanalysis ( 6.1%)** | Highly related fields | 334 | 121 | 0.070 | 2.77 | 2.38 | 0.258 |
| **Social Work ( 6%)** | Other fields | 697 | 295 | 0.009 | 2.98 | 2.72 | 0.034 |
| **Polymers ( 5.9%)** | Other fields | 1743 | 861 | 0.004 | 3 | 2.9 | 0.902 |
| **General Clinical Medicine ( 5.8%)** | Highly related fields | 697 | 295 | 0.017 | 2.57 | 2.36 | 0.499 |
| **Entomology ( 5.8%)** | Other fields | 1093 | 526 | <0.001 | 3.24 | 2.82 | <0.001 |
| **Religions & Theology ( 4.9%)** | Other fields | 98.5 | 70 | 0.185 | 2.24 | 2.28 | 0.532 |
| **Distributed Computing ( 4.9%)** | Other fields | 602 | 461 | 0.565 | 2.43 | 2.4 | 0.365 |
| **Computer Hardware & Architecture ( 4.4%)** | Other fields | 707 | 497 | 0.197 | 2.69 | 2.52 | 0.213 |
| **Information & Library Sciences ( 4.2%)** | Other fields | 147 | 181 | 0.875 | 2.71 | 2.57 | 0.266 |
| **Education ( 4%)** | Other fields | 704 | 417 | 0.001 | 2.99 | 2.93 | 0.674 |
| **Logistics & Transportation ( 4%)** | Other fields | 839 | 595 | 0.399 | 2.93 | 2.96 | 0.647 |
| **Social Psychology ( 3.9%)** | Highly related fields | 1303 | 942 | 0.210 | 3.49 | 3.27 | 0.138 |
| **Applied Mathematics ( 3.8%)** | Other fields | 1331 | 942 | 0.112 | 3.3 | 3.15 | 0.499 |
| **Legal & Forensic Medicine ( 3.8%)** | Highly related fields | 172 | 211 | 0.608 | 2.3 | 2.3 | 0.963 |
| **Physical Chemistry ( 3.7%)** | Other fields | 1724 | 1358 | 0.490 | 2.96 | 2.98 | 0.743 |
| **Development Studies ( 3.7%)** | Other fields | 843 | 601 | 0.521 | 3.08 | 3.16 | 0.608 |
| **Economics ( 3.6%)** | Other fields | 1404 | 609 | 0.001 | 3.6 | 3.14 | 0.006 |
| **Anatomy & Morphology ( 3.6%)** | Highly related fields | 373 | 184 | 1.000 | 2.39 | 2.4 | 0.895 |
| **Food Science ( 3.6%)** | Other fields | 1605 | 853 | 0.238 | 2.92 | 2.99 | 0.912 |
| **Ornithology ( 3.4%)** | Other fields | 401 | 353 | 0.811 | 2.95 | 2.68 | 0.189 |
| **Geography ( 3.3%)** | Other fields | 336 | 406 | 0.310 | 2.95 | 3.06 | 0.225 |
| **Electrical & Electronic Engineering ( 3.3%)** | Other fields | 470 | 460 | 0.717 | 2.44 | 2.54 | 0.387 |
| **Networking & Telecommunications ( 3.3%)** | Other fields | 559 | 561 | 0.742 | 2.61 | 2.63 | 0.436 |
| **Dairy & Animal Science ( 3.3%)** | Other fields | 665 | 493 | 0.054 | 2.49 | 2.65 | 0.164 |
| **General Psychology & Cognitive Sciences ( 3.1%)** | Highly related fields | 359 | 583 | 0.203 | 2.74 | 2.99 | 0.083 |
| **Fluids & Plasmas ( 3.1%)** | Other fields | 1146 | 802 | 0.116 | 3.28 | 3.08 | 0.245 |
| **Social Sciences Methods ( 3%)** | Other fields | 3951 | 760 | 0.059 | 4.25 | 3.23 | 0.033 |
| **Numerical & Computational Mathematics ( 3%)** | Other fields | 1112 | 512 | 0.036 | 3.57 | 2.98 | 0.222 |
| **Building & Construction ( 2.9%)** | Other fields | 1504 | 879 | 0.140 | 3.08 | 3.05 | 0.976 |
| **Mechanical Engineering & Transports ( 2.9%)** | Other fields | 759 | 554 | 0.803 | 2.78 | 2.86 | 0.298 |
| **Science Studies ( 2.9%)** | Other fields | 583 | 546 | 0.621 | 3.03 | 3.16 | 0.488 |
| **Cultural Studies ( 2.8%)** | Other fields | 100 | 124 | 0.810 | 2.52 | 2.54 | 0.962 |
| **Marketing ( 2.8%)** | Other fields | 724 | 686 | 0.721 | 3.23 | 3.09 | 0.442 |
| **Anthropology ( 2.6%)** | Other fields | 630 | 411 | 0.113 | 2.88 | 3.01 | 0.334 |
| **Materials ( 2.4%)** | Other fields | 1308 | 721 | 0.004 | 2.98 | 2.82 | 0.142 |
| **Operations Research ( 2.4%)** | Other fields | 819 | 693 | 0.601 | 3.29 | 3.14 | 0.565 |
| **Mining & Metallurgy ( 2.3%)** | Other fields | 202 | 253 | 0.573 | 2.31 | 2.36 | 0.809 |
| **Optoelectronics & Photonics ( 2.3%)** | Other fields | 639 | 373 | 0.104 | 2.57 | 2.41 | 0.164 |
| **Human Factors ( 2.3%)** | Highly related fields | 1129 | 739 | 0.056 | 3.37 | 3.01 | 0.136 |
| **Software Engineering ( 2.2%)** | Other fields | 719 | 622 | 0.746 | 2.65 | 2.8 | 0.149 |
| **Finance ( 2.2%)** | Other fields | 698 | 738 | 0.550 | 3.19 | 3.11 | 0.556 |
| **Meteorology & Atmospheric Sciences ( 2.2%)** | Other fields | 1558 | 1357 | 0.404 | 3.27 | 3.12 | 0.128 |
| **Communication & Media Studies ( 2.1%)** | Other fields | 826 | 450 | 0.062 | 3.22 | 3.07 | 0.406 |
| **Econometrics ( 2%)** | Other fields | 2443 | 1044 | 0.289 | 3.3 | 3.47 | 0.621 |
| **Political Science & Public Administration ( 2%)** | Other fields | 1694 | 446 | 0.044 | 3.43 | 3.07 | 0.059 |
| **Languages & Linguistics ( 2%)** | Other fields | 484 | 360 | 0.412 | 3.32 | 3.05 | 0.196 |
| **General Physics ( 2%)** | Other fields | 629 | 688 | 0.648 | 2.83 | 2.83 | 0.520 |
| **Archaeology ( 2%)** | Other fields | 447 | 360 | 0.567 | 2.91 | 2.78 | 0.609 |
| **Sport, Leisure & Tourism ( 1.9%)** | Other fields | 790 | 653 | 0.776 | 3.25 | 3.09 | 0.478 |
| **Environmental Engineering ( 1.9%)** | Other fields | 1266 | 787 | 0.089 | 2.9 | 2.97 | 0.825 |
| **Energy ( 1.9%)** | Other fields | 1030 | 785 | 0.023 | 2.85 | 2.87 | 0.974 |
| **Marine Biology & Hydrobiology ( 1.7%)** | Other fields | 1053 | 929 | 0.158 | 3.12 | 3.07 | 0.832 |
| **Ecology ( 1.6%)** | Other fields | 951 | 1224 | 0.703 | 3.18 | 3.16 | 0.482 |
| **Chemical Engineering ( 1.6%)** | Other fields | 1314 | 878 | 0.802 | 3.35 | 2.96 | 0.173 |
| **Philosophy ( 1.6%)** | Other fields | 736 | 200 | 0.023 | 3.65 | 2.86 | 0.020 |
| **Aerospace & Aeronautics ( 1.6%)** | Other fields | 371 | 290 | 0.660 | 2.52 | 2.5 | 0.732 |
| **Inorganic & Nuclear Chemistry ( 1.5%)** | Other fields | 920 | 757 | 0.769 | 2.81 | 2.78 | 0.784 |
| **Agricultural Economics & Policy ( 1.4%)** | Other fields | 911 | 732 | 0.651 | 3.23 | 3.08 | 0.547 |
| **Geological & Geomatics Engineering ( 1.4%)** | Other fields | 1457 | 963 | 0.562 | 3.3 | 3.07 | 0.248 |
| **Accounting ( 1.4%)** | Other fields | 494 | 713 | 0.537 | 3.1 | 3.09 | 1.000 |
| **Applied Physics ( 1.2%)** | Other fields | 945 | 886 | 0.262 | 2.71 | 2.82 | 0.686 |
| **Business & Management ( 1.2%)** | Other fields | 1052 | 791 | 0.223 | 3.15 | 3.17 | 0.682 |
| **Information Systems ( 1.2%)** | Other fields | 1793 | 898 | 0.269 | 3.49 | 3.06 | 0.110 |
| **Computation Theory & Mathematics ( 1.1%)** | Other fields | 607 | 517 | 0.665 | 3.11 | 2.94 | 0.485 |
| **Astronomy & Astrophysics ( .94%)** | Other fields | 2954 | 1751 | 0.397 | 3.11 | 3.14 | 0.820 |
| **Geochemistry & Geophysics ( .81%)** | Other fields | 755 | 868 | 0.694 | 3.14 | 3.15 | 0.653 |
| **Strategic, Defence & Security Studies ( .77%)** | Other fields | 159 | 408 | 0.139 | 2.84 | 2.86 | 0.926 |
| **Paleontology ( .74%)** | Other fields | 496 | 803 | 0.279 | 2.88 | 3.1 | 0.143 |
| **Literary Studies ( .72%)** | Other fields | 143 | 44 | 0.163 | 2.75 | 2 | 0.129 |
| **General Mathematics ( .71%)** | Other fields | 267 | 298 | 0.703 | 2.95 | 2.85 | 0.295 |
| **Nuclear & Particle Physics ( .63%)** | Other fields | 3204 | 759 | 0.085 | 3.2 | 2.9 | 0.062 |
| **Oceanography ( .6%)** | Other fields | 1077 | 654 | 0.307 | 2.71 | 2.93 | 0.184 |
| **Agronomy & Agriculture ( .55%)** | Other fields | 1898 | 771 | 0.268 | 3.34 | 2.89 | 0.078 |
| **Fisheries ( .53%)** | Other fields | 1170 | 593 | 0.201 | 2.99 | 2.8 | 0.481 |
| **Zoology ( 0%)** | Other fields | . | 337 |  | . | 2.58 |  |
| **Urban & Regional Planning ( 0%)** | Other fields | . | 428 |  | . | 3.16 |  |
| **Music ( 0%)** | Other fields | . | 52 |  | . | 2.06 |  |
| **Mathematical Physics ( 0%)** | Other fields | . | 358 |  | . | 2.88 |  |
| **Law ( 0%)** | Other fields | . | 138 |  | . | 2.48 |  |
| **International Relations ( 0%)** | Other fields | . | 343 |  | . | 3.01 |  |
| **Industrial Relations ( 0%)** | Other fields | . | 218 |  | . | 2.87 |  |
| **Horticulture ( 0%)** | Other fields | . | 396 |  | . | 2.64 |  |
| **History of Social Sciences ( 0%)** | Other fields | . | 197 |  | . | 2.75 |  |
| **History of Science, Technology & Medicine ( 0%)** | Other fields | . | 68 |  | . | 2.27 |  |
| **History ( 0%)** | Other fields | . | 50 |  | . | 2.08 |  |
| **Geology ( 0%)** | Other fields | . | 490 |  | . | 2.89 |  |
| **Gender Studies ( 0%)** | Highly related fields | . | 360 |  | . | 3.02 |  |
| **Forestry ( 0%)** | Other fields | . | 492 |  | . | 2.65 |  |
| **Folklore ( 0%)** | Other fields | . | 45 |  | . | 2.1 |  |
| **Economic Theory ( 0%)** | Other fields | . | 323 |  | . | 2.93 |  |
| **Classics ( 0%)** | Other fields | . | 41 |  | . | 2.04 |  |
| **Civil Engineering ( 0%)** | Other fields | . | 564 |  | . | 2.84 |  |
| **Automobile Design & Engineering ( 0%)** | Other fields | . | 122 |  | . | 2.18 |  |
| **Art Practice, History & Theory ( 0%)** | Other fields | . | 36 |  | . | 1.88 |  |
| **Architecture ( 0%)** | Other fields | . | 74 |  | . | 2.12 |  |

**Supplementary Table 3.1 : Career-long impact, Funding time any funding Linear Regressions for each subfield (ordered by percentage funded)**

| **Top-cited US-based researchers: Subfield (perc. funded)** | **Classification** | **Dependent Variable** | **Constant (p-val)** | **Funded (p-val)** | **Years since first pub (p-val)** |
| --- | --- | --- | --- | --- | --- |
| **Developmental Biology ( 89%)** | **Highly related fields** | Raw citations | 16567 (<0.001) | 4342 ( 0.026) | 17.1 ( 0.773) |
|  |  | Composite | 3.53 (<0.001) | .0816 ( 0.001) | .00752 (<0.001) |
| **Substance Abuse ( 87%)** | **Highly related fields** | Raw citations | 2560 ( 0.207) | 2396 ( 0.055) | 134 ( 0.001) |
|  |  | Composite | 3.52 (<0.001) | .0959 ( 0.035) | .00466 ( 0.001) |
| **Immunology ( 85%)** | **Highly related fields** | Raw citations | 14839 (<0.001) | 3606 ( 0.004) | 32.7 ( 0.412) |
|  |  | Composite | 3.6 (<0.001) | .129 (<0.001) | .00406 (<0.001) |
| **Geriatrics ( 85%)** | **Highly related fields** | Raw citations | 5149 ( 0.361) | 6804 ( 0.052) | 50.5 ( 0.711) |
|  |  | Composite | 3.33 (<0.001) | .125 ( 0.179) | .0075 ( 0.041) |
| **Biochemistry & Molecular Biology ( 81%)** | **Highly related fields** | Raw citations | 9394 (<0.001) | 3582 (<0.001) | 25.5 ( 0.287) |
|  |  | Composite | 3.57 (<0.001) | .118 (<0.001) | .00382 (<0.001) |
| **Endocrinology & Metabolism ( 81%)** | **Highly related fields** | Raw citations | 10291 (<0.001) | 4340 (<0.001) | 50 ( 0.226) |
|  |  | Composite | 3.6 (<0.001) | .11 (<0.001) | .00405 (<0.001) |
| **Gerontology ( 80%)** | **Highly related fields** | Raw citations | 6911 ( 0.466) | 8101 ( 0.087) | -39.8 ( 0.837) |
|  |  | Composite | 3.34 (<0.001) | .205 ( 0.003) | .00533 ( 0.060) |
| **Virology ( 80%)** | **Highly related fields** | Raw citations | 6774 (<0.001) | 3764 (<0.001) | 78.1 ( 0.027) |
|  |  | Composite | 3.39 (<0.001) | .1 (<0.001) | .00534 (<0.001) |
| **Neurology & Neurosurgery ( 79%)** | **Highly related fields** | Raw citations | 9504 (<0.001) | 5083 (<0.001) | 46.2 ( 0.043) |
|  |  | Composite | 3.55 (<0.001) | .131 (<0.001) | .00482 (<0.001) |
| **Genetics & Heredity ( 79%)** | **Highly related fields** | Raw citations | 17597 (<0.001) | 4701 ( 0.019) | -97.5 ( 0.216) |
|  |  | Composite | 3.51 (<0.001) | .0698 ( 0.047) | .00414 ( 0.003) |
| **Bioinformatics ( 77%)** | **Highly related fields** | Raw citations | 15469 ( 0.024) | 6025 ( 0.149) | -27.7 ( 0.871) |
|  |  | Composite | 3.37 (<0.001) | .127 ( 0.016) | .00574 ( 0.008) |
| **Psychiatry ( 76%)** | **Highly related fields** | Raw citations | 4549 ( 0.067) | 7887 (<0.001) | 149 ( 0.003) |
|  |  | Composite | 3.56 (<0.001) | .15 (<0.001) | .00527 (<0.001) |
| **Oncology & Carcinogenesis ( 74%)** | **Highly related fields** | Raw citations | 14805 (<0.001) | 4412 (<0.001) | 18.4 ( 0.558) |
|  |  | Composite | 3.41 (<0.001) | .119 (<0.001) | .00628 (<0.001) |
| **Public Health ( 73%)** | **Highly related fields** | Raw citations | 4933 ( 0.004) | 872 ( 0.355) | 142 (<0.001) |
|  |  | Composite | 3.45 (<0.001) | .0795 ( 0.001) | .00587 (<0.001) |
| **Medical Informatics ( 71%)** | **Highly related fields** | Raw citations | 3995 ( 0.098) | 2189 ( 0.151) | 5.3 ( 0.926) |
|  |  | Composite | 3.18 (<0.001) | .142 ( 0.025) | .00323 ( 0.170) |
| **Demography ( 71%)** | **Highly related fields** | Raw citations | -107 ( 0.968) | 467 ( 0.750) | 103 ( 0.061) |
|  |  | Composite | 3.32 (<0.001) | .0289 ( 0.739) | .00681 ( 0.037) |
| **Physiology ( 71%)** | **Highly related fields** | Raw citations | 2908 ( 0.028) | 3168 (<0.001) | 58.6 ( 0.015) |
|  |  | Composite | 3.63 (<0.001) | .0918 ( 0.007) | .00195 ( 0.095) |
| **Epidemiology ( 71%)** | **Highly related fields** | Raw citations | -4950 ( 0.668) | 11292 ( 0.058) | 483 ( 0.055) |
|  |  | Composite | 3.5 (<0.001) | .0814 ( 0.173) | .00859 ( 0.001) |
| **Biomedical Engineering ( 71%)** | **Highly related fields** | Raw citations | 1657 ( 0.373) | 5018 (<0.001) | 84.3 ( 0.034) |
|  |  | Composite | 3.07 (<0.001) | .167 (<0.001) | .00764 (<0.001) |
| **Developmental & Child Psychology ( 70%)** | **Highly related fields** | Raw citations | 1724 ( 0.326) | 5275 (<0.001) | 119 ( 0.002) |
|  |  | Composite | 3.48 (<0.001) | .164 (<0.001) | .00591 (<0.001) |
| **Arthritis & Rheumatology ( 70%)** | **Highly related fields** | Raw citations | 5392 ( 0.189) | 5538 ( 0.006) | 199 ( 0.021) |
|  |  | Composite | 3.49 (<0.001) | .139 ( 0.003) | .00647 ( 0.001) |
| **Allergy ( 69%)** | **Highly related fields** | Raw citations | 4591 ( 0.205) | 5965 ( 0.001) | 86.9 ( 0.238) |
|  |  | Composite | 3.48 (<0.001) | .162 ( 0.004) | .00481 ( 0.032) |
| **Biophysics ( 69%)** | **Highly related fields** | Raw citations | 654 ( 0.840) | 6523 (<0.001) | 91.3 ( 0.161) |
|  |  | Composite | 3.1 (<0.001) | .297 (<0.001) | .007 (<0.001) |
| **Respiratory System ( 66%)** | **Highly related fields** | Raw citations | 8992 (<0.001) | 2276 ( 0.022) | 89.3 ( 0.047) |
|  |  | Composite | 3.53 (<0.001) | .0332 ( 0.180) | .00476 (<0.001) |
| **Experimental Psychology ( 65%)** | **Highly related fields** | Raw citations | 4684 (<0.001) | 3123 (<0.001) | 53.5 ( 0.023) |
|  |  | Composite | 3.62 (<0.001) | .0967 (<0.001) | .00366 (<0.001) |
| **Gastroenterology & Hepatology ( 65%)** | **Highly related fields** | Raw citations | 9502 (<0.001) | 3471 (<0.001) | 42.2 ( 0.300) |
|  |  | Composite | 3.55 (<0.001) | .107 (<0.001) | .00381 (<0.001) |
| **Cardiovascular System & Hematology ( 64%)** | **Highly related fields** | Raw citations | 18376 (<0.001) | 3143 ( 0.002) | 6.02 ( 0.886) |
|  |  | Composite | 3.55 (<0.001) | .102 (<0.001) | .00485 (<0.001) |
| **Health Policy & Services ( 63%)** | **Highly related fields** | Raw citations | -284 ( 0.914) | 3052 ( 0.026) | 222 ( 0.001) |
|  |  | Composite | 3.23 (<0.001) | .117 ( 0.003) | .0104 (<0.001) |
| **Urology & Nephrology ( 63%)** | **Highly related fields** | Raw citations | 10978 (<0.001) | 4202 (<0.001) | -29 ( 0.431) |
|  |  | Composite | 3.49 (<0.001) | .138 (<0.001) | .00345 (<0.001) |
| **Speech-Language Pathology & Audiology ( 61%)** | **Highly related fields** | Raw citations | 6842 (<0.001) | -278 ( 0.724) | -49.7 ( 0.243) |
|  |  | Composite | 3.38 (<0.001) | -.0379 ( 0.429) | .00447 ( 0.085) |
| **Clinical Psychology ( 60%)** | **Highly related fields** | Raw citations | 1024 ( 0.705) | 6703 (<0.001) | 154 ( 0.004) |
|  |  | Composite | 3.57 (<0.001) | .167 (<0.001) | .00476 ( 0.006) |
| **Pediatrics ( 60%)** | **Highly related fields** | Raw citations | 4647 (<0.001) | 2414 (<0.001) | 27 ( 0.140) |
|  |  | Composite | 3.21 (<0.001) | .13 (<0.001) | .00486 (<0.001) |
| **Microbiology ( 60%)** | **Highly related fields** | Raw citations | 11977 (<0.001) | 2368 (<0.001) | -39.4 ( 0.119) |
|  |  | Composite | 3.5 (<0.001) | .0787 (<0.001) | .00453 (<0.001) |
| **Family Studies ( 59%)** | **Other fields** | Raw citations | -751 ( 0.596) | 1672 ( 0.027) | 98.9 ( 0.004) |
|  |  | Composite | 3.11 (<0.001) | .111 ( 0.052) | .0101 (<0.001) |
| **Applied Ethics ( 58%)** | **Highly related fields** | Raw citations | 5693 (<0.001) | 1671 ( 0.042) | -44.2 ( 0.245) |
|  |  | Composite | 3.47 (<0.001) | .0812 ( 0.170) | .00286 ( 0.298) |
| **Nutrition & Dietetics ( 57%)** | **Highly related fields** | Raw citations | 6617 ( 0.002) | 2723 ( 0.013) | 48.5 ( 0.282) |
|  |  | Composite | 3.42 (<0.001) | .137 (<0.001) | .00607 (<0.001) |
| **Ophthalmology & Optometry ( 57%)** | **Highly related fields** | Raw citations | 8300 (<0.001) | 3543 (<0.001) | -41.7 ( 0.098) |
|  |  | Composite | 3.43 (<0.001) | .122 (<0.001) | .00222 ( 0.012) |
| **Organic Chemistry ( 57%)** | **Other fields** | Raw citations | 7114 (<0.001) | 3134 (<0.001) | 23.3 ( 0.409) |
|  |  | Composite | 3.41 (<0.001) | .165 (<0.001) | .00458 (<0.001) |
| **Environmental & Occupational Health ( 57%)** | **Highly related fields** | Raw citations | 3908 ( 0.081) | 750 ( 0.389) | 21.3 ( 0.674) |
|  |  | Composite | 3.1 (<0.001) | .122 ( 0.004) | .00503 ( 0.039) |
| **Toxicology ( 56%)** | **Highly related fields** | Raw citations | 8041 (<0.001) | 2954 (<0.001) | -49 ( 0.110) |
|  |  | Composite | 3.32 (<0.001) | .115 (<0.001) | .00365 ( 0.003) |
| **Nursing ( 55%)** | **Highly related fields** | Raw citations | 770 ( 0.045) | 1412 (<0.001) | 29.4 ( 0.001) |
|  |  | Composite | 2.97 (<0.001) | .0966 (<0.001) | .00355 (<0.001) |
| **Obstetrics & Reproductive Medicine ( 54%)** | **Highly related fields** | Raw citations | 7477 (<0.001) | 3282 (<0.001) | -28.4 ( 0.199) |
|  |  | Composite | 3.41 (<0.001) | .141 (<0.001) | .0026 ( 0.004) |
| **Emergency & Critical Care Medicine ( 53%)** | **Highly related fields** | Raw citations | 2569 ( 0.322) | 6546 (<0.001) | 124 ( 0.051) |
|  |  | Composite | 3.22 (<0.001) | .149 (<0.001) | .00724 (<0.001) |
| **Rehabilitation ( 53%)** | **Highly related fields** | Raw citations | 3170 ( 0.012) | 1469 ( 0.009) | 39.9 ( 0.173) |
|  |  | Composite | 3.22 (<0.001) | .0989 ( 0.004) | .00559 ( 0.002) |
| **Pharmacology & Pharmacy ( 51%)** | **Highly related fields** | Raw citations | 3751 (<0.001) | 3041 (<0.001) | 30 ( 0.097) |
|  |  | Composite | 3.24 (<0.001) | .148 (<0.001) | .00362 (<0.001) |
| **Behavioral Science & Comparative Psychology ( 50%)** | **Highly related fields** | Raw citations | 3640 ( 0.004) | 2043 ( 0.001) | 29.3 ( 0.276) |
|  |  | Composite | 3.55 (<0.001) | .0796 ( 0.021) | .0042 ( 0.008) |
| **Analytical Chemistry ( 49%)** | **Other fields** | Raw citations | 3742 ( 0.001) | 4361 (<0.001) | 30.1 ( 0.213) |
|  |  | Composite | 3.18 (<0.001) | .169 (<0.001) | .00555 (<0.001) |
| **Nuclear Medicine & Medical Imaging ( 47%)** | **Highly related fields** | Raw citations | 5043 (<0.001) | 4177 (<0.001) | 37.7 ( 0.078) |
|  |  | Composite | 3.32 (<0.001) | .163 (<0.001) | .00282 (<0.001) |
| **Otorhinolaryngology ( 44%)** | **Highly related fields** | Raw citations | 5626 (<0.001) | 759 ( 0.022) | -31.3 ( 0.023) |
|  |  | Composite | 3.28 (<0.001) | .0652 ( 0.002) | .00245 ( 0.005) |
| **Dentistry ( 44%)** | **Highly related fields** | Raw citations | 4145 (<0.001) | 2233 (<0.001) | -4.44 ( 0.786) |
|  |  | Composite | 3.3 (<0.001) | .101 (<0.001) | .00277 ( 0.006) |
| **Tropical Medicine ( 44%)** | **Highly related fields** | Raw citations | 6474 (<0.001) | 3029 ( 0.001) | -22.9 ( 0.478) |
|  |  | Composite | 3.31 (<0.001) | .128 ( 0.001) | .00207 ( 0.153) |
| **Mycology & Parasitology ( 43%)** | **Highly related fields** | Raw citations | 8260 (<0.001) | -105 ( 0.922) | -41.5 ( 0.324) |
|  |  | Composite | 3.4 (<0.001) | .00709 ( 0.876) | .00229 ( 0.201) |
| **Statistics & Probability ( 43%)** | **Other fields** | Raw citations | 9377 ( 0.004) | 4827 ( 0.009) | 13.3 ( 0.840) |
|  |  | Composite | 3.65 (<0.001) | .122 ( 0.002) | .00221 ( 0.114) |
| **Medicinal & Biomolecular Chemistry ( 43%)** | **Highly related fields** | Raw citations | 814 ( 0.356) | 2946 (<0.001) | 108 (<0.001) |
|  |  | Composite | 2.87 (<0.001) | .186 (<0.001) | .00879 (<0.001) |
| **Anesthesiology ( 43%)** | **Highly related fields** | Raw citations | 4603 (<0.001) | 3137 (<0.001) | 20.7 ( 0.425) |
|  |  | Composite | 3.32 (<0.001) | .148 (<0.001) | .00319 ( 0.017) |
| **Dermatology & Venereal Diseases ( 42%)** | **Highly related fields** | Raw citations | 7447 (<0.001) | 4780 (<0.001) | -19.5 ( 0.513) |
|  |  | Composite | 3.56 (<0.001) | .132 (<0.001) | .000994 ( 0.383) |
| **Biotechnology ( 41%)** | **Highly related fields** | Raw citations | 2604 ( 0.117) | 3341 ( 0.001) | 104 ( 0.016) |
|  |  | Composite | 3.06 (<0.001) | .151 (<0.001) | .00882 (<0.001) |
| **Social Psychology ( 41%)** | **Highly related fields** | Raw citations | 4642 ( 0.005) | 5428 (<0.001) | 97.2 ( 0.004) |
|  |  | Composite | 3.63 (<0.001) | .158 (<0.001) | .00462 (<0.001) |
| **Surgery ( 40%)** | **Highly related fields** | Raw citations | 6990 (<0.001) | 4518 (<0.001) | -1.68 ( 0.920) |
|  |  | Composite | 3.37 (<0.001) | .129 (<0.001) | .00171 ( 0.006) |
| **General & Internal Medicine ( 40%)** | **Highly related fields** | Raw citations | 10778 (<0.001) | 4012 (<0.001) | -87.6 (<0.001) |
|  |  | Composite | 3.16 (<0.001) | .224 (<0.001) | .00147 ( 0.010) |
| **Complementary & Alternative Medicine ( 40%)** | **Highly related fields** | Raw citations | 1654 ( 0.048) | 2040 (<0.001) | -3.94 ( 0.869) |
|  |  | Composite | 2.96 (<0.001) | .163 ( 0.037) | -.000779 ( 0.833) |
| **Sport Sciences ( 37%)** | **Highly related fields** | Raw citations | 2289 ( 0.288) | 3834 ( 0.003) | 111 ( 0.046) |
|  |  | Composite | 3.41 (<0.001) | .0891 ( 0.041) | .00419 ( 0.026) |
| **Acoustics ( 36%)** | **Other fields** | Raw citations | 5495 (<0.001) | 885 ( 0.035) | -35.8 ( 0.013) |
|  |  | Composite | 3.51 (<0.001) | .0277 ( 0.371) | -.000713 ( 0.499) |
| **History of Social Sciences ( 35%)** | **Other fields** | Raw citations | 194 ( 0.941) | -1365 ( 0.290) | 67.3 ( 0.232) |
|  |  | Composite | 2.87 (<0.001) | .0155 ( 0.901) | .012 ( 0.035) |
| **Nanoscience & Nanotechnology ( 35%)** | **Other fields** | Raw citations | 6113 ( 0.039) | 4857 ( 0.031) | 493 (<0.001) |
|  |  | Composite | 3.28 (<0.001) | .0836 ( 0.021) | .0142 (<0.001) |
| **General Chemistry ( 34%)** | **Other fields** | Raw citations | 9904 (<0.001) | 7159 (<0.001) | -43.9 ( 0.222) |
|  |  | Composite | 3.18 (<0.001) | .358 (<0.001) | .00393 (<0.001) |
| **Microscopy ( 33%)** | **Highly related fields** | Raw citations | 5132 ( 0.173) | 3678 ( 0.057) | 21 ( 0.803) |
|  |  | Composite | 3.26 (<0.001) | .149 ( 0.220) | .00483 ( 0.376) |
| **Criminology ( 31%)** | **Other fields** | Raw citations | 2645 ( 0.017) | 1543 ( 0.022) | 51.3 ( 0.068) |
|  |  | Composite | 3.49 (<0.001) | .0521 ( 0.206) | .00419 ( 0.016) |
| **Sociology ( 31%)** | **Other fields** | Raw citations | 2403 ( 0.014) | 778 ( 0.228) | 60.8 ( 0.007) |
|  |  | Composite | 3.59 (<0.001) | -.0261 ( 0.487) | .00395 ( 0.003) |
| **Optics ( 30%)** | **Other fields** | Raw citations | 7771 (<0.001) | 2447 ( 0.016) | .845 ( 0.983) |
|  |  | Composite | 3.23 (<0.001) | .0743 ( 0.043) | .00524 (<0.001) |
| **Pathology ( 30%)** | **Highly related fields** | Raw citations | 14703 (<0.001) | 910 ( 0.494) | -68.1 ( 0.196) |
|  |  | Composite | 3.57 (<0.001) | .0283 ( 0.477) | .00272 ( 0.084) |
| **Social Work ( 30%)** | **Other fields** | Raw citations | 907 ( 0.155) | 856 ( 0.018) | 27.3 ( 0.124) |
|  |  | Composite | 3.2 (<0.001) | .0656 ( 0.161) | .00199 ( 0.389) |
| **Evolutionary Biology ( 29%)** | **Other fields** | Raw citations | 3332 ( 0.154) | 6642 (<0.001) | 136 ( 0.018) |
|  |  | Composite | 3.59 (<0.001) | .116 (<0.001) | .0064 (<0.001) |
| **Orthopedics ( 28%)** | **Highly related fields** | Raw citations | 6404 (<0.001) | 4295 (<0.001) | 2.44 ( 0.896) |
|  |  | Composite | 3.34 (<0.001) | .193 (<0.001) | .00449 (<0.001) |
| **Social Sciences Methods ( 27%)** | **Other fields** | Raw citations | 1935 ( 0.652) | 13613 (<0.001) | 89.3 ( 0.285) |
|  |  | Composite | 3.53 (<0.001) | .317 (<0.001) | .00454 ( 0.015) |
| **Chemical Physics ( 27%)** | **Other fields** | Raw citations | 9130 (<0.001) | 7215 (<0.001) | 63.1 ( 0.069) |
|  |  | Composite | 3.73 (<0.001) | .123 (<0.001) | .00235 ( 0.001) |
| **General Clinical Medicine ( 24%)** | **Highly related fields** | Raw citations | 3799 ( 0.019) | 4497 (<0.001) | -11.6 ( 0.734) |
|  |  | Composite | 2.91 (<0.001) | .334 (<0.001) | .00431 ( 0.060) |
| **Plant Biology & Botany ( 24%)** | **Other fields** | Raw citations | 7253 (<0.001) | 4304 (<0.001) | -8.13 ( 0.695) |
|  |  | Composite | 3.41 (<0.001) | .181 (<0.001) | .00327 (<0.001) |
| **Inorganic & Nuclear Chemistry ( 24%)** | **Other fields** | Raw citations | 4970 ( 0.013) | 2307 ( 0.071) | 72.1 ( 0.070) |
|  |  | Composite | 3.33 (<0.001) | .151 (<0.001) | .00539 (<0.001) |
| **Veterinary Sciences ( 23%)** | **Highly related fields** | Raw citations | 3034 (<0.001) | 1192 (<0.001) | 17.2 ( 0.175) |
|  |  | Composite | 3.23 (<0.001) | .0519 ( 0.014) | .00282 ( 0.002) |
| **Environmental Sciences ( 21%)** | **Other fields** | Raw citations | 5054 (<0.001) | 3869 (<0.001) | 26.2 ( 0.423) |
|  |  | Composite | 3.25 (<0.001) | .166 (<0.001) | .00519 (<0.001) |
| **Human Factors ( 19%)** | **Highly related fields** | Raw citations | 4463 (<0.001) | 226 ( 0.807) | 61.7 ( 0.080) |
|  |  | Composite | 3.41 (<0.001) | .00886 ( 0.871) | .00705 ( 0.001) |
| **Ornithology ( 18%)** | **Other fields** | Raw citations | 3738 ( 0.001) | -922 ( 0.243) | -1.63 ( 0.941) |
|  |  | Composite | 3.27 (<0.001) | .00966 ( 0.902) | .00468 ( 0.041) |
| **Urban & Regional Planning ( 18%)** | **Other fields** | Raw citations | 3631 ( 0.007) | 758 ( 0.438) | -3.28 ( 0.920) |
|  |  | Composite | 3.56 (<0.001) | .101 ( 0.278) | .000996 ( 0.747) |
| **Economics ( 18%)** | **Other fields** | Raw citations | 6258 (<0.001) | 898 ( 0.283) | 13.1 ( 0.591) |
|  |  | Composite | 3.61 (<0.001) | .0616 ( 0.080) | .00437 (<0.001) |
| **Artificial Intelligence & Image Processing ( 17%)** | **Other fields** | Raw citations | 5478 (<0.001) | 2503 ( 0.001) | 59.9 ( 0.019) |
|  |  | Composite | 3.18 (<0.001) | .0877 ( 0.001) | .0081 (<0.001) |
| **Polymers ( 17%)** | **Other fields** | Raw citations | 8319 (<0.001) | 3998 ( 0.002) | -5.84 ( 0.855) |
|  |  | Composite | 3.49 (<0.001) | .0735 ( 0.080) | .0019 ( 0.073) |
| **Economic Theory ( 17%)** | **Other fields** | Raw citations | 2779 ( 0.139) | -472 ( 0.692) | 21.1 ( 0.588) |
|  |  | Composite | 3.52 (<0.001) | -.0314 ( 0.705) | .00233 ( 0.392) |
| **Development Studies ( 17%)** | **Other fields** | Raw citations | 5171 ( 0.023) | -32.7 ( 0.977) | -36.8 ( 0.498) |
|  |  | Composite | 3.9 (<0.001) | -.114 ( 0.390) | -.00629 ( 0.323) |
| **Anatomy & Morphology ( 17%)** | **Highly related fields** | Raw citations | 2427 ( 0.015) | 871 ( 0.373) | -7.35 ( 0.737) |
|  |  | Composite | 2.97 (<0.001) | -.0567 ( 0.556) | .00418 ( 0.057) |
| **Distributed Computing ( 17%)** | **Other fields** | Raw citations | 2065 ( 0.205) | 5045 (<0.001) | 61.3 ( 0.206) |
|  |  | Composite | 3.13 (<0.001) | .148 ( 0.019) | .00228 ( 0.413) |
| **Entomology ( 16%)** | **Other fields** | Raw citations | 4528 (<0.001) | 2884 (<0.001) | -13.2 ( 0.357) |
|  |  | Composite | 3.49 (<0.001) | .126 ( 0.001) | .000296 ( 0.791) |
| **Gender Studies ( 15%)** | **Highly related fields** | Raw citations | 3224 ( 0.175) | 149 ( 0.898) | -10.6 ( 0.851) |
|  |  | Composite | 3.35 (<0.001) | .0864 ( 0.506) | .00425 ( 0.503) |
| **History of Science, Technology & Medicine ( 15%)** | **Other fields** | Raw citations | 350 ( 0.673) | -115 ( 0.852) | 15.6 ( 0.325) |
|  |  | Composite | 3 (<0.001) | -.00475 ( 0.974) | .00379 ( 0.310) |
| **Software Engineering ( 15%)** | **Other fields** | Raw citations | 7404 (<0.001) | 907 ( 0.164) | -62.1 ( 0.012) |
|  |  | Composite | 3.51 (<0.001) | .00865 ( 0.853) | .000547 ( 0.757) |
| **Education ( 15%)** | **Other fields** | Raw citations | 1643 (<0.001) | 1753 (<0.001) | 42 (<0.001) |
|  |  | Composite | 3.24 (<0.001) | .0881 ( 0.001) | .00489 (<0.001) |
| **Food Science ( 15%)** | **Other fields** | Raw citations | 4993 (<0.001) | 2336 ( 0.041) | 11.4 ( 0.708) |
|  |  | Composite | 3.31 (<0.001) | .166 ( 0.002) | .0033 ( 0.020) |
| **Industrial Engineering & Automation ( 14%)** | **Other fields** | Raw citations | 5629 (<0.001) | 1686 ( 0.013) | 6.56 ( 0.752) |
|  |  | Composite | 3.38 (<0.001) | .0786 ( 0.046) | .00349 ( 0.004) |
| **Logistics & Transportation ( 14%)** | **Other fields** | Raw citations | 3226 (<0.001) | -771 ( 0.226) | 14.3 ( 0.469) |
|  |  | Composite | 3.23 (<0.001) | -.117 ( 0.065) | .00419 ( 0.033) |
| **Geography ( 14%)** | **Other fields** | Raw citations | 1986 ( 0.073) | 2635 ( 0.004) | 52.1 ( 0.062) |
|  |  | Composite | 3.51 (<0.001) | .228 ( 0.001) | .00512 ( 0.014) |
| **Sport, Leisure & Tourism ( 13%)** | **Other fields** | Raw citations | 3113 ( 0.010) | -1394 ( 0.197) | 52.1 ( 0.184) |
|  |  | Composite | 3.44 (<0.001) | -.0131 ( 0.882) | .00616 ( 0.058) |
| **Numerical & Computational Mathematics ( 13%)** | **Other fields** | Raw citations | 1444 ( 0.357) | 2179 ( 0.066) | 79 ( 0.025) |
|  |  | Composite | 3.58 (<0.001) | -.0119 ( 0.864) | .00121 ( 0.562) |
| **Communication & Media Studies ( 12%)** | **Other fields** | Raw citations | 1049 ( 0.258) | -254 ( 0.807) | 81.3 ( 0.002) |
|  |  | Composite | 3.45 (<0.001) | .0128 ( 0.821) | .00336 ( 0.016) |
| **General Psychology & Cognitive Sciences ( 12%)** | **Highly related fields** | Raw citations | 2790 ( 0.007) | 547 ( 0.553) | 28.1 ( 0.254) |
|  |  | Composite | 3.36 (<0.001) | .0861 ( 0.349) | .0043 ( 0.082) |
| **Fluids & Plasmas ( 12%)** | **Other fields** | Raw citations | 13332 (<0.001) | 954 ( 0.625) | -79.2 ( 0.131) |
|  |  | Composite | 3.73 (<0.001) | .0502 ( 0.271) | .00145 ( 0.236) |
| **Information & Library Sciences ( 12%)** | **Other fields** | Raw citations | 422 ( 0.471) | 1575 ( 0.011) | 46.9 ( 0.005) |
|  |  | Composite | 2.99 (<0.001) | .187 ( 0.021) | .00712 ( 0.001) |
| **Dairy & Animal Science ( 12%)** | **Other fields** | Raw citations | 4387 (<0.001) | 1320 ( 0.004) | -11 ( 0.357) |
|  |  | Composite | 3.26 (<0.001) | .0617 ( 0.123) | .00129 ( 0.223) |
| **Optoelectronics & Photonics ( 12%)** | **Other fields** | Raw citations | 3186 (<0.001) | 1654 ( 0.001) | 27.5 ( 0.064) |
|  |  | Composite | 2.99 (<0.001) | .0624 ( 0.068) | .00574 (<0.001) |
| **Anthropology ( 11%)** | **Other fields** | Raw citations | 2312 ( 0.012) | 299 ( 0.648) | 39 ( 0.058) |
|  |  | Composite | 3.48 (<0.001) | .0231 ( 0.691) | .00405 ( 0.027) |
| **Religions & Theology ( 11%)** | **Other fields** | Raw citations | 694 ( 0.094) | 1209 ( 0.017) | 1.88 ( 0.856) |
|  |  | Composite | 2.9 (<0.001) | .135 ( 0.189) | .000388 ( 0.855) |
| **Drama & Theater ( 11%)** | **Other fields** | Raw citations | -188 ( 0.513) | 379 ( 0.116) | 21.8 ( 0.051) |
|  |  | Composite | 2.24 (<0.001) | .246 ( 0.204) | .0169 ( 0.065) |
| **Design Practice & Management ( 11%)** | **Other fields** | Raw citations | 3428 (<0.001) | 984 ( 0.267) | 8.29 ( 0.679) |
|  |  | Composite | 3.26 (<0.001) | .0088 ( 0.920) | .00201 ( 0.311) |
| **Environmental Engineering ( 11%)** | **Other fields** | Raw citations | 5204 (<0.001) | 2518 ( 0.001) | .271 ( 0.989) |
|  |  | Composite | 3.39 (<0.001) | .0792 ( 0.066) | .00295 ( 0.015) |
| **Chemical Engineering ( 11%)** | **Other fields** | Raw citations | 7541 (<0.001) | 1228 ( 0.213) | -40.3 ( 0.055) |
|  |  | Composite | 3.45 (<0.001) | .0426 ( 0.454) | .000511 ( 0.672) |
| **Applied Mathematics ( 11%)** | **Other fields** | Raw citations | 3721 ( 0.224) | 2946 ( 0.293) | 99.7 ( 0.149) |
|  |  | Composite | 3.51 (<0.001) | .0474 ( 0.637) | .0051 ( 0.040) |
| **Marketing ( 11%)** | **Other fields** | Raw citations | 2260 ( 0.276) | 760 ( 0.654) | 157 ( 0.008) |
|  |  | Composite | 3.55 (<0.001) | .083 ( 0.218) | .0048 ( 0.040) |
| **Zoology ( 10%)** | **Other fields** | Raw citations | 2160 ( 0.012) | 547 ( 0.489) | 14.3 ( 0.523) |
|  |  | Composite | 3.17 (<0.001) | .0279 ( 0.706) | .00178 ( 0.396) |
| **Architecture ( 10%)** | **Other fields** | Raw citations | -1503 ( 0.446) | -1246 ( 0.608) | 82.4 ( 0.166) |
|  |  | Composite | 2.63 (<0.001) | -.112 ( 0.706) | -.00186 ( 0.784) |
| **Ecology ( 9.7%)** | **Other fields** | Raw citations | 6330 (<0.001) | 1125 ( 0.370) | 116 ( 0.003) |
|  |  | Composite | 3.55 (<0.001) | .114 ( 0.001) | .00723 (<0.001) |
| **Marine Biology & Hydrobiology ( 9.7%)** | **Other fields** | Raw citations | 7932 (<0.001) | 882 ( 0.410) | -4.67 ( 0.889) |
|  |  | Composite | 3.67 (<0.001) | .0126 ( 0.776) | .00217 ( 0.117) |
| **Networking & Telecommunications ( 9.5%)** | **Other fields** | Raw citations | 5869 (<0.001) | -49 ( 0.935) | -7.6 ( 0.548) |
|  |  | Composite | 3.18 (<0.001) | .0276 ( 0.383) | .00459 (<0.001) |
| **Computer Hardware & Architecture ( 9.3%)** | **Other fields** | Raw citations | 5123 (<0.001) | 308 ( 0.749) | 11.5 ( 0.674) |
|  |  | Composite | 3.17 (<0.001) | -.0273 ( 0.685) | .00665 ( 0.001) |
| **Electrical & Electronic Engineering ( 9.2%)** | **Other fields** | Raw citations | 3198 (<0.001) | 1177 ( 0.158) | 22.7 ( 0.192) |
|  |  | Composite | 3.02 (<0.001) | .0495 ( 0.412) | .00391 ( 0.002) |
| **Geological & Geomatics Engineering ( 9%)** | **Other fields** | Raw citations | 5987 (<0.001) | 1388 ( 0.166) | 5.72 ( 0.833) |
|  |  | Composite | 3.3 (<0.001) | .12 ( 0.029) | .00521 ( 0.001) |
| **Information Systems ( 8.6%)** | **Other fields** | Raw citations | 5740 ( 0.003) | 632 ( 0.731) | 64.4 ( 0.244) |
|  |  | Composite | 3.49 (<0.001) | -.0374 ( 0.585) | .00482 ( 0.020) |
| **Law ( 8.5%)** | **Other fields** | Raw citations | 728 ( 0.455) | -198 ( 0.804) | 33.4 ( 0.310) |
|  |  | Composite | 3.11 (<0.001) | .0375 ( 0.633) | .00577 ( 0.077) |
| **Mathematical Physics ( 8.3%)** | **Other fields** | Raw citations | 4734 ( 0.442) | -2710 ( 0.608) | 8.12 ( 0.939) |
|  |  | Composite | 3.98 (<0.001) | -.388 ( 0.163) | -.00584 ( 0.289) |
| **Strategic, Defence & Security Studies ( 8.2%)** | **Other fields** | Raw citations | 1156 ( 0.209) | 1403 ( 0.181) | 53.4 ( 0.030) |
|  |  | Composite | 3.13 (<0.001) | .189 ( 0.021) | .00552 ( 0.004) |
| **Agricultural Economics & Policy ( 8.2%)** | **Other fields** | Raw citations | 6112 (<0.001) | 2081 ( 0.156) | -26.5 ( 0.494) |
|  |  | Composite | 3.68 (<0.001) | -.0732 ( 0.474) | -.00042 ( 0.877) |
| **General Physics ( 8.2%)** | **Other fields** | Raw citations | 9805 (<0.001) | 2778 ( 0.071) | -31.9 ( 0.247) |
|  |  | Composite | 3.51 (<0.001) | .0254 ( 0.653) | .00247 ( 0.015) |
| **Languages & Linguistics ( 8.2%)** | **Other fields** | Raw citations | 1228 ( 0.026) | 1095 ( 0.050) | 30.6 ( 0.034) |
|  |  | Composite | 3.34 (<0.001) | .165 ( 0.036) | .00355 ( 0.079) |
| **Mining & Metallurgy ( 8.1%)** | **Other fields** | Raw citations | -646 ( 0.497) | 1779 ( 0.087) | 60 ( 0.010) |
|  |  | Composite | 2.34 (<0.001) | .151 ( 0.384) | .014 ( 0.001) |
| **Psychoanalysis ( 8%)** | **Highly related fields** | Raw citations | 1159 ( 0.431) | -16.9 ( 0.990) | 13.1 ( 0.590) |
|  |  | Composite | 3.44 (<0.001) | -.0642 ( 0.641) | -.000979 ( 0.685) |
| **Political Science & Public Administration ( 7.9%)** | **Other fields** | Raw citations | 3338 ( 0.001) | 2392 ( 0.043) | 34.6 ( 0.172) |
|  |  | Composite | 3.62 (<0.001) | .0806 ( 0.174) | .00205 ( 0.108) |
| **Building & Construction ( 7.8%)** | **Other fields** | Raw citations | 2249 ( 0.019) | 925 ( 0.417) | 52.5 ( 0.058) |
|  |  | Composite | 3.12 (<0.001) | .0654 ( 0.496) | .00747 ( 0.002) |
| **Oceanography ( 7.6%)** | **Other fields** | Raw citations | 5640 (<0.001) | 1001 ( 0.391) | 2.28 ( 0.938) |
|  |  | Composite | 3.39 (<0.001) | .0701 ( 0.260) | .00529 ( 0.001) |
| **Literary Studies ( 7.5%)** | **Other fields** | Raw citations | 271 ( 0.242) | -82.3 ( 0.768) | 10.4 ( 0.160) |
|  |  | Composite | 2.63 (<0.001) | .0347 ( 0.669) | .00521 ( 0.016) |
| **Cultural Studies ( 7.4%)** | **Other fields** | Raw citations | 1086 ( 0.052) | -164 ( 0.778) | 2.14 ( 0.863) |
|  |  | Composite | 3.25 (<0.001) | -.143 ( 0.228) | -.0012 ( 0.632) |
| **Materials ( 7.3%)** | **Other fields** | Raw citations | 6710 (<0.001) | 2386 ( 0.003) | -25.5 ( 0.059) |
|  |  | Composite | 3.27 (<0.001) | .13 ( 0.001) | .00237 ( 0.001) |
| **Applied Physics ( 7.2%)** | **Other fields** | Raw citations | 10274 (<0.001) | 2334 ( 0.030) | -13.5 ( 0.460) |
|  |  | Composite | 3.52 (<0.001) | .095 ( 0.001) | .00198 (<0.001) |
| **Econometrics ( 7.1%)** | **Other fields** | Raw citations | 3666 ( 0.469) | 6447 ( 0.215) | 169 ( 0.199) |
|  |  | Composite | 3.57 (<0.001) | .0774 ( 0.692) | .0101 ( 0.045) |
| **Mechanical Engineering & Transports ( 7.1%)** | **Other fields** | Raw citations | 4650 (<0.001) | 2431 ( 0.008) | -3.61 ( 0.839) |
|  |  | Composite | 3.29 (<0.001) | .0471 ( 0.382) | .00319 ( 0.003) |
| **Operations Research ( 6.8%)** | **Other fields** | Raw citations | 974 ( 0.479) | -1618 ( 0.231) | 123 (<0.001) |
|  |  | Composite | 3.45 (<0.001) | -.0941 ( 0.205) | .00547 ( 0.003) |
| **Finance ( 6.8%)** | **Other fields** | Raw citations | 3630 ( 0.041) | -1836 ( 0.343) | 87.8 ( 0.057) |
|  |  | Composite | 3.48 (<0.001) | -.09 ( 0.340) | .00547 ( 0.015) |
| **Business & Management ( 6.5%)** | **Other fields** | Raw citations | 5584 (<0.001) | -398 ( 0.737) | 67.8 ( 0.011) |
|  |  | Composite | 3.63 (<0.001) | .0236 ( 0.611) | .00359 ( 0.001) |
| **Meteorology & Atmospheric Sciences ( 6.4%)** | **Other fields** | Raw citations | 10819 (<0.001) | -539 ( 0.652) | 17.7 ( 0.528) |
|  |  | Composite | 3.68 (<0.001) | -.00165 ( 0.961) | .00293 (<0.001) |
| **General Mathematics ( 6.4%)** | **Other fields** | Raw citations | 2958 (<0.001) | 1782 ( 0.001) | -.659 ( 0.936) |
|  |  | Composite | 3.47 (<0.001) | .0336 ( 0.448) | .000721 ( 0.271) |
| **Computation Theory & Mathematics ( 6.2%)** | **Other fields** | Raw citations | 5038 (<0.001) | 805 ( 0.595) | 34.3 ( 0.285) |
|  |  | Composite | 3.53 (<0.001) | .035 ( 0.675) | .00466 ( 0.009) |
| **Fisheries ( 6.2%)** | **Other fields** | Raw citations | 4249 (<0.001) | 542 ( 0.544) | 5.02 ( 0.824) |
|  |  | Composite | 3.42 (<0.001) | .0137 ( 0.845) | .00128 ( 0.471) |
| **Philosophy ( 6.2%)** | **Other fields** | Raw citations | 1544 (<0.001) | 1032 ( 0.007) | 3.66 ( 0.588) |
|  |  | Composite | 3.4 (<0.001) | .141 ( 0.065) | .000488 ( 0.718) |
| **Archaeology ( 6.1%)** | **Other fields** | Raw citations | 2115 ( 0.003) | -748 ( 0.429) | 15.8 ( 0.366) |
|  |  | Composite | 3.23 (<0.001) | -.154 ( 0.102) | .00427 ( 0.015) |
| **Paleontology ( 6%)** | **Other fields** | Raw citations | 7104 (<0.001) | -108 ( 0.957) | 4.64 ( 0.915) |
|  |  | Composite | 3.64 (<0.001) | .0608 ( 0.365) | .00263 ( 0.071) |
| **Energy ( 5.9%)** | **Other fields** | Raw citations | 4517 (<0.001) | 1099 ( 0.079) | 7.36 ( 0.497) |
|  |  | Composite | 3.2 (<0.001) | .031 ( 0.436) | .00373 (<0.001) |
| **Agronomy & Agriculture ( 5.9%)** | **Other fields** | Raw citations | 5482 (<0.001) | 825 ( 0.405) | -1.73 ( 0.927) |
|  |  | Composite | 3.36 (<0.001) | .0335 ( 0.549) | .0027 ( 0.012) |
| **Aerospace & Aeronautics ( 5.8%)** | **Other fields** | Raw citations | 2312 (<0.001) | 249 ( 0.513) | 8.15 ( 0.289) |
|  |  | Composite | 3.08 (<0.001) | -.0169 ( 0.728) | .0029 ( 0.003) |
| **Astronomy & Astrophysics ( 5.6%)** | **Other fields** | Raw citations | 18634 (<0.001) | -353 ( 0.876) | -85.6 ( 0.064) |
|  |  | Composite | 3.75 (<0.001) | .0537 ( 0.140) | .00118 ( 0.113) |
| **Classics ( 5.6%)** | **Other fields** | Raw citations | 1694 ( 0.058) | -197 ( 0.851) | -23.7 ( 0.301) |
|  |  | Composite | 2.94 (<0.001) | -.143 ( 0.505) | .00159 ( 0.729) |
| **Geology ( 5.4%)** | **Other fields** | Raw citations | 6016 (<0.001) | -128 ( 0.910) | -40 ( 0.061) |
|  |  | Composite | 3.64 (<0.001) | .0227 ( 0.830) | -.00215 ( 0.276) |
| **Forestry ( 5.3%)** | **Other fields** | Raw citations | 2705 (<0.001) | 389 ( 0.608) | 18.2 ( 0.333) |
|  |  | Composite | 3.14 (<0.001) | .0618 ( 0.442) | .00393 ( 0.049) |
| **Geochemistry & Geophysics ( 4.9%)** | **Other fields** | Raw citations | 7287 (<0.001) | 72.3 ( 0.931) | 15.7 ( 0.336) |
|  |  | Composite | 3.66 (<0.001) | .00353 ( 0.929) | .00313 (<0.001) |
| **Nuclear & Particle Physics ( 4.7%)** | **Other fields** | Raw citations | 18813 (<0.001) | -5775 ( 0.033) | -102 ( 0.002) |
|  |  | Composite | 3.64 (<0.001) | -.0187 ( 0.689) | .0012 ( 0.037) |
| **History ( 4.5%)** | **Other fields** | Raw citations | 833 ( 0.001) | -82.9 ( 0.826) | -.847 ( 0.877) |
|  |  | Composite | 2.91 (<0.001) | .00468 ( 0.961) | .000996 ( 0.470) |
| **International Relations ( 4.4%)** | **Other fields** | Raw citations | 2196 ( 0.017) | 340 ( 0.801) | 27.3 ( 0.234) |
|  |  | Composite | 3.42 (<0.001) | .0887 ( 0.515) | .00402 ( 0.084) |
| **Physical Chemistry ( 4.4%)** | **Other fields** | Raw citations | 11444 (<0.001) | 16389 (<0.001) | -51.5 ( 0.323) |
|  |  | Composite | 3.49 (<0.001) | .277 ( 0.106) | .00337 ( 0.137) |
| **Science Studies ( 3.8%)** | **Other fields** | Raw citations | 4042 ( 0.329) | 270 ( 0.950) | 27 ( 0.772) |
|  |  | Composite | 3.7 (<0.001) | -.141 ( 0.648) | .00151 ( 0.821) |
| **Civil Engineering ( 3.6%)** | **Other fields** | Raw citations | 1981 ( 0.011) | -789 ( 0.511) | 34.5 ( 0.072) |
|  |  | Composite | 3.1 (<0.001) | -.104 ( 0.330) | .00447 ( 0.009) |
| **Legal & Forensic Medicine ( 3%)** | **Highly related fields** | Raw citations | 1687 ( 0.120) | -1261 ( 0.493) | 13.4 ( 0.628) |
|  |  | Composite | 2.92 (<0.001) | -.228 ( 0.240) | .00299 ( 0.307) |
| **Music ( 2.4%)** | **Other fields** | Raw citations | 658 ( 0.029) | 41.2 ( 0.939) | -3.65 ( 0.654) |
|  |  | Composite | 2.76 (<0.001) | .227 ( 0.150) | .00133 ( 0.569) |
| **Accounting ( 2.4%)** | **Other fields** | Raw citations | 5000 ( 0.014) | 4808 ( 0.124) | 24.2 ( 0.655) |
|  |  | Composite | 3.44 (<0.001) | .286 ( 0.165) | .00512 ( 0.156) |
| **Horticulture ( 1.3%)** | **Other fields** | Raw citations | 1138 ( 0.267) | -827 ( 0.650) | 38.6 ( 0.170) |
|  |  | Composite | 2.98 (<0.001) | -.0732 ( 0.764) | .00666 ( 0.078) |
| **Industrial Relations ( 0%)** | **Other fields** | Raw citations |  |  |  |
|  |  | Composite |  |  |  |
| **Folklore ( 0%)** | **Other fields** | Raw citations |  |  |  |
|  |  | Composite |  |  |  |
| **Automobile Design & Engineering ( 0%)** | **Other fields** | Raw citations |  |  |  |
|  |  | Composite |  |  |  |
| **Art Practice, History & Theory ( 0%)** | **Other fields** | Raw citations |  |  |  |
|  |  | Composite |  |  |  |

**Supplementary Table 3.2 : Career-long impact, Funding time recent funding Linear Regressions for each subfield (ordered by percentage funded)**

| **Top-cited US-based researchers: Subfield (perc. funded)** | **Classification** | **Dependent Variable** | **Constant (p-val)** | **Funded (p-val)** | **Years since first pub (p-val)** |
| --- | --- | --- | --- | --- | --- |
| **Developmental Biology ( 42%)** | **Highly related fields** | Raw citations | 18380 (<0.001) | 2703 ( 0.037) | 40.4 ( 0.505) |
|  |  | Composite | 3.58 (<0.001) | .0292 ( 0.069) | .00773 (<0.001) |
| **Bioinformatics ( 41%)** | **Highly related fields** | Raw citations | 13708 ( 0.037) | 8132 ( 0.030) | 66.5 ( 0.707) |
|  |  | Composite | 3.38 (<0.001) | .119 ( 0.012) | .00696 ( 0.002) |
| **Geriatrics ( 40%)** | **Highly related fields** | Raw citations | 5149 ( 0.386) | 4288 ( 0.119) | 161 ( 0.271) |
|  |  | Composite | 3.26 (<0.001) | .146 ( 0.043) | .0108 ( 0.006) |
| **Substance Abuse ( 36%)** | **Highly related fields** | Raw citations | 2670 ( 0.129) | 2710 ( 0.003) | 159 (<0.001) |
|  |  | Composite | 3.59 (<0.001) | .0219 ( 0.513) | .00461 ( 0.002) |
| **Biomedical Engineering ( 34%)** | **Highly related fields** | Raw citations | 4129 ( 0.017) | 3388 ( 0.002) | 82.3 ( 0.046) |
|  |  | Composite | 3.18 (<0.001) | .0682 ( 0.034) | .00698 (<0.001) |
| **Medical Informatics ( 34%)** | **Highly related fields** | Raw citations | 4045 ( 0.082) | 2430 ( 0.107) | 25.5 ( 0.666) |
|  |  | Composite | 3.2 (<0.001) | .133 ( 0.034) | .00425 ( 0.083) |
| **Immunology ( 34%)** | **Highly related fields** | Raw citations | 16270 (<0.001) | 2368 ( 0.015) | 52.6 ( 0.201) |
|  |  | Composite | 3.66 (<0.001) | .064 (<0.001) | .00456 (<0.001) |
| **Epidemiology ( 34%)** | **Highly related fields** | Raw citations | 2782 ( 0.801) | 3730 ( 0.521) | 458 ( 0.075) |
|  |  | Composite | 3.58 (<0.001) | -.00789 ( 0.892) | .00811 ( 0.002) |
| **Virology ( 34%)** | **Highly related fields** | Raw citations | 8988 (<0.001) | 1275 ( 0.126) | 87 ( 0.017) |
|  |  | Composite | 3.45 (<0.001) | .0338 ( 0.132) | .00558 (<0.001) |
| **Neurology & Neurosurgery ( 32%)** | **Highly related fields** | Raw citations | 13756 (<0.001) | 1062 ( 0.067) | 32.3 ( 0.174) |
|  |  | Composite | 3.66 (<0.001) | .0268 ( 0.026) | .00446 (<0.001) |
| **Public Health ( 31%)** | **Highly related fields** | Raw citations | 4510 ( 0.007) | 1653 ( 0.075) | 157 (<0.001) |
|  |  | Composite | 3.46 (<0.001) | .0781 ( 0.001) | .00649 (<0.001) |
| **Gerontology ( 30%)** | **Highly related fields** | Raw citations | 6012 ( 0.469) | 11949 ( 0.004) | 56.4 ( 0.771) |
|  |  | Composite | 3.49 (<0.001) | .114 ( 0.068) | .00496 ( 0.093) |
| **Oncology & Carcinogenesis ( 30%)** | **Highly related fields** | Raw citations | 16904 (<0.001) | 2264 ( 0.006) | 31.5 ( 0.344) |
|  |  | Composite | 3.49 (<0.001) | .0333 ( 0.014) | .00627 (<0.001) |
| **Genetics & Heredity ( 29%)** | **Highly related fields** | Raw citations | 18062 (<0.001) | 4592 ( 0.016) | -51.7 ( 0.533) |
|  |  | Composite | 3.52 (<0.001) | .064 ( 0.055) | .00476 ( 0.001) |
| **Demography ( 26%)** | **Highly related fields** | Raw citations | -2967 ( 0.289) | 3338 ( 0.042) | 156 ( 0.009) |
|  |  | Composite | 3.25 (<0.001) | .104 ( 0.295) | .00844 ( 0.020) |
| **Health Policy & Services ( 26%)** | **Highly related fields** | Raw citations | 925 ( 0.720) | 2001 ( 0.193) | 228 ( 0.001) |
|  |  | Composite | 3.26 (<0.001) | .104 ( 0.019) | .0108 (<0.001) |
| **Developmental & Child Psychology ( 25%)** | **Highly related fields** | Raw citations | 3284 ( 0.051) | 4979 (<0.001) | 141 (<0.001) |
|  |  | Composite | 3.53 (<0.001) | .152 (<0.001) | .00658 (<0.001) |
| **Applied Ethics ( 25%)** | **Highly related fields** | Raw citations | 4954 ( 0.002) | 2486 ( 0.008) | -16.5 ( 0.660) |
|  |  | Composite | 3.45 (<0.001) | .0944 ( 0.166) | .00399 ( 0.155) |
| **Gastroenterology & Hepatology ( 25%)** | **Highly related fields** | Raw citations | 11100 (<0.001) | 2220 ( 0.046) | 44.6 ( 0.286) |
|  |  | Composite | 3.6 (<0.001) | .0773 ( 0.008) | .00397 (<0.001) |
| **Endocrinology & Metabolism ( 25%)** | **Highly related fields** | Raw citations | 14061 (<0.001) | 625 ( 0.584) | 39.9 ( 0.355) |
|  |  | Composite | 3.7 (<0.001) | .0014 ( 0.954) | .00364 (<0.001) |
| **Biophysics ( 25%)** | **Highly related fields** | Raw citations | 5070 ( 0.143) | 1652 ( 0.427) | 83.4 ( 0.255) |
|  |  | Composite | 3.3 (<0.001) | .0786 ( 0.197) | .00669 ( 0.002) |
| **Psychiatry ( 25%)** | **Highly related fields** | Raw citations | 10288 (<0.001) | 3928 ( 0.003) | 133 ( 0.010) |
|  |  | Composite | 3.67 (<0.001) | .0758 ( 0.004) | .00497 (<0.001) |
| **Arthritis & Rheumatology ( 24%)** | **Highly related fields** | Raw citations | 11215 ( 0.004) | 700 ( 0.756) | 146 ( 0.105) |
|  |  | Composite | 3.64 (<0.001) | .011 ( 0.831) | .00503 ( 0.015) |
| **Emergency & Critical Care Medicine ( 24%)** | **Highly related fields** | Raw citations | 3871 ( 0.129) | 7100 (<0.001) | 138 ( 0.035) |
|  |  | Composite | 3.27 (<0.001) | .13 ( 0.002) | .00711 (<0.001) |
| **Microbiology ( 23%)** | **Highly related fields** | Raw citations | 12835 (<0.001) | 1461 ( 0.042) | -34.2 ( 0.191) |
|  |  | Composite | 3.53 (<0.001) | .0507 ( 0.008) | .00472 (<0.001) |
| **Allergy ( 23%)** | **Highly related fields** | Raw citations | 7706 ( 0.035) | 3713 ( 0.092) | 90.3 ( 0.262) |
|  |  | Composite | 3.6 (<0.001) | .0464 ( 0.486) | .00412 ( 0.093) |
| **Respiratory System ( 23%)** | **Highly related fields** | Raw citations | 10474 (<0.001) | 1032 ( 0.369) | 83.8 ( 0.070) |
|  |  | Composite | 3.55 (<0.001) | .0191 ( 0.504) | .00472 (<0.001) |
| **Clinical Psychology ( 23%)** | **Highly related fields** | Raw citations | 6159 ( 0.010) | 4980 ( 0.003) | 100 ( 0.059) |
|  |  | Composite | 3.69 (<0.001) | .139 ( 0.009) | .00347 ( 0.039) |
| **Cardiovascular System & Hematology ( 23%)** | **Highly related fields** | Raw citations | 17864 (<0.001) | 4871 (<0.001) | 41.2 ( 0.342) |
|  |  | Composite | 3.59 (<0.001) | .0717 (<0.001) | .00509 (<0.001) |
| **Biochemistry & Molecular Biology ( 22%)** | **Highly related fields** | Raw citations | 13024 (<0.001) | 757 ( 0.281) | 6.57 ( 0.787) |
|  |  | Composite | 3.69 (<0.001) | .0155 ( 0.342) | .00312 (<0.001) |
| **Physiology ( 22%)** | **Highly related fields** | Raw citations | 5511 (<0.001) | 1054 ( 0.186) | 46 ( 0.065) |
|  |  | Composite | 3.7 (<0.001) | .0309 ( 0.413) | .00158 ( 0.180) |
| **Ophthalmology & Optometry ( 21%)** | **Highly related fields** | Raw citations | 9174 (<0.001) | 3663 (<0.001) | -32.9 ( 0.206) |
|  |  | Composite | 3.46 (<0.001) | .127 (<0.001) | .00253 ( 0.006) |
| **Analytical Chemistry ( 21%)** | **Other fields** | Raw citations | 5155 (<0.001) | 3633 (<0.001) | 28.4 ( 0.260) |
|  |  | Composite | 3.24 (<0.001) | .14 (<0.001) | .00548 (<0.001) |
| **Urology & Nephrology ( 20%)** | **Highly related fields** | Raw citations | 14037 (<0.001) | 1128 ( 0.290) | -45.5 ( 0.237) |
|  |  | Composite | 3.57 (<0.001) | .0734 ( 0.007) | .00328 ( 0.001) |
| **Rehabilitation ( 20%)** | **Highly related fields** | Raw citations | 4311 (<0.001) | 915 ( 0.183) | 25.6 ( 0.377) |
|  |  | Composite | 3.31 (<0.001) | .038 ( 0.369) | .00441 ( 0.014) |
| **Microscopy ( 20%)** | **Highly related fields** | Raw citations | 4673 ( 0.196) | 5392 ( 0.016) | 35.6 ( 0.662) |
|  |  | Composite | 3.23 (<0.001) | .238 ( 0.094) | .00552 ( 0.304) |
| **Tropical Medicine ( 20%)** | **Highly related fields** | Raw citations | 7901 (<0.001) | 1618 ( 0.144) | -33 ( 0.320) |
|  |  | Composite | 3.37 (<0.001) | .0669 ( 0.175) | .00164 ( 0.269) |
| **Statistics & Probability ( 20%)** | **Other fields** | Raw citations | 10637 ( 0.001) | 5233 ( 0.024) | 8.11 ( 0.902) |
|  |  | Composite | 3.69 (<0.001) | .11 ( 0.027) | .00189 ( 0.181) |
| **Family Studies ( 20%)** | **Other fields** | Raw citations | -123 ( 0.927) | 2036 ( 0.030) | 97.8 ( 0.005) |
|  |  | Composite | 3.15 (<0.001) | .13 ( 0.065) | .0101 (<0.001) |
| **Toxicology ( 20%)** | **Highly related fields** | Raw citations | 8420 (<0.001) | 3264 (<0.001) | -33.9 ( 0.285) |
|  |  | Composite | 3.34 (<0.001) | .113 ( 0.001) | .0041 ( 0.001) |
| **Nutrition & Dietetics ( 19%)** | **Highly related fields** | Raw citations | 7523 (<0.001) | 2664 ( 0.058) | 52 ( 0.262) |
|  |  | Composite | 3.5 (<0.001) | .072 ( 0.088) | .00566 (<0.001) |
| **Medicinal & Biomolecular Chemistry ( 19%)** | **Highly related fields** | Raw citations | 1298 ( 0.149) | 2568 (<0.001) | 115 (<0.001) |
|  |  | Composite | 2.91 (<0.001) | .132 (<0.001) | .00907 (<0.001) |
| **Nuclear Medicine & Medical Imaging ( 19%)** | **Highly related fields** | Raw citations | 6170 (<0.001) | 3041 (<0.001) | 44.7 ( 0.050) |
|  |  | Composite | 3.38 (<0.001) | .106 (<0.001) | .00295 (<0.001) |
| **Nanoscience & Nanotechnology ( 19%)** | **Other fields** | Raw citations | 6951 ( 0.019) | 3115 ( 0.258) | 504 (<0.001) |
|  |  | Composite | 3.29 (<0.001) | .044 ( 0.320) | .0144 (<0.001) |
| **Pediatrics ( 19%)** | **Highly related fields** | Raw citations | 5342 (<0.001) | 2847 (<0.001) | 32 ( 0.086) |
|  |  | Composite | 3.27 (<0.001) | .117 (<0.001) | .00485 (<0.001) |
| **Pharmacology & Pharmacy ( 18%)** | **Highly related fields** | Raw citations | 4335 (<0.001) | 3030 (<0.001) | 40.1 ( 0.031) |
|  |  | Composite | 3.28 (<0.001) | .117 (<0.001) | .00393 (<0.001) |
| **Optics ( 18%)** | **Other fields** | Raw citations | 7761 (<0.001) | 3540 ( 0.004) | 4.19 ( 0.913) |
|  |  | Composite | 3.25 (<0.001) | .0607 ( 0.166) | .00502 (<0.001) |
| **Anesthesiology ( 18%)** | **Highly related fields** | Raw citations | 5756 (<0.001) | 2786 (<0.001) | 12.5 ( 0.638) |
|  |  | Composite | 3.39 (<0.001) | .0998 ( 0.010) | .00254 ( 0.063) |
| **Obstetrics & Reproductive Medicine ( 18%)** | **Highly related fields** | Raw citations | 9194 (<0.001) | 1693 ( 0.015) | -33.9 ( 0.143) |
|  |  | Composite | 3.49 (<0.001) | .0569 ( 0.044) | .00223 ( 0.018) |
| **Experimental Psychology ( 18%)** | **Highly related fields** | Raw citations | 6479 (<0.001) | 2860 ( 0.001) | 47.3 ( 0.045) |
|  |  | Composite | 3.68 (<0.001) | .0928 ( 0.004) | .00349 (<0.001) |
| **Speech-Language Pathology & Audiology ( 17%)** | **Highly related fields** | Raw citations | 6410 (<0.001) | 662 ( 0.521) | -45.9 ( 0.286) |
|  |  | Composite | 3.36 (<0.001) | .00525 ( 0.934) | .00439 ( 0.096) |
| **Biotechnology ( 17%)** | **Highly related fields** | Raw citations | 3864 ( 0.021) | 1784 ( 0.168) | 98.4 ( 0.027) |
|  |  | Composite | 3.1 (<0.001) | .119 ( 0.037) | .00877 (<0.001) |
| **Organic Chemistry ( 16%)** | **Other fields** | Raw citations | 8186 (<0.001) | 4348 (<0.001) | 23.7 ( 0.400) |
|  |  | Composite | 3.51 (<0.001) | .126 ( 0.001) | .00406 (<0.001) |
| **Nursing ( 16%)** | **Highly related fields** | Raw citations | 1148 ( 0.001) | 2076 (<0.001) | 31.1 ( 0.001) |
|  |  | Composite | 3 (<0.001) | .126 (<0.001) | .00359 (<0.001) |
| **Environmental & Occupational Health ( 15%)** | **Highly related fields** | Raw citations | 4784 ( 0.026) | -324 ( 0.786) | 11.4 ( 0.821) |
|  |  | Composite | 3.21 (<0.001) | .0191 ( 0.744) | .0039 ( 0.119) |
| **Behavioral Science & Comparative Psychology ( 13%)** | **Highly related fields** | Raw citations | 3705 ( 0.004) | 1873 ( 0.035) | 45.1 ( 0.099) |
|  |  | Composite | 3.56 (<0.001) | .0386 ( 0.450) | .00472 ( 0.003) |
| **Mycology & Parasitology ( 13%)** | **Highly related fields** | Raw citations | 8399 (<0.001) | -564 ( 0.724) | -44 ( 0.303) |
|  |  | Composite | 3.41 (<0.001) | -.000403 ( 0.995) | .00227 ( 0.212) |
| **General & Internal Medicine ( 13%)** | **Highly related fields** | Raw citations | 12625 (<0.001) | 3604 ( 0.003) | -104 (<0.001) |
|  |  | Composite | 3.29 (<0.001) | .136 (<0.001) | .000181 ( 0.753) |
| **Otorhinolaryngology ( 13%)** | **Highly related fields** | Raw citations | 5666 (<0.001) | 963 ( 0.055) | -27.1 ( 0.054) |
|  |  | Composite | 3.28 (<0.001) | .0898 ( 0.005) | .00284 ( 0.002) |
| **Sociology ( 13%)** | **Other fields** | Raw citations | 2549 ( 0.007) | 1086 ( 0.221) | 59.6 ( 0.008) |
|  |  | Composite | 3.58 (<0.001) | -.0241 ( 0.641) | .00402 ( 0.002) |
| **Surgery ( 13%)** | **Highly related fields** | Raw citations | 8909 (<0.001) | 3918 (<0.001) | -16.3 ( 0.340) |
|  |  | Composite | 3.43 (<0.001) | .0975 (<0.001) | .00122 ( 0.052) |
| **Development Studies ( 13%)** | **Other fields** | Raw citations | 5361 ( 0.015) | -764 ( 0.540) | -39.5 ( 0.458) |
|  |  | Composite | 3.88 (<0.001) | -.167 ( 0.253) | -.00588 ( 0.342) |
| **Drama & Theater ( 11%)** | **Other fields** | Raw citations | -188 ( 0.513) | 379 ( 0.116) | 21.8 ( 0.051) |
|  |  | Composite | 2.24 (<0.001) | .246 ( 0.204) | .0169 ( 0.065) |
| **General Chemistry ( 11%)** | **Other fields** | Raw citations | 13581 (<0.001) | 3801 ( 0.148) | -74.8 ( 0.038) |
|  |  | Composite | 3.34 (<0.001) | .285 (<0.001) | .00272 ( 0.005) |
| **Dermatology & Venereal Diseases ( 11%)** | **Highly related fields** | Raw citations | 8923 (<0.001) | 5520 (<0.001) | -20.9 ( 0.494) |
|  |  | Composite | 3.61 (<0.001) | .117 ( 0.018) | .000838 ( 0.471) |
| **Acoustics ( 11%)** | **Other fields** | Raw citations | 5868 (<0.001) | 299 ( 0.657) | -37.6 ( 0.011) |
|  |  | Composite | 3.54 (<0.001) | -.0497 ( 0.315) | -.00109 ( 0.315) |
| **Criminology ( 10%)** | **Other fields** | Raw citations | 2832 ( 0.012) | 1150 ( 0.263) | 55.8 ( 0.051) |
|  |  | Composite | 3.49 (<0.001) | .0599 ( 0.338) | .00439 ( 0.012) |
| **Evolutionary Biology ( 10%)** | **Other fields** | Raw citations | 4856 ( 0.041) | 3072 ( 0.196) | 139 ( 0.019) |
|  |  | Composite | 3.62 (<0.001) | .0489 ( 0.307) | .00645 (<0.001) |
| **Orthopedics ( 10%)** | **Highly related fields** | Raw citations | 7241 (<0.001) | 3837 (<0.001) | 1.31 ( 0.946) |
|  |  | Composite | 3.38 (<0.001) | .171 (<0.001) | .00443 (<0.001) |
| **Environmental Sciences ( 9.7%)** | **Other fields** | Raw citations | 5136 (<0.001) | 4222 ( 0.001) | 34.9 ( 0.294) |
|  |  | Composite | 3.26 (<0.001) | .136 ( 0.008) | .00541 (<0.001) |
| **Distributed Computing ( 9.4%)** | **Other fields** | Raw citations | 1607 ( 0.331) | 5988 (<0.001) | 83.7 ( 0.087) |
|  |  | Composite | 3.13 (<0.001) | .106 ( 0.188) | .00286 ( 0.309) |
| **Social Work ( 9.1%)** | **Other fields** | Raw citations | 945 ( 0.120) | 1935 ( 0.001) | 28.5 ( 0.097) |
|  |  | Composite | 3.2 (<0.001) | .163 ( 0.027) | .0021 ( 0.355) |
| **Social Sciences Methods ( 9.1%)** | **Other fields** | Raw citations | 5776 ( 0.216) | 8448 ( 0.141) | 69.6 ( 0.451) |
|  |  | Composite | 3.57 (<0.001) | .38 ( 0.002) | .00476 ( 0.017) |
| **Artificial Intelligence & Image Processing ( 8.9%)** | **Other fields** | Raw citations | 5549 (<0.001) | 1884 ( 0.055) | 66.1 ( 0.010) |
|  |  | Composite | 3.18 (<0.001) | .0864 ( 0.011) | .00835 (<0.001) |
| **Complementary & Alternative Medicine ( 8.6%)** | **Highly related fields** | Raw citations | 2381 ( 0.006) | 2062 ( 0.029) | -7.06 ( 0.782) |
|  |  | Composite | 3.01 (<0.001) | .205 ( 0.134) | -.00102 ( 0.785) |
| **Dentistry ( 8%)** | **Highly related fields** | Raw citations | 5242 (<0.001) | 1039 ( 0.167) | -8.8 ( 0.605) |
|  |  | Composite | 3.34 (<0.001) | .0697 ( 0.124) | .00266 ( 0.010) |
| **Veterinary Sciences ( 7.9%)** | **Highly related fields** | Raw citations | 3394 (<0.001) | 876 ( 0.062) | 13.5 ( 0.291) |
|  |  | Composite | 3.25 (<0.001) | .0165 ( 0.618) | .00259 ( 0.004) |
| **Pathology ( 7.8%)** | **Highly related fields** | Raw citations | 14924 (<0.001) | 573 ( 0.804) | -68 ( 0.207) |
|  |  | Composite | 3.56 (<0.001) | .065 ( 0.346) | .00296 ( 0.066) |
| **Plant Biology & Botany ( 7.8%)** | **Other fields** | Raw citations | 7649 (<0.001) | 6088 (<0.001) | -4.17 ( 0.843) |
|  |  | Composite | 3.44 (<0.001) | .203 (<0.001) | .00333 (<0.001) |
| **Gender Studies ( 7.7%)** | **Highly related fields** | Raw citations | 3443 ( 0.159) | 542 ( 0.741) | -16.7 ( 0.778) |
|  |  | Composite | 3.41 (<0.001) | .106 ( 0.566) | .00303 ( 0.649) |
| **Social Psychology ( 7.7%)** | **Highly related fields** | Raw citations | 7636 (<0.001) | 2379 ( 0.206) | 74.5 ( 0.031) |
|  |  | Composite | 3.71 (<0.001) | .101 ( 0.080) | .004 (<0.001) |
| **Geography ( 7.6%)** | **Other fields** | Raw citations | 1761 ( 0.127) | 2708 ( 0.027) | 62.5 ( 0.031) |
|  |  | Composite | 3.5 (<0.001) | .167 ( 0.071) | .00574 ( 0.010) |
| **Sport Sciences ( 7.5%)** | **Highly related fields** | Raw citations | 3908 ( 0.073) | 759 ( 0.753) | 103 ( 0.072) |
|  |  | Composite | 3.45 (<0.001) | -.032 ( 0.692) | .00384 ( 0.045) |
| **Industrial Engineering & Automation ( 7.3%)** | **Other fields** | Raw citations | 5873 (<0.001) | 1191 ( 0.197) | 4.07 ( 0.846) |
|  |  | Composite | 3.39 (<0.001) | .0566 ( 0.291) | .00338 ( 0.006) |
| **Chemical Physics ( 7.3%)** | **Other fields** | Raw citations | 11369 (<0.001) | 6593 ( 0.002) | 46.4 ( 0.186) |
|  |  | Composite | 3.76 (<0.001) | .173 (<0.001) | .00223 ( 0.002) |
| **Urban & Regional Planning ( 6.6%)** | **Other fields** | Raw citations | 3722 ( 0.006) | 1254 ( 0.409) | -4.24 ( 0.897) |
|  |  | Composite | 3.58 (<0.001) | .0714 ( 0.622) | .000918 ( 0.768) |
| **Sport, Leisure & Tourism ( 6.5%)** | **Other fields** | Raw citations | 3171 ( 0.010) | -1116 ( 0.449) | 46.2 ( 0.240) |
|  |  | Composite | 3.44 (<0.001) | -.0313 ( 0.793) | .00611 ( 0.058) |
| **Entomology ( 6.1%)** | **Other fields** | Raw citations | 4771 (<0.001) | 3014 (<0.001) | -12.6 ( 0.394) |
|  |  | Composite | 3.49 (<0.001) | .172 ( 0.004) | .000408 ( 0.717) |
| **Human Factors ( 6%)** | **Highly related fields** | Raw citations | 4439 (<0.001) | 540 ( 0.716) | 62.8 ( 0.070) |
|  |  | Composite | 3.41 (<0.001) | .0738 ( 0.399) | .00702 ( 0.001) |
| **Information & Library Sciences ( 6%)** | **Other fields** | Raw citations | 635 ( 0.285) | 792 ( 0.355) | 44.8 ( 0.008) |
|  |  | Composite | 3.02 (<0.001) | .0433 ( 0.699) | .00684 ( 0.002) |
| **Numerical & Computational Mathematics ( 5.8%)** | **Other fields** | Raw citations | 1557 ( 0.312) | 4150 ( 0.012) | 77.1 ( 0.027) |
|  |  | Composite | 3.57 (<0.001) | .0519 ( 0.598) | .00122 ( 0.557) |
| **Economics ( 5.7%)** | **Other fields** | Raw citations | 6202 (<0.001) | 2505 ( 0.069) | 14.9 ( 0.537) |
|  |  | Composite | 3.61 (<0.001) | .162 ( 0.005) | .00448 (<0.001) |
| **Economic Theory ( 5.6%)** | **Other fields** | Raw citations | 2749 ( 0.150) | -9.76 ( 0.996) | 20 ( 0.616) |
|  |  | Composite | 3.49 (<0.001) | -.145 ( 0.287) | .0029 ( 0.285) |
| **Classics ( 5.6%)** | **Other fields** | Raw citations | 1821 ( 0.045) | -692 ( 0.508) | -26.5 ( 0.248) |
|  |  | Composite | 2.97 (<0.001) | -.138 ( 0.522) | .000834 ( 0.857) |
| **Anatomy & Morphology ( 5.6%)** | **Highly related fields** | Raw citations | 2839 ( 0.003) | -414 ( 0.790) | -13.7 ( 0.524) |
|  |  | Composite | 2.98 (<0.001) | -.262 ( 0.082) | .00394 ( 0.058) |
| **General Clinical Medicine ( 5.5%)** | **Highly related fields** | Raw citations | 4671 ( 0.006) | 4784 ( 0.020) | -12.7 ( 0.725) |
|  |  | Composite | 2.98 (<0.001) | .317 ( 0.023) | .00422 ( 0.084) |
| **Logistics & Transportation ( 5.5%)** | **Other fields** | Raw citations | 3164 (<0.001) | -717 ( 0.465) | 14.1 ( 0.480) |
|  |  | Composite | 3.22 (<0.001) | -.0589 ( 0.546) | .00424 ( 0.033) |
| **Computer Hardware & Architecture ( 5.3%)** | **Other fields** | Raw citations | 5136 (<0.001) | 264 ( 0.833) | 11.5 ( 0.674) |
|  |  | Composite | 3.17 (<0.001) | -.0908 ( 0.297) | .00655 ( 0.001) |
| **Polymers ( 5.3%)** | **Other fields** | Raw citations | 9249 (<0.001) | 3006 ( 0.159) | -15.4 ( 0.632) |
|  |  | Composite | 3.51 (<0.001) | .0498 ( 0.477) | .00171 ( 0.106) |
| **Communication & Media Studies ( 5.2%)** | **Other fields** | Raw citations | 998 ( 0.281) | 336 ( 0.827) | 81.3 ( 0.002) |
|  |  | Composite | 3.45 (<0.001) | .0191 ( 0.820) | .00337 ( 0.015) |
| **Fluids & Plasmas ( 5.1%)** | **Other fields** | Raw citations | 12731 (<0.001) | 4258 ( 0.143) | -68.1 ( 0.197) |
|  |  | Composite | 3.71 (<0.001) | .131 ( 0.053) | .00171 ( 0.165) |
| **Software Engineering ( 4.8%)** | **Other fields** | Raw citations | 7307 (<0.001) | 2275 ( 0.033) | -58.5 ( 0.016) |
|  |  | Composite | 3.51 (<0.001) | .0251 ( 0.744) | .000579 ( 0.740) |
| **Electrical & Electronic Engineering ( 4.8%)** | **Other fields** | Raw citations | 3331 (<0.001) | 634 ( 0.574) | 21.1 ( 0.226) |
|  |  | Composite | 3.03 (<0.001) | .0114 ( 0.889) | .00382 ( 0.003) |
| **Optoelectronics & Photonics ( 4.7%)** | **Other fields** | Raw citations | 3129 (<0.001) | 2733 ( 0.001) | 30.6 ( 0.040) |
|  |  | Composite | 2.99 (<0.001) | .105 ( 0.043) | .00586 (<0.001) |
| **Marketing ( 4.7%)** | **Other fields** | Raw citations | 2400 ( 0.242) | -1934 ( 0.440) | 158 ( 0.008) |
|  |  | Composite | 3.56 (<0.001) | -.0956 ( 0.337) | .00476 ( 0.042) |
| **Languages & Linguistics ( 4.5%)** | **Other fields** | Raw citations | 1366 ( 0.015) | 242 ( 0.744) | 29 ( 0.048) |
|  |  | Composite | 3.36 (<0.001) | .0355 ( 0.734) | .00331 ( 0.108) |
| **Education ( 4.5%)** | **Other fields** | Raw citations | 1702 (<0.001) | 2779 (<0.001) | 44.1 (<0.001) |
|  |  | Composite | 3.24 (<0.001) | .179 (<0.001) | .00502 (<0.001) |
| **Networking & Telecommunications ( 4.5%)** | **Other fields** | Raw citations | 5917 (<0.001) | -593 ( 0.489) | -8.36 ( 0.510) |
|  |  | Composite | 3.18 (<0.001) | -.0431 ( 0.339) | .0045 (<0.001) |
| **Religions & Theology ( 4.4%)** | **Other fields** | Raw citations | 1003 ( 0.019) | -52.6 ( 0.946) | -2.99 ( 0.780) |
|  |  | Composite | 2.93 (<0.001) | .0854 ( 0.587) | .0000947 ( 0.965) |
| **Archaeology ( 4.4%)** | **Other fields** | Raw citations | 2107 ( 0.003) | -679 ( 0.540) | 15.6 ( 0.373) |
|  |  | Composite | 3.23 (<0.001) | -.102 ( 0.356) | .00425 ( 0.016) |
| **History of Social Sciences ( 4.3%)** | **Other fields** | Raw citations | -424 ( 0.872) | -814 ( 0.790) | 71.3 ( 0.219) |
|  |  | Composite | 2.89 (<0.001) | -.0518 ( 0.859) | .0119 ( 0.038) |
| **Applied Mathematics ( 4.3%)** | **Other fields** | Raw citations | 2859 ( 0.345) | 10207 ( 0.017) | 117 ( 0.087) |
|  |  | Composite | 3.49 (<0.001) | .229 ( 0.136) | .00551 ( 0.027) |
| **Mathematical Physics ( 4.2%)** | **Other fields** | Raw citations | 4008 ( 0.501) | -2176 ( 0.761) | 18.9 ( 0.855) |
|  |  | Composite | 3.89 (<0.001) | -.358 ( 0.346) | -.00453 ( 0.409) |
| **Agricultural Economics & Policy ( 4.1%)** | **Other fields** | Raw citations | 6524 (<0.001) | -2108 ( 0.301) | -30.6 ( 0.434) |
|  |  | Composite | 3.68 (<0.001) | -.13 ( 0.357) | -.000426 ( 0.875) |
| **Food Science ( 4.1%)** | **Other fields** | Raw citations | 5243 (<0.001) | 3202 ( 0.119) | 10.4 ( 0.736) |
|  |  | Composite | 3.36 (<0.001) | .0863 ( 0.367) | .00278 ( 0.054) |
| **Anthropology ( 4.1%)** | **Other fields** | Raw citations | 2374 ( 0.011) | -19.9 ( 0.985) | 38.4 ( 0.066) |
|  |  | Composite | 3.5 (<0.001) | -.0445 ( 0.639) | .00387 ( 0.037) |
| **Science Studies ( 3.8%)** | **Other fields** | Raw citations | 4042 ( 0.329) | 270 ( 0.950) | 27 ( 0.772) |
|  |  | Composite | 3.7 (<0.001) | -.141 ( 0.648) | .00151 ( 0.821) |
| **Information Systems ( 3.8%)** | **Other fields** | Raw citations | 5783 ( 0.003) | 46.2 ( 0.986) | 64.6 ( 0.243) |
|  |  | Composite | 3.48 (<0.001) | .00696 ( 0.945) | .00482 ( 0.021) |
| **Finance ( 3.8%)** | **Other fields** | Raw citations | 3543 ( 0.046) | -1360 ( 0.597) | 88.2 ( 0.058) |
|  |  | Composite | 3.47 (<0.001) | .00115 ( 0.993) | .00535 ( 0.019) |
| **Cultural Studies ( 3.7%)** | **Other fields** | Raw citations | 1081 ( 0.052) | -366 ( 0.650) | 2.31 ( 0.852) |
|  |  | Composite | 3.24 (<0.001) | -.0615 ( 0.710) | -.0011 ( 0.664) |
| **General Physics ( 3.7%)** | **Other fields** | Raw citations | 10053 (<0.001) | 1077 ( 0.632) | -33.2 ( 0.231) |
|  |  | Composite | 3.5 (<0.001) | .0829 ( 0.314) | .00254 ( 0.013) |
| **Chemical Engineering ( 3.6%)** | **Other fields** | Raw citations | 7632 (<0.001) | 1851 ( 0.260) | -40.8 ( 0.052) |
|  |  | Composite | 3.45 (<0.001) | .0862 ( 0.362) | .000512 ( 0.672) |
| **Mechanical Engineering & Transports ( 3.5%)** | **Other fields** | Raw citations | 4647 (<0.001) | 3195 ( 0.012) | -2.2 ( 0.902) |
|  |  | Composite | 3.29 (<0.001) | .0219 ( 0.771) | .00312 ( 0.003) |
| **Ecology ( 3.5%)** | **Other fields** | Raw citations | 6411 (<0.001) | 505 ( 0.803) | 116 ( 0.003) |
|  |  | Composite | 3.55 (<0.001) | .107 ( 0.054) | .00732 (<0.001) |
| **Operations Research ( 3.4%)** | **Other fields** | Raw citations | 820 ( 0.549) | -1333 ( 0.477) | 125 (<0.001) |
|  |  | Composite | 3.45 (<0.001) | -.0873 ( 0.397) | .00559 ( 0.003) |
| **Dairy & Animal Science ( 3.3%)** | **Other fields** | Raw citations | 4591 (<0.001) | 1254 ( 0.128) | -13.1 ( 0.278) |
|  |  | Composite | 3.26 (<0.001) | .0906 ( 0.211) | .00125 ( 0.237) |
| **Mining & Metallurgy ( 3.2%)** | **Other fields** | Raw citations | -841 ( 0.365) | 4010 ( 0.012) | 65.3 ( 0.005) |
|  |  | Composite | 2.31 (<0.001) | .515 ( 0.054) | .0149 (<0.001) |
| **Building & Construction ( 3.1%)** | **Other fields** | Raw citations | 2248 ( 0.020) | 1421 ( 0.421) | 53.4 ( 0.055) |
|  |  | Composite | 3.13 (<0.001) | .0225 ( 0.880) | .00739 ( 0.002) |
| **Philosophy ( 3.1%)** | **Other fields** | Raw citations | 1531 (<0.001) | 1526 ( 0.004) | 4.39 ( 0.514) |
|  |  | Composite | 3.4 (<0.001) | .181 ( 0.088) | .000582 ( 0.667) |
| **Political Science & Public Administration ( 3.1%)** | **Other fields** | Raw citations | 3454 ( 0.001) | 2617 ( 0.158) | 34.4 ( 0.178) |
|  |  | Composite | 3.63 (<0.001) | .0339 ( 0.715) | .00198 ( 0.124) |
| **Zoology ( 3%)** | **Other fields** | Raw citations | 1876 ( 0.023) | 3975 ( 0.004) | 20.4 ( 0.345) |
|  |  | Composite | 3.17 (<0.001) | .102 ( 0.435) | .00193 ( 0.358) |
| **Ornithology ( 3%)** | **Other fields** | Raw citations | 3524 ( 0.001) | -1396 ( 0.434) | .408 ( 0.985) |
|  |  | Composite | 3.28 (<0.001) | -.0295 ( 0.867) | .00454 ( 0.047) |
| **Design Practice & Management ( 3%)** | **Other fields** | Raw citations | 3386 (<0.001) | 1663 ( 0.313) | 11 ( 0.591) |
|  |  | Composite | 3.27 (<0.001) | -.0199 ( 0.902) | .00196 ( 0.330) |
| **Materials ( 2.9%)** | **Other fields** | Raw citations | 6797 (<0.001) | 2686 ( 0.029) | -25.3 ( 0.062) |
|  |  | Composite | 3.27 (<0.001) | .193 ( 0.002) | .00243 (<0.001) |
| **Environmental Engineering ( 2.9%)** | **Other fields** | Raw citations | 5644 (<0.001) | 1163 ( 0.395) | -4.68 ( 0.822) |
|  |  | Composite | 3.4 (<0.001) | .0259 ( 0.747) | .00278 ( 0.023) |
| **Inorganic & Nuclear Chemistry ( 2.8%)** | **Other fields** | Raw citations | 5716 ( 0.005) | 206 ( 0.951) | 67.7 ( 0.096) |
|  |  | Composite | 3.4 (<0.001) | -.122 ( 0.283) | .0048 ( 0.001) |
| **Geological & Geomatics Engineering ( 2.8%)** | **Other fields** | Raw citations | 5789 (<0.001) | 4426 ( 0.011) | 11 ( 0.684) |
|  |  | Composite | 3.3 (<0.001) | .194 ( 0.043) | .00536 (<0.001) |
| **Oceanography ( 2.7%)** | **Other fields** | Raw citations | 5601 (<0.001) | 2050 ( 0.281) | 3.64 ( 0.901) |
|  |  | Composite | 3.38 (<0.001) | .16 ( 0.114) | .00539 ( 0.001) |
| **Literary Studies ( 2.5%)** | **Other fields** | Raw citations | 260 ( 0.257) | -17.2 ( 0.971) | 10.6 ( 0.151) |
|  |  | Composite | 2.63 (<0.001) | .0703 ( 0.606) | .00518 ( 0.016) |
| **Meteorology & Atmospheric Sciences ( 2.5%)** | **Other fields** | Raw citations | 10959 (<0.001) | -2892 ( 0.123) | 15.1 ( 0.589) |
|  |  | Composite | 3.68 (<0.001) | -.0241 ( 0.648) | .0029 (<0.001) |
| **Applied Physics ( 2.4%)** | **Other fields** | Raw citations | 10388 (<0.001) | 2418 ( 0.183) | -13.5 ( 0.458) |
|  |  | Composite | 3.52 (<0.001) | .104 ( 0.037) | .00198 (<0.001) |
| **Econometrics ( 2.4%)** | **Other fields** | Raw citations | 3434 ( 0.505) | 5372 ( 0.544) | 183 ( 0.169) |
|  |  | Composite | 3.54 (<0.001) | -.211 ( 0.521) | .0111 ( 0.029) |
| **Business & Management ( 2.3%)** | **Other fields** | Raw citations | 5600 (<0.001) | -1132 ( 0.562) | 67.4 ( 0.011) |
|  |  | Composite | 3.63 (<0.001) | -.0147 ( 0.848) | .0036 ( 0.001) |
| **Computation Theory & Mathematics ( 2.3%)** | **Other fields** | Raw citations | 4982 (<0.001) | 1821 ( 0.459) | 36 ( 0.263) |
|  |  | Composite | 3.53 (<0.001) | .082 ( 0.546) | .00473 ( 0.008) |
| **Aerospace & Aeronautics ( 2.3%)** | **Other fields** | Raw citations | 2314 (<0.001) | 379 ( 0.528) | 8.24 ( 0.284) |
|  |  | Composite | 3.08 (<0.001) | -.0131 ( 0.865) | .0029 ( 0.003) |
| **Strategic, Defence & Security Studies ( 2.2%)** | **Other fields** | Raw citations | 1219 ( 0.187) | -102 ( 0.959) | 55 ( 0.026) |
|  |  | Composite | 3.14 (<0.001) | .227 ( 0.141) | .00571 ( 0.003) |
| **General Mathematics ( 2.2%)** | **Other fields** | Raw citations | 3090 (<0.001) | -413 ( 0.660) | -.84 ( 0.920) |
|  |  | Composite | 3.47 (<0.001) | -.0642 ( 0.385) | .000699 ( 0.286) |
| **Civil Engineering ( 2.2%)** | **Other fields** | Raw citations | 1891 ( 0.014) | 198 ( 0.897) | 36 ( 0.059) |
|  |  | Composite | 3.09 (<0.001) | -.00369 ( 0.978) | .00467 ( 0.006) |
| **Geology ( 2.2%)** | **Other fields** | Raw citations | 5985 (<0.001) | -1123 ( 0.524) | -38.9 ( 0.068) |
|  |  | Composite | 3.64 (<0.001) | -.00126 ( 0.994) | -.00216 ( 0.276) |
| **Energy ( 2.1%)** | **Other fields** | Raw citations | 4520 (<0.001) | 1691 ( 0.099) | 8.05 ( 0.458) |
|  |  | Composite | 3.2 (<0.001) | .0679 ( 0.296) | .00376 (<0.001) |
| **Psychoanalysis ( 2%)** | **Highly related fields** | Raw citations | 988 ( 0.497) | 2357 ( 0.380) | 15.2 ( 0.529) |
|  |  | Composite | 3.41 (<0.001) | .29 ( 0.276) | -.000634 ( 0.791) |
| **Law ( 1.9%)** | **Other fields** | Raw citations | 742 ( 0.443) | -514 ( 0.749) | 32.6 ( 0.314) |
|  |  | Composite | 3.11 (<0.001) | -.119 ( 0.453) | .0062 ( 0.054) |
| **Marine Biology & Hydrobiology ( 1.8%)** | **Other fields** | Raw citations | 7982 (<0.001) | 325 ( 0.891) | -4.01 ( 0.905) |
|  |  | Composite | 3.67 (<0.001) | .029 ( 0.767) | .0022 ( 0.113) |
| **Astronomy & Astrophysics ( 1.8%)** | **Other fields** | Raw citations | 18941 (<0.001) | -6369 ( 0.107) | -91.4 ( 0.048) |
|  |  | Composite | 3.75 (<0.001) | .0463 ( 0.466) | .00118 ( 0.113) |
| **Fisheries ( 1.8%)** | **Other fields** | Raw citations | 4259 (<0.001) | 519 ( 0.751) | 5.37 ( 0.812) |
|  |  | Composite | 3.42 (<0.001) | .0749 ( 0.559) | .00126 ( 0.478) |
| **Legal & Forensic Medicine ( 1.5%)** | **Highly related fields** | Raw citations | 1711 ( 0.119) | -1184 ( 0.649) | 12.3 ( 0.661) |
|  |  | Composite | 2.93 (<0.001) | -.158 ( 0.565) | .00263 ( 0.376) |
| **International Relations ( 1.5%)** | **Other fields** | Raw citations | 2208 ( 0.016) | -216 ( 0.926) | 27.4 ( 0.237) |
|  |  | Composite | 3.43 (<0.001) | .0318 ( 0.892) | .00393 ( 0.096) |
| **Nuclear & Particle Physics ( 1.3%)** | **Other fields** | Raw citations | 18699 (<0.001) | -7166 ( 0.161) | -103 ( 0.002) |
|  |  | Composite | 3.65 (<0.001) | -.0646 ( 0.464) | .00118 ( 0.042) |
| **Geochemistry & Geophysics ( 1.2%)** | **Other fields** | Raw citations | 7325 (<0.001) | -1353 ( 0.415) | 15.3 ( 0.348) |
|  |  | Composite | 3.66 (<0.001) | -.12 ( 0.125) | .00309 (<0.001) |
| **Forestry ( .96%)** | **Other fields** | Raw citations | 2792 (<0.001) | -1115 ( 0.520) | 16.6 ( 0.374) |
|  |  | Composite | 3.15 (<0.001) | -.0613 ( 0.739) | .00375 ( 0.060) |
| **Physical Chemistry ( .88%)** | **Other fields** | Raw citations | 11640 (<0.001) | -5354 ( 0.568) | -38.2 ( 0.499) |
|  |  | Composite | 3.49 (<0.001) | -.247 ( 0.517) | .00373 ( 0.107) |
| **Agronomy & Agriculture ( .88%)** | **Other fields** | Raw citations | 5470 (<0.001) | 1660 ( 0.508) | -.603 ( 0.975) |
|  |  | Composite | 3.36 (<0.001) | .106 ( 0.453) | .00276 ( 0.010) |
| **Paleontology ( 0%)** | **Other fields** | Raw citations |  |  |  |
|  |  | Composite |  |  |  |
| **Music ( 0%)** | **Other fields** | Raw citations |  |  |  |
|  |  | Composite |  |  |  |
| **Industrial Relations ( 0%)** | **Other fields** | Raw citations |  |  |  |
|  |  | Composite |  |  |  |
| **Horticulture ( 0%)** | **Other fields** | Raw citations |  |  |  |
|  |  | Composite |  |  |  |
| **History of Science, Technology & Medicine ( 0%)** | **Other fields** | Raw citations |  |  |  |
|  |  | Composite |  |  |  |
| **History ( 0%)** | **Other fields** | Raw citations |  |  |  |
|  |  | Composite |  |  |  |
| **General Psychology & Cognitive Sciences ( 0%)** | **Highly related fields** | Raw citations |  |  |  |
|  |  | Composite |  |  |  |
| **Folklore ( 0%)** | **Other fields** | Raw citations |  |  |  |
|  |  | Composite |  |  |  |
| **Automobile Design & Engineering ( 0%)** | **Other fields** | Raw citations |  |  |  |
|  |  | Composite |  |  |  |
| **Art Practice, History & Theory ( 0%)** | **Other fields** | Raw citations |  |  |  |
|  |  | Composite |  |  |  |
| **Architecture ( 0%)** | **Other fields** | Raw citations |  |  |  |
|  |  | Composite |  |  |  |
| **Accounting ( 0%)** | **Other fields** | Raw citations |  |  |  |
|  |  | Composite |  |  |  |

**Supplementary Table 3.3 : Career-long impact, Funding time current funding Linear Regressions for each subfield (ordered by percentage funded)**

| **Top-cited US-based researchers: Subfield (perc. funded)** | **Classification** | **Dependent Variable** | **Constant (p-val)** | **Funded (p-val)** | **Years since first pub (p-val)** |
| --- | --- | --- | --- | --- | --- |
| **Geriatrics ( 31%)** | **Highly related fields** | Raw citations | 7240 ( 0.221) | 2666 ( 0.364) | 128 ( 0.384) |
|  |  | Composite | 3.33 (<0.001) | .0879 ( 0.257) | .00962 ( 0.015) |
| **Bioinformatics ( 30%)** | **Highly related fields** | Raw citations | 18202 ( 0.005) | 3599 ( 0.376) | -2.28 ( 0.990) |
|  |  | Composite | 3.41 (<0.001) | .102 ( 0.045) | .00664 ( 0.003) |
| **Developmental Biology ( 29%)** | **Highly related fields** | Raw citations | 19299 (<0.001) | 1961 ( 0.165) | 31.3 ( 0.607) |
|  |  | Composite | 3.6 (<0.001) | .0117 ( 0.505) | .00753 (<0.001) |
| **Substance Abuse ( 23%)** | **Highly related fields** | Raw citations | 3955 ( 0.023) | 1569 ( 0.131) | 142 (<0.001) |
|  |  | Composite | 3.63 (<0.001) | -.0224 ( 0.555) | .00412 ( 0.005) |
| **Medical Informatics ( 23%)** | **Highly related fields** | Raw citations | 3976 ( 0.073) | 3468 ( 0.039) | 28.4 ( 0.625) |
|  |  | Composite | 3.21 (<0.001) | .18 ( 0.010) | .00432 ( 0.073) |
| **Virology ( 22%)** | **Highly related fields** | Raw citations | 9012 (<0.001) | 1496 ( 0.119) | 89 ( 0.015) |
|  |  | Composite | 3.45 (<0.001) | .0466 ( 0.072) | .00569 (<0.001) |
| **Biomedical Engineering ( 22%)** | **Highly related fields** | Raw citations | 4808 ( 0.003) | 3856 ( 0.002) | 72.1 ( 0.073) |
|  |  | Composite | 3.21 (<0.001) | .0523 ( 0.144) | .00655 (<0.001) |
| **Neurology & Neurosurgery ( 21%)** | **Highly related fields** | Raw citations | 14167 (<0.001) | 729 ( 0.272) | 26.9 ( 0.255) |
|  |  | Composite | 3.67 (<0.001) | .0196 ( 0.154) | .00433 (<0.001) |
| **Immunology ( 20%)** | **Highly related fields** | Raw citations | 16782 (<0.001) | 2421 ( 0.033) | 47.7 ( 0.244) |
|  |  | Composite | 3.67 (<0.001) | .0824 (<0.001) | .00456 (<0.001) |
| **Gerontology ( 20%)** | **Highly related fields** | Raw citations | 15511 ( 0.061) | 1837 ( 0.700) | -102 ( 0.601) |
|  |  | Composite | 3.53 (<0.001) | .09 ( 0.203) | .00419 ( 0.150) |
| **Applied Ethics ( 20%)** | **Highly related fields** | Raw citations | 5381 ( 0.001) | 2541 ( 0.013) | -24.2 ( 0.518) |
|  |  | Composite | 3.49 (<0.001) | .0413 ( 0.578) | .00342 ( 0.222) |
| **Public Health ( 19%)** | **Highly related fields** | Raw citations | 4564 ( 0.005) | 2088 ( 0.054) | 158 (<0.001) |
|  |  | Composite | 3.47 (<0.001) | .0841 ( 0.002) | .00642 (<0.001) |
| **Demography ( 19%)** | **Highly related fields** | Raw citations | -619 ( 0.831) | 1038 ( 0.584) | 117 ( 0.057) |
|  |  | Composite | 3.36 (<0.001) | -.0231 ( 0.837) | .00644 ( 0.078) |
| **Oncology & Carcinogenesis ( 18%)** | **Highly related fields** | Raw citations | 17848 (<0.001) | 1549 ( 0.114) | 17.1 ( 0.603) |
|  |  | Composite | 3.5 (<0.001) | .0143 ( 0.370) | .00598 (<0.001) |
| **Gastroenterology & Hepatology ( 17%)** | **Highly related fields** | Raw citations | 11847 (<0.001) | 1338 ( 0.301) | 34 ( 0.414) |
|  |  | Composite | 3.61 (<0.001) | .0665 ( 0.050) | .00373 ( 0.001) |
| **Allergy ( 16%)** | **Highly related fields** | Raw citations | 10372 ( 0.003) | 681 ( 0.782) | 43.7 ( 0.579) |
|  |  | Composite | 3.63 (<0.001) | .0272 ( 0.714) | .00373 ( 0.118) |
| **Biophysics ( 16%)** | **Highly related fields** | Raw citations | 5031 ( 0.147) | 2001 ( 0.421) | 86.4 ( 0.247) |
|  |  | Composite | 3.33 (<0.001) | .054 ( 0.458) | .00629 ( 0.004) |
| **Developmental & Child Psychology ( 16%)** | **Highly related fields** | Raw citations | 4485 ( 0.006) | 4997 (<0.001) | 123 ( 0.002) |
|  |  | Composite | 3.57 (<0.001) | .148 (<0.001) | .006 (<0.001) |
| **Health Policy & Services ( 16%)** | **Highly related fields** | Raw citations | 1789 ( 0.491) | 577 ( 0.758) | 216 ( 0.001) |
|  |  | Composite | 3.29 (<0.001) | .0579 ( 0.285) | .0104 (<0.001) |
| **Physiology ( 16%)** | **Highly related fields** | Raw citations | 6256 (<0.001) | -514 ( 0.575) | 36.5 ( 0.149) |
|  |  | Composite | 3.72 (<0.001) | -.00535 ( 0.902) | .00137 ( 0.252) |
| **Arthritis & Rheumatology ( 16%)** | **Highly related fields** | Raw citations | 10770 ( 0.004) | 1837 ( 0.478) | 154 ( 0.080) |
|  |  | Composite | 3.64 (<0.001) | .0118 ( 0.842) | .005 ( 0.014) |
| **Epidemiology ( 15%)** | **Highly related fields** | Raw citations | 1001 ( 0.926) | 10141 ( 0.183) | 496 ( 0.053) |
|  |  | Composite | 3.55 (<0.001) | .0412 ( 0.589) | .00847 ( 0.001) |
| **Psychiatry ( 15%)** | **Highly related fields** | Raw citations | 10580 (<0.001) | 4869 ( 0.002) | 131 ( 0.011) |
|  |  | Composite | 3.68 (<0.001) | .0821 ( 0.009) | .00484 (<0.001) |
| **Genetics & Heredity ( 15%)** | **Highly related fields** | Raw citations | 19705 (<0.001) | 4478 ( 0.057) | -75.7 ( 0.353) |
|  |  | Composite | 3.57 (<0.001) | .00232 ( 0.955) | .00386 ( 0.008) |
| **Emergency & Critical Care Medicine ( 15%)** | **Highly related fields** | Raw citations | 6790 ( 0.007) | 4966 ( 0.005) | 82.9 ( 0.205) |
|  |  | Composite | 3.32 (<0.001) | .106 ( 0.030) | .00623 ( 0.001) |
| **Endocrinology & Metabolism ( 15%)** | **Highly related fields** | Raw citations | 13853 (<0.001) | 1278 ( 0.354) | 44 ( 0.306) |
|  |  | Composite | 3.7 (<0.001) | .0075 ( 0.798) | .00369 (<0.001) |
| **Statistics & Probability ( 14%)** | **Other fields** | Raw citations | 11748 (<0.001) | 4219 ( 0.106) | -8.42 ( 0.898) |
|  |  | Composite | 3.72 (<0.001) | .0768 ( 0.169) | .00147 ( 0.298) |
| **Ophthalmology & Optometry ( 14%)** | **Highly related fields** | Raw citations | 10708 (<0.001) | 1751 ( 0.043) | -55.9 ( 0.033) |
|  |  | Composite | 3.5 (<0.001) | .0825 ( 0.007) | .00191 ( 0.038) |
| **Microbiology ( 14%)** | **Highly related fields** | Raw citations | 12953 (<0.001) | 1918 ( 0.026) | -35.4 ( 0.171) |
|  |  | Composite | 3.54 (<0.001) | .0542 ( 0.017) | .0046 (<0.001) |
| **Cardiovascular System & Hematology ( 14%)** | **Highly related fields** | Raw citations | 19446 (<0.001) | 3580 ( 0.014) | 17.2 ( 0.689) |
|  |  | Composite | 3.62 (<0.001) | .0499 ( 0.027) | .00472 (<0.001) |
| **Respiratory System ( 13%)** | **Highly related fields** | Raw citations | 11193 (<0.001) | -24.4 ( 0.986) | 72.1 ( 0.114) |
|  |  | Composite | 3.56 (<0.001) | .00577 ( 0.869) | .00455 (<0.001) |
| **Clinical Psychology ( 13%)** | **Highly related fields** | Raw citations | 7741 ( 0.001) | 1524 ( 0.473) | 84.7 ( 0.116) |
|  |  | Composite | 3.73 (<0.001) | .0574 ( 0.389) | .00307 ( 0.070) |
| **Nuclear Medicine & Medical Imaging ( 13%)** | **Highly related fields** | Raw citations | 6940 (<0.001) | 2480 (<0.001) | 31.7 ( 0.161) |
|  |  | Composite | 3.41 (<0.001) | .0724 ( 0.004) | .00238 ( 0.003) |
| **Analytical Chemistry ( 13%)** | **Other fields** | Raw citations | 5901 (<0.001) | 3309 ( 0.001) | 18.4 ( 0.467) |
|  |  | Composite | 3.29 (<0.001) | .0761 ( 0.071) | .00475 (<0.001) |
| **Medicinal & Biomolecular Chemistry ( 12%)** | **Highly related fields** | Raw citations | 1538 ( 0.083) | 2902 (<0.001) | 113 (<0.001) |
|  |  | Composite | 2.93 (<0.001) | .12 ( 0.002) | .00877 (<0.001) |
| **Nanoscience & Nanotechnology ( 12%)** | **Other fields** | Raw citations | 7634 ( 0.009) | 449 ( 0.891) | 497 (<0.001) |
|  |  | Composite | 3.3 (<0.001) | .0263 ( 0.617) | .0143 (<0.001) |
| **Urology & Nephrology ( 12%)** | **Highly related fields** | Raw citations | 14065 (<0.001) | 1537 ( 0.237) | -45.1 ( 0.238) |
|  |  | Composite | 3.58 (<0.001) | .0714 ( 0.031) | .00308 ( 0.002) |
| **Optics ( 12%)** | **Other fields** | Raw citations | 7949 (<0.001) | 4544 ( 0.001) | 1.31 ( 0.973) |
|  |  | Composite | 3.26 (<0.001) | .0423 ( 0.410) | .00481 ( 0.001) |
| **Biochemistry & Molecular Biology ( 12%)** | **Highly related fields** | Raw citations | 12813 (<0.001) | 1695 ( 0.062) | 10.4 ( 0.666) |
|  |  | Composite | 3.68 (<0.001) | .0445 ( 0.035) | .00326 (<0.001) |
| **Nutrition & Dietetics ( 11%)** | **Highly related fields** | Raw citations | 8095 (<0.001) | 2690 ( 0.118) | 43.3 ( 0.344) |
|  |  | Composite | 3.52 (<0.001) | .047 ( 0.364) | .00527 (<0.001) |
| **Experimental Psychology ( 11%)** | **Highly related fields** | Raw citations | 6337 (<0.001) | 4191 (<0.001) | 51.2 ( 0.030) |
|  |  | Composite | 3.67 (<0.001) | .157 (<0.001) | .0037 (<0.001) |
| **Rehabilitation ( 11%)** | **Highly related fields** | Raw citations | 3920 ( 0.001) | 2184 ( 0.012) | 34.2 ( 0.235) |
|  |  | Composite | 3.3 (<0.001) | .0716 ( 0.182) | .00462 ( 0.010) |
| **Toxicology ( 11%)** | **Highly related fields** | Raw citations | 8361 (<0.001) | 4376 (<0.001) | -29 ( 0.363) |
|  |  | Composite | 3.35 (<0.001) | .114 ( 0.009) | .00394 ( 0.002) |
| **Drama & Theater ( 11%)** | **Other fields** | Raw citations | -188 ( 0.513) | 379 ( 0.116) | 21.8 ( 0.051) |
|  |  | Composite | 2.24 (<0.001) | .246 ( 0.204) | .0169 ( 0.065) |
| **Obstetrics & Reproductive Medicine ( 11%)** | **Highly related fields** | Raw citations | 9867 (<0.001) | 592 ( 0.485) | -44.4 ( 0.054) |
|  |  | Composite | 3.51 (<0.001) | .0377 ( 0.273) | .00197 ( 0.034) |
| **Pediatrics ( 11%)** | **Highly related fields** | Raw citations | 6378 (<0.001) | 1513 ( 0.037) | 16.2 ( 0.382) |
|  |  | Composite | 3.3 (<0.001) | .0813 ( 0.016) | .00428 (<0.001) |
| **Anesthesiology ( 11%)** | **Highly related fields** | Raw citations | 6214 (<0.001) | 3010 ( 0.001) | 5.13 ( 0.845) |
|  |  | Composite | 3.41 (<0.001) | .0984 ( 0.041) | .00223 ( 0.099) |
| **Pharmacology & Pharmacy ( 10%)** | **Highly related fields** | Raw citations | 5187 (<0.001) | 1667 ( 0.031) | 28.8 ( 0.125) |
|  |  | Composite | 3.32 (<0.001) | .0433 ( 0.245) | .00339 (<0.001) |
| **Speech-Language Pathology & Audiology ( 9.9%)** | **Highly related fields** | Raw citations | 6996 (<0.001) | -1163 ( 0.368) | -55.1 ( 0.197) |
|  |  | Composite | 3.36 (<0.001) | -.0243 ( 0.758) | .00426 ( 0.103) |
| **Organic Chemistry ( 9.8%)** | **Other fields** | Raw citations | 8871 (<0.001) | 3838 ( 0.007) | 15.3 ( 0.587) |
|  |  | Composite | 3.52 (<0.001) | .121 ( 0.007) | .00385 (<0.001) |
| **Environmental & Occupational Health ( 9.8%)** | **Highly related fields** | Raw citations | 5081 ( 0.019) | -1098 ( 0.453) | 5.49 ( 0.914) |
|  |  | Composite | 3.22 (<0.001) | -.000389 ( 0.996) | .00377 ( 0.136) |
| **Family Studies ( 8.9%)** | **Other fields** | Raw citations | 180 ( 0.895) | 1669 ( 0.209) | 96.4 ( 0.007) |
|  |  | Composite | 3.18 (<0.001) | .0128 ( 0.898) | .00977 (<0.001) |
| **Development Studies ( 8.3%)** | **Other fields** | Raw citations | 5159 ( 0.019) | -48.8 ( 0.974) | -36.5 ( 0.494) |
|  |  | Composite | 3.86 (<0.001) | -.177 ( 0.310) | -.00532 ( 0.389) |
| **Mycology & Parasitology ( 8.2%)** | **Highly related fields** | Raw citations | 8329 (<0.001) | -479 ( 0.807) | -43.2 ( 0.313) |
|  |  | Composite | 3.4 (<0.001) | .0122 ( 0.883) | .00232 ( 0.202) |
| **Nursing ( 7.9%)** | **Highly related fields** | Raw citations | 1304 (<0.001) | 2989 (<0.001) | 29.4 ( 0.001) |
|  |  | Composite | 3.01 (<0.001) | .173 (<0.001) | .00347 (<0.001) |
| **Surgery ( 7.9%)** | **Highly related fields** | Raw citations | 9513 (<0.001) | 3110 ( 0.001) | -24.9 ( 0.147) |
|  |  | Composite | 3.45 (<0.001) | .0473 ( 0.152) | .000912 ( 0.147) |
| **Tropical Medicine ( 7.8%)** | **Highly related fields** | Raw citations | 8018 (<0.001) | 2713 ( 0.097) | -33.2 ( 0.314) |
|  |  | Composite | 3.38 (<0.001) | .0795 ( 0.276) | .00152 ( 0.304) |
| **General Chemistry ( 7.8%)** | **Other fields** | Raw citations | 13531 (<0.001) | 5432 ( 0.075) | -74 ( 0.039) |
|  |  | Composite | 3.36 (<0.001) | .302 (<0.001) | .00251 ( 0.009) |
| **Complementary & Alternative Medicine ( 7.4%)** | **Highly related fields** | Raw citations | 2244 ( 0.010) | 2381 ( 0.019) | -2.69 ( 0.916) |
|  |  | Composite | 3 (<0.001) | .259 ( 0.078) | -.00054 ( 0.885) |
| **General & Internal Medicine ( 7.1%)** | **Highly related fields** | Raw citations | 13326 (<0.001) | 2641 ( 0.097) | -114 (<0.001) |
|  |  | Composite | 3.31 (<0.001) | .113 ( 0.006) | -.000135 ( 0.813) |
| **Biotechnology ( 7%)** | **Highly related fields** | Raw citations | 4261 ( 0.009) | 1527 ( 0.415) | 92.5 ( 0.036) |
|  |  | Composite | 3.12 (<0.001) | .159 ( 0.053) | .00851 (<0.001) |
| **Microscopy ( 6.7%)** | **Highly related fields** | Raw citations | 6035 ( 0.129) | 3451 ( 0.362) | 23.3 ( 0.796) |
|  |  | Composite | 3.26 (<0.001) | .322 ( 0.163) | .0056 ( 0.307) |
| **Acoustics ( 6.5%)** | **Other fields** | Raw citations | 5840 (<0.001) | 570 ( 0.497) | -37.1 ( 0.012) |
|  |  | Composite | 3.53 (<0.001) | -.0568 ( 0.357) | -.00103 ( 0.336) |
| **Sociology ( 6.4%)** | **Other fields** | Raw citations | 2704 ( 0.004) | 609 ( 0.617) | 58.2 ( 0.010) |
|  |  | Composite | 3.57 (<0.001) | .00735 ( 0.917) | .0041 ( 0.002) |
| **Otorhinolaryngology ( 6.3%)** | **Highly related fields** | Raw citations | 5652 (<0.001) | 1622 ( 0.018) | -26.2 ( 0.061) |
|  |  | Composite | 3.29 (<0.001) | .0981 ( 0.026) | .00272 ( 0.002) |
| **Behavioral Science & Comparative Psychology ( 6.3%)** | **Highly related fields** | Raw citations | 3888 ( 0.003) | 1817 ( 0.142) | 43.9 ( 0.110) |
|  |  | Composite | 3.57 (<0.001) | .0329 ( 0.644) | .00469 ( 0.003) |
| **Orthopedics ( 5.9%)** | **Highly related fields** | Raw citations | 7766 (<0.001) | 3284 ( 0.001) | -6.91 ( 0.723) |
|  |  | Composite | 3.4 (<0.001) | .157 (<0.001) | .0041 (<0.001) |
| **Sport Sciences ( 5.9%)** | **Highly related fields** | Raw citations | 3800 ( 0.079) | 1673 ( 0.534) | 105 ( 0.066) |
|  |  | Composite | 3.45 (<0.001) | -.0187 ( 0.836) | .0039 ( 0.041) |
| **Social Work ( 5.7%)** | **Other fields** | Raw citations | 1135 ( 0.074) | 1393 ( 0.052) | 25.7 ( 0.151) |
|  |  | Composite | 3.21 (<0.001) | .117 ( 0.205) | .00187 ( 0.419) |
| **Economic Theory ( 5.6%)** | **Other fields** | Raw citations | 2749 ( 0.150) | -9.76 ( 0.996) | 20 ( 0.616) |
|  |  | Composite | 3.49 (<0.001) | -.145 ( 0.287) | .0029 ( 0.285) |
| **Evolutionary Biology ( 5.5%)** | **Other fields** | Raw citations | 4994 ( 0.035) | 3554 ( 0.261) | 138 ( 0.019) |
|  |  | Composite | 3.62 (<0.001) | .0811 ( 0.203) | .00648 (<0.001) |
| **Dermatology & Venereal Diseases ( 5.5%)** | **Highly related fields** | Raw citations | 9162 (<0.001) | 6568 (<0.001) | -21.1 ( 0.493) |
|  |  | Composite | 3.62 (<0.001) | .0743 ( 0.268) | .000661 ( 0.573) |
| **Artificial Intelligence & Image Processing ( 5.5%)** | **Other fields** | Raw citations | 5687 (<0.001) | 1298 ( 0.290) | 64.8 ( 0.011) |
|  |  | Composite | 3.19 (<0.001) | .0637 ( 0.136) | .00829 (<0.001) |
| **Environmental Sciences ( 5.5%)** | **Other fields** | Raw citations | 5263 (<0.001) | 4227 ( 0.012) | 36.3 ( 0.282) |
|  |  | Composite | 3.27 (<0.001) | .152 ( 0.024) | .00552 (<0.001) |
| **Plant Biology & Botany ( 5.2%)** | **Other fields** | Raw citations | 7870 (<0.001) | 6208 (<0.001) | -5.94 ( 0.779) |
|  |  | Composite | 3.44 (<0.001) | .198 (<0.001) | .00326 ( 0.001) |
| **Pathology ( 5.1%)** | **Highly related fields** | Raw citations | 15555 (<0.001) | -2297 ( 0.413) | -79.6 ( 0.138) |
|  |  | Composite | 3.59 (<0.001) | -.0478 ( 0.569) | .00245 ( 0.126) |
| **Chemical Physics ( 4.8%)** | **Other fields** | Raw citations | 12058 (<0.001) | 4285 ( 0.101) | 37.7 ( 0.284) |
|  |  | Composite | 3.77 (<0.001) | .133 ( 0.011) | .00204 ( 0.004) |
| **Industrial Engineering & Automation ( 4.8%)** | **Other fields** | Raw citations | 5880 (<0.001) | 1965 ( 0.078) | 3.68 ( 0.859) |
|  |  | Composite | 3.39 (<0.001) | .0694 ( 0.284) | .0033 ( 0.006) |
| **Entomology ( 4.8%)** | **Other fields** | Raw citations | 4855 (<0.001) | 3779 (<0.001) | -14.5 ( 0.322) |
|  |  | Composite | 3.49 (<0.001) | .231 (<0.001) | .000319 ( 0.774) |
| **Criminology ( 4.6%)** | **Other fields** | Raw citations | 2985 ( 0.008) | 756 ( 0.616) | 54 ( 0.060) |
|  |  | Composite | 3.5 (<0.001) | .0849 ( 0.353) | .00435 ( 0.013) |
| **Religions & Theology ( 4.4%)** | **Other fields** | Raw citations | 1003 ( 0.019) | -52.6 ( 0.946) | -2.99 ( 0.780) |
|  |  | Composite | 2.93 (<0.001) | .0854 ( 0.587) | .0000947 ( 0.965) |
| **Sport, Leisure & Tourism ( 4.3%)** | **Other fields** | Raw citations | 3099 ( 0.012) | -972 ( 0.587) | 47.7 ( 0.228) |
|  |  | Composite | 3.44 (<0.001) | -.0256 ( 0.859) | .00615 ( 0.057) |
| **Distributed Computing ( 4.3%)** | **Other fields** | Raw citations | 2342 ( 0.154) | 8703 (<0.001) | 66.8 ( 0.171) |
|  |  | Composite | 3.14 (<0.001) | .217 ( 0.060) | .00249 ( 0.374) |
| **Dentistry ( 4.3%)** | **Highly related fields** | Raw citations | 5429 (<0.001) | 548 ( 0.585) | -11.7 ( 0.488) |
|  |  | Composite | 3.35 (<0.001) | .043 ( 0.476) | .00248 ( 0.015) |
| **Social Sciences Methods ( 3.9%)** | **Other fields** | Raw citations | 6813 ( 0.127) | 14216 ( 0.087) | 51.6 ( 0.565) |
|  |  | Composite | 3.63 (<0.001) | .514 ( 0.005) | .00383 ( 0.049) |
| **Science Studies ( 3.8%)** | **Other fields** | Raw citations | 4042 ( 0.329) | 270 ( 0.950) | 27 ( 0.772) |
|  |  | Composite | 3.7 (<0.001) | -.141 ( 0.648) | .00151 ( 0.821) |
| **Logistics & Transportation ( 3.8%)** | **Other fields** | Raw citations | 3206 (<0.001) | -955 ( 0.416) | 12.7 ( 0.525) |
|  |  | Composite | 3.24 (<0.001) | -.195 ( 0.094) | .00382 ( 0.055) |
| **Veterinary Sciences ( 3.8%)** | **Highly related fields** | Raw citations | 3398 (<0.001) | 1524 ( 0.022) | 13.7 ( 0.283) |
|  |  | Composite | 3.24 (<0.001) | .0639 ( 0.173) | .00267 ( 0.003) |
| **Social Psychology ( 3.7%)** | **Highly related fields** | Raw citations | 7827 (<0.001) | 1501 ( 0.570) | 73 ( 0.036) |
|  |  | Composite | 3.71 (<0.001) | .125 ( 0.124) | .004 (<0.001) |
| **Anatomy & Morphology ( 3.7%)** | **Highly related fields** | Raw citations | 2676 ( 0.005) | 858 ( 0.651) | -10.9 ( 0.614) |
|  |  | Composite | 2.96 (<0.001) | -.105 ( 0.571) | .0043 ( 0.046) |
| **Applied Mathematics ( 3.6%)** | **Other fields** | Raw citations | 3765 ( 0.226) | 2805 ( 0.554) | 104 ( 0.139) |
|  |  | Composite | 3.51 (<0.001) | .0465 ( 0.784) | .00517 ( 0.040) |
| **Polymers ( 3.6%)** | **Other fields** | Raw citations | 9496 (<0.001) | 2070 ( 0.418) | -19.1 ( 0.551) |
|  |  | Composite | 3.52 (<0.001) | -.00218 ( 0.979) | .0016 ( 0.128) |
| **Information & Library Sciences ( 3.4%)** | **Other fields** | Raw citations | 682 ( 0.257) | 172 ( 0.879) | 44.6 ( 0.009) |
|  |  | Composite | 3.02 (<0.001) | .0424 ( 0.773) | .00689 ( 0.002) |
| **Communication & Media Studies ( 3.2%)** | **Other fields** | Raw citations | 1000 ( 0.278) | 585 ( 0.762) | 81.2 ( 0.002) |
|  |  | Composite | 3.45 (<0.001) | .0306 ( 0.771) | .00336 ( 0.016) |
| **Mining & Metallurgy ( 3.2%)** | **Other fields** | Raw citations | -841 ( 0.365) | 4010 ( 0.012) | 65.3 ( 0.005) |
|  |  | Composite | 2.31 (<0.001) | .515 ( 0.054) | .0149 (<0.001) |
| **General Clinical Medicine ( 3.1%)** | **Highly related fields** | Raw citations | 4741 ( 0.004) | 7767 ( 0.004) | -13.8 ( 0.698) |
|  |  | Composite | 2.99 (<0.001) | .441 ( 0.015) | .00414 ( 0.089) |
| **Marketing ( 3.1%)** | **Other fields** | Raw citations | 2302 ( 0.264) | -1698 ( 0.580) | 159 ( 0.008) |
|  |  | Composite | 3.56 (<0.001) | -.0515 ( 0.673) | .00476 ( 0.044) |
| **Economics ( 3.1%)** | **Other fields** | Raw citations | 6137 (<0.001) | 4725 ( 0.010) | 16.6 ( 0.492) |
|  |  | Composite | 3.61 (<0.001) | .255 ( 0.001) | .00451 (<0.001) |
| **Software Engineering ( 3.1%)** | **Other fields** | Raw citations | 7314 (<0.001) | 1442 ( 0.279) | -56.8 ( 0.020) |
|  |  | Composite | 3.5 (<0.001) | .058 ( 0.543) | .000601 ( 0.731) |
| **Ornithology ( 3%)** | **Other fields** | Raw citations | 3524 ( 0.001) | -1396 ( 0.434) | .408 ( 0.985) |
|  |  | Composite | 3.28 (<0.001) | -.0295 ( 0.867) | .00454 ( 0.047) |
| **Finance ( 3%)** | **Other fields** | Raw citations | 3568 ( 0.045) | -637 ( 0.824) | 86.6 ( 0.062) |
|  |  | Composite | 3.48 (<0.001) | .0402 ( 0.774) | .00527 ( 0.020) |
| **Human Factors ( 3%)** | **Highly related fields** | Raw citations | 4424 (<0.001) | 451 ( 0.827) | 63.8 ( 0.066) |
|  |  | Composite | 3.4 (<0.001) | .18 ( 0.139) | .00723 (<0.001) |
| **Agricultural Economics & Policy ( 2.7%)** | **Other fields** | Raw citations | 6469 (<0.001) | -2200 ( 0.375) | -29.9 ( 0.446) |
|  |  | Composite | 3.67 (<0.001) | -.077 ( 0.654) | -.000359 ( 0.895) |
| **Computer Hardware & Architecture ( 2.6%)** | **Other fields** | Raw citations | 5135 (<0.001) | 358 ( 0.838) | 11.7 ( 0.670) |
|  |  | Composite | 3.16 (<0.001) | .0267 ( 0.827) | .00673 ( 0.001) |
| **Networking & Telecommunications ( 2.6%)** | **Other fields** | Raw citations | 5889 (<0.001) | -456 ( 0.679) | -7.97 ( 0.529) |
|  |  | Composite | 3.18 (<0.001) | -.00216 ( 0.970) | .00456 (<0.001) |
| **Political Science & Public Administration ( 2.6%)** | **Other fields** | Raw citations | 3341 ( 0.001) | 3546 ( 0.077) | 37.1 ( 0.147) |
|  |  | Composite | 3.62 (<0.001) | .0847 ( 0.400) | .00208 ( 0.107) |
| **Food Science ( 2.6%)** | **Other fields** | Raw citations | 5514 (<0.001) | 3799 ( 0.130) | 4.56 ( 0.880) |
|  |  | Composite | 3.37 (<0.001) | .0365 ( 0.755) | .00255 ( 0.072) |
| **Education ( 2.6%)** | **Other fields** | Raw citations | 1755 (<0.001) | 3119 (<0.001) | 43.9 (<0.001) |
|  |  | Composite | 3.25 (<0.001) | .199 ( 0.001) | .005 (<0.001) |
| **Dairy & Animal Science ( 2.4%)** | **Other fields** | Raw citations | 4689 (<0.001) | 522 ( 0.585) | -14.7 ( 0.224) |
|  |  | Composite | 3.27 (<0.001) | .0237 ( 0.778) | .00112 ( 0.290) |
| **Econometrics ( 2.4%)** | **Other fields** | Raw citations | 3434 ( 0.505) | 5372 ( 0.544) | 183 ( 0.169) |
|  |  | Composite | 3.54 (<0.001) | -.211 ( 0.521) | .0111 ( 0.029) |
| **Mechanical Engineering & Transports ( 2.3%)** | **Other fields** | Raw citations | 5149 (<0.001) | -537 ( 0.734) | -10.7 ( 0.551) |
|  |  | Composite | 3.31 (<0.001) | -.0951 ( 0.310) | .00288 ( 0.007) |
| **Strategic, Defence & Security Studies ( 2.2%)** | **Other fields** | Raw citations | 1213 ( 0.190) | 205 ( 0.917) | 55 ( 0.026) |
|  |  | Composite | 3.14 (<0.001) | .175 ( 0.257) | .00572 ( 0.003) |
| **Electrical & Electronic Engineering ( 2.2%)** | **Other fields** | Raw citations | 3374 (<0.001) | 547 ( 0.739) | 20.3 ( 0.241) |
|  |  | Composite | 3.03 (<0.001) | -.0614 ( 0.604) | .00375 ( 0.003) |
| **Geography ( 2.2%)** | **Other fields** | Raw citations | 2410 ( 0.042) | -36 ( 0.987) | 50.8 ( 0.089) |
|  |  | Composite | 3.55 (<0.001) | -.0617 ( 0.718) | .00485 ( 0.032) |
| **Chemical Engineering ( 2.1%)** | **Other fields** | Raw citations | 7839 (<0.001) | -1467 ( 0.491) | -43.2 ( 0.040) |
|  |  | Composite | 3.46 (<0.001) | -.0154 ( 0.900) | .000427 ( 0.724) |
| **General Physics ( 2%)** | **Other fields** | Raw citations | 9983 (<0.001) | 2744 ( 0.359) | -32.1 ( 0.248) |
|  |  | Composite | 3.5 (<0.001) | .102 ( 0.351) | .00253 ( 0.013) |
| **Optoelectronics & Photonics ( 2%)** | **Other fields** | Raw citations | 3306 (<0.001) | 3117 ( 0.008) | 27.7 ( 0.063) |
|  |  | Composite | 2.99 (<0.001) | .125 ( 0.108) | .00575 (<0.001) |
| **Psychoanalysis ( 2%)** | **Highly related fields** | Raw citations | 988 ( 0.497) | 2357 ( 0.380) | 15.2 ( 0.529) |
|  |  | Composite | 3.41 (<0.001) | .29 ( 0.276) | -.000634 ( 0.791) |
| **Design Practice & Management ( 2%)** | **Other fields** | Raw citations | 3446 (<0.001) | 1632 ( 0.415) | 9.81 ( 0.629) |
|  |  | Composite | 3.26 (<0.001) | .0583 ( 0.767) | .00209 ( 0.297) |
| **Operations Research ( 2%)** | **Other fields** | Raw citations | 758 ( 0.580) | -544 ( 0.825) | 126 (<0.001) |
|  |  | Composite | 3.44 (<0.001) | .0117 ( 0.931) | .00566 ( 0.002) |
| **Environmental Engineering ( 2%)** | **Other fields** | Raw citations | 5584 (<0.001) | 2746 ( 0.098) | -3.67 ( 0.859) |
|  |  | Composite | 3.4 (<0.001) | .0912 ( 0.349) | .00283 ( 0.020) |
| **Information Systems ( 1.9%)** | **Other fields** | Raw citations | 5690 ( 0.004) | 2803 ( 0.455) | 65.8 ( 0.233) |
|  |  | Composite | 3.48 (<0.001) | .0686 ( 0.625) | .00484 ( 0.020) |
| **Literary Studies ( 1.9%)** | **Other fields** | Raw citations | 258 ( 0.258) | 15.9 ( 0.976) | 10.6 ( 0.150) |
|  |  | Composite | 2.63 (<0.001) | .104 ( 0.508) | .00514 ( 0.016) |
| **Cultural Studies ( 1.9%)** | **Other fields** | Raw citations | 1070 ( 0.054) | -72.8 ( 0.949) | 2.28 ( 0.854) |
|  |  | Composite | 3.23 (<0.001) | .11 ( 0.637) | -.00117 ( 0.647) |
| **Languages & Linguistics ( 1.8%)** | **Other fields** | Raw citations | 1367 ( 0.015) | 264 ( 0.820) | 29.1 ( 0.047) |
|  |  | Composite | 3.36 (<0.001) | .155 ( 0.341) | .0034 ( 0.097) |
| **Fisheries ( 1.8%)** | **Other fields** | Raw citations | 4290 (<0.001) | 3379 ( 0.038) | 3.33 ( 0.882) |
|  |  | Composite | 3.42 (<0.001) | .218 ( 0.089) | .00115 ( 0.516) |
| **Archaeology ( 1.8%)** | **Other fields** | Raw citations | 2083 ( 0.003) | -452 ( 0.795) | 15.6 ( 0.373) |
|  |  | Composite | 3.23 (<0.001) | -.0732 ( 0.673) | .00425 ( 0.017) |
| **Computation Theory & Mathematics ( 1.7%)** | **Other fields** | Raw citations | 5046 (<0.001) | -1823 ( 0.520) | 36.2 ( 0.261) |
|  |  | Composite | 3.53 (<0.001) | -.0894 ( 0.567) | .00475 ( 0.008) |
| **Numerical & Computational Mathematics ( 1.7%)** | **Other fields** | Raw citations | 1528 ( 0.329) | 5377 ( 0.080) | 81.3 ( 0.022) |
|  |  | Composite | 3.56 (<0.001) | .335 ( 0.062) | .00149 ( 0.467) |
| **Urban & Regional Planning ( 1.6%)** | **Other fields** | Raw citations | 3774 ( 0.005) | -145 ( 0.961) | -3.4 ( 0.918) |
|  |  | Composite | 3.57 (<0.001) | -.265 ( 0.350) | .00131 ( 0.675) |
| **Anthropology ( 1.6%)** | **Other fields** | Raw citations | 2335 ( 0.011) | 647 ( 0.694) | 39.1 ( 0.058) |
|  |  | Composite | 3.49 (<0.001) | -.0296 ( 0.840) | .00399 ( 0.030) |
| **Materials ( 1.6%)** | **Other fields** | Raw citations | 6837 (<0.001) | 3504 ( 0.034) | -25.7 ( 0.058) |
|  |  | Composite | 3.28 (<0.001) | .245 ( 0.004) | .00239 ( 0.001) |
| **Fluids & Plasmas ( 1.6%)** | **Other fields** | Raw citations | 13116 (<0.001) | 6896 ( 0.170) | -74.3 ( 0.154) |
|  |  | Composite | 3.73 (<0.001) | .223 ( 0.058) | .00153 ( 0.208) |
| **General Mathematics ( 1.6%)** | **Other fields** | Raw citations | 3092 (<0.001) | -777 ( 0.478) | -.816 ( 0.922) |
|  |  | Composite | 3.47 (<0.001) | -.149 ( 0.084) | .000699 ( 0.285) |
| **Building & Construction ( 1.6%)** | **Other fields** | Raw citations | 2339 ( 0.014) | 1702 ( 0.489) | 51.2 ( 0.064) |
|  |  | Composite | 3.13 (<0.001) | .107 ( 0.606) | .00737 ( 0.002) |
| **Philosophy ( 1.5%)** | **Other fields** | Raw citations | 1638 (<0.001) | 3821 (<0.001) | 1.48 ( 0.813) |
|  |  | Composite | 3.42 (<0.001) | .595 (<0.001) | .000139 ( 0.914) |
| **Meteorology & Atmospheric Sciences ( 1.5%)** | **Other fields** | Raw citations | 10829 (<0.001) | -1970 ( 0.409) | 17.3 ( 0.536) |
|  |  | Composite | 3.68 (<0.001) | -.0122 ( 0.856) | .00292 (<0.001) |
| **Aerospace & Aeronautics ( 1.5%)** | **Other fields** | Raw citations | 2328 (<0.001) | 625 ( 0.400) | 7.86 ( 0.306) |
|  |  | Composite | 3.08 (<0.001) | -.0421 ( 0.659) | .00292 ( 0.003) |
| **Energy ( 1.4%)** | **Other fields** | Raw citations | 4530 (<0.001) | 2329 ( 0.065) | 7.86 ( 0.468) |
|  |  | Composite | 3.2 (<0.001) | .0433 ( 0.588) | .00374 (<0.001) |
| **Geological & Geomatics Engineering ( 1.4%)** | **Other fields** | Raw citations | 5922 (<0.001) | 5192 ( 0.033) | 8.83 ( 0.744) |
|  |  | Composite | 3.31 (<0.001) | .214 ( 0.112) | .00526 ( 0.001) |
| **Ecology ( 1.3%)** | **Other fields** | Raw citations | 6424 (<0.001) | 914 ( 0.777) | 116 ( 0.003) |
|  |  | Composite | 3.55 (<0.001) | .176 ( 0.048) | .00727 (<0.001) |
| **Oceanography ( 1.1%)** | **Other fields** | Raw citations | 5634 (<0.001) | 4612 ( 0.121) | 3 ( 0.918) |
|  |  | Composite | 3.39 (<0.001) | .163 ( 0.306) | .00534 ( 0.001) |
| **Business & Management ( .95%)** | **Other fields** | Raw citations | 5565 (<0.001) | 216 ( 0.943) | 67.6 ( 0.011) |
|  |  | Composite | 3.63 (<0.001) | .0771 ( 0.511) | .0036 ( 0.001) |
| **Law ( .94%)** | **Other fields** | Raw citations | 737 ( 0.447) | -826 ( 0.715) | 32.7 ( 0.312) |
|  |  | Composite | 3.11 (<0.001) | -.0805 ( 0.719) | .00612 ( 0.057) |
| **Applied Physics ( .94%)** | **Other fields** | Raw citations | 10460 (<0.001) | 1586 ( 0.582) | -14.2 ( 0.436) |
|  |  | Composite | 3.53 (<0.001) | .124 ( 0.118) | .00196 (<0.001) |
| **Marine Biology & Hydrobiology ( .91%)** | **Other fields** | Raw citations | 7907 (<0.001) | 4376 ( 0.190) | -3.06 ( 0.927) |
|  |  | Composite | 3.66 (<0.001) | .246 ( 0.073) | .00225 ( 0.104) |
| **Physical Chemistry ( .88%)** | **Other fields** | Raw citations | 11640 (<0.001) | -5354 ( 0.568) | -38.2 ( 0.499) |
|  |  | Composite | 3.49 (<0.001) | -.247 ( 0.517) | .00373 ( 0.107) |
| **Nuclear & Particle Physics ( .78%)** | **Other fields** | Raw citations | 18451 (<0.001) | -417 ( 0.949) | -99.6 ( 0.003) |
|  |  | Composite | 3.64 (<0.001) | .038 ( 0.735) | .00122 ( 0.034) |
| **Astronomy & Astrophysics ( .76%)** | **Other fields** | Raw citations | 18707 (<0.001) | -5138 ( 0.391) | -87.1 ( 0.060) |
|  |  | Composite | 3.75 (<0.001) | .149 ( 0.121) | .00118 ( 0.110) |
| **Civil Engineering ( .73%)** | **Other fields** | Raw citations | 1891 ( 0.014) | 336 ( 0.898) | 36 ( 0.059) |
|  |  | Composite | 3.08 (<0.001) | .151 ( 0.519) | .00471 ( 0.006) |
| **Geochemistry & Geophysics ( .71%)** | **Other fields** | Raw citations | 7325 (<0.001) | -1900 ( 0.374) | 15.2 ( 0.350) |
|  |  | Composite | 3.66 (<0.001) | -.176 ( 0.081) | .00308 (<0.001) |
| **Agronomy & Agriculture ( .66%)** | **Other fields** | Raw citations | 5473 (<0.001) | 3108 ( 0.282) | -.802 ( 0.966) |
|  |  | Composite | 3.36 (<0.001) | .218 ( 0.181) | .00275 ( 0.010) |
| **Inorganic & Nuclear Chemistry ( .63%)** | **Other fields** | Raw citations | 5850 ( 0.003) | -3669 ( 0.595) | 65.5 ( 0.102) |
|  |  | Composite | 3.38 (<0.001) | -.147 ( 0.531) | .00501 (<0.001) |
| **Forestry ( .48%)** | **Other fields** | Raw citations | 2816 (<0.001) | -2168 ( 0.376) | 16 ( 0.395) |
|  |  | Composite | 3.15 (<0.001) | -.171 ( 0.510) | .00368 ( 0.065) |
| **Zoology ( 0%)** | **Other fields** | Raw citations |  |  |  |
|  |  | Composite |  |  |  |
| **Paleontology ( 0%)** | **Other fields** | Raw citations |  |  |  |
|  |  | Composite |  |  |  |
| **Music ( 0%)** | **Other fields** | Raw citations |  |  |  |
|  |  | Composite |  |  |  |
| **Mathematical Physics ( 0%)** | **Other fields** | Raw citations |  |  |  |
|  |  | Composite |  |  |  |
| **Legal & Forensic Medicine ( 0%)** | **Highly related fields** | Raw citations |  |  |  |
|  |  | Composite |  |  |  |
| **International Relations ( 0%)** | **Other fields** | Raw citations |  |  |  |
|  |  | Composite |  |  |  |
| **Industrial Relations ( 0%)** | **Other fields** | Raw citations |  |  |  |
|  |  | Composite |  |  |  |
| **Horticulture ( 0%)** | **Other fields** | Raw citations |  |  |  |
|  |  | Composite |  |  |  |
| **History of Social Sciences ( 0%)** | **Other fields** | Raw citations |  |  |  |
|  |  | Composite |  |  |  |
| **History of Science, Technology & Medicine ( 0%)** | **Other fields** | Raw citations |  |  |  |
|  |  | Composite |  |  |  |
| **History ( 0%)** | **Other fields** | Raw citations |  |  |  |
|  |  | Composite |  |  |  |
| **Geology ( 0%)** | **Other fields** | Raw citations |  |  |  |
|  |  | Composite |  |  |  |
| **General Psychology & Cognitive Sciences ( 0%)** | **Highly related fields** | Raw citations |  |  |  |
|  |  | Composite |  |  |  |
| **Gender Studies ( 0%)** | **Highly related fields** | Raw citations |  |  |  |
|  |  | Composite |  |  |  |
| **Folklore ( 0%)** | **Other fields** | Raw citations |  |  |  |
|  |  | Composite |  |  |  |
| **Classics ( 0%)** | **Other fields** | Raw citations |  |  |  |
|  |  | Composite |  |  |  |
| **Automobile Design & Engineering ( 0%)** | **Other fields** | Raw citations |  |  |  |
|  |  | Composite |  |  |  |
| **Art Practice, History & Theory ( 0%)** | **Other fields** | Raw citations |  |  |  |
|  |  | Composite |  |  |  |
| **Architecture ( 0%)** | **Other fields** | Raw citations |  |  |  |
|  |  | Composite |  |  |  |
| **Accounting ( 0%)** | **Other fields** | Raw citations |  |  |  |
|  |  | Composite |  |  |  |

**Supplementary Table 3.4 : Recent year impact, Funding time any funding Linear Regressions for each subfield (ordered by percentage funded)**

| **Top-cited US-based researchers: Subfield (perc. funded)** | **Classification** | **Dependent Variable** | **Constant (p-val)** | **Funded (p-val)** | **Years since first pub (p-val)** |
| --- | --- | --- | --- | --- | --- |
| **Geriatrics ( 88%)** | **Highly related fields** | Raw citations | 768 ( 0.390) | 1029 ( 0.122) | 9.23 ( 0.632) |
|  |  | Composite | 2.84 (<0.001) | .25 ( 0.038) | .00409 ( 0.240) |
| **Gerontology ( 87%)** | **Highly related fields** | Raw citations | 392 ( 0.752) | 1370 ( 0.138) | 11.2 ( 0.680) |
|  |  | Composite | 2.87 (<0.001) | .174 ( 0.048) | .005 ( 0.053) |
| **Substance Abuse ( 86%)** | **Highly related fields** | Raw citations | 719 ( 0.003) | 351 ( 0.062) | 10.1 ( 0.076) |
|  |  | Composite | 2.98 (<0.001) | .061 ( 0.251) | .00411 ( 0.011) |
| **Developmental Biology ( 86%)** | **Highly related fields** | Raw citations | 4258 (<0.001) | -45.7 ( 0.886) | -29.7 ( 0.002) |
|  |  | Composite | 2.99 (<0.001) | .0564 ( 0.017) | .00616 (<0.001) |
| **Endocrinology & Metabolism ( 83%)** | **Highly related fields** | Raw citations | 1543 (<0.001) | 363 ( 0.068) | 6.25 ( 0.297) |
|  |  | Composite | 2.98 (<0.001) | .0971 ( 0.003) | .00481 (<0.001) |
| **Immunology ( 83%)** | **Highly related fields** | Raw citations | 2095 (<0.001) | 10.3 ( 0.952) | 13.7 ( 0.011) |
|  |  | Composite | 2.96 (<0.001) | .0632 ( 0.014) | .0053 (<0.001) |
| **Neurology & Neurosurgery ( 82%)** | **Highly related fields** | Raw citations | 1765 (<0.001) | 478 (<0.001) | 1.27 ( 0.709) |
|  |  | Composite | 3 (<0.001) | .0889 (<0.001) | .00399 (<0.001) |
| **Biochemistry & Molecular Biology ( 81%)** | **Highly related fields** | Raw citations | 1317 (<0.001) | 346 (<0.001) | -5.45 ( 0.032) |
|  |  | Composite | 2.76 (<0.001) | .123 (<0.001) | .00408 (<0.001) |
| **Virology ( 81%)** | **Highly related fields** | Raw citations | 1723 (<0.001) | 120 ( 0.501) | -1.37 ( 0.809) |
|  |  | Composite | 2.75 (<0.001) | .0491 ( 0.094) | .00491 (<0.001) |
| **Psychiatry ( 81%)** | **Highly related fields** | Raw citations | 1030 ( 0.002) | 785 (<0.001) | 19 ( 0.009) |
|  |  | Composite | 3 (<0.001) | .0841 ( 0.008) | .00582 (<0.001) |
| **Genetics & Heredity ( 79%)** | **Highly related fields** | Raw citations | 2348 (<0.001) | 414 ( 0.115) | -16.2 ( 0.079) |
|  |  | Composite | 2.82 (<0.001) | .0564 ( 0.107) | .0018 ( 0.144) |
| **Allergy ( 75%)** | **Highly related fields** | Raw citations | 740 ( 0.052) | 606 ( 0.015) | 19.5 ( 0.030) |
|  |  | Composite | 2.87 (<0.001) | .145 ( 0.019) | .00609 ( 0.007) |
| **Biophysics ( 74%)** | **Highly related fields** | Raw citations | 689 ( 0.057) | 521 ( 0.040) | .219 ( 0.978) |
|  |  | Composite | 2.5 (<0.001) | .198 (<0.001) | .00546 (<0.001) |
| **Epidemiology ( 74%)** | **Highly related fields** | Raw citations | 1537 ( 0.105) | 217 ( 0.733) | 28.8 ( 0.207) |
|  |  | Composite | 3.15 (<0.001) | -.0773 ( 0.286) | .0041 ( 0.115) |
| **Biomedical Engineering ( 74%)** | **Highly related fields** | Raw citations | 501 ( 0.031) | 658 (<0.001) | 11 ( 0.078) |
|  |  | Composite | 2.64 (<0.001) | .121 (<0.001) | .00673 (<0.001) |
| **Arthritis & Rheumatology ( 73%)** | **Highly related fields** | Raw citations | 1440 ( 0.007) | 869 ( 0.010) | 10.2 ( 0.409) |
|  |  | Composite | 2.98 (<0.001) | .121 ( 0.021) | .00408 ( 0.033) |
| **Oncology & Carcinogenesis ( 72%)** | **Highly related fields** | Raw citations | 3185 (<0.001) | 410 ( 0.014) | -10.9 ( 0.088) |
|  |  | Composite | 2.91 (<0.001) | .0791 (<0.001) | .00381 (<0.001) |
| **Developmental & Child Psychology ( 72%)** | **Highly related fields** | Raw citations | 466 ( 0.005) | 572 (<0.001) | 14.1 (<0.001) |
|  |  | Composite | 3.02 (<0.001) | .0938 ( 0.005) | .0053 (<0.001) |
| **Medical Informatics ( 71%)** | **Highly related fields** | Raw citations | 750 ( 0.003) | 243 ( 0.217) | .0172 ( 0.998) |
|  |  | Composite | 2.71 (<0.001) | -.000266 ( 0.997) | .00417 ( 0.084) |
| **Physiology ( 71%)** | **Highly related fields** | Raw citations | 474 (<0.001) | 253 ( 0.001) | 4.07 ( 0.080) |
|  |  | Composite | 2.83 (<0.001) | .0537 ( 0.171) | .00379 ( 0.003) |
| **Public Health ( 71%)** | **Highly related fields** | Raw citations | 962 ( 0.003) | -94.5 ( 0.672) | 27.4 ( 0.002) |
|  |  | Composite | 3 (<0.001) | .0397 ( 0.117) | .00487 (<0.001) |
| **Gastroenterology & Hepatology ( 69%)** | **Highly related fields** | Raw citations | 1618 (<0.001) | 405 ( 0.012) | .268 ( 0.965) |
|  |  | Composite | 2.9 (<0.001) | .0949 ( 0.003) | .00355 ( 0.004) |
| **Urology & Nephrology ( 67%)** | **Highly related fields** | Raw citations | 1708 (<0.001) | 370 ( 0.009) | -7.9 ( 0.150) |
|  |  | Composite | 2.79 (<0.001) | .129 (<0.001) | .0028 ( 0.004) |
| **Respiratory System ( 67%)** | **Highly related fields** | Raw citations | 1285 (<0.001) | 455 ( 0.001) | 10.8 ( 0.059) |
|  |  | Composite | 2.82 (<0.001) | .0622 ( 0.015) | .00506 (<0.001) |
| **Pediatrics ( 67%)** | **Highly related fields** | Raw citations | 699 (<0.001) | 257 (<0.001) | 2.61 ( 0.233) |
|  |  | Composite | 2.58 (<0.001) | .0925 (<0.001) | .00346 (<0.001) |
| **Bioinformatics ( 67%)** | **Highly related fields** | Raw citations | 3520 (<0.001) | 765 ( 0.250) | -19.3 ( 0.496) |
|  |  | Composite | 3 (<0.001) | .0686 ( 0.252) | .00289 ( 0.259) |
| **Cardiovascular System & Hematology ( 66%)** | **Highly related fields** | Raw citations | 3008 (<0.001) | 267 ( 0.098) | -7.59 ( 0.199) |
|  |  | Composite | 2.95 (<0.001) | .0794 (<0.001) | .00337 (<0.001) |
| **Health Policy & Services ( 64%)** | **Highly related fields** | Raw citations | 685 ( 0.019) | 223 ( 0.223) | 17.2 ( 0.028) |
|  |  | Composite | 2.8 (<0.001) | .0999 ( 0.026) | .00665 ( 0.001) |
| **Experimental Psychology ( 64%)** | **Highly related fields** | Raw citations | 909 (<0.001) | 302 ( 0.001) | 2.58 ( 0.382) |
|  |  | Composite | 3.1 (<0.001) | .0719 ( 0.012) | .00368 (<0.001) |
| **Emergency & Critical Care Medicine ( 63%)** | **Highly related fields** | Raw citations | 714 ( 0.087) | 1077 (<0.001) | 16.9 ( 0.157) |
|  |  | Composite | 2.75 (<0.001) | .188 (<0.001) | .00366 ( 0.048) |
| **Clinical Psychology ( 63%)** | **Highly related fields** | Raw citations | 541 ( 0.045) | 723 (<0.001) | 16.9 ( 0.006) |
|  |  | Composite | 3.03 (<0.001) | .0951 ( 0.056) | .00482 ( 0.005) |
| **Ophthalmology & Optometry ( 62%)** | **Highly related fields** | Raw citations | 1209 (<0.001) | 359 (<0.001) | -5.75 ( 0.137) |
|  |  | Composite | 2.7 (<0.001) | .132 (<0.001) | .00307 ( 0.001) |
| **Family Studies ( 62%)** | **Other fields** | Raw citations | 215 ( 0.126) | 214 ( 0.022) | 6.25 ( 0.078) |
|  |  | Composite | 2.85 (<0.001) | .0286 ( 0.626) | .00579 ( 0.013) |
| **Speech-Language Pathology & Audiology ( 61%)** | **Highly related fields** | Raw citations | 1132 (<0.001) | -84.7 ( 0.615) | -9.25 ( 0.198) |
|  |  | Composite | 2.81 (<0.001) | -.000538 ( 0.991) | .00419 ( 0.044) |
| **Rehabilitation ( 61%)** | **Highly related fields** | Raw citations | 439 ( 0.003) | 159 ( 0.071) | 10 ( 0.013) |
|  |  | Composite | 2.7 (<0.001) | -.0105 ( 0.764) | .00662 (<0.001) |
| **Obstetrics & Reproductive Medicine ( 60%)** | **Highly related fields** | Raw citations | 817 (<0.001) | 412 (<0.001) | .233 ( 0.944) |
|  |  | Composite | 2.72 (<0.001) | .118 (<0.001) | .00254 ( 0.011) |
| **Toxicology ( 60%)** | **Highly related fields** | Raw citations | 1148 (<0.001) | 536 ( 0.001) | -5.14 ( 0.418) |
|  |  | Composite | 2.67 (<0.001) | .133 (<0.001) | .00463 (<0.001) |
| **Environmental & Occupational Health ( 59%)** | **Highly related fields** | Raw citations | 469 ( 0.208) | -167 ( 0.446) | 12.7 ( 0.216) |
|  |  | Composite | 2.53 (<0.001) | .0602 ( 0.230) | .00361 ( 0.125) |
| **Nutrition & Dietetics ( 59%)** | **Highly related fields** | Raw citations | 1066 (<0.001) | 350 ( 0.015) | 4.67 ( 0.393) |
|  |  | Composite | 2.95 (<0.001) | .0863 ( 0.016) | .00441 ( 0.001) |
| **Microbiology ( 59%)** | **Highly related fields** | Raw citations | 2613 (<0.001) | 161 ( 0.228) | -24.5 (<0.001) |
|  |  | Composite | 2.92 (<0.001) | .0543 ( 0.002) | .00369 (<0.001) |
| **Nursing ( 59%)** | **Highly related fields** | Raw citations | 129 ( 0.029) | 179 (<0.001) | 7 (<0.001) |
|  |  | Composite | 2.41 (<0.001) | .0303 ( 0.183) | .00467 (<0.001) |
| **Demography ( 58%)** | **Highly related fields** | Raw citations | 58.1 ( 0.858) | 192 ( 0.369) | 13.6 ( 0.094) |
|  |  | Composite | 2.8 (<0.001) | .0392 ( 0.691) | .00702 ( 0.065) |
| **Analytical Chemistry ( 58%)** | **Other fields** | Raw citations | 810 (<0.001) | 606 (<0.001) | -1.48 ( 0.732) |
|  |  | Composite | 2.71 (<0.001) | .0844 ( 0.011) | .00318 ( 0.006) |
| **Nuclear Medicine & Medical Imaging ( 57%)** | **Highly related fields** | Raw citations | 1004 (<0.001) | 324 (<0.001) | -1.93 ( 0.465) |
|  |  | Composite | 2.66 (<0.001) | .121 (<0.001) | .00171 ( 0.018) |
| **Organic Chemistry ( 56%)** | **Other fields** | Raw citations | 1316 (<0.001) | 179 ( 0.095) | -2.69 ( 0.415) |
|  |  | Composite | 2.82 (<0.001) | .112 (<0.001) | .00348 (<0.001) |
| **Pharmacology & Pharmacy ( 53%)** | **Highly related fields** | Raw citations | 623 (<0.001) | 339 (<0.001) | 1.09 ( 0.641) |
|  |  | Composite | 2.58 (<0.001) | .0988 (<0.001) | .00414 (<0.001) |
| **Medicinal & Biomolecular Chemistry ( 51%)** | **Highly related fields** | Raw citations | 517 (<0.001) | 337 ( 0.001) | 8.9 ( 0.019) |
|  |  | Composite | 2.44 (<0.001) | .12 (<0.001) | .00677 (<0.001) |
| **Complementary & Alternative Medicine ( 50%)** | **Highly related fields** | Raw citations | 242 ( 0.044) | 257 ( 0.001) | 1.81 ( 0.663) |
|  |  | Composite | 2.52 (<0.001) | .0584 ( 0.483) | .00107 ( 0.813) |
| **Behavioral Science & Comparative Psychology ( 49%)** | **Highly related fields** | Raw citations | 692 (<0.001) | 236 ( 0.012) | .129 ( 0.970) |
|  |  | Composite | 2.93 (<0.001) | .0766 ( 0.085) | .0045 ( 0.007) |
| **Anesthesiology ( 48%)** | **Highly related fields** | Raw citations | 885 (<0.001) | 256 ( 0.023) | -1.98 ( 0.684) |
|  |  | Composite | 2.6 (<0.001) | .135 (<0.001) | .00423 ( 0.002) |
| **Dentistry ( 48%)** | **Highly related fields** | Raw citations | 787 (<0.001) | 263 (<0.001) | -4.62 ( 0.061) |
|  |  | Composite | 2.69 (<0.001) | .0963 ( 0.001) | .00173 ( 0.107) |
| **Otorhinolaryngology ( 47%)** | **Highly related fields** | Raw citations | 893 (<0.001) | 27.6 ( 0.734) | -5.1 ( 0.105) |
|  |  | Composite | 2.6 (<0.001) | .0578 ( 0.011) | .00161 ( 0.066) |
| **Tropical Medicine ( 46%)** | **Highly related fields** | Raw citations | 1264 ( 0.001) | 460 ( 0.084) | -8.21 ( 0.381) |
|  |  | Composite | 2.56 (<0.001) | .106 ( 0.004) | .00463 (<0.001) |
| **Applied Ethics ( 46%)** | **Highly related fields** | Raw citations | 804 (<0.001) | 264 ( 0.051) | -5.64 ( 0.300) |
|  |  | Composite | 3.02 (<0.001) | .058 ( 0.410) | .000272 ( 0.924) |
| **General & Internal Medicine ( 46%)** | **Highly related fields** | Raw citations | 3316 (<0.001) | -197 ( 0.465) | -35.2 (<0.001) |
|  |  | Composite | 2.55 (<0.001) | .209 (<0.001) | .00181 ( 0.007) |
| **Statistics & Probability ( 45%)** | **Other fields** | Raw citations | 942 ( 0.026) | 606 ( 0.039) | 12.8 ( 0.170) |
|  |  | Composite | 3.07 (<0.001) | .0876 ( 0.068) | .00331 ( 0.030) |
| **Surgery ( 44%)** | **Highly related fields** | Raw citations | 943 (<0.001) | 395 (<0.001) | 2.83 ( 0.355) |
|  |  | Composite | 2.6 (<0.001) | .104 (<0.001) | .0032 (<0.001) |
| **Mycology & Parasitology ( 44%)** | **Highly related fields** | Raw citations | 1424 (<0.001) | -270 ( 0.104) | -8.83 ( 0.129) |
|  |  | Composite | 2.82 (<0.001) | -.0937 ( 0.121) | .00172 ( 0.414) |
| **Dermatology & Venereal Diseases ( 42%)** | **Highly related fields** | Raw citations | 1145 (<0.001) | 683 (<0.001) | -4.48 ( 0.478) |
|  |  | Composite | 2.73 (<0.001) | .161 (<0.001) | .00273 ( 0.046) |
| **History of Social Sciences ( 41%)** | **Other fields** | Raw citations | -31.4 ( 0.881) | -172 ( 0.189) | 10.1 ( 0.041) |
|  |  | Composite | 2.29 (<0.001) | -.0882 ( 0.519) | .0138 ( 0.012) |
| **Biotechnology ( 41%)** | **Highly related fields** | Raw citations | 692 ( 0.004) | 164 ( 0.359) | 20 ( 0.008) |
|  |  | Composite | 2.64 (<0.001) | .052 ( 0.260) | .0104 (<0.001) |
| **General Chemistry ( 40%)** | **Other fields** | Raw citations | 1304 (<0.001) | 385 ( 0.111) | -.0811 ( 0.986) |
|  |  | Composite | 2.59 (<0.001) | .171 (<0.001) | .00529 (<0.001) |
| **Sport Sciences ( 38%)** | **Highly related fields** | Raw citations | 784 ( 0.013) | 506 ( 0.016) | 10.4 ( 0.265) |
|  |  | Composite | 3.02 (<0.001) | .0572 ( 0.226) | .00204 ( 0.335) |
| **Social Psychology ( 38%)** | **Highly related fields** | Raw citations | 961 (<0.001) | 706 (<0.001) | 9.51 ( 0.047) |
|  |  | Composite | 3.14 (<0.001) | .178 (<0.001) | .00434 (<0.001) |
| **Acoustics ( 37%)** | **Other fields** | Raw citations | 963 (<0.001) | -5.23 ( 0.954) | -7.81 ( 0.005) |
|  |  | Composite | 2.84 (<0.001) | -.0267 ( 0.508) | .000611 ( 0.621) |
| **Optics ( 32%)** | **Other fields** | Raw citations | 1066 (<0.001) | 75.7 ( 0.669) | 13.1 ( 0.052) |
|  |  | Composite | 2.67 (<0.001) | .0451 ( 0.213) | .00779 (<0.001) |
| **Orthopedics ( 31%)** | **Highly related fields** | Raw citations | 1110 (<0.001) | 365 (<0.001) | -2.37 ( 0.388) |
|  |  | Composite | 2.79 (<0.001) | .116 (<0.001) | .00336 (<0.001) |
| **Chemical Physics ( 30%)** | **Other fields** | Raw citations | 1439 (<0.001) | 795 (<0.001) | 6.04 ( 0.223) |
|  |  | Composite | 3 (<0.001) | .11 (<0.001) | .00415 (<0.001) |
| **Evolutionary Biology ( 30%)** | **Other fields** | Raw citations | 960 ( 0.001) | 769 (<0.001) | 6.19 ( 0.419) |
|  |  | Composite | 2.98 (<0.001) | .0813 ( 0.016) | .00686 (<0.001) |
| **Nanoscience & Nanotechnology ( 29%)** | **Other fields** | Raw citations | 1728 (<0.001) | -28.7 ( 0.940) | 98.9 (<0.001) |
|  |  | Composite | 2.98 (<0.001) | .071 ( 0.030) | .0127 (<0.001) |
| **Sociology ( 28%)** | **Other fields** | Raw citations | 384 (<0.001) | 40.1 ( 0.513) | 6.23 ( 0.002) |
|  |  | Composite | 3.06 (<0.001) | -.0314 ( 0.427) | .00385 ( 0.003) |
| **Pathology ( 28%)** | **Highly related fields** | Raw citations | 1763 (<0.001) | 404 ( 0.056) | -7.32 ( 0.342) |
|  |  | Composite | 2.83 (<0.001) | .0621 ( 0.142) | .00202 ( 0.192) |
| **Plant Biology & Botany ( 27%)** | **Other fields** | Raw citations | 1194 (<0.001) | 418 (<0.001) | -3.61 ( 0.258) |
|  |  | Composite | 2.87 (<0.001) | .0835 ( 0.001) | .0041 (<0.001) |
| **General Clinical Medicine ( 27%)** | **Highly related fields** | Raw citations | 289 ( 0.034) | 465 (<0.001) | 1.84 ( 0.611) |
|  |  | Composite | 2.2 (<0.001) | .175 ( 0.015) | .00594 ( 0.006) |
| **Veterinary Sciences ( 27%)** | **Highly related fields** | Raw citations | 456 (<0.001) | 125 ( 0.001) | .87 ( 0.563) |
|  |  | Composite | 2.5 (<0.001) | .0669 ( 0.005) | .0029 ( 0.002) |
| **Environmental Sciences ( 26%)** | **Other fields** | Raw citations | 1664 (<0.001) | 665 ( 0.023) | -1.15 ( 0.909) |
|  |  | Composite | 3.07 (<0.001) | .101 ( 0.037) | .00295 ( 0.080) |
| **Social Sciences Methods ( 25%)** | **Other fields** | Raw citations | 487 ( 0.391) | 1982 (<0.001) | 11.3 ( 0.357) |
|  |  | Composite | 3.1 (<0.001) | .322 ( 0.001) | .00332 ( 0.113) |
| **Criminology ( 25%)** | **Other fields** | Raw citations | 379 (<0.001) | 311 (<0.001) | 8.31 ( 0.008) |
|  |  | Composite | 2.98 (<0.001) | .0943 ( 0.033) | .00485 ( 0.002) |
| **Microscopy ( 24%)** | **Highly related fields** | Raw citations | 656 ( 0.236) | 235 ( 0.577) | 9.4 ( 0.521) |
|  |  | Composite | 2.42 (<0.001) | .102 ( 0.497) | .0104 ( 0.053) |
| **Inorganic & Nuclear Chemistry ( 23%)** | **Other fields** | Raw citations | 934 (<0.001) | 32.1 ( 0.892) | 4.53 ( 0.433) |
|  |  | Composite | 2.75 (<0.001) | .111 ( 0.048) | .00287 ( 0.036) |
| **Social Work ( 23%)** | **Other fields** | Raw citations | 190 ( 0.035) | 321 (<0.001) | 5.37 ( 0.092) |
|  |  | Composite | 2.75 (<0.001) | .146 ( 0.006) | .00126 ( 0.522) |
| **Polymers ( 21%)** | **Other fields** | Raw citations | 1200 (<0.001) | 543 ( 0.001) | -1.22 ( 0.760) |
|  |  | Composite | 2.86 (<0.001) | .074 ( 0.081) | .00315 ( 0.004) |
| **Drama & Theater ( 20%)** | **Other fields** | Raw citations | 89.6 ( 0.160) | -18 ( 0.687) | -.776 ( 0.706) |
|  |  | Composite | 1.99 (<0.001) | .0288 ( 0.831) | .00431 ( 0.495) |
| **Economics ( 19%)** | **Other fields** | Raw citations | 739 (<0.001) | 216 ( 0.033) | 4.18 ( 0.143) |
|  |  | Composite | 3.08 (<0.001) | .0832 ( 0.028) | .00502 (<0.001) |
| **Food Science ( 19%)** | **Other fields** | Raw citations | 1061 ( 0.003) | 184 ( 0.578) | 3.59 ( 0.701) |
|  |  | Composite | 2.86 (<0.001) | .0976 ( 0.124) | .0054 ( 0.003) |
| **Artificial Intelligence & Image Processing ( 19%)** | **Other fields** | Raw citations | 1773 (<0.001) | 132 ( 0.499) | -4.51 ( 0.470) |
|  |  | Composite | 2.78 (<0.001) | .0109 ( 0.697) | .00714 (<0.001) |
| **Distributed Computing ( 18%)** | **Other fields** | Raw citations | 363 ( 0.266) | 663 ( 0.016) | 9.86 ( 0.374) |
|  |  | Composite | 2.37 (<0.001) | .052 ( 0.519) | .0042 ( 0.204) |
| **Industrial Engineering & Automation ( 17%)** | **Other fields** | Raw citations | 1046 (<0.001) | 272 ( 0.022) | -2.38 ( 0.489) |
|  |  | Composite | 2.87 (<0.001) | .0638 ( 0.133) | .00302 ( 0.015) |
| **Entomology ( 17%)** | **Other fields** | Raw citations | 962 (<0.001) | 392 (<0.001) | -8.84 (<0.001) |
|  |  | Composite | 2.91 (<0.001) | .218 (<0.001) | -.00174 ( 0.207) |
| **Design Practice & Management ( 16%)** | **Other fields** | Raw citations | 638 (<0.001) | 259 ( 0.137) | -.528 ( 0.899) |
|  |  | Composite | 2.79 (<0.001) | .0166 ( 0.876) | .00122 ( 0.635) |
| **Human Factors ( 16%)** | **Highly related fields** | Raw citations | 745 (<0.001) | 73.4 ( 0.647) | 6.59 ( 0.204) |
|  |  | Composite | 2.88 (<0.001) | .0226 ( 0.702) | .00743 (<0.001) |
| **Information & Library Sciences ( 16%)** | **Other fields** | Raw citations | 216 ( 0.004) | 253 ( 0.006) | 2.05 ( 0.373) |
|  |  | Composite | 2.6 (<0.001) | .185 ( 0.026) | .00199 ( 0.345) |
| **Fluids & Plasmas ( 16%)** | **Other fields** | Raw citations | 1646 (<0.001) | 89.2 ( 0.718) | -10.5 ( 0.098) |
|  |  | Composite | 3.12 (<0.001) | .0232 ( 0.662) | .001 ( 0.460) |
| **General Psychology & Cognitive Sciences ( 16%)** | **Highly related fields** | Raw citations | 660 (<0.001) | 87.6 ( 0.610) | .427 ( 0.927) |
|  |  | Composite | 3 (<0.001) | -.0246 ( 0.813) | .000894 ( 0.752) |
| **Urban & Regional Planning ( 15%)** | **Other fields** | Raw citations | 549 ( 0.001) | 82.1 ( 0.655) | 1.56 ( 0.748) |
|  |  | Composite | 3.12 (<0.001) | .121 ( 0.220) | .00247 ( 0.344) |
| **Education ( 15%)** | **Other fields** | Raw citations | 340 (<0.001) | 216 (<0.001) | 6.95 (<0.001) |
|  |  | Composite | 2.89 (<0.001) | .0149 ( 0.615) | .00417 (<0.001) |
| **Psychoanalysis ( 15%)** | **Highly related fields** | Raw citations | 82.3 ( 0.574) | 12.2 ( 0.926) | 2.19 ( 0.413) |
|  |  | Composite | 2.56 (<0.001) | -.0927 ( 0.607) | .000313 ( 0.931) |
| **Development Studies ( 15%)** | **Other fields** | Raw citations | 550 ( 0.024) | 129 ( 0.515) | 2.3 ( 0.719) |
|  |  | Composite | 3.14 (<0.001) | -.0142 ( 0.920) | .0018 ( 0.696) |
| **Anatomy & Morphology ( 14%)** | **Highly related fields** | Raw citations | 330 (<0.001) | 89.4 ( 0.379) | -1.81 ( 0.380) |
|  |  | Composite | 2.46 (<0.001) | -.0209 ( 0.841) | -.000409 ( 0.847) |
| **Geography ( 14%)** | **Other fields** | Raw citations | 300 ( 0.014) | 239 ( 0.066) | 7.74 ( 0.030) |
|  |  | Composite | 2.98 (<0.001) | .185 ( 0.013) | .00461 ( 0.023) |
| **Ornithology ( 14%)** | **Other fields** | Raw citations | 403 ( 0.003) | -2.79 ( 0.985) | .55 ( 0.850) |
|  |  | Composite | 2.57 (<0.001) | .075 ( 0.480) | .00349 ( 0.096) |
| **History of Science, Technology & Medicine ( 13%)** | **Other fields** | Raw citations | 48.4 ( 0.342) | -3.1 ( 0.954) | 1.18 ( 0.262) |
|  |  | Composite | 2.29 (<0.001) | -.0496 ( 0.811) | .00217 ( 0.587) |
| **Gender Studies ( 13%)** | **Highly related fields** | Raw citations | 417 ( 0.064) | -153 ( 0.468) | 2.06 ( 0.723) |
|  |  | Composite | 2.9 (<0.001) | -.0419 ( 0.769) | .00499 ( 0.225) |
| **Dairy & Animal Science ( 13%)** | **Other fields** | Raw citations | 738 (<0.001) | 136 ( 0.066) | -3.79 ( 0.042) |
|  |  | Composite | 2.63 (<0.001) | .0375 ( 0.430) | .00236 ( 0.050) |
| **Logistics & Transportation ( 13%)** | **Other fields** | Raw citations | 698 (<0.001) | -93.9 ( 0.615) | 6.69 ( 0.202) |
|  |  | Composite | 2.97 (<0.001) | -.121 ( 0.142) | .00342 ( 0.140) |
| **Economic Theory ( 13%)** | **Other fields** | Raw citations | 220 ( 0.217) | -137 ( 0.369) | 5.24 ( 0.228) |
|  |  | Composite | 2.77 (<0.001) | .0322 ( 0.815) | .00461 ( 0.246) |
| **Optoelectronics & Photonics ( 12%)** | **Other fields** | Raw citations | 487 (<0.001) | 85.4 ( 0.264) | 1.57 ( 0.395) |
|  |  | Composite | 2.32 (<0.001) | .0124 ( 0.746) | .0057 (<0.001) |
| **Software Engineering ( 12%)** | **Other fields** | Raw citations | 1091 (<0.001) | 42.9 ( 0.695) | -13.3 (<0.001) |
|  |  | Composite | 2.79 (<0.001) | -.0816 ( 0.153) | .00156 ( 0.380) |
| **Marine Biology & Hydrobiology ( 12%)** | **Other fields** | Raw citations | 1526 (<0.001) | 54.8 ( 0.739) | -8.98 ( 0.088) |
|  |  | Composite | 3.06 (<0.001) | -.0403 ( 0.429) | .00224 ( 0.168) |
| **Anthropology ( 12%)** | **Other fields** | Raw citations | 297 (<0.001) | 33 ( 0.660) | 4.7 ( 0.023) |
|  |  | Composite | 2.97 (<0.001) | -.0705 ( 0.293) | .00293 ( 0.109) |
| **Numerical & Computational Mathematics ( 12%)** | **Other fields** | Raw citations | 525 ( 0.008) | 421 ( 0.029) | 5.09 ( 0.281) |
|  |  | Composite | 3.1 (<0.001) | .0294 ( 0.752) | -.0000629 ( 0.978) |
| **Geological & Geomatics Engineering ( 11%)** | **Other fields** | Raw citations | 1364 (<0.001) | 185 ( 0.368) | -2.54 ( 0.640) |
|  |  | Composite | 2.97 (<0.001) | .072 ( 0.233) | .0049 ( 0.002) |
| **Electrical & Electronic Engineering ( 11%)** | **Other fields** | Raw citations | 399 (<0.001) | 223 ( 0.113) | 10.2 ( 0.003) |
|  |  | Composite | 2.45 (<0.001) | .0931 ( 0.125) | .00632 (<0.001) |
| **Physical Chemistry ( 10%)** | **Other fields** | Raw citations | 1703 (<0.001) | 1296 ( 0.007) | 2.67 ( 0.754) |
|  |  | Composite | 2.91 (<0.001) | .0543 ( 0.641) | .00678 ( 0.002) |
| **Econometrics ( 10%)** | **Other fields** | Raw citations | 582 ( 0.488) | 111 ( 0.889) | 27.4 ( 0.217) |
|  |  | Composite | 3.28 (<0.001) | -.171 ( 0.434) | .00635 ( 0.298) |
| **Computer Hardware & Architecture ( 10%)** | **Other fields** | Raw citations | 594 (<0.001) | 138 ( 0.248) | 2.65 ( 0.382) |
|  |  | Composite | 2.46 (<0.001) | .0834 ( 0.168) | .00469 ( 0.003) |
| **Communication & Media Studies ( 10%)** | **Other fields** | Raw citations | 113 ( 0.419) | -127 ( 0.548) | 20.9 (<0.001) |
|  |  | Composite | 3.04 (<0.001) | -.0157 ( 0.812) | .00428 ( 0.006) |
| **Environmental Engineering ( 10%)** | **Other fields** | Raw citations | 1011 (<0.001) | 825 (<0.001) | -1.98 ( 0.631) |
|  |  | Composite | 2.95 (<0.001) | .148 ( 0.006) | .00205 ( 0.109) |
| **Networking & Telecommunications ( 10%)** | **Other fields** | Raw citations | 882 (<0.001) | -92.8 ( 0.367) | -1.8 ( 0.371) |
|  |  | Composite | 2.62 (<0.001) | -.0311 ( 0.404) | .00377 (<0.001) |
| **Archaeology ( 9.9%)** | **Other fields** | Raw citations | 423 (<0.001) | -37.7 ( 0.788) | .935 ( 0.741) |
|  |  | Composite | 2.82 (<0.001) | -.0845 ( 0.326) | .0015 ( 0.384) |
| **Zoology ( 9.8%)** | **Other fields** | Raw citations | 444 ( 0.001) | -45 ( 0.729) | .508 ( 0.891) |
|  |  | Composite | 2.65 (<0.001) | -.0371 ( 0.664) | -.00103 ( 0.671) |
| **Religions & Theology ( 9.8%)** | **Other fields** | Raw citations | 139 ( 0.002) | 174 ( 0.013) | -.768 ( 0.593) |
|  |  | Composite | 2.33 (<0.001) | .177 ( 0.178) | .000108 ( 0.968) |
| **Chemical Engineering ( 9.6%)** | **Other fields** | Raw citations | 1492 (<0.001) | 508 ( 0.046) | -8.9 ( 0.054) |
|  |  | Composite | 2.98 (<0.001) | .12 ( 0.145) | .00205 ( 0.170) |
| **Ecology ( 9.4%)** | **Other fields** | Raw citations | 1294 (<0.001) | 14.6 ( 0.941) | 11.6 ( 0.025) |
|  |  | Composite | 3.07 (<0.001) | .0896 ( 0.024) | .00531 (<0.001) |
| **Materials ( 9.3%)** | **Other fields** | Raw citations | 1231 (<0.001) | 148 ( 0.407) | -1.59 ( 0.606) |
|  |  | Composite | 2.76 (<0.001) | .0408 ( 0.331) | .00499 (<0.001) |
| **General Physics ( 9.1%)** | **Other fields** | Raw citations | 1256 (<0.001) | 259 ( 0.178) | -6.79 ( 0.031) |
|  |  | Composite | 2.78 (<0.001) | .0217 ( 0.721) | .00334 ( 0.001) |
| **Oceanography ( 8.9%)** | **Other fields** | Raw citations | 1002 (<0.001) | 141 ( 0.341) | -5.64 ( 0.120) |
|  |  | Composite | 2.91 (<0.001) | .0801 ( 0.269) | .0016 ( 0.366) |
| **Building & Construction ( 8.8%)** | **Other fields** | Raw citations | 759 ( 0.001) | 304 ( 0.295) | 12.3 ( 0.074) |
|  |  | Composite | 2.93 (<0.001) | .0546 ( 0.601) | .00731 ( 0.004) |
| **Sport, Leisure & Tourism ( 8.7%)** | **Other fields** | Raw citations | 698 (<0.001) | -248 ( 0.172) | 5.28 ( 0.303) |
|  |  | Composite | 3.08 (<0.001) | -.0635 ( 0.481) | .00458 ( 0.075) |
| **Applied Mathematics ( 8.7%)** | **Other fields** | Raw citations | 953 ( 0.016) | 1278 ( 0.009) | 6.49 ( 0.488) |
|  |  | Composite | 3.05 (<0.001) | .262 ( 0.052) | .0046 ( 0.078) |
| **Mechanical Engineering & Transports ( 8.6%)** | **Other fields** | Raw citations | 984 (<0.001) | 394 ( 0.026) | -3.76 ( 0.259) |
|  |  | Composite | 2.83 (<0.001) | -.0361 ( 0.566) | .00344 ( 0.004) |
| **Strategic, Defence & Security Studies ( 8.5%)** | **Other fields** | Raw citations | 240 ( 0.113) | 137 ( 0.509) | 10.7 ( 0.017) |
|  |  | Composite | 2.7 (<0.001) | .0471 ( 0.640) | .00767 ( 0.001) |
| **Languages & Linguistics ( 8.1%)** | **Other fields** | Raw citations | 408 (<0.001) | 163 ( 0.115) | .872 ( 0.713) |
|  |  | Composite | 3.02 (<0.001) | .222 ( 0.022) | .00292 ( 0.190) |
| **Computation Theory & Mathematics ( 8%)** | **Other fields** | Raw citations | 695 (<0.001) | -7.46 ( 0.970) | -.302 ( 0.940) |
|  |  | Composite | 2.93 (<0.001) | .0198 ( 0.814) | .00231 ( 0.176) |
| **Marketing ( 7.9%)** | **Other fields** | Raw citations | 191 ( 0.388) | 50.4 ( 0.840) | 27.5 (<0.001) |
|  |  | Composite | 3.01 (<0.001) | .108 ( 0.133) | .00564 ( 0.005) |
| **Law ( 7.6%)** | **Other fields** | Raw citations | 72.5 ( 0.722) | -104 ( 0.609) | 6.67 ( 0.369) |
|  |  | Composite | 2.45 (<0.001) | -.0012 ( 0.992) | .00448 ( 0.278) |
| **Meteorology & Atmospheric Sciences ( 7.4%)** | **Other fields** | Raw citations | 1768 (<0.001) | 108 ( 0.512) | 1.25 ( 0.730) |
|  |  | Composite | 3.09 (<0.001) | .0183 ( 0.595) | .00334 (<0.001) |
| **Applied Physics ( 7.3%)** | **Other fields** | Raw citations | 1446 (<0.001) | 246 ( 0.160) | -2.46 ( 0.354) |
|  |  | Composite | 2.84 (<0.001) | .0402 ( 0.268) | .00261 (<0.001) |
| **Operations Research ( 7.2%)** | **Other fields** | Raw citations | 553 ( 0.032) | -250 ( 0.446) | 12.4 ( 0.062) |
|  |  | Composite | 3.1 (<0.001) | -.0278 ( 0.771) | .00308 ( 0.112) |
| **Finance ( 7.2%)** | **Other fields** | Raw citations | 582 ( 0.003) | -373 ( 0.151) | 14.3 ( 0.016) |
|  |  | Composite | 3.01 (<0.001) | -.195 ( 0.038) | .00732 ( 0.001) |
| **Political Science & Public Administration ( 7%)** | **Other fields** | Raw citations | 465 (<0.001) | 471 ( 0.002) | 5.7 ( 0.059) |
|  |  | Composite | 3.05 (<0.001) | .11 ( 0.088) | .00392 ( 0.002) |
| **Forestry ( 6.9%)** | **Other fields** | Raw citations | 651 (<0.001) | 65.8 ( 0.626) | -.962 ( 0.777) |
|  |  | Composite | 2.62 (<0.001) | .0768 ( 0.368) | .00343 ( 0.113) |
| **Business & Management ( 6.6%)** | **Other fields** | Raw citations | 762 (<0.001) | -25.2 ( 0.885) | 12.4 ( 0.001) |
|  |  | Composite | 3.11 (<0.001) | .00783 ( 0.872) | .00466 (<0.001) |
| **Literary Studies ( 6.5%)** | **Other fields** | Raw citations | 33.6 ( 0.370) | -20.5 ( 0.688) | 1.9 ( 0.156) |
|  |  | Composite | 1.94 (<0.001) | -.00612 ( 0.962) | .00787 ( 0.021) |
| **General Mathematics ( 6.1%)** | **Other fields** | Raw citations | 442 (<0.001) | 190 ( 0.019) | -.916 ( 0.398) |
|  |  | Composite | 2.9 (<0.001) | -.00709 ( 0.899) | .000617 ( 0.411) |
| **Aerospace & Aeronautics ( 6.1%)** | **Other fields** | Raw citations | 398 (<0.001) | 82.2 ( 0.179) | -.0663 ( 0.952) |
|  |  | Composite | 2.44 (<0.001) | -.0296 ( 0.581) | .00446 (<0.001) |
| **Geology ( 6.1%)** | **Other fields** | Raw citations | 908 (<0.001) | -69.5 ( 0.730) | -6.24 ( 0.131) |
|  |  | Composite | 3.02 (<0.001) | -.0697 ( 0.599) | -.000583 ( 0.829) |
| **History ( 5.9%)** | **Other fields** | Raw citations | 71.9 ( 0.028) | 11.9 ( 0.837) | .286 ( 0.738) |
|  |  | Composite | 2.11 (<0.001) | .082 ( 0.433) | .00162 ( 0.300) |
| **Information Systems ( 5.9%)** | **Other fields** | Raw citations | 1214 (<0.001) | -63.9 ( 0.882) | -1.4 ( 0.888) |
|  |  | Composite | 3.17 (<0.001) | -.0083 ( 0.942) | .000141 ( 0.957) |
| **Energy ( 5.9%)** | **Other fields** | Raw citations | 1018 (<0.001) | 138 ( 0.367) | .526 ( 0.830) |
|  |  | Composite | 2.85 (<0.001) | .0144 ( 0.758) | .00394 (<0.001) |
| **Science Studies ( 5.7%)** | **Other fields** | Raw citations | 244 ( 0.310) | -101 ( 0.767) | 12.1 ( 0.066) |
|  |  | Composite | 3.03 (<0.001) | -.185 ( 0.456) | .00637 ( 0.174) |
| **Legal & Forensic Medicine ( 5.7%)** | **Highly related fields** | Raw citations | 272 ( 0.004) | -22.3 ( 0.883) | .685 ( 0.828) |
|  |  | Composite | 2.22 (<0.001) | .0124 ( 0.934) | .00552 ( 0.082) |
| **Cultural Studies ( 5.6%)** | **Other fields** | Raw citations | 78.7 ( 0.289) | -88.9 ( 0.450) | 2.51 ( 0.253) |
|  |  | Composite | 2.44 (<0.001) | -.193 ( 0.338) | .00556 ( 0.141) |
| **Astronomy & Astrophysics ( 5.5%)** | **Other fields** | Raw citations | 3131 (<0.001) | -293 ( 0.433) | -23.5 ( 0.001) |
|  |  | Composite | 3.17 (<0.001) | -.00394 ( 0.929) | .000616 ( 0.441) |
| **Accounting ( 5.5%)** | **Other fields** | Raw citations | 427 ( 0.023) | 176 ( 0.473) | 13 ( 0.024) |
|  |  | Composite | 2.92 (<0.001) | .042 ( 0.766) | .00809 ( 0.016) |
| **Music ( 5.4%)** | **Other fields** | Raw citations | 54.8 ( 0.008) | -18.7 ( 0.557) | .573 ( 0.492) |
|  |  | Composite | 2.17 (<0.001) | .0175 ( 0.906) | -.00137 ( 0.724) |
| **Geochemistry & Geophysics ( 5.4%)** | **Other fields** | Raw citations | 1303 (<0.001) | -52 ( 0.678) | -4.8 ( 0.042) |
|  |  | Composite | 3.15 (<0.001) | -.0317 ( 0.500) | .00175 ( 0.049) |
| **Agronomy & Agriculture ( 5.3%)** | **Other fields** | Raw citations | 1178 (<0.001) | 106 ( 0.670) | -2.93 ( 0.475) |
|  |  | Composite | 2.88 (<0.001) | .0747 ( 0.325) | .00287 ( 0.022) |
| **Nuclear & Particle Physics ( 4.9%)** | **Other fields** | Raw citations | 3823 (<0.001) | -419 ( 0.433) | -40.4 (<0.001) |
|  |  | Composite | 2.89 (<0.001) | .0439 ( 0.390) | .00253 (<0.001) |
| **Fisheries ( 4.8%)** | **Other fields** | Raw citations | 710 (<0.001) | -75.4 ( 0.658) | 1.11 ( 0.751) |
|  |  | Composite | 2.77 (<0.001) | -.0803 ( 0.408) | .00303 ( 0.131) |
| **Classics ( 4.8%)** | **Other fields** | Raw citations | 40.1 ( 0.022) | 17 ( 0.536) | .248 ( 0.593) |
|  |  | Composite | 2.04 (<0.001) | .216 ( 0.279) | .00159 ( 0.633) |
| **Mining & Metallurgy ( 4.7%)** | **Other fields** | Raw citations | 223 ( 0.141) | -133 ( 0.605) | 4.36 ( 0.253) |
|  |  | Composite | 2.3 (<0.001) | .0316 ( 0.895) | .006 ( 0.094) |
| **Paleontology ( 4.4%)** | **Other fields** | Raw citations | 1174 (<0.001) | -209 ( 0.609) | -4.07 ( 0.558) |
|  |  | Composite | 3.13 (<0.001) | -.0634 ( 0.488) | .000257 ( 0.869) |
| **Agricultural Economics & Policy ( 4.3%)** | **Other fields** | Raw citations | 976 (<0.001) | 200 ( 0.586) | -2.55 ( 0.713) |
|  |  | Composite | 3.24 (<0.001) | -.00151 ( 0.993) | -.00163 ( 0.634) |
| **Philosophy ( 3.9%)** | **Other fields** | Raw citations | 232 (<0.001) | 177 ( 0.026) | .676 ( 0.568) |
|  |  | Composite | 2.89 (<0.001) | .288 ( 0.012) | .00113 ( 0.504) |
| **International Relations ( 3.6%)** | **Other fields** | Raw citations | 118 ( 0.154) | -153 ( 0.350) | 9.87 (<0.001) |
|  |  | Composite | 2.82 (<0.001) | -.276 ( 0.112) | .00882 ( 0.002) |
| **Civil Engineering ( 2.9%)** | **Other fields** | Raw citations | 536 (<0.001) | -9.04 ( 0.975) | 6.69 ( 0.071) |
|  |  | Composite | 2.77 (<0.001) | -.0232 ( 0.872) | .00494 ( 0.008) |
| **Mathematical Physics ( 0%)** | **Other fields** | Raw citations |  |  |  |
|  |  | Composite |  |  |  |
| **Industrial Relations ( 0%)** | **Other fields** | Raw citations |  |  |  |
|  |  | Composite |  |  |  |
| **Horticulture ( 0%)** | **Other fields** | Raw citations |  |  |  |
|  |  | Composite |  |  |  |
| **Folklore ( 0%)** | **Other fields** | Raw citations |  |  |  |
|  |  | Composite |  |  |  |
| **Automobile Design & Engineering ( 0%)** | **Other fields** | Raw citations |  |  |  |
|  |  | Composite |  |  |  |
| **Art Practice, History & Theory ( 0%)** | **Other fields** | Raw citations |  |  |  |
|  |  | Composite |  |  |  |
| **Architecture ( 0%)** | **Other fields** | Raw citations |  |  |  |
|  |  | Composite |  |  |  |

**Supplementary Table 3.5 : Recent year impact, Funding time recent funding Linear Regressions for each subfield (ordered by percentage funded)**

| **Top-cited US-based researchers: Subfield (perc. funded)** | **Classification** | **Dependent Variable** | **Constant (p-val)** | **Funded (p-val)** | **Years since first pub (p-val)** |
| --- | --- | --- | --- | --- | --- |
| **Geriatrics ( 55%)** | **Highly related fields** | Raw citations | 1138 ( 0.217) | 421 ( 0.390) | 18.2 ( 0.404) |
|  |  | Composite | 2.91 (<0.001) | .125 ( 0.160) | .00672 ( 0.091) |
| **Substance Abuse ( 50%)** | **Highly related fields** | Raw citations | 758 ( 0.001) | 270 ( 0.045) | 14 ( 0.017) |
|  |  | Composite | 3.03 (<0.001) | -.00696 ( 0.855) | .00419 ( 0.012) |
| **Developmental Biology ( 49%)** | **Highly related fields** | Raw citations | 4063 (<0.001) | 213 ( 0.346) | -28.2 ( 0.004) |
|  |  | Composite | 3 (<0.001) | .0355 ( 0.034) | .00674 (<0.001) |
| **Medical Informatics ( 48%)** | **Highly related fields** | Raw citations | 749 ( 0.003) | 220 ( 0.226) | 2.61 ( 0.728) |
|  |  | Composite | 2.69 (<0.001) | .0314 ( 0.594) | .00444 ( 0.071) |
| **Bioinformatics ( 46%)** | **Highly related fields** | Raw citations | 3212 ( 0.001) | 1178 ( 0.063) | -9.15 ( 0.748) |
|  |  | Composite | 2.99 (<0.001) | .0779 ( 0.173) | .00361 ( 0.163) |
| **Virology ( 45%)** | **Highly related fields** | Raw citations | 1694 (<0.001) | 160 ( 0.264) | .291 ( 0.960) |
|  |  | Composite | 2.76 (<0.001) | .0343 ( 0.146) | .00534 (<0.001) |
| **Immunology ( 45%)** | **Highly related fields** | Raw citations | 2053 (<0.001) | 69.6 ( 0.586) | 14.3 ( 0.008) |
|  |  | Composite | 2.97 (<0.001) | .0445 ( 0.021) | .00604 (<0.001) |
| **Gerontology ( 44%)** | **Highly related fields** | Raw citations | 153 ( 0.889) | 1644 ( 0.012) | 31.9 ( 0.250) |
|  |  | Composite | 2.96 (<0.001) | .0662 ( 0.295) | .00593 ( 0.030) |
| **Biomedical Engineering ( 44%)** | **Highly related fields** | Raw citations | 579 ( 0.011) | 531 ( 0.001) | 17 ( 0.008) |
|  |  | Composite | 2.66 (<0.001) | .0817 ( 0.005) | .00769 (<0.001) |
| **Neurology & Neurosurgery ( 42%)** | **Highly related fields** | Raw citations | 1995 (<0.001) | 198 ( 0.027) | 3.48 ( 0.325) |
|  |  | Composite | 3.06 (<0.001) | .0074 ( 0.581) | .00411 (<0.001) |
| **Oncology & Carcinogenesis ( 40%)** | **Highly related fields** | Raw citations | 3170 (<0.001) | 335 ( 0.030) | -5.15 ( 0.425) |
|  |  | Composite | 2.92 (<0.001) | .044 ( 0.001) | .00473 (<0.001) |
| **Public Health ( 38%)** | **Highly related fields** | Raw citations | 984 ( 0.002) | -124 ( 0.562) | 26 ( 0.004) |
|  |  | Composite | 3.02 (<0.001) | .0108 ( 0.658) | .00504 (<0.001) |
| **Genetics & Heredity ( 38%)** | **Highly related fields** | Raw citations | 2182 (<0.001) | 636 ( 0.005) | -9.39 ( 0.318) |
|  |  | Composite | 2.81 (<0.001) | .076 ( 0.012) | .00263 ( 0.037) |
| **Emergency & Critical Care Medicine ( 38%)** | **Highly related fields** | Raw citations | 721 ( 0.088) | 1059 (<0.001) | 26.2 ( 0.035) |
|  |  | Composite | 2.78 (<0.001) | .147 ( 0.001) | .00482 ( 0.014) |
| **Epidemiology ( 36%)** | **Highly related fields** | Raw citations | 1546 ( 0.090) | 237 ( 0.686) | 30.6 ( 0.184) |
|  |  | Composite | 3.09 (<0.001) | .00261 ( 0.969) | .00395 ( 0.134) |
| **Psychiatry ( 36%)** | **Highly related fields** | Raw citations | 1237 (<0.001) | 610 ( 0.002) | 24.9 ( 0.001) |
|  |  | Composite | 3.03 (<0.001) | .0527 ( 0.054) | .0063 (<0.001) |
| **Gastroenterology & Hepatology ( 35%)** | **Highly related fields** | Raw citations | 1700 (<0.001) | 259 ( 0.104) | 3.67 ( 0.560) |
|  |  | Composite | 2.91 (<0.001) | .0798 ( 0.012) | .00449 (<0.001) |
| **Analytical Chemistry ( 35%)** | **Other fields** | Raw citations | 901 (<0.001) | 476 ( 0.001) | 1.37 ( 0.765) |
|  |  | Composite | 2.72 (<0.001) | .0665 ( 0.064) | .00358 ( 0.003) |
| **Arthritis & Rheumatology ( 34%)** | **Highly related fields** | Raw citations | 2017 (<0.001) | 176 ( 0.610) | 10.2 ( 0.451) |
|  |  | Composite | 3.06 (<0.001) | .0323 ( 0.541) | .0042 ( 0.044) |
| **Allergy ( 34%)** | **Highly related fields** | Raw citations | 803 ( 0.028) | 600 ( 0.013) | 25.2 ( 0.008) |
|  |  | Composite | 2.93 (<0.001) | .0852 ( 0.163) | .00665 ( 0.006) |
| **Developmental & Child Psychology ( 33%)** | **Highly related fields** | Raw citations | 561 (<0.001) | 546 (<0.001) | 18 (<0.001) |
|  |  | Composite | 3.04 (<0.001) | .0724 ( 0.025) | .00582 (<0.001) |
| **Endocrinology & Metabolism ( 33%)** | **Highly related fields** | Raw citations | 1785 (<0.001) | 49.4 ( 0.764) | 7.44 ( 0.234) |
|  |  | Composite | 3.06 (<0.001) | -.00906 ( 0.738) | .00488 (<0.001) |
| **Respiratory System ( 33%)** | **Highly related fields** | Raw citations | 1545 (<0.001) | 110 ( 0.443) | 11.1 ( 0.063) |
|  |  | Composite | 2.86 (<0.001) | .00952 ( 0.720) | .00504 (<0.001) |
| **Nuclear Medicine & Medical Imaging ( 32%)** | **Highly related fields** | Raw citations | 968 (<0.001) | 332 (<0.001) | 1.75 ( 0.520) |
|  |  | Composite | 2.65 (<0.001) | .118 (<0.001) | .00304 (<0.001) |
| **Cardiovascular System & Hematology ( 32%)** | **Highly related fields** | Raw citations | 2767 (<0.001) | 642 (<0.001) | -1.31 ( 0.830) |
|  |  | Composite | 2.95 (<0.001) | .0824 (<0.001) | .00418 (<0.001) |
| **Health Policy & Services ( 31%)** | **Highly related fields** | Raw citations | 749 ( 0.008) | 177 ( 0.354) | 17.9 ( 0.023) |
|  |  | Composite | 2.82 (<0.001) | .1 ( 0.032) | .00711 (<0.001) |
| **Microbiology ( 31%)** | **Highly related fields** | Raw citations | 2694 (<0.001) | 8.58 ( 0.952) | -24.2 (<0.001) |
|  |  | Composite | 2.94 (<0.001) | .0275 ( 0.150) | .00397 (<0.001) |
| **Toxicology ( 31%)** | **Highly related fields** | Raw citations | 1040 (<0.001) | 729 (<0.001) | .942 ( 0.885) |
|  |  | Composite | 2.69 (<0.001) | .121 ( 0.001) | .00558 (<0.001) |
| **Pediatrics ( 31%)** | **Highly related fields** | Raw citations | 704 (<0.001) | 294 (<0.001) | 4.8 ( 0.033) |
|  |  | Composite | 2.6 (<0.001) | .0875 (<0.001) | .00409 (<0.001) |
| **Biophysics ( 31%)** | **Highly related fields** | Raw citations | 1228 ( 0.001) | -204 ( 0.443) | -2.28 ( 0.796) |
|  |  | Composite | 2.57 (<0.001) | .0794 ( 0.128) | .00663 (<0.001) |
| **Rehabilitation ( 30%)** | **Highly related fields** | Raw citations | 428 ( 0.004) | 197 ( 0.038) | 11.5 ( 0.005) |
|  |  | Composite | 2.7 (<0.001) | -.0139 ( 0.714) | .00651 (<0.001) |
| **Urology & Nephrology ( 30%)** | **Highly related fields** | Raw citations | 1878 (<0.001) | 99.8 ( 0.505) | -6.43 ( 0.257) |
|  |  | Composite | 2.81 (<0.001) | .106 (<0.001) | .00398 (<0.001) |
| **Ophthalmology & Optometry ( 30%)** | **Highly related fields** | Raw citations | 1186 (<0.001) | 409 (<0.001) | -2.45 ( 0.537) |
|  |  | Composite | 2.69 (<0.001) | .154 (<0.001) | .00432 (<0.001) |
| **Physiology ( 29%)** | **Highly related fields** | Raw citations | 603 (<0.001) | 136 ( 0.071) | 4.35 ( 0.072) |
|  |  | Composite | 2.86 (<0.001) | .0303 ( 0.448) | .00386 ( 0.003) |
| **Biochemistry & Molecular Biology ( 29%)** | **Highly related fields** | Raw citations | 1535 (<0.001) | 113 ( 0.149) | -4.76 ( 0.071) |
|  |  | Composite | 2.85 (<0.001) | .0271 ( 0.140) | .00422 (<0.001) |
| **Clinical Psychology ( 28%)** | **Highly related fields** | Raw citations | 890 (<0.001) | 471 ( 0.018) | 16.1 ( 0.011) |
|  |  | Composite | 3.08 (<0.001) | .0664 ( 0.219) | .00474 ( 0.006) |
| **Medicinal & Biomolecular Chemistry ( 28%)** | **Highly related fields** | Raw citations | 588 (<0.001) | 190 ( 0.093) | 10.4 ( 0.007) |
|  |  | Composite | 2.47 (<0.001) | .0491 ( 0.174) | .0072 (<0.001) |
| **Obstetrics & Reproductive Medicine ( 28%)** | **Highly related fields** | Raw citations | 917 (<0.001) | 202 ( 0.035) | 2.97 ( 0.395) |
|  |  | Composite | 2.75 (<0.001) | .0575 ( 0.045) | .00332 ( 0.002) |
| **Anesthesiology ( 27%)** | **Highly related fields** | Raw citations | 891 (<0.001) | 244 ( 0.061) | -.264 ( 0.958) |
|  |  | Composite | 2.61 (<0.001) | .11 ( 0.003) | .00495 (<0.001) |
| **Tropical Medicine ( 27%)** | **Highly related fields** | Raw citations | 1428 (<0.001) | 145 ( 0.639) | -7.89 ( 0.416) |
|  |  | Composite | 2.59 (<0.001) | .0525 ( 0.224) | .00484 (<0.001) |
| **Pharmacology & Pharmacy ( 25%)** | **Highly related fields** | Raw citations | 605 (<0.001) | 358 (<0.001) | 4.33 ( 0.064) |
|  |  | Composite | 2.58 (<0.001) | .106 (<0.001) | .0051 (<0.001) |
| **Nutrition & Dietetics ( 25%)** | **Highly related fields** | Raw citations | 995 (<0.001) | 502 ( 0.003) | 9 ( 0.107) |
|  |  | Composite | 2.95 (<0.001) | .0784 ( 0.060) | .00515 (<0.001) |
| **Nursing ( 25%)** | **Highly related fields** | Raw citations | 140 ( 0.015) | 226 (<0.001) | 8.23 (<0.001) |
|  |  | Composite | 2.39 (<0.001) | .0803 ( 0.002) | .00504 (<0.001) |
| **Demography ( 25%)** | **Highly related fields** | Raw citations | -165 ( 0.590) | 609 ( 0.009) | 18.5 ( 0.016) |
|  |  | Composite | 2.74 (<0.001) | .148 ( 0.184) | .00817 ( 0.030) |
| **Speech-Language Pathology & Audiology ( 25%)** | **Highly related fields** | Raw citations | 1059 (<0.001) | 28 ( 0.886) | -8.84 ( 0.232) |
|  |  | Composite | 2.81 (<0.001) | .00573 ( 0.919) | .00424 ( 0.047) |
| **Statistics & Probability ( 24%)** | **Other fields** | Raw citations | 942 ( 0.021) | 843 ( 0.015) | 14.7 ( 0.120) |
|  |  | Composite | 3.09 (<0.001) | .0833 ( 0.141) | .0033 ( 0.034) |
| **Family Studies ( 24%)** | **Other fields** | Raw citations | 245 ( 0.052) | 325 ( 0.002) | 7 ( 0.042) |
|  |  | Composite | 2.81 (<0.001) | .161 ( 0.014) | .00634 ( 0.005) |
| **Environmental & Occupational Health ( 23%)** | **Highly related fields** | Raw citations | 462 ( 0.218) | -153 ( 0.547) | 11.1 ( 0.280) |
|  |  | Composite | 2.56 (<0.001) | -.0146 ( 0.802) | .00391 ( 0.098) |
| **Experimental Psychology ( 22%)** | **Highly related fields** | Raw citations | 980 (<0.001) | 330 ( 0.003) | 3.88 ( 0.197) |
|  |  | Composite | 3.12 (<0.001) | .0657 ( 0.051) | .00392 (<0.001) |
| **Applied Ethics ( 22%)** | **Highly related fields** | Raw citations | 798 (<0.001) | 292 ( 0.069) | -3.71 ( 0.489) |
|  |  | Composite | 3.02 (<0.001) | .0736 ( 0.375) | .000696 ( 0.804) |
| **Organic Chemistry ( 22%)** | **Other fields** | Raw citations | 1298 (<0.001) | 324 ( 0.012) | -1.26 ( 0.702) |
|  |  | Composite | 2.85 (<0.001) | .0937 ( 0.006) | .00407 (<0.001) |
| **Optics ( 22%)** | **Other fields** | Raw citations | 1038 (<0.001) | 181 ( 0.367) | 13.6 ( 0.045) |
|  |  | Composite | 2.67 (<0.001) | .0303 ( 0.461) | .0078 (<0.001) |
| **Microscopy ( 21%)** | **Highly related fields** | Raw citations | 636 ( 0.253) | 260 ( 0.554) | 10 ( 0.491) |
|  |  | Composite | 2.42 (<0.001) | .0882 ( 0.574) | .0107 ( 0.047) |
| **General & Internal Medicine ( 21%)** | **Highly related fields** | Raw citations | 3261 (<0.001) | -114 ( 0.736) | -35.6 (<0.001) |
|  |  | Composite | 2.58 (<0.001) | .176 (<0.001) | .00249 (<0.001) |
| **Biotechnology ( 21%)** | **Highly related fields** | Raw citations | 715 ( 0.003) | 60 ( 0.782) | 21.1 ( 0.005) |
|  |  | Composite | 2.63 (<0.001) | .0562 ( 0.316) | .0109 (<0.001) |
| **Otorhinolaryngology ( 20%)** | **Highly related fields** | Raw citations | 901 (<0.001) | .501 ( 0.996) | -4.95 ( 0.116) |
|  |  | Composite | 2.6 (<0.001) | .0495 ( 0.080) | .00214 ( 0.015) |
| **Surgery ( 20%)** | **Highly related fields** | Raw citations | 928 (<0.001) | 478 (<0.001) | 5.87 ( 0.058) |
|  |  | Composite | 2.6 (<0.001) | .0936 (<0.001) | .00384 (<0.001) |
| **General Chemistry ( 19%)** | **Other fields** | Raw citations | 1459 (<0.001) | -43.1 ( 0.889) | .159 ( 0.973) |
|  |  | Composite | 2.62 (<0.001) | .126 ( 0.029) | .00576 (<0.001) |
| **Mycology & Parasitology ( 18%)** | **Highly related fields** | Raw citations | 1377 (<0.001) | -247 ( 0.255) | -9.55 ( 0.107) |
|  |  | Composite | 2.78 (<0.001) | -.00229 ( 0.977) | .00178 ( 0.408) |
| **Nanoscience & Nanotechnology ( 18%)** | **Other fields** | Raw citations | 1719 (<0.001) | 29.8 ( 0.947) | 98.7 (<0.001) |
|  |  | Composite | 2.98 (<0.001) | .0545 ( 0.157) | .0131 (<0.001) |
| **Behavioral Science & Comparative Psychology ( 17%)** | **Highly related fields** | Raw citations | 715 (<0.001) | 204 ( 0.109) | 1.51 ( 0.671) |
|  |  | Composite | 2.94 (<0.001) | .0557 ( 0.353) | .0049 ( 0.004) |
| **Dermatology & Venereal Diseases ( 16%)** | **Highly related fields** | Raw citations | 1168 (<0.001) | 708 ( 0.002) | .0314 ( 0.996) |
|  |  | Composite | 2.76 (<0.001) | .0719 ( 0.146) | .00366 ( 0.009) |
| **Acoustics ( 16%)** | **Other fields** | Raw citations | 958 (<0.001) | 5.66 ( 0.963) | -7.77 ( 0.007) |
|  |  | Composite | 2.85 (<0.001) | -.0544 ( 0.321) | .000337 ( 0.791) |
| **Orthopedics ( 15%)** | **Highly related fields** | Raw citations | 1128 (<0.001) | 313 ( 0.002) | -.821 ( 0.768) |
|  |  | Composite | 2.79 (<0.001) | .114 (<0.001) | .0039 (<0.001) |
| **Environmental Sciences ( 15%)** | **Other fields** | Raw citations | 1593 (<0.001) | 923 ( 0.010) | 2.43 ( 0.809) |
|  |  | Composite | 3.06 (<0.001) | .113 ( 0.057) | .00346 ( 0.040) |
| **Sociology ( 13%)** | **Other fields** | Raw citations | 385 (<0.001) | 57 ( 0.483) | 6.31 ( 0.002) |
|  |  | Composite | 3.05 (<0.001) | .00144 ( 0.978) | .00394 ( 0.003) |
| **Complementary & Alternative Medicine ( 12%)** | **Highly related fields** | Raw citations | 298 ( 0.019) | 284 ( 0.024) | 3.2 ( 0.461) |
|  |  | Composite | 2.54 (<0.001) | .195 ( 0.120) | .000627 ( 0.887) |
| **Dentistry ( 12%)** | **Highly related fields** | Raw citations | 796 (<0.001) | 391 (<0.001) | -2.76 ( 0.271) |
|  |  | Composite | 2.69 (<0.001) | .15 ( 0.001) | .00244 ( 0.025) |
| **Distributed Computing ( 12%)** | **Other fields** | Raw citations | 314 ( 0.343) | 627 ( 0.056) | 13.4 ( 0.228) |
|  |  | Composite | 2.38 (<0.001) | -.0301 ( 0.754) | .00461 ( 0.160) |
| **Pathology ( 11%)** | **Highly related fields** | Raw citations | 1747 (<0.001) | 418 ( 0.169) | -4.81 ( 0.539) |
|  |  | Composite | 2.82 (<0.001) | .0974 ( 0.108) | .00255 ( 0.104) |
| **Evolutionary Biology ( 11%)** | **Other fields** | Raw citations | 1049 (<0.001) | 465 ( 0.145) | 8.82 ( 0.257) |
|  |  | Composite | 2.99 (<0.001) | .0804 ( 0.101) | .00719 (<0.001) |
| **Artificial Intelligence & Image Processing ( 11%)** | **Other fields** | Raw citations | 1797 (<0.001) | -95.5 ( 0.694) | -4.07 ( 0.512) |
|  |  | Composite | 2.78 (<0.001) | -.0148 ( 0.670) | .00718 (<0.001) |
| **Veterinary Sciences ( 11%)** | **Highly related fields** | Raw citations | 464 (<0.001) | 158 ( 0.004) | 1.12 ( 0.462) |
|  |  | Composite | 2.5 (<0.001) | .0864 ( 0.013) | .00304 ( 0.001) |
| **Plant Biology & Botany ( 10%)** | **Other fields** | Raw citations | 1183 (<0.001) | 578 (<0.001) | -1.7 ( 0.593) |
|  |  | Composite | 2.87 (<0.001) | .114 ( 0.003) | .00448 (<0.001) |
| **Chemical Physics ( 10%)** | **Other fields** | Raw citations | 1676 (<0.001) | 315 ( 0.274) | 5.26 ( 0.297) |
|  |  | Composite | 3.02 (<0.001) | .112 ( 0.014) | .0042 (<0.001) |
| **Industrial Engineering & Automation ( 10%)** | **Other fields** | Raw citations | 1099 (<0.001) | 55.1 ( 0.709) | -2.81 ( 0.420) |
|  |  | Composite | 2.89 (<0.001) | -.019 ( 0.720) | .00283 ( 0.024) |
| **Sport Sciences ( 10%)** | **Highly related fields** | Raw citations | 895 ( 0.005) | 388 ( 0.261) | 11.8 ( 0.218) |
|  |  | Composite | 3.04 (<0.001) | .0182 ( 0.814) | .00208 ( 0.333) |
| **Drama & Theater ( 10%)** | **Other fields** | Raw citations | 82.1 ( 0.212) | 3.11 ( 0.960) | -.644 ( 0.765) |
|  |  | Composite | 1.95 (<0.001) | .146 ( 0.424) | .00562 ( 0.376) |
| **Social Work ( 9.6%)** | **Other fields** | Raw citations | 143 ( 0.105) | 551 (<0.001) | 8.07 ( 0.010) |
|  |  | Composite | 2.74 (<0.001) | .198 ( 0.010) | .00226 ( 0.262) |
| **Criminology ( 9.6%)** | **Other fields** | Raw citations | 395 (<0.001) | 295 ( 0.023) | 9.47 ( 0.003) |
|  |  | Composite | 2.98 (<0.001) | .121 ( 0.063) | .00524 ( 0.001) |
| **Design Practice & Management ( 9%)** | **Other fields** | Raw citations | 606 (<0.001) | 437 ( 0.055) | .541 ( 0.897) |
|  |  | Composite | 2.79 (<0.001) | -.0125 ( 0.929) | .00115 ( 0.657) |
| **Polymers ( 8.5%)** | **Other fields** | Raw citations | 1292 (<0.001) | 556 ( 0.016) | -1.92 ( 0.633) |
|  |  | Composite | 2.88 (<0.001) | .0689 ( 0.266) | .00304 ( 0.005) |
| **General Clinical Medicine ( 7.8%)** | **Highly related fields** | Raw citations | 282 ( 0.050) | 404 ( 0.051) | 4.72 ( 0.204) |
|  |  | Composite | 2.2 (<0.001) | .0323 ( 0.786) | .00712 ( 0.001) |
| **Geography ( 7.5%)** | **Other fields** | Raw citations | 283 ( 0.020) | 356 ( 0.037) | 8.5 ( 0.016) |
|  |  | Composite | 2.98 (<0.001) | .169 ( 0.085) | .00521 ( 0.011) |
| **Social Psychology ( 7.5%)** | **Highly related fields** | Raw citations | 1188 (<0.001) | 424 ( 0.146) | 9.77 ( 0.045) |
|  |  | Composite | 3.19 (<0.001) | .139 ( 0.042) | .00442 (<0.001) |
| **Development Studies ( 7.4%)** | **Other fields** | Raw citations | 588 ( 0.014) | -109 ( 0.685) | 1.96 ( 0.759) |
|  |  | Composite | 3.13 (<0.001) | -.199 ( 0.293) | .00232 ( 0.604) |
| **Economics ( 7.2%)** | **Other fields** | Raw citations | 743 (<0.001) | 387 ( 0.012) | 4.46 ( 0.119) |
|  |  | Composite | 3.08 (<0.001) | .161 ( 0.005) | .00515 (<0.001) |
| **Fluids & Plasmas ( 7.1%)** | **Other fields** | Raw citations | 1573 (<0.001) | 505 ( 0.153) | -9.19 ( 0.148) |
|  |  | Composite | 3.1 (<0.001) | .158 ( 0.037) | .00142 ( 0.297) |
| **Social Sciences Methods ( 7%)** | **Other fields** | Raw citations | 964 ( 0.103) | 1194 ( 0.212) | 9.64 ( 0.459) |
|  |  | Composite | 3.14 (<0.001) | .485 ( 0.002) | .00348 ( 0.103) |
| **Entomology ( 6.9%)** | **Other fields** | Raw citations | 984 (<0.001) | 440 (<0.001) | -8.51 ( 0.001) |
|  |  | Composite | 2.92 (<0.001) | .27 (<0.001) | -.00155 ( 0.266) |
| **Numerical & Computational Mathematics ( 6.9%)** | **Other fields** | Raw citations | 538 ( 0.006) | 556 ( 0.024) | 5.04 ( 0.285) |
|  |  | Composite | 3.1 (<0.001) | .0774 ( 0.514) | -.0000828 ( 0.971) |
| **Electrical & Electronic Engineering ( 6.9%)** | **Other fields** | Raw citations | 416 (<0.001) | 170 ( 0.330) | 10 ( 0.003) |
|  |  | Composite | 2.46 (<0.001) | .0702 ( 0.350) | .00625 (<0.001) |
| **Gender Studies ( 6.7%)** | **Highly related fields** | Raw citations | 395 ( 0.084) | -186 ( 0.531) | 2.48 ( 0.681) |
|  |  | Composite | 2.9 (<0.001) | .0222 ( 0.912) | .00468 ( 0.268) |
| **Computer Hardware & Architecture ( 6.6%)** | **Other fields** | Raw citations | 597 (<0.001) | 119 ( 0.413) | 2.76 ( 0.363) |
|  |  | Composite | 2.46 (<0.001) | .0401 ( 0.587) | .00471 ( 0.003) |
| **Logistics & Transportation ( 6.6%)** | **Other fields** | Raw citations | 667 (<0.001) | 102 ( 0.683) | 7.13 ( 0.174) |
|  |  | Composite | 2.96 (<0.001) | -.0995 ( 0.366) | .00357 ( 0.124) |
| **Food Science ( 6.5%)** | **Other fields** | Raw citations | 1030 ( 0.003) | 528 ( 0.317) | 4.53 ( 0.630) |
|  |  | Composite | 2.87 (<0.001) | .0813 ( 0.424) | .00537 ( 0.003) |
| **Information & Library Sciences ( 6.3%)** | **Other fields** | Raw citations | 253 ( 0.001) | 172 ( 0.218) | 1.77 ( 0.457) |
|  |  | Composite | 2.62 (<0.001) | .235 ( 0.061) | .00172 ( 0.417) |
| **Psychoanalysis ( 6.1%)** | **Highly related fields** | Raw citations | 51 ( 0.729) | 182 ( 0.366) | 2.62 ( 0.329) |
|  |  | Composite | 2.5 (<0.001) | .236 ( 0.389) | .000928 ( 0.799) |
| **Education ( 6%)** | **Other fields** | Raw citations | 346 (<0.001) | 233 ( 0.004) | 7.41 (<0.001) |
|  |  | Composite | 2.89 (<0.001) | .0232 ( 0.606) | .0042 (<0.001) |
| **Archaeology ( 5.9%)** | **Other fields** | Raw citations | 417 (<0.001) | 3.67 ( 0.984) | .996 ( 0.725) |
|  |  | Composite | 2.81 (<0.001) | -.0678 ( 0.535) | .00151 ( 0.386) |
| **Optoelectronics & Photonics ( 5.9%)** | **Other fields** | Raw citations | 477 (<0.001) | 222 ( 0.038) | 1.78 ( 0.333) |
|  |  | Composite | 2.31 (<0.001) | .0967 ( 0.072) | .00576 (<0.001) |
| **History of Social Sciences ( 5.9%)** | **Other fields** | Raw citations | -114 ( 0.589) | -126 ( 0.655) | 10.6 ( 0.043) |
|  |  | Composite | 2.26 (<0.001) | -.237 ( 0.403) | .014 ( 0.011) |
| **Science Studies ( 5.7%)** | **Other fields** | Raw citations | 244 ( 0.310) | -101 ( 0.767) | 12.1 ( 0.066) |
|  |  | Composite | 3.03 (<0.001) | -.185 ( 0.456) | .00637 ( 0.174) |
| **Human Factors ( 5.6%)** | **Highly related fields** | Raw citations | 739 (<0.001) | 45.7 ( 0.854) | 7.17 ( 0.152) |
|  |  | Composite | 2.88 (<0.001) | .00908 ( 0.921) | .00762 (<0.001) |
| **Urban & Regional Planning ( 5.6%)** | **Other fields** | Raw citations | 561 ( 0.001) | 332 ( 0.249) | .96 ( 0.842) |
|  |  | Composite | 3.13 (<0.001) | .195 ( 0.210) | .00245 ( 0.346) |
| **Networking & Telecommunications ( 5.2%)** | **Other fields** | Raw citations | 886 (<0.001) | -177 ( 0.208) | -1.95 ( 0.335) |
|  |  | Composite | 2.63 (<0.001) | -.0825 ( 0.105) | .0037 (<0.001) |
| **Dairy & Animal Science ( 5.1%)** | **Other fields** | Raw citations | 730 (<0.001) | 233 ( 0.040) | -3.4 ( 0.070) |
|  |  | Composite | 2.63 (<0.001) | .0804 ( 0.271) | .0025 ( 0.039) |
| **Languages & Linguistics ( 5.1%)** | **Other fields** | Raw citations | 427 (<0.001) | -57.6 ( 0.656) | .771 ( 0.749) |
|  |  | Composite | 3.05 (<0.001) | -.00505 ( 0.967) | .00283 ( 0.216) |
| **Building & Construction ( 4.9%)** | **Other fields** | Raw citations | 752 ( 0.001) | 449 ( 0.239) | 12.7 ( 0.065) |
|  |  | Composite | 2.94 (<0.001) | .0109 ( 0.937) | .00728 ( 0.004) |
| **Religions & Theology ( 4.9%)** | **Other fields** | Raw citations | 143 ( 0.002) | 12.9 ( 0.895) | -.325 ( 0.827) |
|  |  | Composite | 2.33 (<0.001) | .0544 ( 0.763) | .000519 ( 0.850) |
| **Inorganic & Nuclear Chemistry ( 4.9%)** | **Other fields** | Raw citations | 909 ( 0.001) | 196 ( 0.677) | 5.08 ( 0.390) |
|  |  | Composite | 2.77 (<0.001) | -.000647 ( 0.995) | .00296 ( 0.037) |
| **Applied Mathematics ( 4.8%)** | **Other fields** | Raw citations | 839 ( 0.030) | 2290 (<0.001) | 9.42 ( 0.301) |
|  |  | Composite | 3.03 (<0.001) | .355 ( 0.045) | .00509 ( 0.052) |
| **Mechanical Engineering & Transports ( 4.7%)** | **Other fields** | Raw citations | 971 (<0.001) | 653 ( 0.005) | -3.37 ( 0.311) |
|  |  | Composite | 2.83 (<0.001) | -.045 ( 0.589) | .00343 ( 0.004) |
| **General Psychology & Cognitive Sciences ( 4.7%)** | **Highly related fields** | Raw citations | 664 (<0.001) | 185 ( 0.530) | .446 ( 0.924) |
|  |  | Composite | 3 (<0.001) | -.184 ( 0.299) | .000869 ( 0.757) |
| **Mining & Metallurgy ( 4.7%)** | **Other fields** | Raw citations | 154 ( 0.286) | 528 ( 0.036) | 5.41 ( 0.141) |
|  |  | Composite | 2.25 (<0.001) | .438 ( 0.062) | .0068 ( 0.051) |
| **Software Engineering ( 4.5%)** | **Other fields** | Raw citations | 1084 (<0.001) | 293 ( 0.084) | -13.3 (<0.001) |
|  |  | Composite | 2.8 (<0.001) | -.0675 ( 0.447) | .00108 ( 0.535) |
| **Anthropology ( 4.3%)** | **Other fields** | Raw citations | 291 (<0.001) | 113 ( 0.344) | 4.83 ( 0.018) |
|  |  | Composite | 2.97 (<0.001) | -.0875 ( 0.415) | .00273 ( 0.135) |
| **Materials ( 4.3%)** | **Other fields** | Raw citations | 1235 (<0.001) | 154 ( 0.550) | -1.51 ( 0.626) |
|  |  | Composite | 2.75 (<0.001) | .103 ( 0.087) | .00508 (<0.001) |
| **Chemical Engineering ( 4.3%)** | **Other fields** | Raw citations | 1517 (<0.001) | 355 ( 0.340) | -8.67 ( 0.063) |
|  |  | Composite | 2.98 (<0.001) | .0989 ( 0.410) | .00212 ( 0.159) |
| **Accounting ( 4.1%)** | **Other fields** | Raw citations | 457 ( 0.017) | -35.1 ( 0.902) | 12.4 ( 0.033) |
|  |  | Composite | 2.94 (<0.001) | -.125 ( 0.443) | .00754 ( 0.024) |
| **Econometrics ( 4.1%)** | **Other fields** | Raw citations | 591 ( 0.479) | -366 ( 0.763) | 27.9 ( 0.209) |
|  |  | Composite | 3.27 (<0.001) | -.44 ( 0.187) | .00673 ( 0.265) |
| **Ecology ( 3.9%)** | **Other fields** | Raw citations | 1298 (<0.001) | -72.3 ( 0.806) | 11.6 ( 0.025) |
|  |  | Composite | 3.07 (<0.001) | .0478 ( 0.422) | .00549 (<0.001) |
| **Sport, Leisure & Tourism ( 3.8%)** | **Other fields** | Raw citations | 701 (<0.001) | -147 ( 0.576) | 4.48 ( 0.382) |
|  |  | Composite | 3.09 (<0.001) | -.0594 ( 0.650) | .00439 ( 0.086) |
| **Legal & Forensic Medicine ( 3.8%)** | **Highly related fields** | Raw citations | 277 ( 0.003) | -118 ( 0.515) | .588 ( 0.850) |
|  |  | Composite | 2.22 (<0.001) | -.037 ( 0.837) | .00542 ( 0.084) |
| **Communication & Media Studies ( 3.7%)** | **Other fields** | Raw citations | 108 ( 0.441) | 6.53 ( 0.984) | 20.6 (<0.001) |
|  |  | Composite | 3.04 (<0.001) | -.0162 ( 0.877) | .00426 ( 0.006) |
| **Physical Chemistry ( 3.7%)** | **Other fields** | Raw citations | 1889 (<0.001) | -162 ( 0.837) | 1.19 ( 0.893) |
|  |  | Composite | 2.92 (<0.001) | -.0478 ( 0.798) | .00667 ( 0.002) |
| **Operations Research ( 3.6%)** | **Other fields** | Raw citations | 523 ( 0.040) | -82 ( 0.857) | 12.8 ( 0.054) |
|  |  | Composite | 3.09 (<0.001) | .0494 ( 0.709) | .00316 ( 0.102) |
| **Anatomy & Morphology ( 3.6%)** | **Highly related fields** | Raw citations | 344 (<0.001) | 60.3 ( 0.756) | -1.92 ( 0.357) |
|  |  | Composite | 2.46 (<0.001) | -.0653 ( 0.743) | -.000483 ( 0.821) |
| **Information Systems ( 3.6%)** | **Other fields** | Raw citations | 1212 (<0.001) | -39.3 ( 0.943) | -1.41 ( 0.887) |
|  |  | Composite | 3.16 (<0.001) | .0141 ( 0.923) | .00017 ( 0.948) |
| **Marketing ( 3.5%)** | **Other fields** | Raw citations | 195 ( 0.377) | -72.8 ( 0.841) | 27.6 (<0.001) |
|  |  | Composite | 3.01 (<0.001) | .00894 ( 0.932) | .00571 ( 0.005) |
| **Ornithology ( 3.4%)** | **Other fields** | Raw citations | 404 ( 0.002) | -16.7 ( 0.952) | .54 ( 0.850) |
|  |  | Composite | 2.57 (<0.001) | .287 ( 0.139) | .00354 ( 0.077) |
| **General Physics ( 3.3%)** | **Other fields** | Raw citations | 1266 (<0.001) | 344 ( 0.265) | -6.75 ( 0.032) |
|  |  | Composite | 2.78 (<0.001) | .0854 ( 0.379) | .00337 ( 0.001) |
| **Finance ( 3.3%)** | **Other fields** | Raw citations | 571 ( 0.004) | -336 ( 0.372) | 14.1 ( 0.017) |
|  |  | Composite | 3 (<0.001) | -.145 ( 0.286) | .00721 ( 0.001) |
| **Zoology ( 3.3%)** | **Other fields** | Raw citations | 428 ( 0.001) | 257 ( 0.234) | .604 ( 0.869) |
|  |  | Composite | 2.65 (<0.001) | .0414 ( 0.772) | -.0011 ( 0.653) |
| **Oceanography ( 3%)** | **Other fields** | Raw citations | 1007 (<0.001) | 299 ( 0.229) | -5.66 ( 0.119) |
|  |  | Composite | 2.92 (<0.001) | .211 ( 0.082) | .00157 ( 0.373) |
| **Meteorology & Atmospheric Sciences ( 3%)** | **Other fields** | Raw citations | 1771 (<0.001) | 138 ( 0.587) | 1.29 ( 0.723) |
|  |  | Composite | 3.09 (<0.001) | .0817 ( 0.124) | .00339 (<0.001) |
| **Applied Physics ( 2.8%)** | **Other fields** | Raw citations | 1450 (<0.001) | 299 ( 0.275) | -2.3 ( 0.387) |
|  |  | Composite | 2.84 (<0.001) | .0262 ( 0.645) | .00263 (<0.001) |
| **Geological & Geomatics Engineering ( 2.8%)** | **Other fields** | Raw citations | 1323 (<0.001) | 912 ( 0.019) | -1.47 ( 0.784) |
|  |  | Composite | 2.96 (<0.001) | .271 ( 0.017) | .00517 ( 0.001) |
| **Aerospace & Aeronautics ( 2.8%)** | **Other fields** | Raw citations | 401 (<0.001) | 93.9 ( 0.289) | -.0888 ( 0.936) |
|  |  | Composite | 2.44 (<0.001) | -.0482 ( 0.534) | .00448 (<0.001) |
| **Marine Biology & Hydrobiology ( 2.8%)** | **Other fields** | Raw citations | 1529 (<0.001) | 61.8 ( 0.850) | -8.91 ( 0.090) |
|  |  | Composite | 3.06 (<0.001) | -.0715 ( 0.480) | .00219 ( 0.179) |
| **Cultural Studies ( 2.8%)** | **Other fields** | Raw citations | 74.7 ( 0.319) | -92.6 ( 0.578) | 2.56 ( 0.252) |
|  |  | Composite | 2.44 (<0.001) | -.17 ( 0.551) | .0056 ( 0.147) |
| **Energy ( 2.6%)** | **Other fields** | Raw citations | 1012 (<0.001) | 304 ( 0.182) | .717 ( 0.770) |
|  |  | Composite | 2.85 (<0.001) | .0828 ( 0.233) | .00398 (<0.001) |
| **Environmental Engineering ( 2.5%)** | **Other fields** | Raw citations | 1064 (<0.001) | 1045 ( 0.002) | -1.87 ( 0.658) |
|  |  | Composite | 2.97 (<0.001) | .111 ( 0.282) | .002 ( 0.123) |
| **Geology ( 2.4%)** | **Other fields** | Raw citations | 926 (<0.001) | -295 ( 0.341) | -6.59 ( 0.108) |
|  |  | Composite | 3.04 (<0.001) | -.168 ( 0.410) | -.000855 ( 0.750) |
| **Business & Management ( 2.3%)** | **Other fields** | Raw citations | 761 (<0.001) | -18.9 ( 0.948) | 12.4 ( 0.001) |
|  |  | Composite | 3.11 (<0.001) | .0511 ( 0.525) | .00466 (<0.001) |
| **Strategic, Defence & Security Studies ( 2.3%)** | **Other fields** | Raw citations | 246 ( 0.105) | -146 ( 0.704) | 11 ( 0.014) |
|  |  | Composite | 2.7 (<0.001) | .128 ( 0.490) | .00777 (<0.001) |
| **Computation Theory & Mathematics ( 2.3%)** | **Other fields** | Raw citations | 686 (<0.001) | 344 ( 0.340) | -.292 ( 0.942) |
|  |  | Composite | 2.93 (<0.001) | .203 ( 0.184) | .00233 ( 0.169) |
| **Law ( 2.2%)** | **Other fields** | Raw citations | 81.5 ( 0.691) | -4.51 ( 0.990) | 6.04 ( 0.411) |
|  |  | Composite | 2.45 (<0.001) | -.00751 ( 0.970) | .00446 ( 0.274) |
| **Political Science & Public Administration ( 2%)** | **Other fields** | Raw citations | 463 (<0.001) | 1092 (<0.001) | 6.15 ( 0.040) |
|  |  | Composite | 3.05 (<0.001) | .312 ( 0.007) | .00406 ( 0.001) |
| **History ( 2%)** | **Other fields** | Raw citations | 72.9 ( 0.026) | -33.6 ( 0.729) | .299 ( 0.726) |
|  |  | Composite | 2.12 (<0.001) | -.0867 ( 0.623) | .00174 ( 0.262) |
| **International Relations ( 1.8%)** | **Other fields** | Raw citations | 120 ( 0.151) | -104 ( 0.655) | 9.67 (<0.001) |
|  |  | Composite | 2.82 (<0.001) | -.296 ( 0.231) | .00875 ( 0.002) |
| **Astronomy & Astrophysics ( 1.7%)** | **Other fields** | Raw citations | 3149 (<0.001) | -813 ( 0.213) | -24.2 (<0.001) |
|  |  | Composite | 3.17 (<0.001) | .0159 ( 0.837) | .000621 ( 0.438) |
| **General Mathematics ( 1.7%)** | **Other fields** | Raw citations | 452 (<0.001) | 82.7 ( 0.590) | -.899 ( 0.410) |
|  |  | Composite | 2.9 (<0.001) | -.0362 ( 0.732) | .000608 ( 0.418) |
| **Fisheries ( 1.6%)** | **Other fields** | Raw citations | 713 (<0.001) | 394 ( 0.172) | .749 ( 0.830) |
|  |  | Composite | 2.77 (<0.001) | .158 ( 0.337) | .00279 ( 0.162) |
| **Philosophy ( 1.6%)** | **Other fields** | Raw citations | 242 (<0.001) | 476 (<0.001) | .346 ( 0.761) |
|  |  | Composite | 2.91 (<0.001) | .712 (<0.001) | .000684 ( 0.676) |
| **Literary Studies ( 1.4%)** | **Other fields** | Raw citations | 30.4 ( 0.415) | 24 ( 0.820) | 1.96 ( 0.143) |
|  |  | Composite | 1.93 (<0.001) | .34 ( 0.201) | .00816 ( 0.016) |
| **Agricultural Economics & Policy ( 1.4%)** | **Other fields** | Raw citations | 990 (<0.001) | -8.71 ( 0.989) | -2.71 ( 0.698) |
|  |  | Composite | 3.24 (<0.001) | .033 ( 0.915) | -.0016 ( 0.641) |
| **Civil Engineering ( 1.4%)** | **Other fields** | Raw citations | 533 (<0.001) | 218 ( 0.591) | 6.66 ( 0.072) |
|  |  | Composite | 2.77 (<0.001) | -.00115 ( 0.995) | .00495 ( 0.007) |
| **Geochemistry & Geophysics ( 1.3%)** | **Other fields** | Raw citations | 1303 (<0.001) | -225 ( 0.359) | -4.81 ( 0.042) |
|  |  | Composite | 3.15 (<0.001) | -.101 ( 0.271) | .00174 ( 0.050) |
| **Forestry ( 1.1%)** | **Other fields** | Raw citations | 652 (<0.001) | -132 ( 0.681) | -.812 ( 0.812) |
|  |  | Composite | 2.62 (<0.001) | -.0327 ( 0.873) | .00351 ( 0.106) |
| **Nuclear & Particle Physics ( 1.1%)** | **Other fields** | Raw citations | 3809 (<0.001) | -755 ( 0.501) | -40.4 (<0.001) |
|  |  | Composite | 2.89 (<0.001) | .0162 ( 0.880) | .00251 (<0.001) |
| **Agronomy & Agriculture ( .83%)** | **Other fields** | Raw citations | 1177 (<0.001) | 432 ( 0.482) | -2.86 ( 0.486) |
|  |  | Composite | 2.88 (<0.001) | .226 ( 0.227) | .00291 ( 0.020) |
| **Paleontology ( .74%)** | **Other fields** | Raw citations | 1198 (<0.001) | -586 ( 0.551) | -4.82 ( 0.489) |
|  |  | Composite | 3.14 (<0.001) | -.258 ( 0.241) | -.0000303 ( 0.984) |
| **Music ( 0%)** | **Other fields** | Raw citations |  |  |  |
|  |  | Composite |  |  |  |
| **Mathematical Physics ( 0%)** | **Other fields** | Raw citations |  |  |  |
|  |  | Composite |  |  |  |
| **Industrial Relations ( 0%)** | **Other fields** | Raw citations |  |  |  |
|  |  | Composite |  |  |  |
| **Horticulture ( 0%)** | **Other fields** | Raw citations |  |  |  |
|  |  | Composite |  |  |  |
| **History of Science, Technology & Medicine ( 0%)** | **Other fields** | Raw citations |  |  |  |
|  |  | Composite |  |  |  |
| **Folklore ( 0%)** | **Other fields** | Raw citations |  |  |  |
|  |  | Composite |  |  |  |
| **Economic Theory ( 0%)** | **Other fields** | Raw citations |  |  |  |
|  |  | Composite |  |  |  |
| **Classics ( 0%)** | **Other fields** | Raw citations |  |  |  |
|  |  | Composite |  |  |  |
| **Automobile Design & Engineering ( 0%)** | **Other fields** | Raw citations |  |  |  |
|  |  | Composite |  |  |  |
| **Art Practice, History & Theory ( 0%)** | **Other fields** | Raw citations |  |  |  |
|  |  | Composite |  |  |  |
| **Architecture ( 0%)** | **Other fields** | Raw citations |  |  |  |
|  |  | Composite |  |  |  |

**Supplementary Table 3.6 : Recent year impact, Funding time current funding Linear Regressions for each subfield (ordered by percentage funded)**

| **Top-cited US-based researchers: Subfield (perc. funded)** | **Classification** | **Dependent Variable** | **Constant (p-val)** | **Funded (p-val)** | **Years since first pub (p-val)** |
| --- | --- | --- | --- | --- | --- |
| **Geriatrics ( 43%)** | **Highly related fields** | Raw citations | 1293 ( 0.144) | 330 ( 0.501) | 16.3 ( 0.454) |
|  |  | Composite | 2.97 (<0.001) | .083 ( 0.352) | .00586 ( 0.141) |
| **Developmental Biology ( 36%)** | **Highly related fields** | Raw citations | 4064 (<0.001) | 253 ( 0.282) | -27.8 ( 0.004) |
|  |  | Composite | 3.02 (<0.001) | .0166 ( 0.342) | .00658 (<0.001) |
| **Medical Informatics ( 34%)** | **Highly related fields** | Raw citations | 790 ( 0.001) | 204 ( 0.286) | 2.4 ( 0.749) |
|  |  | Composite | 2.66 (<0.001) | .0811 ( 0.190) | .00485 ( 0.048) |
| **Bioinformatics ( 34%)** | **Highly related fields** | Raw citations | 3646 (<0.001) | 633 ( 0.345) | -13 ( 0.650) |
|  |  | Composite | 3 (<0.001) | .0606 ( 0.315) | .00348 ( 0.180) |
| **Substance Abuse ( 31%)** | **Highly related fields** | Raw citations | 804 (<0.001) | 306 ( 0.033) | 13.8 ( 0.018) |
|  |  | Composite | 3.03 (<0.001) | -.00341 ( 0.933) | .00423 ( 0.010) |
| **Gerontology ( 31%)** | **Highly related fields** | Raw citations | 1530 ( 0.155) | 18.8 ( 0.979) | 12.4 ( 0.657) |
|  |  | Composite | 3.02 (<0.001) | -.00842 ( 0.900) | .00507 ( 0.060) |
| **Biomedical Engineering ( 30%)** | **Highly related fields** | Raw citations | 684 ( 0.002) | 526 ( 0.003) | 15.9 ( 0.013) |
|  |  | Composite | 2.69 (<0.001) | .0568 ( 0.071) | .00733 (<0.001) |
| **Virology ( 30%)** | **Highly related fields** | Raw citations | 1648 (<0.001) | 289 ( 0.064) | 1.28 ( 0.823) |
|  |  | Composite | 2.75 (<0.001) | .0665 ( 0.010) | .00559 (<0.001) |
| **Immunology ( 30%)** | **Highly related fields** | Raw citations | 2113 (<0.001) | -20.8 ( 0.881) | 13.6 ( 0.011) |
|  |  | Composite | 2.98 (<0.001) | .0336 ( 0.110) | .00596 (<0.001) |
| **Neurology & Neurosurgery ( 29%)** | **Highly related fields** | Raw citations | 2058 (<0.001) | 147 ( 0.130) | 2.82 ( 0.424) |
|  |  | Composite | 3.06 (<0.001) | .0234 ( 0.109) | .00424 (<0.001) |
| **Allergy ( 27%)** | **Highly related fields** | Raw citations | 1238 ( 0.001) | 56.5 ( 0.830) | 17.6 ( 0.072) |
|  |  | Composite | 2.97 (<0.001) | .0356 ( 0.587) | .00594 ( 0.016) |
| **Emergency & Critical Care Medicine ( 27%)** | **Highly related fields** | Raw citations | 1029 ( 0.014) | 782 ( 0.011) | 22.2 ( 0.078) |
|  |  | Composite | 2.82 (<0.001) | .102 ( 0.033) | .0042 ( 0.034) |
| **Oncology & Carcinogenesis ( 26%)** | **Highly related fields** | Raw citations | 3201 (<0.001) | 381 ( 0.026) | -5.11 ( 0.429) |
|  |  | Composite | 2.93 (<0.001) | .0316 ( 0.031) | .00459 (<0.001) |
| **Public Health ( 26%)** | **Highly related fields** | Raw citations | 881 ( 0.005) | 38.7 ( 0.870) | 27.5 ( 0.002) |
|  |  | Composite | 3.02 (<0.001) | .0222 ( 0.412) | .00512 (<0.001) |
| **Gastroenterology & Hepatology ( 24%)** | **Highly related fields** | Raw citations | 1731 (<0.001) | 269 ( 0.128) | 3.51 ( 0.577) |
|  |  | Composite | 2.92 (<0.001) | .0809 ( 0.021) | .00443 (<0.001) |
| **Psychiatry ( 23%)** | **Highly related fields** | Raw citations | 1246 (<0.001) | 783 (<0.001) | 25.6 ( 0.001) |
|  |  | Composite | 3.05 (<0.001) | .0432 ( 0.161) | .0061 (<0.001) |
| **Arthritis & Rheumatology ( 23%)** | **Highly related fields** | Raw citations | 1826 ( 0.001) | 496 ( 0.197) | 14.2 ( 0.293) |
|  |  | Composite | 3.06 (<0.001) | .0397 ( 0.502) | .00425 ( 0.042) |
| **Epidemiology ( 22%)** | **Highly related fields** | Raw citations | 1526 ( 0.099) | 289 ( 0.677) | 31.8 ( 0.178) |
|  |  | Composite | 3.1 (<0.001) | -.011 ( 0.889) | .00384 ( 0.155) |
| **Genetics & Heredity ( 22%)** | **Highly related fields** | Raw citations | 2360 (<0.001) | 521 ( 0.052) | -10.8 ( 0.255) |
|  |  | Composite | 2.86 (<0.001) | .0139 ( 0.698) | .00201 ( 0.114) |
| **Analytical Chemistry ( 22%)** | **Other fields** | Raw citations | 1.0e+03 (<0.001) | 425 ( 0.008) | .584 ( 0.900) |
|  |  | Composite | 2.75 (<0.001) | .0411 ( 0.323) | .00331 ( 0.006) |
| **Ophthalmology & Optometry ( 22%)** | **Highly related fields** | Raw citations | 1306 (<0.001) | 245 ( 0.041) | -3.83 ( 0.340) |
|  |  | Composite | 2.73 (<0.001) | .0999 ( 0.001) | .00386 (<0.001) |
| **Developmental & Child Psychology ( 22%)** | **Highly related fields** | Raw citations | 712 (<0.001) | 430 ( 0.001) | 16.2 (<0.001) |
|  |  | Composite | 3.07 (<0.001) | .0305 ( 0.404) | .00545 (<0.001) |
| **Nuclear Medicine & Medical Imaging ( 22%)** | **Highly related fields** | Raw citations | 1014 (<0.001) | 333 (<0.001) | 1.32 ( 0.627) |
|  |  | Composite | 2.68 (<0.001) | .0934 (<0.001) | .00269 (<0.001) |
| **Endocrinology & Metabolism ( 21%)** | **Highly related fields** | Raw citations | 1804 (<0.001) | 33.3 ( 0.858) | 7.17 ( 0.246) |
|  |  | Composite | 3.04 (<0.001) | .0171 ( 0.577) | .00512 (<0.001) |
| **Biophysics ( 21%)** | **Highly related fields** | Raw citations | 1169 ( 0.002) | -156 ( 0.605) | -1.46 ( 0.869) |
|  |  | Composite | 2.59 (<0.001) | .0786 ( 0.184) | .00653 (<0.001) |
| **Toxicology ( 21%)** | **Highly related fields** | Raw citations | 1144 (<0.001) | 671 ( 0.001) | .418 ( 0.950) |
|  |  | Composite | 2.71 (<0.001) | .105 ( 0.012) | .00544 (<0.001) |
| **Cardiovascular System & Hematology ( 21%)** | **Highly related fields** | Raw citations | 2934 (<0.001) | 511 ( 0.009) | -3.39 ( 0.578) |
|  |  | Composite | 2.97 (<0.001) | .0645 ( 0.003) | .00391 (<0.001) |
| **Respiratory System ( 21%)** | **Highly related fields** | Raw citations | 1614 (<0.001) | 5.34 ( 0.974) | 10 ( 0.091) |
|  |  | Composite | 2.87 (<0.001) | -.00743 ( 0.807) | .00489 (<0.001) |
| **Physiology ( 21%)** | **Highly related fields** | Raw citations | 662 (<0.001) | 39.5 ( 0.642) | 3.65 ( 0.134) |
|  |  | Composite | 2.86 (<0.001) | .036 ( 0.424) | .00388 ( 0.003) |
| **Rehabilitation ( 21%)** | **Highly related fields** | Raw citations | 398 ( 0.005) | 316 ( 0.003) | 12.3 ( 0.003) |
|  |  | Composite | 2.7 (<0.001) | -.0147 ( 0.732) | .00652 (<0.001) |
| **Microbiology ( 20%)** | **Highly related fields** | Raw citations | 2630 (<0.001) | 173 ( 0.297) | -23.2 (<0.001) |
|  |  | Composite | 2.93 (<0.001) | .0481 ( 0.028) | .00407 (<0.001) |
| **Urology & Nephrology ( 20%)** | **Highly related fields** | Raw citations | 1917 (<0.001) | 47.8 ( 0.780) | -7 ( 0.216) |
|  |  | Composite | 2.83 (<0.001) | .0954 ( 0.002) | .00371 (<0.001) |
| **Medicinal & Biomolecular Chemistry ( 20%)** | **Highly related fields** | Raw citations | 564 (<0.001) | 295 ( 0.021) | 11 ( 0.005) |
|  |  | Composite | 2.47 (<0.001) | .0665 ( 0.102) | .00731 (<0.001) |
| **Health Policy & Services ( 20%)** | **Highly related fields** | Raw citations | 806 ( 0.004) | 80.9 ( 0.717) | 17.4 ( 0.029) |
|  |  | Composite | 2.84 (<0.001) | .061 ( 0.266) | .00691 (<0.001) |
| **Pediatrics ( 19%)** | **Highly related fields** | Raw citations | 834 (<0.001) | 122 ( 0.109) | 3 ( 0.185) |
|  |  | Composite | 2.63 (<0.001) | .0545 ( 0.051) | .00367 (<0.001) |
| **Obstetrics & Reproductive Medicine ( 19%)** | **Highly related fields** | Raw citations | 1012 (<0.001) | 46.9 ( 0.670) | 1.57 ( 0.654) |
|  |  | Composite | 2.77 (<0.001) | .0234 ( 0.478) | .003 ( 0.004) |
| **Anesthesiology ( 18%)** | **Highly related fields** | Raw citations | 949 (<0.001) | 205 ( 0.172) | -1.18 ( 0.813) |
|  |  | Composite | 2.64 (<0.001) | .0743 ( 0.079) | .00441 ( 0.002) |
| **Statistics & Probability ( 17%)** | **Other fields** | Raw citations | 1055 ( 0.008) | 811 ( 0.036) | 13.3 ( 0.157) |
|  |  | Composite | 3.1 (<0.001) | .093 ( 0.143) | .00324 ( 0.036) |
| **Biochemistry & Molecular Biology ( 17%)** | **Highly related fields** | Raw citations | 1481 (<0.001) | 282 ( 0.003) | -3.82 ( 0.146) |
|  |  | Composite | 2.84 (<0.001) | .0613 ( 0.006) | .0044 (<0.001) |
| **Speech-Language Pathology & Audiology ( 17%)** | **Highly related fields** | Raw citations | 1188 (<0.001) | -254 ( 0.262) | -11.2 ( 0.130) |
|  |  | Composite | 2.82 (<0.001) | -.0227 ( 0.728) | .004 ( 0.062) |
| **Demography ( 17%)** | **Highly related fields** | Raw citations | -70.6 ( 0.826) | 504 ( 0.072) | 17.8 ( 0.028) |
|  |  | Composite | 2.79 (<0.001) | .0537 ( 0.684) | .00757 ( 0.050) |
| **Experimental Psychology ( 16%)** | **Highly related fields** | Raw citations | 969 (<0.001) | 476 (<0.001) | 4.14 ( 0.166) |
|  |  | Composite | 3.11 (<0.001) | .12 ( 0.002) | .00407 (<0.001) |
| **Pharmacology & Pharmacy ( 15%)** | **Highly related fields** | Raw citations | 692 (<0.001) | 195 ( 0.034) | 3.56 ( 0.134) |
|  |  | Composite | 2.6 (<0.001) | .0519 ( 0.145) | .00485 (<0.001) |
| **Clinical Psychology ( 15%)** | **Highly related fields** | Raw citations | 1058 (<0.001) | 217 ( 0.386) | 14.2 ( 0.025) |
|  |  | Composite | 3.11 (<0.001) | .0157 ( 0.817) | .00442 ( 0.010) |
| **Applied Ethics ( 15%)** | **Highly related fields** | Raw citations | 816 (<0.001) | 328 ( 0.081) | -3.75 ( 0.486) |
|  |  | Composite | 3.02 (<0.001) | .0772 ( 0.427) | .000686 ( 0.807) |
| **Organic Chemistry ( 15%)** | **Other fields** | Raw citations | 1337 (<0.001) | 263 ( 0.080) | -1.46 ( 0.659) |
|  |  | Composite | 2.86 (<0.001) | .0742 ( 0.062) | .004 (<0.001) |
| **Nutrition & Dietetics ( 14%)** | **Highly related fields** | Raw citations | 1129 (<0.001) | 397 ( 0.052) | 7.14 ( 0.199) |
|  |  | Composite | 2.98 (<0.001) | .0591 ( 0.247) | .00485 ( 0.001) |
| **Mycology & Parasitology ( 14%)** | **Highly related fields** | Raw citations | 1379 (<0.001) | -244 ( 0.312) | -9.84 ( 0.101) |
|  |  | Composite | 2.8 (<0.001) | -.062 ( 0.480) | .00148 ( 0.494) |
| **Nursing ( 14%)** | **Highly related fields** | Raw citations | 158 ( 0.005) | 271 (<0.001) | 8.23 (<0.001) |
|  |  | Composite | 2.4 (<0.001) | .0782 ( 0.015) | .00498 (<0.001) |
| **Optics ( 14%)** | **Other fields** | Raw citations | 1011 (<0.001) | 388 ( 0.103) | 14 ( 0.038) |
|  |  | Composite | 2.68 (<0.001) | .0331 ( 0.498) | .00778 (<0.001) |
| **General Chemistry ( 13%)** | **Other fields** | Raw citations | 1421 (<0.001) | 127 ( 0.717) | .507 ( 0.915) |
|  |  | Composite | 2.62 (<0.001) | .171 ( 0.009) | .00576 (<0.001) |
| **General & Internal Medicine ( 13%)** | **Highly related fields** | Raw citations | 3268 (<0.001) | -184 ( 0.648) | -35.8 (<0.001) |
|  |  | Composite | 2.61 (<0.001) | .159 (<0.001) | .00227 ( 0.001) |
| **Surgery ( 13%)** | **Highly related fields** | Raw citations | 1002 (<0.001) | 380 ( 0.002) | 4.97 ( 0.110) |
|  |  | Composite | 2.62 (<0.001) | .0801 ( 0.002) | .00369 (<0.001) |
| **Nanoscience & Nanotechnology ( 13%)** | **Other fields** | Raw citations | 1746 (<0.001) | -224 ( 0.664) | 99 (<0.001) |
|  |  | Composite | 2.99 (<0.001) | .0258 ( 0.561) | .0132 (<0.001) |
| **Otorhinolaryngology ( 12%)** | **Highly related fields** | Raw citations | 880 (<0.001) | 76.9 ( 0.537) | -4.58 ( 0.148) |
|  |  | Composite | 2.6 (<0.001) | .0574 ( 0.101) | .00219 ( 0.014) |
| **Environmental & Occupational Health ( 12%)** | **Highly related fields** | Raw citations | 429 ( 0.240) | -174 ( 0.594) | 11.6 ( 0.256) |
|  |  | Composite | 2.56 (<0.001) | -.007 ( 0.926) | .00397 ( 0.092) |
| **Tropical Medicine ( 12%)** | **Highly related fields** | Raw citations | 1405 (<0.001) | 391 ( 0.343) | -7.44 ( 0.436) |
|  |  | Composite | 2.6 (<0.001) | .0677 ( 0.239) | .00472 (<0.001) |
| **Microscopy ( 12%)** | **Highly related fields** | Raw citations | 684 ( 0.229) | 16 ( 0.977) | 10.2 ( 0.490) |
|  |  | Composite | 2.42 (<0.001) | .0482 ( 0.808) | .0108 ( 0.046) |
| **Biotechnology ( 12%)** | **Highly related fields** | Raw citations | 720 ( 0.003) | 56.2 ( 0.836) | 21.1 ( 0.005) |
|  |  | Composite | 2.64 (<0.001) | .0649 ( 0.355) | .0109 (<0.001) |
| **Drama & Theater ( 10%)** | **Other fields** | Raw citations | 82.1 ( 0.212) | 3.11 ( 0.960) | -.644 ( 0.765) |
|  |  | Composite | 1.95 (<0.001) | .146 ( 0.424) | .00562 ( 0.376) |
| **Complementary & Alternative Medicine ( 10%)** | **Highly related fields** | Raw citations | 266 ( 0.035) | 297 ( 0.027) | 4.52 ( 0.295) |
|  |  | Composite | 2.52 (<0.001) | .272 ( 0.041) | .00146 ( 0.732) |
| **Acoustics ( 9.9%)** | **Other fields** | Raw citations | 962 (<0.001) | -7.41 ( 0.960) | -7.83 ( 0.006) |
|  |  | Composite | 2.84 (<0.001) | -.0526 ( 0.428) | .000464 ( 0.711) |
| **Dermatology & Venereal Diseases ( 9.7%)** | **Highly related fields** | Raw citations | 1159 (<0.001) | 841 ( 0.003) | 1.27 ( 0.843) |
|  |  | Composite | 2.76 (<0.001) | .0544 ( 0.380) | .0037 ( 0.009) |
| **Orthopedics ( 9.5%)** | **Highly related fields** | Raw citations | 1159 (<0.001) | 275 ( 0.023) | -1.15 ( 0.681) |
|  |  | Composite | 2.8 (<0.001) | .111 ( 0.001) | .00381 (<0.001) |
| **Family Studies ( 9.1%)** | **Other fields** | Raw citations | 294 ( 0.025) | 352 ( 0.027) | 6.89 ( 0.056) |
|  |  | Composite | 2.87 (<0.001) | .0212 ( 0.833) | .00579 ( 0.014) |
| **Environmental Sciences ( 9%)** | **Other fields** | Raw citations | 1628 (<0.001) | 1024 ( 0.023) | 2.79 ( 0.783) |
|  |  | Composite | 3.06 (<0.001) | .147 ( 0.047) | .00353 ( 0.036) |
| **Behavioral Science & Comparative Psychology ( 8.7%)** | **Highly related fields** | Raw citations | 758 (<0.001) | 180 ( 0.292) | .93 ( 0.793) |
|  |  | Composite | 2.95 (<0.001) | .0981 ( 0.219) | .00487 ( 0.004) |
| **Sociology ( 7.8%)** | **Other fields** | Raw citations | 408 (<0.001) | -53.7 ( 0.606) | 5.98 ( 0.003) |
|  |  | Composite | 3.06 (<0.001) | -.0488 ( 0.467) | .00379 ( 0.004) |
| **Pathology ( 7.7%)** | **Highly related fields** | Raw citations | 1786 (<0.001) | 363 ( 0.314) | -5.4 ( 0.489) |
|  |  | Composite | 2.84 (<0.001) | .0321 ( 0.657) | .00223 ( 0.154) |
| **Design Practice & Management ( 7.5%)** | **Other fields** | Raw citations | 619 (<0.001) | 510 ( 0.037) | .181 ( 0.965) |
|  |  | Composite | 2.79 (<0.001) | .0271 ( 0.858) | .00125 ( 0.628) |
| **Dentistry ( 7.3%)** | **Highly related fields** | Raw citations | 837 (<0.001) | 335 ( 0.013) | -3.29 ( 0.192) |
|  |  | Composite | 2.72 (<0.001) | .0978 ( 0.094) | .00214 ( 0.051) |
| **Plant Biology & Botany ( 7.2%)** | **Other fields** | Raw citations | 1188 (<0.001) | 588 (<0.001) | -1.3 ( 0.684) |
|  |  | Composite | 2.87 (<0.001) | .0966 ( 0.031) | .00453 (<0.001) |
| **Evolutionary Biology ( 7.1%)** | **Other fields** | Raw citations | 1068 (<0.001) | 519 ( 0.185) | 8.71 ( 0.263) |
|  |  | Composite | 2.99 (<0.001) | .115 ( 0.056) | .00721 (<0.001) |
| **Artificial Intelligence & Image Processing ( 6.7%)** | **Other fields** | Raw citations | 1792 (<0.001) | -76.2 ( 0.800) | -4.08 ( 0.511) |
|  |  | Composite | 2.78 (<0.001) | -.0237 ( 0.583) | .00717 (<0.001) |
| **Chemical Physics ( 6.7%)** | **Other fields** | Raw citations | 1697 (<0.001) | 264 ( 0.449) | 5.11 ( 0.312) |
|  |  | Composite | 3.03 (<0.001) | .12 ( 0.029) | .0042 (<0.001) |
| **Industrial Engineering & Automation ( 6.7%)** | **Other fields** | Raw citations | 1097 (<0.001) | 108 ( 0.543) | -2.79 ( 0.421) |
|  |  | Composite | 2.89 (<0.001) | -.0181 ( 0.777) | .00285 ( 0.022) |
| **Criminology ( 6.4%)** | **Other fields** | Raw citations | 416 (<0.001) | 170 ( 0.282) | 9.35 ( 0.004) |
|  |  | Composite | 2.99 (<0.001) | .0653 ( 0.408) | .00519 ( 0.001) |
| **Sport Sciences ( 6.3%)** | **Highly related fields** | Raw citations | 912 ( 0.003) | 742 ( 0.079) | 11 ( 0.242) |
|  |  | Composite | 3.04 (<0.001) | .0609 ( 0.522) | .00208 ( 0.328) |
| **Veterinary Sciences ( 6.2%)** | **Highly related fields** | Raw citations | 479 (<0.001) | 126 ( 0.076) | .954 ( 0.533) |
|  |  | Composite | 2.51 (<0.001) | .0793 ( 0.074) | .00298 ( 0.002) |
| **Psychoanalysis ( 6.1%)** | **Highly related fields** | Raw citations | 51 ( 0.729) | 182 ( 0.366) | 2.62 ( 0.329) |
|  |  | Composite | 2.5 (<0.001) | .236 ( 0.389) | .000928 ( 0.799) |
| **Social Work ( 6%)** | **Other fields** | Raw citations | 167 ( 0.045) | 764 (<0.001) | 7.4 ( 0.013) |
|  |  | Composite | 2.75 (<0.001) | .233 ( 0.013) | .00193 ( 0.336) |
| **Polymers ( 5.9%)** | **Other fields** | Raw citations | 1339 (<0.001) | 384 ( 0.160) | -2.5 ( 0.536) |
|  |  | Composite | 2.89 (<0.001) | -.0114 ( 0.877) | .00287 ( 0.009) |
| **General Clinical Medicine ( 5.8%)** | **Highly related fields** | Raw citations | 285 ( 0.045) | 578 ( 0.014) | 4.57 ( 0.214) |
|  |  | Composite | 2.21 (<0.001) | .0189 ( 0.889) | .00713 ( 0.001) |
| **Entomology ( 5.8%)** | **Other fields** | Raw citations | 989 (<0.001) | 461 (<0.001) | -8.53 ( 0.001) |
|  |  | Composite | 2.92 (<0.001) | .316 (<0.001) | -.00156 ( 0.262) |
| **Religions & Theology ( 4.9%)** | **Other fields** | Raw citations | 143 ( 0.002) | 12.9 ( 0.895) | -.325 ( 0.827) |
|  |  | Composite | 2.33 (<0.001) | .0544 ( 0.763) | .000519 ( 0.850) |
| **Distributed Computing ( 4.9%)** | **Other fields** | Raw citations | 352 ( 0.296) | 142 ( 0.774) | 14.3 ( 0.203) |
|  |  | Composite | 2.37 (<0.001) | .159 ( 0.267) | .0045 ( 0.167) |
| **Computer Hardware & Architecture ( 4.4%)** | **Other fields** | Raw citations | 605 (<0.001) | 80.3 ( 0.647) | 2.63 ( 0.387) |
|  |  | Composite | 2.46 (<0.001) | .0563 ( 0.528) | .00468 ( 0.003) |
| **Information & Library Sciences ( 4.2%)** | **Other fields** | Raw citations | 258 ( 0.001) | 69.6 ( 0.682) | 1.9 ( 0.429) |
|  |  | Composite | 2.62 (<0.001) | .174 ( 0.254) | .00193 ( 0.368) |
| **Education ( 4%)** | **Other fields** | Raw citations | 350 (<0.001) | 263 ( 0.008) | 7.37 (<0.001) |
|  |  | Composite | 2.89 (<0.001) | -.00199 ( 0.971) | .0042 (<0.001) |
| **Logistics & Transportation ( 4%)** | **Other fields** | Raw citations | 614 (<0.001) | 497 ( 0.121) | 8.55 ( 0.107) |
|  |  | Composite | 2.95 (<0.001) | -.0441 ( 0.757) | .00361 ( 0.128) |
| **Social Psychology ( 3.9%)** | **Highly related fields** | Raw citations | 1214 (<0.001) | 269 ( 0.497) | 9.64 ( 0.048) |
|  |  | Composite | 3.2 (<0.001) | .0938 ( 0.311) | .00439 (<0.001) |
| **Applied Mathematics ( 3.8%)** | **Other fields** | Raw citations | 908 ( 0.026) | 1266 ( 0.082) | 9.22 ( 0.337) |
|  |  | Composite | 3.05 (<0.001) | .156 ( 0.436) | .005 ( 0.061) |
| **Legal & Forensic Medicine ( 3.8%)** | **Highly related fields** | Raw citations | 277 ( 0.003) | -118 ( 0.515) | .588 ( 0.850) |
|  |  | Composite | 2.22 (<0.001) | -.037 ( 0.837) | .00542 ( 0.084) |
| **Physical Chemistry ( 3.7%)** | **Other fields** | Raw citations | 1889 (<0.001) | -162 ( 0.837) | 1.19 ( 0.893) |
|  |  | Composite | 2.92 (<0.001) | -.0478 ( 0.798) | .00667 ( 0.002) |
| **Development Studies ( 3.7%)** | **Other fields** | Raw citations | 598 ( 0.013) | 194 ( 0.603) | 1.21 ( 0.849) |
|  |  | Composite | 3.13 (<0.001) | -.137 ( 0.606) | .00221 ( 0.630) |
| **Economics ( 3.6%)** | **Other fields** | Raw citations | 739 (<0.001) | 767 (<0.001) | 4.58 ( 0.106) |
|  |  | Composite | 3.08 (<0.001) | .289 (<0.001) | .00517 (<0.001) |
| **Anatomy & Morphology ( 3.6%)** | **Highly related fields** | Raw citations | 344 (<0.001) | 60.3 ( 0.756) | -1.92 ( 0.357) |
|  |  | Composite | 2.46 (<0.001) | -.0653 ( 0.743) | -.000483 ( 0.821) |
| **Food Science ( 3.6%)** | **Other fields** | Raw citations | 1047 ( 0.002) | 882 ( 0.206) | 4.11 ( 0.659) |
|  |  | Composite | 2.88 (<0.001) | .0576 ( 0.669) | .00521 ( 0.004) |
| **Ornithology ( 3.4%)** | **Other fields** | Raw citations | 404 ( 0.002) | -16.7 ( 0.952) | .54 ( 0.850) |
|  |  | Composite | 2.57 (<0.001) | .287 ( 0.139) | .00354 ( 0.077) |
| **Geography ( 3.3%)** | **Other fields** | Raw citations | 318 ( 0.010) | -236 ( 0.348) | 8.48 ( 0.018) |
|  |  | Composite | 3 (<0.001) | -.149 ( 0.302) | .00519 ( 0.012) |
| **Electrical & Electronic Engineering ( 3.3%)** | **Other fields** | Raw citations | 426 (<0.001) | 130 ( 0.603) | 9.93 ( 0.004) |
|  |  | Composite | 2.47 (<0.001) | .0131 ( 0.903) | .00613 (<0.001) |
| **Networking & Telecommunications ( 3.3%)** | **Other fields** | Raw citations | 880 (<0.001) | -151 ( 0.387) | -1.87 ( 0.354) |
|  |  | Composite | 2.62 (<0.001) | -.0672 ( 0.289) | .00373 (<0.001) |
| **Dairy & Animal Science ( 3.3%)** | **Other fields** | Raw citations | 735 (<0.001) | 248 ( 0.076) | -3.44 ( 0.067) |
|  |  | Composite | 2.64 (<0.001) | -.0237 ( 0.792) | .0023 ( 0.058) |
| **General Psychology & Cognitive Sciences ( 3.1%)** | **Highly related fields** | Raw citations | 699 (<0.001) | -340 ( 0.344) | -.0267 ( 0.995) |
|  |  | Composite | 3.02 (<0.001) | -.286 ( 0.185) | .000522 ( 0.852) |
| **Fluids & Plasmas ( 3.1%)** | **Other fields** | Raw citations | 1638 (<0.001) | 291 ( 0.579) | -10.1 ( 0.111) |
|  |  | Composite | 3.11 (<0.001) | .157 ( 0.163) | .00125 ( 0.360) |
| **Social Sciences Methods ( 3%)** | **Other fields** | Raw citations | 1036 ( 0.073) | 2227 ( 0.117) | 8.28 ( 0.520) |
|  |  | Composite | 3.18 (<0.001) | .701 ( 0.003) | .00289 ( 0.173) |
| **Numerical & Computational Mathematics ( 3%)** | **Other fields** | Raw citations | 490 ( 0.014) | 829 ( 0.025) | 6.65 ( 0.163) |
|  |  | Composite | 3.07 (<0.001) | .364 ( 0.040) | .000555 ( 0.807) |
| **Building & Construction ( 2.9%)** | **Other fields** | Raw citations | 780 (<0.001) | 363 ( 0.457) | 12.2 ( 0.078) |
|  |  | Composite | 2.94 (<0.001) | -.0224 ( 0.898) | .00725 ( 0.004) |
| **Mechanical Engineering & Transports ( 2.9%)** | **Other fields** | Raw citations | 1048 (<0.001) | -.278 ( 0.999) | -4.51 ( 0.178) |
|  |  | Composite | 2.83 (<0.001) | -.092 ( 0.384) | .0034 ( 0.004) |
| **Science Studies ( 2.9%)** | **Other fields** | Raw citations | 217 ( 0.353) | 62.4 ( 0.894) | 12.7 ( 0.051) |
|  |  | Composite | 3 (<0.001) | -.136 ( 0.688) | .00692 ( 0.136) |
| **Cultural Studies ( 2.8%)** | **Other fields** | Raw citations | 74.7 ( 0.319) | -92.6 ( 0.578) | 2.56 ( 0.252) |
|  |  | Composite | 2.44 (<0.001) | -.17 ( 0.551) | .0056 ( 0.147) |
| **Marketing ( 2.8%)** | **Other fields** | Raw citations | 194 ( 0.380) | -19.8 ( 0.962) | 27.6 (<0.001) |
|  |  | Composite | 3.01 (<0.001) | .04 ( 0.735) | .00568 ( 0.005) |
| **Anthropology ( 2.6%)** | **Other fields** | Raw citations | 288 (<0.001) | 184 ( 0.229) | 4.94 ( 0.016) |
|  |  | Composite | 2.97 (<0.001) | -.129 ( 0.348) | .00266 ( 0.146) |
| **Materials ( 2.4%)** | **Other fields** | Raw citations | 1235 (<0.001) | 275 ( 0.415) | -1.51 ( 0.626) |
|  |  | Composite | 2.76 (<0.001) | .131 ( 0.098) | .00504 (<0.001) |
| **Operations Research ( 2.4%)** | **Other fields** | Raw citations | 518 ( 0.042) | 10.5 ( 0.985) | 12.9 ( 0.053) |
|  |  | Composite | 3.09 (<0.001) | .0935 ( 0.562) | .00317 ( 0.101) |
| **Mining & Metallurgy ( 2.3%)** | **Other fields** | Raw citations | 225 ( 0.148) | -103 ( 0.778) | 4.22 ( 0.278) |
|  |  | Composite | 2.31 (<0.001) | -.105 ( 0.755) | .00577 ( 0.113) |
| **Optoelectronics & Photonics ( 2.3%)** | **Other fields** | Raw citations | 486 (<0.001) | 327 ( 0.052) | 1.69 ( 0.360) |
|  |  | Composite | 2.31 (<0.001) | .166 ( 0.050) | .00571 (<0.001) |
| **Human Factors ( 2.3%)** | **Highly related fields** | Raw citations | 731 (<0.001) | 266 ( 0.487) | 7.31 ( 0.141) |
|  |  | Composite | 2.88 (<0.001) | .218 ( 0.121) | .00767 (<0.001) |
| **Software Engineering ( 2.2%)** | **Other fields** | Raw citations | 1082 (<0.001) | 240 ( 0.313) | -12.9 (<0.001) |
|  |  | Composite | 2.8 (<0.001) | -.162 ( 0.189) | .000988 ( 0.568) |
| **Finance ( 2.2%)** | **Other fields** | Raw citations | 568 ( 0.004) | -345 ( 0.451) | 14.1 ( 0.018) |
|  |  | Composite | 3 (<0.001) | -.0545 ( 0.743) | .00713 ( 0.001) |
| **Meteorology & Atmospheric Sciences ( 2.2%)** | **Other fields** | Raw citations | 1773 (<0.001) | 157 ( 0.592) | 1.23 ( 0.733) |
|  |  | Composite | 3.09 (<0.001) | .0653 ( 0.287) | .00335 (<0.001) |
| **Communication & Media Studies ( 2.1%)** | **Other fields** | Raw citations | 107 ( 0.444) | 129 ( 0.770) | 20.5 (<0.001) |
|  |  | Composite | 3.04 (<0.001) | .00959 ( 0.945) | .00424 ( 0.006) |
| **Econometrics ( 2%)** | **Other fields** | Raw citations | 624 ( 0.460) | 505 ( 0.769) | 26.3 ( 0.243) |
|  |  | Composite | 3.25 (<0.001) | -.296 ( 0.534) | .00695 ( 0.264) |
| **Political Science & Public Administration ( 2%)** | **Other fields** | Raw citations | 463 (<0.001) | 1092 (<0.001) | 6.15 ( 0.040) |
|  |  | Composite | 3.05 (<0.001) | .312 ( 0.007) | .00406 ( 0.001) |
| **Languages & Linguistics ( 2%)** | **Other fields** | Raw citations | 422 (<0.001) | 35.6 ( 0.860) | .804 ( 0.738) |
|  |  | Composite | 3.04 (<0.001) | .182 ( 0.340) | .00281 ( 0.218) |
| **General Physics ( 2%)** | **Other fields** | Raw citations | 1270 (<0.001) | 381 ( 0.335) | -6.75 ( 0.032) |
|  |  | Composite | 2.78 (<0.001) | .101 ( 0.419) | .00337 ( 0.001) |
| **Archaeology ( 2%)** | **Other fields** | Raw citations | 418 (<0.001) | -1.75 ( 0.995) | .99 ( 0.726) |
|  |  | Composite | 2.81 (<0.001) | .0493 ( 0.789) | .00164 ( 0.344) |
| **Sport, Leisure & Tourism ( 1.9%)** | **Other fields** | Raw citations | 696 (<0.001) | -46.8 ( 0.900) | 4.47 ( 0.387) |
|  |  | Composite | 3.08 (<0.001) | .0348 ( 0.850) | .00429 ( 0.096) |
| **Environmental Engineering ( 1.9%)** | **Other fields** | Raw citations | 1071 (<0.001) | 1489 (<0.001) | -2.14 ( 0.608) |
|  |  | Composite | 2.97 (<0.001) | .156 ( 0.188) | .00197 ( 0.127) |
| **Energy ( 1.9%)** | **Other fields** | Raw citations | 1012 (<0.001) | 454 ( 0.087) | .673 ( 0.784) |
|  |  | Composite | 2.85 (<0.001) | .0496 ( 0.540) | .00395 (<0.001) |
| **Marine Biology & Hydrobiology ( 1.7%)** | **Other fields** | Raw citations | 1519 (<0.001) | 379 ( 0.355) | -8.78 ( 0.094) |
|  |  | Composite | 3.05 (<0.001) | .0472 ( 0.711) | .00222 ( 0.172) |
| **Ecology ( 1.6%)** | **Other fields** | Raw citations | 1288 (<0.001) | 263 ( 0.563) | 11.7 ( 0.024) |
|  |  | Composite | 3.07 (<0.001) | .154 ( 0.095) | .00552 (<0.001) |
| **Chemical Engineering ( 1.6%)** | **Other fields** | Raw citations | 1543 (<0.001) | -113 ( 0.850) | -8.93 ( 0.056) |
|  |  | Composite | 2.98 (<0.001) | .215 ( 0.265) | .00205 ( 0.170) |
| **Philosophy ( 1.6%)** | **Other fields** | Raw citations | 242 (<0.001) | 476 (<0.001) | .346 ( 0.761) |
|  |  | Composite | 2.91 (<0.001) | .712 (<0.001) | .000684 ( 0.676) |
| **Aerospace & Aeronautics ( 1.6%)** | **Other fields** | Raw citations | 403 (<0.001) | 142 ( 0.229) | -.121 ( 0.913) |
|  |  | Composite | 2.44 (<0.001) | -.0368 ( 0.723) | .00447 (<0.001) |
| **Inorganic & Nuclear Chemistry ( 1.5%)** | **Other fields** | Raw citations | 951 (<0.001) | -182 ( 0.828) | 4.34 ( 0.458) |
|  |  | Composite | 2.77 (<0.001) | -.0123 ( 0.951) | .00294 ( 0.036) |
| **Agricultural Economics & Policy ( 1.4%)** | **Other fields** | Raw citations | 990 (<0.001) | -8.71 ( 0.989) | -2.71 ( 0.698) |
|  |  | Composite | 3.24 (<0.001) | .033 ( 0.915) | -.0016 ( 0.641) |
| **Geological & Geomatics Engineering ( 1.4%)** | **Other fields** | Raw citations | 1385 (<0.001) | 399 ( 0.462) | -2.71 ( 0.616) |
|  |  | Composite | 2.98 (<0.001) | .19 ( 0.233) | .00485 ( 0.002) |
| **Accounting ( 1.4%)** | **Other fields** | Raw citations | 465 ( 0.014) | -178 ( 0.713) | 12.2 ( 0.036) |
|  |  | Composite | 2.92 (<0.001) | .0437 ( 0.875) | .00806 ( 0.017) |
| **Applied Physics ( 1.2%)** | **Other fields** | Raw citations | 1454 (<0.001) | 257 ( 0.535) | -2.28 ( 0.392) |
|  |  | Composite | 2.84 (<0.001) | .0137 ( 0.874) | .00263 (<0.001) |
| **Business & Management ( 1.2%)** | **Other fields** | Raw citations | 761 (<0.001) | 14.3 ( 0.971) | 12.4 ( 0.001) |
|  |  | Composite | 3.11 (<0.001) | .0147 ( 0.894) | .00466 (<0.001) |
| **Information Systems ( 1.2%)** | **Other fields** | Raw citations | 1196 (<0.001) | 629 ( 0.504) | -1.17 ( 0.906) |
|  |  | Composite | 3.16 (<0.001) | .321 ( 0.195) | .000241 ( 0.926) |
| **Computation Theory & Mathematics ( 1.1%)** | **Other fields** | Raw citations | 696 (<0.001) | -77.8 ( 0.878) | -.305 ( 0.939) |
|  |  | Composite | 2.93 (<0.001) | .0902 ( 0.675) | .00232 ( 0.173) |
| **Astronomy & Astrophysics ( .94%)** | **Other fields** | Raw citations | 3121 (<0.001) | 113 ( 0.898) | -23.8 (<0.001) |
|  |  | Composite | 3.17 (<0.001) | .101 ( 0.332) | .000634 ( 0.428) |
| **Geochemistry & Geophysics ( .81%)** | **Other fields** | Raw citations | 1302 (<0.001) | -173 ( 0.583) | -4.81 ( 0.042) |
|  |  | Composite | 3.15 (<0.001) | -.0982 ( 0.408) | .00175 ( 0.049) |
| **Strategic, Defence & Security Studies ( .77%)** | **Other fields** | Raw citations | 244 ( 0.106) | -462 ( 0.483) | 11.1 ( 0.013) |
|  |  | Composite | 2.7 (<0.001) | -.125 ( 0.695) | .00778 (<0.001) |
| **Paleontology ( .74%)** | **Other fields** | Raw citations | 1198 (<0.001) | -586 ( 0.551) | -4.82 ( 0.489) |
|  |  | Composite | 3.14 (<0.001) | -.258 ( 0.241) | -.0000303 ( 0.984) |
| **Literary Studies ( .72%)** | **Other fields** | Raw citations | 30.3 ( 0.414) | 67.7 ( 0.648) | 1.96 ( 0.143) |
|  |  | Composite | 1.93 (<0.001) | .635 ( 0.089) | .00804 ( 0.017) |
| **General Mathematics ( .71%)** | **Other fields** | Raw citations | 447 (<0.001) | 391 ( 0.094) | -.82 ( 0.452) |
|  |  | Composite | 2.9 (<0.001) | .0651 ( 0.685) | .000633 ( 0.399) |
| **Nuclear & Particle Physics ( .63%)** | **Other fields** | Raw citations | 3770 (<0.001) | 2194 ( 0.129) | -39.9 (<0.001) |
|  |  | Composite | 2.89 (<0.001) | .224 ( 0.106) | .00254 (<0.001) |
| **Oceanography ( .6%)** | **Other fields** | Raw citations | 1004 (<0.001) | 262 ( 0.634) | -5.42 ( 0.136) |
|  |  | Composite | 2.92 (<0.001) | -.267 ( 0.320) | .00165 ( 0.352) |
| **Agronomy & Agriculture ( .55%)** | **Other fields** | Raw citations | 1183 (<0.001) | 839 ( 0.265) | -3.06 ( 0.455) |
|  |  | Composite | 2.88 (<0.001) | .343 ( 0.133) | .00282 ( 0.024) |
| **Fisheries ( .53%)** | **Other fields** | Raw citations | 710 (<0.001) | 425 ( 0.393) | .938 ( 0.788) |
|  |  | Composite | 2.77 (<0.001) | .109 ( 0.701) | .00287 ( 0.151) |
| **Zoology ( 0%)** | **Other fields** | Raw citations |  |  |  |
|  |  | Composite |  |  |  |
| **Urban & Regional Planning ( 0%)** | **Other fields** | Raw citations |  |  |  |
|  |  | Composite |  |  |  |
| **Music ( 0%)** | **Other fields** | Raw citations |  |  |  |
|  |  | Composite |  |  |  |
| **Mathematical Physics ( 0%)** | **Other fields** | Raw citations |  |  |  |
|  |  | Composite |  |  |  |
| **Law ( 0%)** | **Other fields** | Raw citations |  |  |  |
|  |  | Composite |  |  |  |
| **International Relations ( 0%)** | **Other fields** | Raw citations |  |  |  |
|  |  | Composite |  |  |  |
| **Industrial Relations ( 0%)** | **Other fields** | Raw citations |  |  |  |
|  |  | Composite |  |  |  |
| **Horticulture ( 0%)** | **Other fields** | Raw citations |  |  |  |
|  |  | Composite |  |  |  |
| **History of Social Sciences ( 0%)** | **Other fields** | Raw citations |  |  |  |
|  |  | Composite |  |  |  |
| **History of Science, Technology & Medicine ( 0%)** | **Other fields** | Raw citations |  |  |  |
|  |  | Composite |  |  |  |
| **History ( 0%)** | **Other fields** | Raw citations |  |  |  |
|  |  | Composite |  |  |  |
| **Geology ( 0%)** | **Other fields** | Raw citations |  |  |  |
|  |  | Composite |  |  |  |
| **Gender Studies ( 0%)** | **Highly related fields** | Raw citations |  |  |  |
|  |  | Composite |  |  |  |
| **Forestry ( 0%)** | **Other fields** | Raw citations |  |  |  |
|  |  | Composite |  |  |  |
| **Folklore ( 0%)** | **Other fields** | Raw citations |  |  |  |
|  |  | Composite |  |  |  |
| **Economic Theory ( 0%)** | **Other fields** | Raw citations |  |  |  |
|  |  | Composite |  |  |  |
| **Classics ( 0%)** | **Other fields** | Raw citations |  |  |  |
|  |  | Composite |  |  |  |
| **Civil Engineering ( 0%)** | **Other fields** | Raw citations |  |  |  |
|  |  | Composite |  |  |  |
| **Automobile Design & Engineering ( 0%)** | **Other fields** | Raw citations |  |  |  |
|  |  | Composite |  |  |  |
| **Art Practice, History & Theory ( 0%)** | **Other fields** | Raw citations |  |  |  |
|  |  | Composite |  |  |  |
| **Architecture ( 0%)** | **Other fields** | Raw citations |  |  |  |
|  |  | Composite |  |  |  |
